# Big Data Analysis of Electronic Health Records: Clinically interpretable representations of older adult inpatient trajectories using time-series numerical data and Hidden Markov Models

**DOI:** 10.1101/2021.06.18.21258885

**Authors:** Maria Herrero-Zazo, Tomas Fitzgerald, Vince Taylor, Helen Street, Afzal N Chaudhry, John Bradley, Ewan Birney, Victoria L Keevil

## Abstract

The implementation of Electronic Health Records (EHR) in UK hospitals provides new opportunities for clinical ‘big data’ analysis. The representation of observations routinely recorded in clinical practice is the first step to use these data in several research tasks. Anonymised data were extracted from 11 158 first emergency admission episodes (AE) in older adults. Irregular records from 23 laboratory blood tests and vital signs were normalized and regularised into daily bins and represented as numerical multivariate time-series (MVTS). Unsupervised Hidden Markov Models (HMM) were trained to represent each day of each AE as one of 17 state spaces. The visual clinical interpretation of these states showed remarkable differences between patients who died at the end of the AE and those who were discharged. All states had marked features that allowed their clinical interpretation and differentiation between those associated with the patients’ disease burden, their physiological response to this burden or the stage of admission. The most evident relationships with hold-out clinical information were also confirmed by Chi-square tests, with two states strongly associated with inpatient mortality (IM) and 12 states (71%) associated with at least one admission diagnosis. The potential of these data representations on prediction of hospital outcomes was also explored using Logistic Regression (LR) and Random Forest (RF) models, with higher prediction performance observed when models were trained with MVTS data compared to HMM state spaces. However, the outputs of generative and discriminative analyses were complementary. For example, highest ranking features of the best performing RF model for IM (ROC-AUC 0.85) resembled the laboratory blood test and vital sign variables characterising the *‘Early Inflammatory Response-like’* state, itself strongly associated with IM. These results provide evidence of the capability of generative models to extract biological signals from routinely collected clinical data and their potential to represent interpretable patients’ trajectories for future research in hypothesis generation or prediction modelling.

## 1. INTRODUCTION

The National Health Service (NHS) provides medical care to 66 million UK residents from ‘cradle to grave’ and is undergoing digital transformation. This is presenting opportunities to improve the delivery, quality and safety of healthcare [1,2] and the research potential of real world healthcare data is well recognised [3,4]. However, although key descriptors of inpatient clinical activity, such as procedure or diagnostic codes, have been available for some time [5], hospital records have been largely digitally inaccessible. Thus, inpatient treatment decisions and service developments have not benefited from research using the wealth of routinely collected healthcare data available.

The implementation of Electronic Health Records (EHRs) in NHS hospitals heralds an opportunity to address this. Comprehensive real-time clinical information is recorded for every patient from admission to discharge. This generates large and detailed datasets, with thousands of data pertaining to thousands of patients collected and stored every day. Machine Learning (ML) can unlock the research potential of this ‘big data’ by unravelling hidden relationships within its large and complex structure. In selected settings, hospital EHRs have already been used to explore how ML could augment development of stratified medicine, automated medical image analysis, and the prediction of clinical diagnoses and outcomes [6–9].

This potential for ML to transform healthcare delivery [10] has not yet been realised. Ethical and legal guidance, balancing the benefits of using routinely collected healthcare data for research against potential harm through breaches in patient confidentiality, is evolving. Data security can limit access to and sharing of data for ML, and data quality varies across organisations and other features of collection framework [11,12]. Data are collected with the primary purpose of supporting clinical care and different mechanisms for entering information can complicate data extraction. Additionally, datasets often contain heterogeneous entries. Not only is there unstructured, semi-structured and structured data, even amongst structured numerical variables there is variation in scale, frequency, regularity and completeness. Therefore, generation of a research ready dataset from hospital EHRs requires clinical, technical, ethical and information governance expertise.

Numerical time-series data, such as commonly measured laboratory blood tests and bedside vital signs, is a sub-group of hospital EHR data that presents a rich longitudinal account of patients’ metabolic and physiologic profiles throughout an admission. However, given the irregularity and heterogeneity in these data it is not clear how data should be optimised for ML analyses. Although different ML techniques require different degrees of feature engineering [13], most ML methods benefit from an intermediate, regularised representation of the raw data, with this representation used as input data to train prediction models [14–16]. A particular task in real world data is how to handle missing entries. While some ML techniques can handle missing data implicitly in their scheme, most of them require a complete dataset and, therefore, an imputation step prior to model training. The missingness, distribution and normalisation of these data in general inpatient populations is also infrequently studied, partly because there are few publicly available hospital EHR datasets. The ‘Medical Information Mart for Intensive Care’ database is a notable exception, but focuses on intensive care patients only [17]. Furthermore, some approaches ignore test values and only include the occurrence of a test, reflecting a previous reliance on code based representations of EHR data [8].

Representations of EHR data are usually evaluated on prediction tasks and by the performance of ML models, measured by metrics such as recall and precision [9,18,19]. However, clinicians often want to evaluate associations between features and outcomes to make hypotheses about causal pathways. This is hard to do from outcome based ML models, which by design have been developed to maximise prediction, and this ‘black box’ of ML limits the clinical translation of research findings [14,20,21]. Successful representations of EHR data need to address the interpretability of ML models as well as their performance on prediction tasks. Additionally, less emphasis has been placed on simply exploring hospital EHR data, and thus the inherent variation in patient populations, which will also be of value to clinicians trying to understand relationships between health determinants and outcomes or disease clusters.

Considering this current landscape, we formed a collaboration between clinicians, data scientists, research governance experts and clinical informatics specialists. Our shared aim was to extract comprehensive, anonymised, high quality data from an EHR at a tertiary care NHS hospital and analyse it for exploration of the inherent variation in the dataset and certain straightforward prediction tasks. We focus on data from older adult inpatients admitted as an emergency under any hospital specialty. Older adults place a disproportionately high demand on emergency inpatient services [22] and relatively modest gains in healthcare delivery and effectiveness can have large system wide effects. We aim to develop methods of representing hospital EHR data from this heterogenous inpatient group for ML analyses, particularly exploring pre-processing approaches that work across a range of modern and classical ML methods. We also explore the interpretable representation of inpatient trajectories, derived from irregular time-series numerical data including 23 commonly measured laboratory blood test and vital sign values. Thus, our main goal is to use generative, unbiased ML techniques (Hidden Markov Models), which make only broad assumptions of the data, and to compare the outcome of these models to hold-out clinical diagnoses and outcome information for clinical interpretation. We also evaluate the potential of these representations as input variables for hypothesis driven discriminative models, preregistering our hypotheses explicitly using the Open Science Foundation scheme (https://osf.io/6zp3d).

## 2. METHODS

### 2.1. Collaboration

The primary collaboration was between clinician scientists at Cambridge University Hospitals NHS Foundation Trust (CUHNFT), and technical experts in artificial intelligence at the European Molecular Biology Laboratory’s European Bioinformatics Institute (EMBL-EBI). This collaboration was supported by Clinical Informatics and Research Governance experts at CUHNFT.

### 2.2 Ethics

The project was approved by the NHS Health Research Authority (HRA) (IRAS: 253457), North East – Newcastle & North Tyneside 1 Research Ethics Committee (REC) (REC reference: 19/NE/0013) and by the EMBL Scientific Advisory Committee (BIAC).

### 2.3 Data

Data from all patients aged 65 years or older admitted to Addenbrooke’s Hospital as an emergency between January 2015 and December 2019 were retrieved by the hospital’s Clinical Informatics team from the Epic EHR system introduced in October 2014. Supplementary Table 1 summarises the variables retrieved and the anonymisation process, which was developed by clinical and research governance experts. In brief, patients were characterised by five-year age groups, sex, month of admission, hospital admission and discharge service, clinical frailty scale (CFS) score [23], disease phenotype information at admission and discharge (top-level ICD-10 category class), laboratory blood test results (comprised of clinically relevant and commonly requested tests), bedside vital signs (indicative of a patient’s physiological status and illness acuity), length of inpatient stay and clinical outcomes of the admission episode (AE) including: discharge alive (DA), inpatient mortality (IM, referred to as Inpatient Death [ID] in multiclass outcome analyses), 30-day post-discharge readmission (PDR), post-discharge readmission and 30-day post-discharge mortality (PDRM) and 30-day post-discharge mortality (PDM). As explained below, after data pre-processing a ‘hold-out validation’ dataset was kept aside for confirmatory analyses.

### 2.4 Data transfer and access

A bespoke data sharing agreement was established between CUH and EMBL-EBI. Anonymised data was uploaded to a private and password protected STP site hosted by the EBI server and stored using electronic protection from UNIX security models. Data is shared through a dataset specific UNIX group and all individuals granted access to the data receive internal training and acknowledge their responsibilities before being granted access to the data. The lead technical and clinical analysts also hold honorary contracts with CUHNFT and EMBL-EBI respectively. No data is stored outside of the controlled, project storage volumes at EMBL-EBI.

### 2.5. Pre-processing: data cleaning, normalization, and regularization for time-series numerical data

23 different variables denoted laboratory blood test results (17/23) and vital signs (5/23). Data were first normalized for each variable using inverse rank normalization (IRN) to reduce the impact of outliers without having to define maximum or minimum reference levels. We defined 24-hour bins running from midnight to midnight and aimed to select a unique observation for each lab test or vital sign in each daily bin. Blood tests are not often measured more than once per day but if multiple observations were recorded on the same day, the earliest record was selected as the unique value. The time the blood sample was taken from the patient was defined as the ‘reference time’ for each daily bin for each patient and the closest vital sign measurements to this reference time were selected, since vital signs are usually measured several times per day. Observations not recorded for a given daily bin were considered ‘missing observations’. Values for laboratory tests were also discarded if the sample collection time was recorded as later than the time test results were reported, if they contained non-numeric symbols, or were duplicate entries. Point-of-care (POC) and non-POC results for the same laboratory test were considered as the same variable, although it is recognised that the laboratory procedures are different. A similar process of data cleaning and normalisation was conducted for vital signs.

After data cleaning, normalisation and regularisation some daily bins had none or a very small number of non-recorded laboratory test and vital sign values, whilst others had a large proportion of missingness. Daily bins were defined as *‘rich-information’* or *‘poor-information’* days depending on the number of recorded and missing observations. A *‘rich-information’* day was defined as a day with information for at least 4 vital signs and 14 laboratory test values.

### 2.6 Patient cohort

AEs were included if patients were admitted to a hospital ward (i.e., not discharged from the Emergency Department) and if information on disease phenotype on admission (admission diagnosis) or discharge was available. Only the first AE within the study period for each patient was included and the length of stay was set at a minimum of 3 days, since our aim was to focus on time-series numerical data.

Additional inclusion criteria were defined to avoid potential biases due to imputation of a large number of missing values whilst not restricting the cohort to a very closely monitored sub-sample. AEs were included if the first and last days were *‘rich-information’* days and if *‘rich-information’* days accounted for at least 2/3 of the length of the time series. The minimum number of recorded laboratory test results (≥ 14 out of 18) used to define a *‘rich-information’* day was chosen in an iterative process as the largest number leading to the desired sample size (around 10,000 patients).

The final dataset was randomly divided into a ‘training and test’ set (80% AE) for exploratory analyses and a ‘hold-out validation’ set (20% AE) for confirmatory analyses.

### 2.7 Imputation of missing values

Imputation was a two-step process: multiple imputation (MI) of *‘rich-information’* days and linear interpolation (LI) imputation of *‘poor-information’* days. MI is the method of choice for complex incomplete data, and is commonly used in clinical research [24]. MI was implemented with Predictive Mean Matching (PMM) and conducted for each time interval (i.e., it does not specifically include information on the time series) using information from the same and the other patients in the cohort. Explanatory variables were defined for each predicted variable excluding 1) variables with the largest fractions of missingness in the dataset (glucose, alkaline phosphatase [alp], alanine transaminase [alt], bilirubin, urea and respiratory rate); 2) variables with highly correlated missingness patterns (usually variables requested as a batch, i.e., alt and alp); and 3) highly correlated variables (e.g., neutrophils and white blood cell counts [WBC]). Age, sex, discharge specialty and primary diagnosis at admission were also included as predictors. The predictors matrix is shown in Supplementary Figure 1D. The dataset was imputed ten times.

LI imputation assumes a linear relationship between data points in the time series and relies on non-missing values from adjacent observations to compute the missing value [25]. LI imputation was conducted at the patient and variable level meaning the imputed value is independent of other variables and other patients. The R packages *mice* and *imputeTS* were used for MI and LI imputation, respectively.

Imputation was evaluated by comparing the distribution of original and imputed values and analysing the convergence plots for MI. The distributions of randomly inserted missing values before imputation versus original missing and non-missing values for MI and LI were also compared. The potential impact of imputed values in the output of the HMM model was assessed in the visual interpretation analysis of the HMM states representation.

### 2.8 Generative models for multivariate time-series representation: HMM

An unsupervised HMM was trained using expectation maximisation with the numeric multivariate time series (23 laboratory test results and vital signs) as input data. The HMM framework makes unrealistic assumptions about human biology, such as that a small number of disjoint states are an adequate model for a patient’s internal physiological state, but it requires a minimum of parameters (in effect, only the number states to train). Additionally, being a generative model, it does not require an explicit discriminatory function and no diagnosis or outcome data was used in the model, or in its training or assessment. One can consider the HMM technique as a dimensionality reduction of the multivariate space of patients, which has a natural time dependence. In this case, transforming MVTS data into a simpler state space which maximises the likelihood of the observed data and captures some dominant aspects of the patient’s condition on each day. Thus, inpatients’ trajectories are represented as a univariate discrete time series, with each day of each AE represented as a unique state space, instead of 23 numeric variables.

An HMM with Gaussian emissions, full covariance matrix and 17 states was trained and fitted in the entire ‘training and test’ dataset with the Python package *hmmlearn*. The number of states set in the model was selected in a 2-fold cross validation process where the ‘training and test’ set was randomly divided into two subsets, which were separately used to train a model and then fit the entire dataset. The output of both models was compared on a concordance matrix of co-occurrences for patient and day with the aim to find an association between states in both output datasets (i.e., a model trained with n states in two different datasets would lead to similar results or predicted states), while selecting a number of states large enough to capture the heterogeneity of the data. Chi-square tests showed that the p-values for all tested numbers of states (2 to 50) were significantly different (p-value <0.0001). The visualization of Pearson residuals and contribution plots (the contribution of a given cell to the total Chi-square score) showed that 17 states provided good concordance between states pairs without being too restrictive (Supplementary Figure 2).

A clinician (VLK) first inspected the distribution (mean values and variance) of each of the vital signs and laboratory test values stratified by state. This information was combined with a visual evaluation of: 1) the temporal distribution of the state within AE; 2) the distribution of the state across AE organised by the primary admission diagnosis; and 3) the distribution of the state in those discharged alive compared to those who died during the inpatient episode. Then formal association analyses were conducted using Chi-square tests to confirm the expert interpretation.

Reproducibility of the results was assessed by re-training the HMM algorithm in the ‘training and test’ dataset and fitting the model to the ‘hold-out validation’ dataset. The distribution of the variables was compared to map the resulting states to those in the initial training and Pearson correlation between their means was calculated.

### 2.9 Prediction models in the first days of admission

In addition to the generative modelling, we also wanted to explore the best timeframe and dataset for making discriminatory decisions. This is an obvious use of EHR data and clinically it is of interest to identify patients at higher risk early after admission. Two well established fixed frame ML techniques were employed, logistic regression (LR) and random forest (RF). LR and RF models were trained to evaluate their performance on the prediction of AE outcome.

Specifically, the impact of combining patients’ representations during the first three days of admission (either the MVTS data or the HMM state spaces) with information on age, sex, CFS score and primary admission diagnosis was assessed and compared. LR models were trained for binary classification only (i.e., prediction of IM) and RF models for binary, multiclass (prediction of five disjoint outcome classes: ID, DA, PDR, PDRM and PDM; and prediction of primary diagnosis at admission [PDA]) and multilabel classification (prediction of diagnoses at discharge [DD]).

The ‘training and test’ dataset was divided into a training set (80%) for 5-fold cross validation hyperparameter tuning and a test set (20%) to evaluate the performance of the model with the selected best parameters. The model was trained in the training set and evaluated in the test set ten times to calculate the mean of the performance metrics. All the trained models were then run against the ‘hold-out validation’ dataset without additional training.

## 3. RESULTS

### 3.1. Patient cohort: characteristics and evaluation of data processing

The initial dataset included more than 146 000 AE from 61 513 patients. After inclusion criteria, the final cohort comprised 11 158 unique AE equating to the same number of unique patients. The cohort characteristics are summarised in Supplementary Table 2.

Data was imputed for 25.5% of laboratory test values (7.8% MI; 17.7% LI) and 0.3% (0.26% MI; 0.04% LI) of vital sign values in the ‘training and test’ dataset. Kernel density estimates of the imputed and observed data for each variable are shown in Supplementary Figure 3A. The distributions match well for all variables except those with very small numbers of missing observations (i.e., 4 to 36 imputed values). MI is an iterative process and convergence plots show the mean and standard deviation of the synthetic values plotted against iteration number for the imputed data (Supplementary Figure 3B). The lack of trends and the mixing of streams are indicative of convergence of the algorithm.

To further assess the imputation method, we randomly introduced missing observations before imputation. For MI, the number of randomly inserted missing observations was selected to increase the total fraction of missingness to 5% for each variable, while for LI missingness was increased to 1/3 of each time series while always maintaining the first and last observations. The distributions after imputation match well for inserted missing observations and real (non-missing) observations for all variables, even those with very low number of actual missing values (Supplementary Figure 3C). The same conclusions were observed in the evaluation of LI imputation (Supplementary Figure 3D).

Lastly, we inspected the distributions of imputed and original observations for each of the 17 HMM states (Supplementary Figures 4). Distributions of imputed and original observations were similar across all states for most variables, except variables with very low numbers of imputed values. These did not alter the key defining characteristics of each state.

### 3.2. Clinical interpretation of HMM states

We modelled the time-dependent multivariate signal for each patient using the well-established machine learning approach of Hidden Markov Models (HMM) so that each day of each AE is represented by one of 17 possible state spaces (see methods section 2.5). Using the HMM framework only requires selecting the number of states (see methods) and requires no other additional training data (e.g., outcomes or diagnoses).

An important aspect was understanding how the HMM organised the data and facilitating clinical interpretation. Figure 3 shows the overall proportion of HMM states grouped by inpatient mortality and AE trajectories represented as the different states. Figure 3a and 3b demonstrate that some states were far more common in patients discharged alive (e.g., state q) rather than dead (e.g., state a). This overall pattern also matched the time trajectory of patients (Figure 3C), with states associated with patients finally being discharged alive enriched at the end of the admission period in those discharged alive.

**Figure 1.**
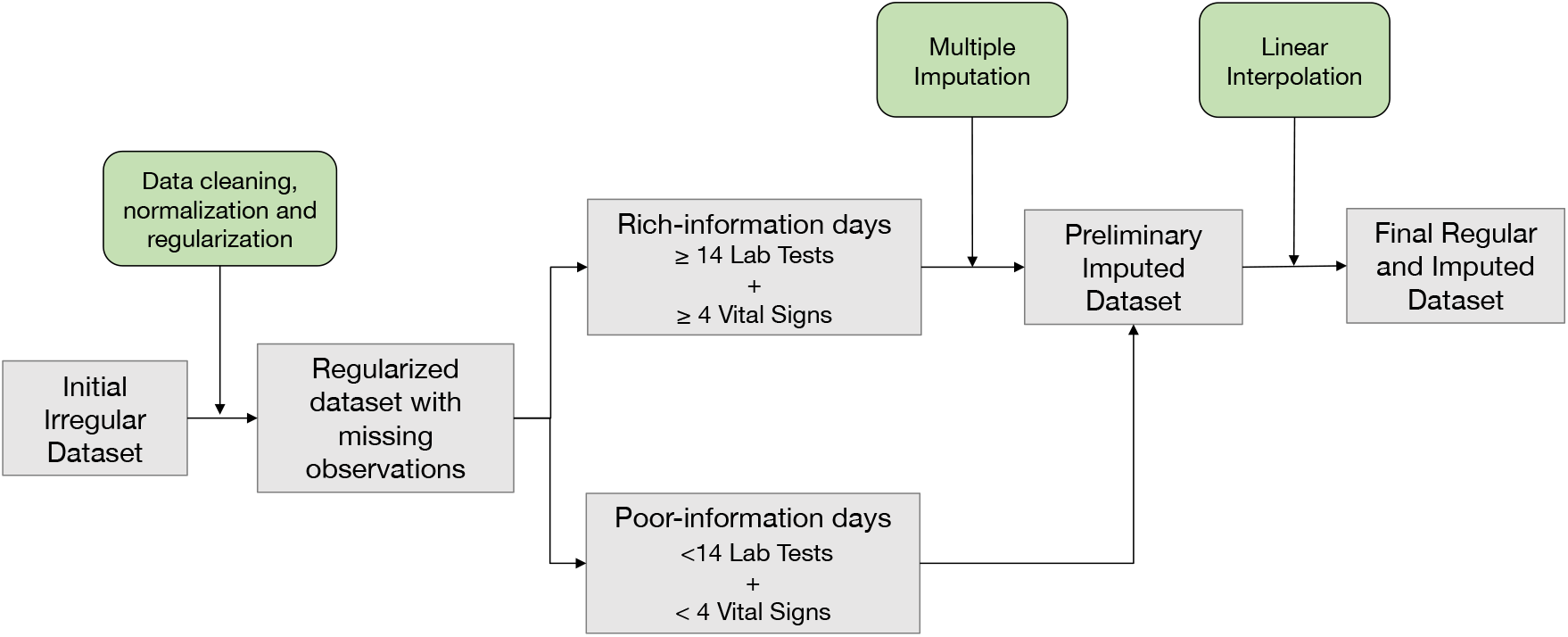
Summary of data pre-processing and imputation

**Figure 2.**
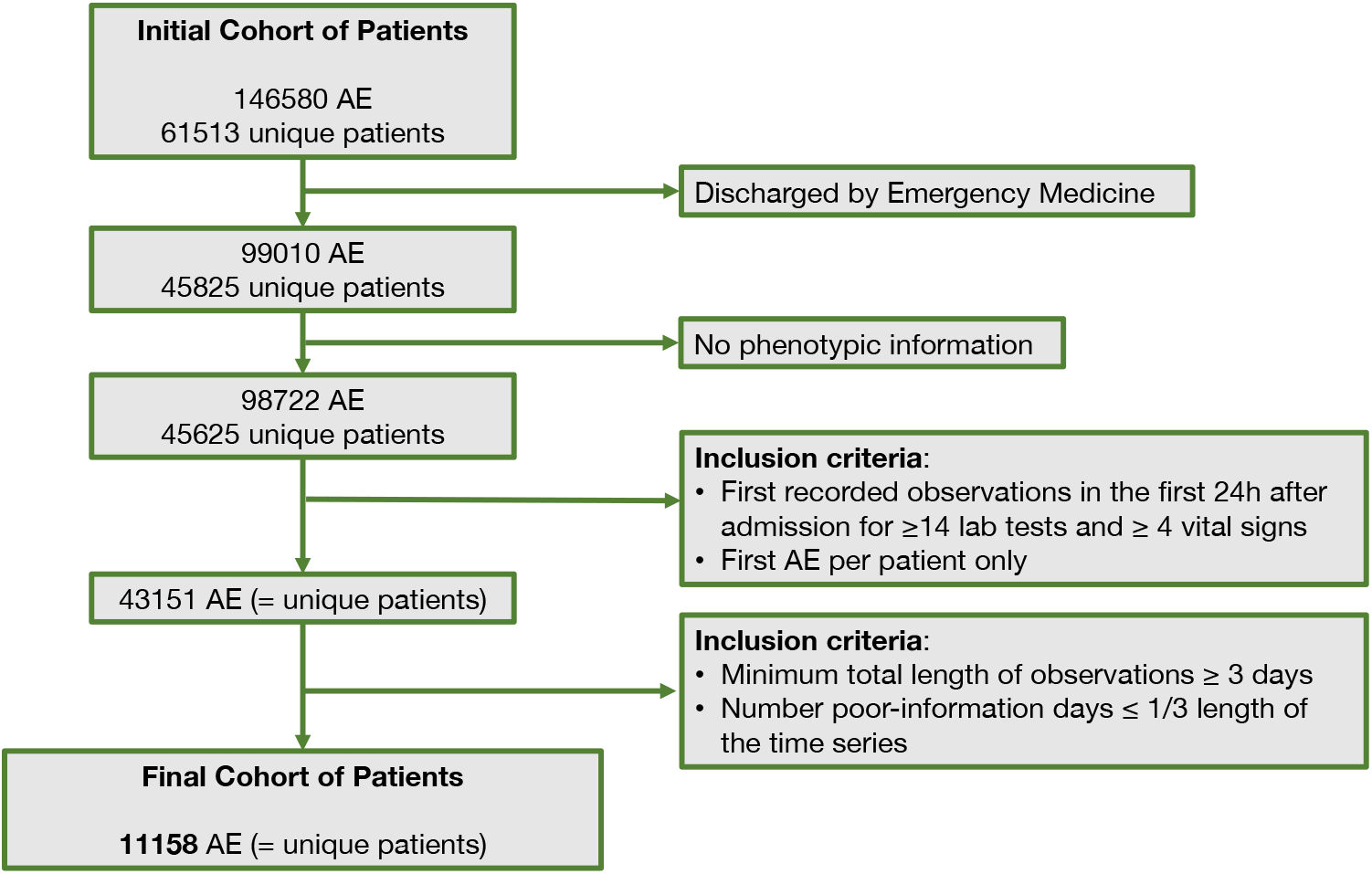
Description of inclusion criteria to select the final cohort of patients; AE: Admission episode.

**Figure 3.**
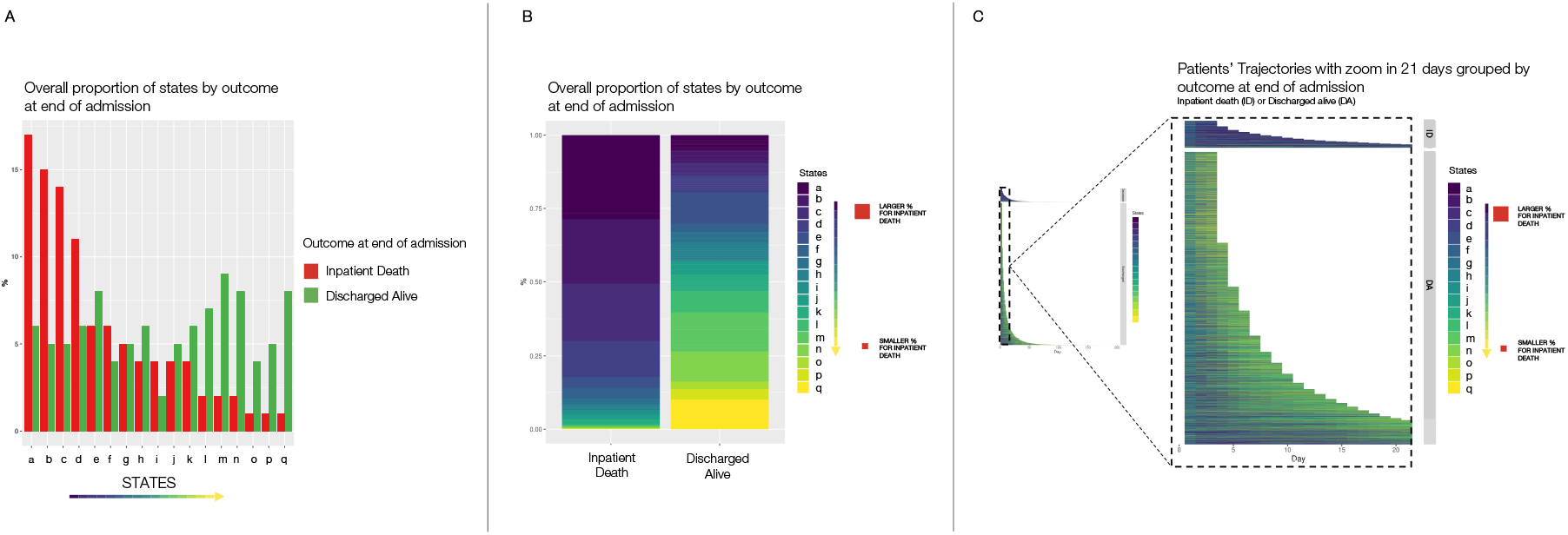
Representation of the relationship between states and outcome at the end of admission (inpatient death [ID] or discharged alive [DA]). A and B: Overall proportion of states by outcome at end of admission episode ranked by higher proportion on the inpatient death group. C: Representation of patients’ trajectories as the different states for each day with a zoom on the first 21 days of admission.

Figures 4A and 4B similarly visualise the first day of admission and show that most patients shared the same state on Day 1 (state h), whether they were discharged alive or died during the inpatient episode. However, on Day 2 patients transitioned to a greater range of states and the distributions of these differed depending on whether patients died during the inpatient episode or not (Figure 4C). These differences were even greater when examining the last day of admission (Figure 4D).

**Figure 4.**
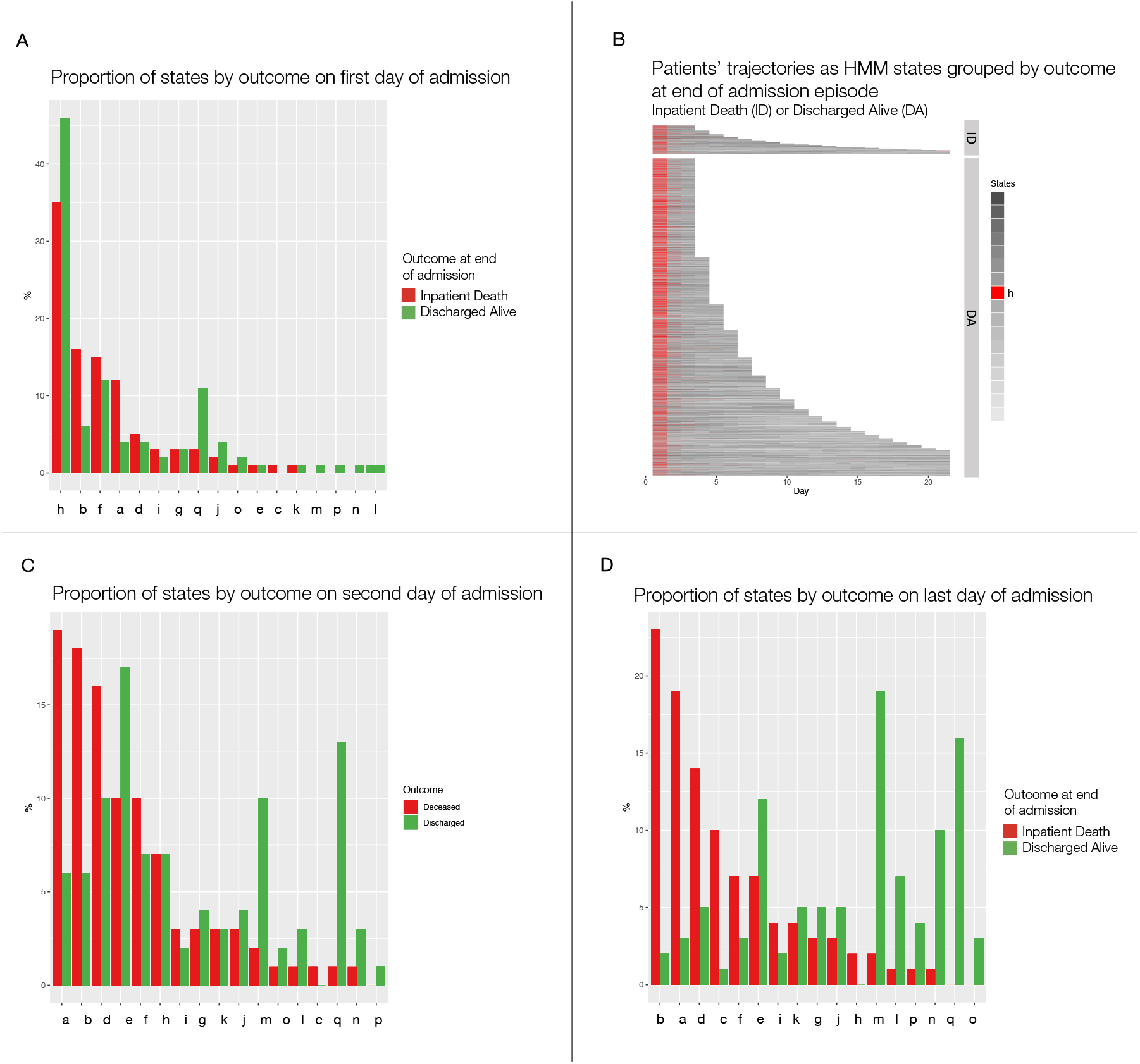
Overall proportion of states on first (A), second (C) and last day of admission (D). The most common state on day one of admission is highlighted on the representation of patients’ trajectories during the first 21 days and grouped by outcome at the end of admission (B).

An expert clinician provided an overall broad clinical interpretation of each state using output information from the HMM detailed in section 2.5 and Supplementary Figure 5, with explanatory visualisations shown in Figures 5-7. The clinician also provided an overall classification of the states, dividing them into three groups based upon the predominant feature that delineated them from other states: *‘Disease-like’, ‘Admission-like’* and *‘Physiological-like’*. The HMM state spaces are summarised in Table 1 and further detailed in Supplementary Table 3.

**Figure 5.**
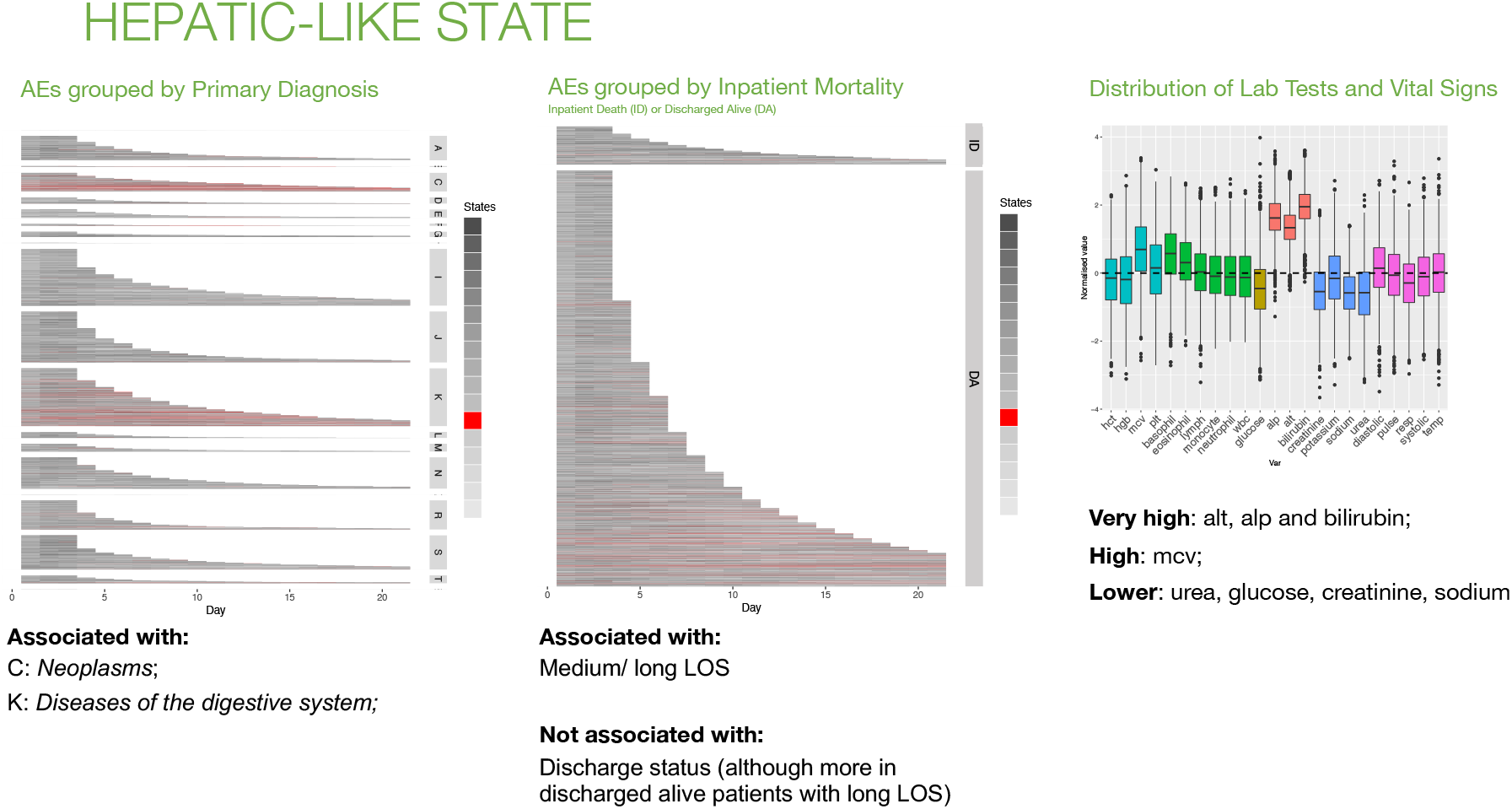
Hepatic-Like state. Patients’ trajectories (up to 21 days) grouped by primary diagnosis at admission (left) and by outcome at discharge (centre). Distribution of laboratory test results and vital signs for days represented as this state by the HMM model (right). AE: admission episode; ID: inpatient death; DA: discharged alive; LOS: length of stay.

**Table 1.** Summary of states and their most relevant features

|  | State | Key features | Key associations |
| --- | --- | --- | --- |
| Disease-like states |  |  |  |
| o | Hepatic | Abnormal liver function tests | Primary diagnosis: digestive system (K) & neoplasms (C) |
| j | Stable renal | Abnormal renal function tests | Primary diagnosis: genitourinary system (N) |
| a | Unstable renal | Abnormal renal function tests | Primary diagnosis: genitourinary system (N)<br>Clinical outcome: inpatient death |
| l | Stable but static renal | Abnormal renal function tests | Clinical outcome: Discharge alive |
| i | Blood dyscrasia | Abnormal full blood count (wide variance for most values) | Primary diagnosis: neoplasms (C) & diseases of the blood (D) |
| g | Bone marrow suppression | Abnormal full blood count (low values) | Primary diagnosis: neoplasms (C) & diseases of the blood (D) |
| Admission-like states |  |  |  |
| h | Acute presentation | High WBCs, Haemoglobin, Haematocrit and abnormal vital signs | Stage of Admission: Day 1 |
| e | Treatment response (1) | All values nearer group mean compared to Acute Presentation-like state | Stage of Admission: Day 2 to Day 8 (or discharge) |
| d | Treatment response (2) | Low values of some WBCs with other values around group mean | Stage of Admission: Day 2 to Day 8 (or discharge) |
| m | Early discharge | All values near group mean | Stage of Admission: last days of short admission episodes |
| n | Pre-discharge | All values near group mean except creatinine (lower), urea (lower) and platelets (higher) | Stage of admission: last days of long admission episodes |
| Physiological-like states |  |  |  |
| b | Early inflammatory response | Markedly high WBCs, urea, and abnormal vital signs | Clinical outcome: inpatient death |
| k | Resolving inflammatory response | High WBCs but vital signs nearer to group mean | Clinical outcome: long length of stay |
| p | Autoimmune/ atopic | High basophils, eosinophils and lymphocytes | Clinical outcome: Weakly associated with discharge alive |
| q | Acute thrombotic | High haemoglobin, haematocrit and lymphocytes | Clinical outcome: short length of stay and discharge alive<br>Primary diagnosis: Diseases of the circulatory system (I) and symptoms/ signs/ clinical findings (R) |
| c | Prolonged illness | Most values abnormal but especially respiratory rate (high) and haemoglobin (low) | Admission stage: more common after Day 4 |
| f | Other illness presentation | All parameters at or near group mean | Admission stage: More common in first 3-4 days |

*‘Disease-like’* states are characterised by over-representation of the state in patients sharing a common primary admission diagnosis and/ or a pattern of abnormality in the laboratory blood test and vital signs information that reflects dysfunction in a particular organ (either liver, kidney or bone marrow). This is exemplified by *‘Hepatic-like’* state in Figure 5. This state is defined by abnormalities in liver function tests and is over-represented in AEs with an admission diagnosis coded as ‘Neoplasms’ or ‘Diseases of The Digestive System’. Many neoplasms involve the liver as a common site of metastasis and the liver is an organ of the digestive system.

The predominant feature for *‘Admission-like’* states is over-representation of the state at a particular stage of the AE, either at the beginning (Day 1), middle (Day 2 onwards) or in the days leading up to discharge or death. Unlike *‘Disease-like’* states, these states are not overrepresented in a particular diagnostic code (Supplementary Figure 5). However, they have distinct patterns with respect to the distributions of laboratory and physiological variables, and the clinical interpretation of these is consistent with the temporal distribution of the state within the AE. For example, *‘Acute Presentation-like’* state is associated with Day 1 (Figure 6) and values are higher than average for haemoglobin, haematocrit, total WBCs, and neutrophils, and all vital signs are abnormal. This pattern is consistent with common conditions causing older patients to access emergency inpatient treatment, such as acute infections or acute coronary syndromes. For example, high WBCs are associated with pneumonia and urinary tract infections, and a high haematocrit is associated with cardiovascular disease and related risk factors [26–28]. Abnormal vital signs are also consistent with higher illness acuity at the point of emergency hospital admission before medical intervention has occurred. In contrast, *‘Early Discharge-like’* state (Supplementary Figure 5) shows a remarkably ‘normal’ distribution of laboratory test and vital sign values. This would be consistent with good illness recovery and this state is common on Day 2 of the admission onwards, particularly in short AEs for patients discharged alive.

**Figure 6.**
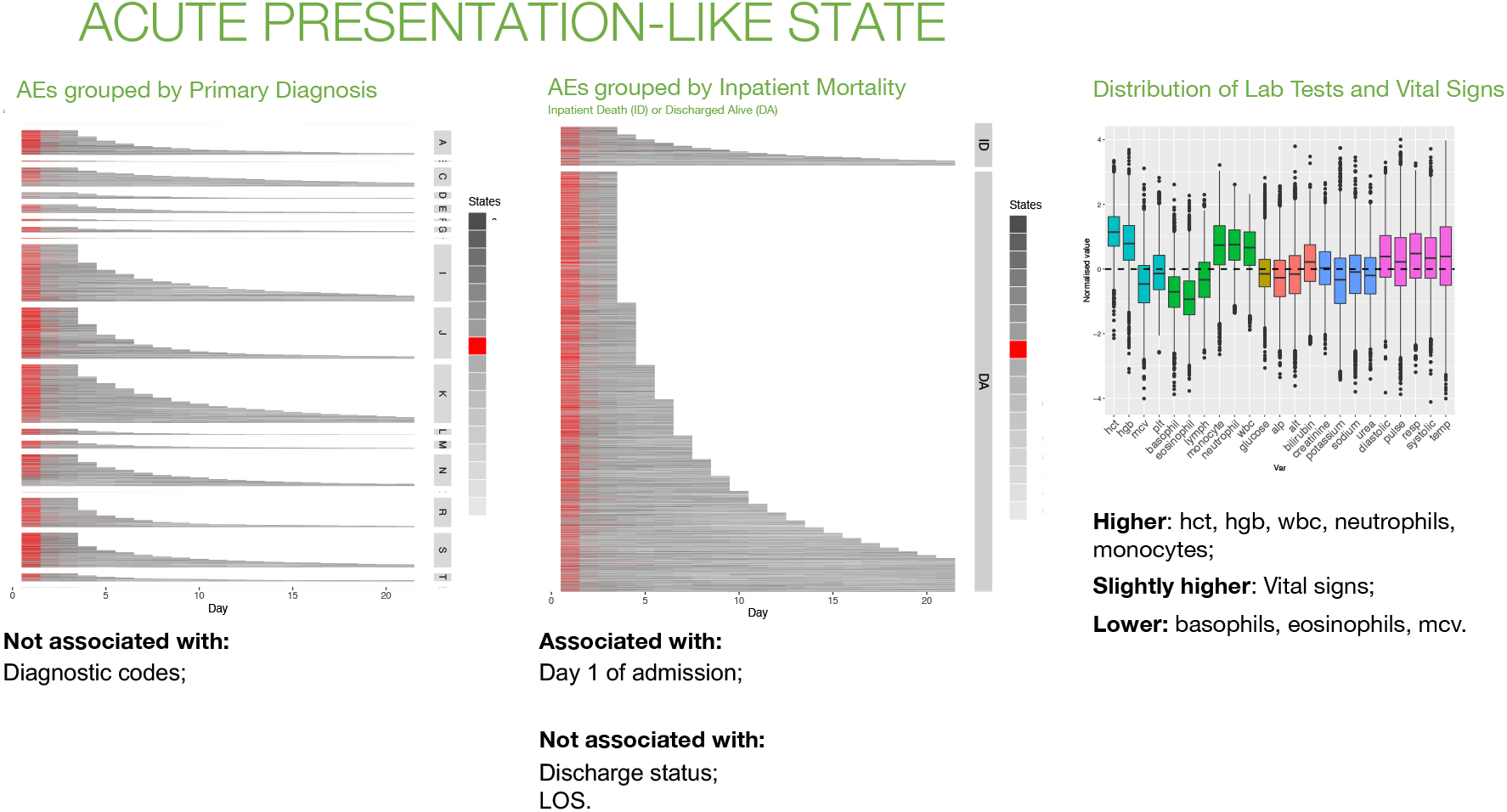
Acute Presentation-like state. Patients’ trajectories (up to 21 days) grouped by primary diagnosis at admission (left) and by outcome at discharge (centre). Distribution of laboratory test results and vital signs for days represented as this state by the HMM model (right). AE: admission episode; ID: inpatient death; DA: discharged alive; LOS: length of stay.

*‘Physiological-like’* states are characterised by patterns of ‘abnormality’ in laboratory and vital sign values. Clinicians might recognise these states as clinical syndromes or the final common pathway of a physiological response to a range of insults e.g., autoimmune or inflammatory responses. For example, *‘Early Inflammatory Response-like’* state is characterised by high respiratory rate, heart rate, WBCs, neutrophils and urea and low blood pressure (Figure 7). This is similar to the clinical description of the Systemic Inflammatory Response Syndrome (SIRS) [29]. In general, these states are present in several diagnostic codes but not strongly over-represented in any one ICD-10 code (Supplementary Figure 5).

**Figure 7.**
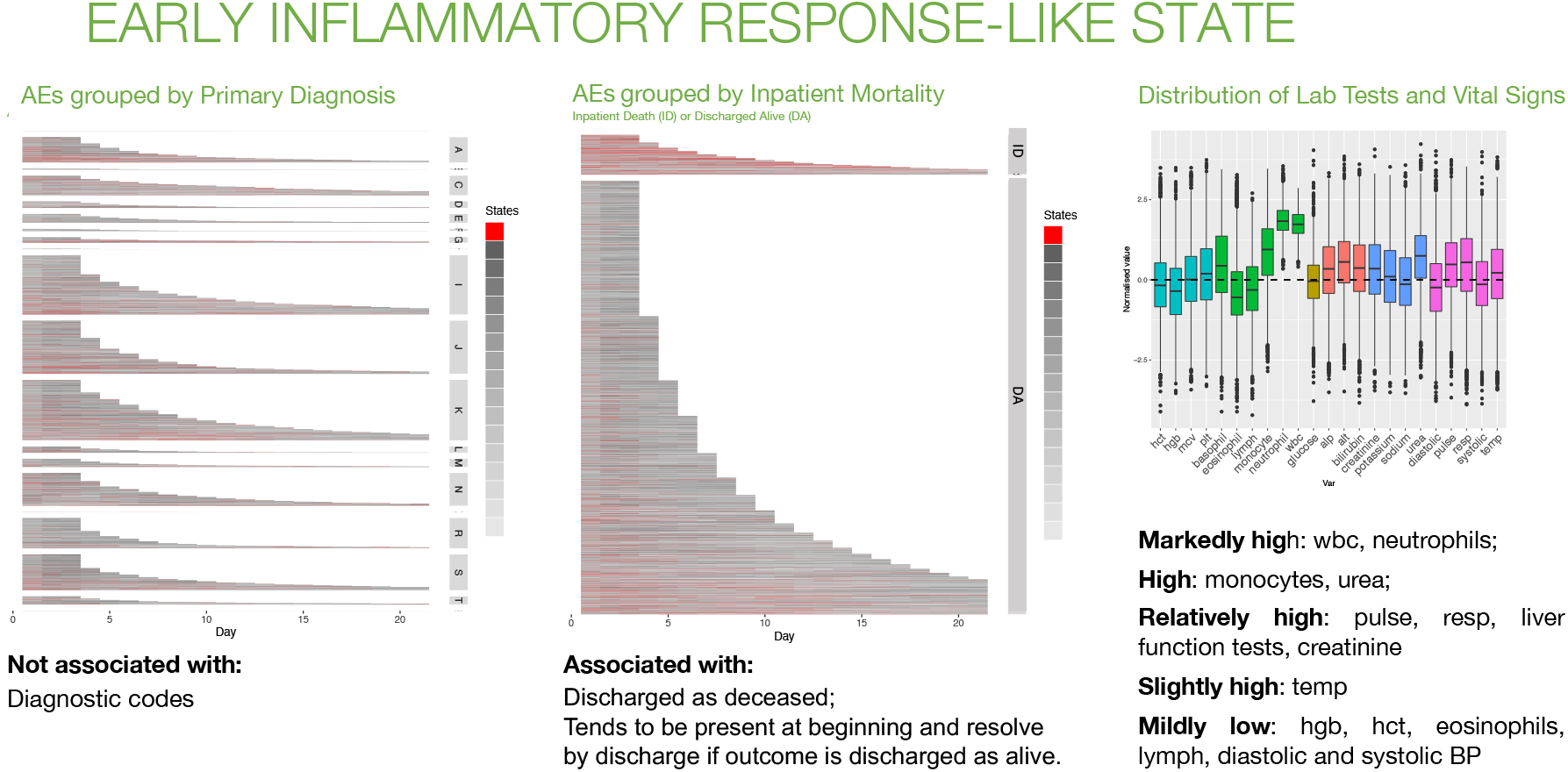
Early Inflammatory Response-like state. Patients trajectories (up to 21 days) grouped by primary diagnosis at admission (left) and by outcome at discharge (centre). Distribution of laboratory test results and vital signs for days represented as this state by the HMM model (right). AE: admission episode; ID: inpatient death; DA: discharged alive; LOS: length of stay.

Not all 17 states could be classified confidently. For example, some states such as *‘Stable but Static Renal-like’* state and *‘Other Illness Presentation-like’* state did not have a clear ‘predominant feature’ (Supplementary Table 3).

The algorithm was trained again in the ‘training and test’ dataset and the resulting model used to assess the ‘hold-out validation’ dataset. The distributions of the laboratory test result and vital signs in both outputs showed similar patterns and allowed the mapping of all states with the exception of *‘Other Illness Presentation-like’* state, for which some variables did not match as well as the for the other states (Supplementary Figure 6). Figure 8 shows the correlation between the means of each variable in the training and validation datasets with a Pearson correlation coefficient of 0.94 and outliers corresponding to variables of this state. HMM are stochastic algorithms and, as such, every time they are trained the resulting model, and subsequently the outputs, are different. The presence of a poorly defined state with heterogeneous patient characteristics and more variance in variable characteristics is expected given the HMM modelling framework, which has to assign a single state to every time point and patient with a limited set of states. Similar to other uses of HMM models, we should expect one state to be less defined and more heterogeneous Patients were assigned to a ‘main state’ if they spent more than half of their time in the same state. The percentages of patients classified in this way were 80% and 86% in the ‘training and test’ and ‘hold-out validation’ datasets, respectively. Formal statistical tests were used to evaluate associations of each state with sex, age-group, CFS category, month of admission, admission diagnosis, discharge specialty, length of stay, number of diagnoses at discharge, and hospital outcomes (IM, PDR, PDM), none of which were variables used to train the HMM. Some trends could be predicted by the initial visual clinical interpretation of the states and supported the expert interpretation. For example, *‘Disease-like’* states showed the strongest associations with admission ICD-10 codes and overall, 12 States (71%) showed at least one association with an ICD-10 code (p-value <0.001 with Bonferroni correction) and a similar pattern of associations was also observed in the ‘hold-out validation’ dataset (Figure 9). *‘Early Inflammatory Response-like’* state was also strongly associated with inpatient and post-discharge mortality (Supplementary Figure 7C). These associations supported the clinical visual interpretation, and it is important to remember that the HMM was trained without any information on ICD-10 codes or outcome data.

**Figure 8.**
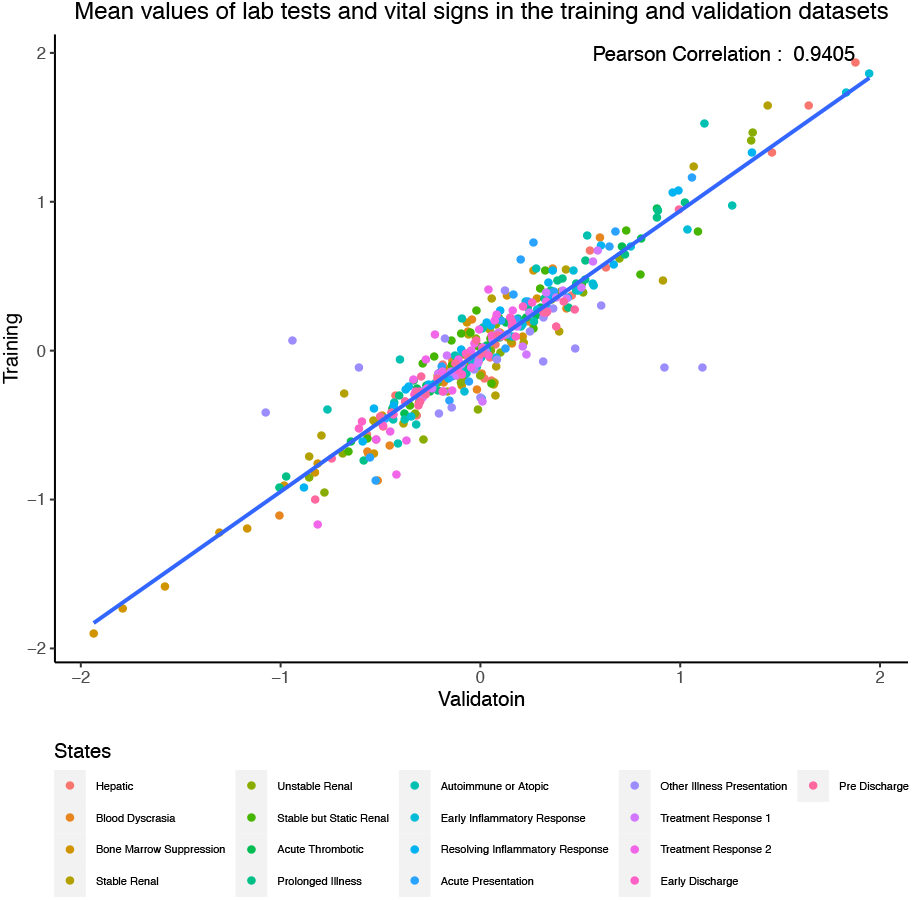
Correlation between mean values of laboratory test results and vital signs variables in the ‘training and test’ and ‘hold-out validation’ datasets.

**Figure 9.**
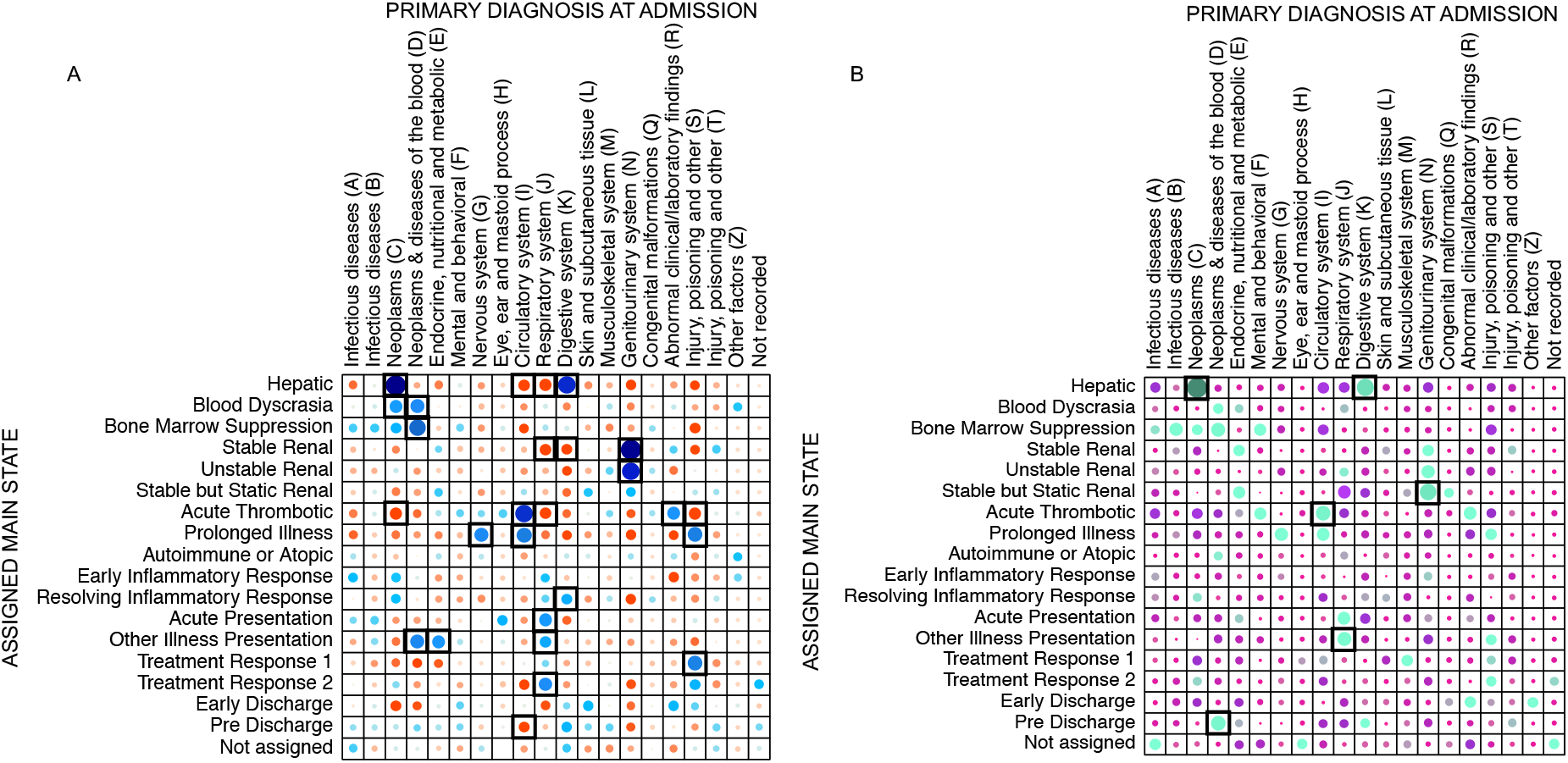
Pearson residuals plots showing associations between patients’ assigned main states and primary diagnosis at admission. A) ‘Training and test’ dataset; B) ‘Hold-out validation’ dataset. Blue and green: positive associations; red and purple: negative associations. Black boxes: statistically significant associations (p-value <0.001 with Bonferroni correction)

Other trends added to clinical interpretations. *‘Prolonged Illness-like’* state generally occurred 4-5 days into an AE and was characterised by low haemoglobin and haematocrit whilst all other parameters were high, especially respiratory rate (Supplementary Figures 5). One hypothesis, after visual clinical interpretation, was that an intervention that influenced the state had occurred during the AE. Consistent with this, *‘Prolonged Illness-like’* state was strongly associated with discharge by a surgical specialty (Supplementary Figure 7A).

The *‘Disease-like’* states also showed strong positive associations with post discharge readmission, particularly *‘Bone Marrow Suppression-like’* and *‘Stable but Static Renal-like’* states, perhaps reflecting the role of chronic disease in increasing vulnerability to hospitalisation. However, associations did not always add to the clinicians’ understanding of the HMM state. For example, *‘Treatment Response-like 1 and 2’* states remained difficult to fully differentiate in clinical terms.

### 3.3. Results from prediction models focusing on first days of admission

We used straightforward discriminative models on this dataset and enumerated a relatively large number of hypotheses (16 × 6 = 96), defined as combinations of: a) prediction model (Random Forests, RF or Logistic Regression, LR), b) input data representation (MVTS or HMM states), c) inclusion or not of phenotypic information (i.e., primary diagnosis at admission), d) different input days lengths, and e) different predicted clinical outcomes (Supplementary Table 4). We registered these hypotheses before running tests at the Open Science Framework. For each outcome, the model with the highest performance by ‘group’ (defined as the combination of model, input data representation and phenotypic information) was identified. We refer to these several combinations as *‘models’* for simplicity. The selected parameters of the best models (based on ROC-AUC or weighted ROC-AUC performance) are summarized in Supplementary Table 5. The trained models were assessed with the ‘hold-out validation’ dataset. For simplicity, in this paper we describe only the performance achieved by the best models as identified in the ‘training and test’ dataset. Detailed results of performance metrics in both datasets are reported in Supplementary Tables (6-15).

Although similar, higher performance on ROC-AUC and weighted ROC-AUC was observed overall for models trained with MVTS compared to states representations (Table 2). Also, similar performances were observed by group, especially in the models trained with states data. Almost all models showed highest performance for all metrics when trained with either Day 3 (D3) or Days 1 to 3 (D1D2D3) input variables and the inclusion of phenotypic information modestly improved the performance of models trained to predict inpatient mortality and clinical outcome at 30-day of admission.

**Table 2.** ROC-AUC (and weighted ROC-AUC) results for the models with highest performance on the prediction of inpatient mortality, 30-day clinical outcome, diagnosis at admission and diagnoses at discharge.

| Predicted Outcome | Data Representation | Algorithm | Input data |  | ROC-AUC |  |
| --- | --- | --- | --- | --- | --- | --- |
|  |  |  | Days | Phenotypic information | Training dataset | Validation dataset |
| Inpatient Mortality | MVTs | RF | D3 | YES | 0.85 | 0.86 |
|  | STATES | RF | D3 | YES | 0.79 | 0.63 |
| Clinical outcome | MVTs | RF | D1D2D3 | YES | 0.68 | 0.68 |
|  | STATES | RF | D3 | YES | 0.67 | 0.58 |
| Diagnosis at admission | MVTs | RF | D1D2D3 | - | 0.76 | 0.75 |
|  | STATES | RF | D1D2D3 | - | 0.65 | 0.54 |
| Diagnosis at discharge | MVTs | RF | D1D2D3 | - | 0.65 | 0.75 |
|  | STATES | RF | D2D3/D1D2D3* | - | 0.6 | 0.54 |

RF: Random Forest; MVTs: Multivariate time series; \* Same performance with both input datasets

Prediction of **inpatient mortality** was assessed comparing the performance of RF and LR models (Supplementary Tables 6-8). While only the RF trained with MVTS data (with or without phenotypic information) achieved reasonable precision compared to other models (0.72 and 0.62, respectively), recall results were very low (highest 0.2). The opposite was observed with the RF models trained with HMM states (highest precision and recall 0.2 and 0.7, respectively). Recall obtained by LR models was higher than by RF models, which explains that the highest F1-score was achieved with a LR model with MVTS data (Supplementary Table 8). Consistent results were observed in the hold-out validation data. However, the low recall of the best model in the test dataset (only 14 patients predicted at higher risk of inpatient death in five or more runs out of 226 real cases) limits the analysis of these results.

Feature ranking for the best RF model (based on ROC-AUC performance) showed that the top five features were respiratory rate, eosinophil count, urea, lymphocytes and neutrophils. These features closely resemble those characterising the *‘Early Inflammatory Response-like’* state (high WBC, neutrophils, urea, respiratory rate, and heart rate; Figure 7), which emerged from the HMM and was associated with inpatient mortality in both the visual clinical interpretation and statistical analysis of HMM state associations (Supplementary Figure 7C).

Prediction of **clinical outcome at 30 days** was assessed using the one-vs-rest classifier approach, where all tasks are binary classification problems with five disjoint classes: ID, DA, PDR, PDRM and PDM. Similar weighted ROC-AUC performance was achieved by the four best models with the highest obtained by the RF model trained with MVTS on D1D2D3 and phenotypic information (Table 2). Weighted recall and precision were also very similar for all the models (Supplementary Table 10). As shown in Supplementary Table 11, the averaged results are mainly driven by the performance achieved on the prediction of the majority class, DA. Results for the class ID did not differ considerably from those observed with the RF model in the binary classification of this outcome, apart from precision achieved with MVTS (0.6 with the best binary RF model vs 0.4 with the best multiclass model). The prediction performance for other classes was quite limited, most likely due to small numbers of outcome events (especially in the case of PDRM and PDM).

Feature ranking for the best performing models showed that highest ranking features differed by outcome class. Features driving prediction of hospital readmission (PDR) differed from the ones driving prediction of mortality. Examination of the best performing model employing the MVTS representation revealed PDR prediction was driven by data on Day 1 rather than either Day 3 or Days 1,2 and 3. When the best performing model employing the HMM states representation of physiological variables was examined, demographics and diagnosis codes were the main drivers of PDR prediction (age, sex, frailty score and ICD-10 code ‘S’).

The prediction of **primary diagnosis at admission** (PDA) differs for the different diagnosis codes. An analysis of F1-score by class showed that prediction performance was usually higher for diagnosis codes with more instances in the dataset, ICD-10 codes J, K, N, I, S, C, with the exception of D (good performance with small number of instances) and A (worse performance than other codes with similar numbers). Prediction of **diagnosis at discharge** (DD) was defined as a multilabel classification problem because one patient can be predicted to have several diagnosis codes. Similar to prediction of PDA, precision, recall and F1-score (Supplementary Figures 8-9) was better for instances with more examples in the dataset (e.g., ICD-10 codes I or N).

## 4. DISCUSSION AND CONCLUSIONS

We have shown a practical way to provide a dataset from an NHS hospital EHR, which is appropriate for ML using the well-established generative time-dependent HMM framework and discriminative RF and LR models. Important aspects of our work were development of the governance structure to facilitate analysis of routinely collected hospital EHR data by technical experts, and development of a protocol to regularise and impute this data into research ready components. We incorporated sufficient anonymisation processes to safeguard privacy, whilst leaving enough information for hidden relationships within the data to be explored and employed secure mechanisms for data transfer and access. We believe it will be critical to store these specific datasets as they are generated, as well as protocols for specific transformations as defined outputs, to progress this field and generate both reproducible research and share expertise.

All modelling requires assumptions and decisions about a variety of topics before the application of methods, including: what data to collect? what regularisation in time and distribution to use? how should missing data be handled? We have provided one approach addressing some of these issues and found the most important aspects of data preprocessing were regularisation and imputation of missing values. Daily bins across time series records of ≥3 days were selected because of our interest in blood biochemistry as input variables, which were recorded once per day for most tests and patients. Regularisation of time series is a necessary step for many ML models but implies loss of information for those tests with multiple recorded observations on a pre-established bin and the introduction of missing values where there are no recorded observations. In future, we will explore different ways of feeding or representing this information to analyse its impact on inpatient representations or outcome prediction [30].

Our approach towards imputation was a two-step process. MI was the preferred method because it takes advantage of relationships between variables in the dataset [31] but this method cannot be used on days with many missing variables. In that case, LI was used at the individual time series level. To reduce the impact of imputed values on our results, strict inclusion criteria were used to select the final cohort of patients, aimed to limit missingness and avoid imputation of large numbers of consecutive missing observations. Imputation evaluation conducted at different stages found no evidence that the distributions of imputed values were different from original observations. Importantly, we showed that the main characteristics driving the HMM states were not influenced by imputed observations. We anticipate our methods can be adapted, replicated and probably improved by others, since we utilised previously validated and commonly used imputation techniques. Future work, with less restrictive inclusion criteria or shorter time-bins might benefit from other approaches, such as deep learning [32] or alternatives to imputation [14].

The representation of how a patient’s condition evolves over time using ML has been predominantly studied from the perspective of temporal disease trajectories through consecutive clinical encounters [19,33,34]. These representations are derived mainly from clinical diagnosis codes, which can be merged with other categorical information such as medications, demographics or non-numeric descriptions of laboratory test results [8,18]. In this work we focus on a narrower time-window, hospital admissions in a general inpatient population, and use laboratory blood tests and vital signs to form a numeric representation of patients’ physiological status and illness acuity over time. We have explored this to discover trends on inpatients’ trajectories that are not necessarily linked to a previously specified outcome.

We were surprised by the ability of the HMM to provide an informative view of inpatient trajectories. The initial analysis of the distribution of states showed remarkable differences between patients who died at the end of the AE and those who were discharged. The expert clinician provided a visual clinical interpretation for each of the 17 HMM state spaces, with most states naturally coalescing into one of three higher level classifications, which made biological and clinical sense. The conceptual ‘state’ of a hospitalised patient at any moment in time will depend on their disease burden (represented by *‘Disease-like’* states), their physiological response to this burden (represented by *‘Physiological-like’* states) and their response to medical intervention (represented by *‘Admission-like’* states). There were also strikingly different state distributions in patients who died during the AE versus those discharged alive and *‘Early Inflammatory Response-like’* state was one of two states strongly over-represented in those who died. This state was interpreted by the clinician as resembling SIRS, a clinical syndrome often precipitated by infection and associated with high mortality in hospitalised patients [29,35,36]. Furthermore, significant numbers of states were associated either with specific diagnosis codes or stages of the AE. These findings are all made with no explicit training data for outcomes or diagnoses, but rather emerge from the generative model of the HMM and strongly suggest the HMM is capturing real patient biology. We acknowledge that the HMM model which insists on a single state being assigned to a patient day here is a crude approximation to clinical reality, but nevertheless reassured that even with this constraint the resulting HMM was a useful model to explore clinical trajectories.

Little emphasis has been placed on simply exploring data extracted from hospital EHR systems. Instead, this data is used to develop prediction models for pre-specified outcomes [8]. Whilst this focus is understandable, given its potential clinical utility, information is lost pertaining to the variability of what happens to patients after emergency hospitalisation. This may help inform modelling. Although not the goal for this analysis, our exploration and description of inpatient trajectories using generative ML prompts questions about exploring better decision support tools or characterisation of patients. Notably, *‘Acute Presentation-like’* state is almost uniquely observed on the first day of admission and shows the well understood need to stabilise patients presenting with acutely deranged physiological status. However, the state on Day 1 of admission showed similar distributions in those discharged alive versus dead. In contrast, patients transitioned to a greater range of states from Day 2 onwards, and there were increasingly different state space distributions as length of stay progressed depending on the final hospital outcome. This highlights how careful modelling of daily, or finer grain, vital signs and laboratory blood tests can be used to better inform clinicians of patient risk across the entirety of an AE. Current decision support tools aimed at identifying the deteriorating patient [37] have not usually been developed considering temporal trends in physiological status and employ simple risk factor categorisations for scoring at the bedside [38,39]. The implementation of hospital EHRs offers an opportunity to develop new tools that can both utilise continuous risk estimates [40] and consider the temporal sequence of data [9], perhaps focusing particularly on Days 2 and 3 of admission as patients’ clinical trajectories diverge.

The discriminative learning models also performed reasonably well. Higher prediction was achieved when the MVTS data was used as input data compared to the HMM states representations, which likely reflects the reduction of input data from a multivariate to univariate time series in the latter case. However, models employing states representations still performed comparably well and there were interesting similarities between generative and discriminative ML outputs that made clinical sense. Both approaches identified states and features representative of a strong inflammatory response as important for inpatient mortality. Additionally, both generative and discriminative modelling suggested factors more indicative of chronic problems such as frailty, diagnosis codes, the *‘Disease-like’* HMM states and vital sign and laboratory test values on Day 1 rather than Day 3 of admission, may be important for risk of hospital readmission. It is notable that ICD-10 code ‘S’ was an important feature driving prediction of PDR in the best performing model employing the states representation of MVTS data. This code includes ‘injuries’ and will capture older patients hospitalised due to the consequences of a fall e.g., hip fracture. Falls and associated injuries are key consequences of frailty, place significant burden on the NHS [41] and are associated with high readmission rates [42]. Thus, although the limited performance achieved by the prediction models prevents establishing associations, these or future analyses could generate hypothesis for further investigation.

Our dataset is extracted from a single NHS hospital and our final patient cohort was subject to strict inclusion criteria meaning our results are not generalisable to all inpatients. Restricting our cohort to patients with an inpatient stay of ≥3 days excluded the most acutely unwell and fittest patients, truncating the range of physiological and metabolic variation and perhaps excluding those whose outcome was easiest to predict. MVTS data were limited to commonly measured variables and we were limited to broad categorisation of some features, such as the highest level of ICD-10 categorisation. Considering our modelling, as noted above, the HMM has strong and unrealistic assumptions and the clinical interpretations and classifications provided require careful consideration. Our prediction work was also limited to exploratory analyses and would benefit from inclusion of additional patient features and use of ML techniques that account for the sequential nature of data. Lastly, although the discriminative learning methods performed reasonably well, similar to other EHR schemes, it is important to realise the relatively poor precision. This should be expected since the goal of healthcare is to change discharge status via a variety of means, but it also emphasises the difference between providing useful summaries of patient data consistent with previous knowledge for decision support, compared to making a hard ‘call’ of outcome.

Nevertheless, we generated a research ready dataset using hospital EHR data that was appropriate for both generative and discriminative ML techniques. The clinical interpretation of the HMM states was novel, with previous attempts limited to interpretation of states as disease stages [19,43,44], and promoted knowledge sharing between clinicians and data scientists. Additionally, the HMM appeared to capture real biological signal. Not only were all 17 states interpreted by the clinician and associated with hold-out clinical information, but generative and discriminative models converged on outputs which made clinical sense, such as associations between inflammation and mortality. Both approaches appeared complementary, discriminative analyses using the raw MVTS variables achieved higher prediction and generative modelling facilitated clinical interpretation, and both can provide a landscape for hypothesis generation. In future we intend to pursue more focused questions, for example capitalising on our characterisation of inpatient trajectories to explore finer grain modelling in the first three days of admission and exploring ML techniques that consider the sequential nature of data. We welcome collaborations from other ML experts, health data scientists and EHR facilities to explore this datasets and others, following governance and ethical review.

## Data Availability

The anonymised data that support this research project are available from Cambridge University Hospitals NHS Foundation Trust. Restrictions apply to the availability of these data, which were used under license for the current study, and so are not publicly available. Reasonable data requests will be considered by the authors with permission of Cambridge University Hospitals NHS Foundation Trust.

## Authors’ contributions

MHZ, VLK, TF and EB designed the research; MHZ performed the data processing and experimental analysis; VLK performed the clinical interpretation of the results; MHZ, VLK, TF and EB wrote the manuscript; AC and VT provided the EHR data; HS advised on data governance and anonymization and gained ethics approval; EB and JB supervised and supported the research. All authors have read and approved the final manuscript.

## Acknowledgements & Funding

This research was supported by the NIHR Cambridge Biomedical Research Centre (BRC-1215-20014). The views expressed are those of the authors and not necessarily those of the NIHR or the Department of Health and Social Care. VLK was funded by a MRC/NIHR Clinical Academic Research Partnership Grant (CARP; grant code: MR/T023902/1). VT is supported by Cancer Research UK. EB and TF were funded by the EMBL European Bioinformatics Institute (EMBL-EBI).

## Consent and Ethics

Informed consent was not required for this study since the routinely collected healthcare data presented were anonymised. The project was approved by the NHS Health Research Authority (HRA) (IRAS: 253457), North East – Newcastle & North Tyneside 1 Research Ethics Committee (REC) (REC reference: 19/NE/0013) and by the EMBL Scientific Advisory Committee (BIAC).

## Software availability

The R and Python code associated with the data pre-processing, imputation and model development is available https://github.com/mariaheza/hmm_inpatient_trajectories.

## References

1. Evans, T.W. Best research for best health: a new national health research strategy. Clin. Med. (Northfield. II). 2006, 6, 435–437, doi:10.7861/clinmedicine.6-5-435.

2. Department of Health and Social Care The future of healthcare: our vision for digital, data and technology in health and care. Dep. Heal. Soc. Care Gov.UK 2018, 1–38.

3. Herrett, E.; Gallagher, A.M.; Bhaskaran, K.; Forbes, H.; Mathur, R.; van Staa, T.; Smeeth, L. Data Resource Profile: Clinical Practice Research Datalink (CPRD). Int. J. Epidemiol. 2015, 44, 827–836, doi:10.1093/ije/dyv098.

4. Denaxas, S.; Gonzalez-Izquierdo, A.; Direk, K.; Fitzpatrick, N.K.; Fatemifar, G.; Banerjee, A.; Dobson, R.J.B.; Howe, L.J.; Kuan, V.; Lumbers, R.T.; et al. UK phenomics platform for developing and validating electronic health record phenotypes: CALIBER. J. Am. Med. Informatics Assoc. 2019, 26, 1545–1559, doi:10.1093/jamia/ocz105.

5. Herbert, A.; Wijlaars, L.; Zylbersztejn, A.; Cromwell, D.; Hardelid, P. Data Resource Profile: Hospital Episode Statistics Admitted Patient Care (HES APC). Int. J. Epidemiol. 2017, 46, 1093–1093i, doi:10.1093/ije/dyx015.

6. Lee, C.K.; Hofer, I.; Gabel, E.; Baldi, P.; Cannesson, M. Development and Validation of a Deep Neural Network Model for Prediction of Postoperative In-hospital Mortality. Anesthesiology 2018, 129, 649–662, doi:10.1097/ALN.0000000000002186.

7. Chilamkurthy, S.; Ghosh, R.; Tanamala, S.; Biviji, M.; Campeau, N.G.; Venugopal, V.K.; Mahajan, V.; Rao, P.; Warier, P. Deep learning algorithms for detection of critical findings in head CT scans: a retrospective study. Lancet 2018, 392, 2388–2396, doi:10.1016/S0140-6736(18)31645-3.

8. Miotto, R.; Li, L.; Kidd, B.A.; Dudley, J.T. Deep Patient: An Unsupervised Representation to Predict the Future of Patients from the Electronic Health Records. Sci. Rep. 2016, 6, 26094, doi:10.1038/srep26094.

9. Rajkomar, A.; Oren, E.; Chen, K.; Dai, A.M.; Hajaj, N.; Liu, P.J.; Liu, X.; Sun, M.; Sundberg, P.; Yee, H.; et al. Scalable and accurate deep learning for electronic health records. npj Digit. Med. 2018, 1–10, doi:10.1038/s41746-018-0029-1.

10. Rajkomar, A.; Dean, J.; Kohane, I. Machine Learning in Medicine. N. Engl. J. Med. 2019, 380, 1347–1358, doi:10.1056/NEJMra1814259.

11. Hemingway, H.; Asselbergs, F.W.; Danesh, J.; Dobson, R.; Maniadakis, N.; Maggioni, A.; van Thiel, G.J.M.; Cronin, M.; Brobert, G.; Vardas, P.; et al. Big data from electronic health records for early and late translational cardiovascular research: challenges and potential. Eur. Heart J. 2018, 39, 1481–1495, doi:10.1093/eurheartj/ehx487.

12. Shickel, B.; Tighe, P.J.; Bihorac, A.; Rashidi, P. Deep EHR: A Survey of Recent Advances in Deep Learning Techniques for Electronic Health Record (EHR) Analysis. IEEE J. Biomed. Heal. Informatics 2018, 22, 1589–1604, doi:10.1109/JBHI.2017.2767063.

13. Purushotham, S.; Meng, C.; Che, Z.; Liu, Y. Benchmarking deep learning models on large healthcare datasets. J. Biomed. Inform. 2018, 83, 112–134, doi:10.1016/j.jbi.2018.04.007.

14. Bianchi, F.M.; Livi, L.; Mikalsen, K.Ø.; Kampffmeyer, M.; Jenssen, R. Learning representations for multivariate time series with missing data using Temporal Kernelized Autoencoders. arXiv Prepr. 1805.0347 2018, 1–18.

15. Zhao, J.; Papapetrou, P.; Asker, L.; Boström, H. Learning from heterogeneous temporal data in electronic health records. J. Biomed. Inform. 2017, 65, 105–119, doi:10.1016/j.jbi.2016.11.006.

16. Harutyunyan, H.; Khachatrian, H.; Kale, D.C.; Ver Steeg, G.; Galstyan, A. Multitask learning and benchmarking with clinical time series data. Sci. Data 2019, 6, 96, doi:10.1038/s41597-019-0103-9.

17. Johnson, A.E.W.; Pollard, T.J.; Shen, L.; Lehman, L.H.; Feng, M.; Ghassemi, M.; Moody, B.; Szolovits, P.; Anthony Celi, L.; Mark, R.G. MIMIC-III, a freely accessible critical care database. Sci. Data 2016, 3, 160035, doi:10.1038/sdata.2016.35.

18. Ruan, T.; Lei, L.; Zhou, Y.; Zhai, J.; Zhang, L.; He, P.; Gao, J. Representation learning for clinical time series prediction tasks in electronic health records. BMC Med. Inform. Decis. Mak. 2019, 19, 1–14, doi:10.1186/s12911-019-0985-7.

19. Pham, T.; Tran, T.; Phung, D.; Venkatesh, S. Predicting healthcare trajectories from medical records: A deep learning approach. J. Biomed. Inform. 2017, 69, 218–229, doi:10.1016/j.jbi.2017.04.001.

20. Lipton, Z.C.; Kale, D.C.; Elkan, C.; Wetzel, R. Learning to Diagnose with LSTM Recurrent Neural Networks. 4th Int. Conf. Learn. Represent. ICLR 2016 - Conf. Track Proc. 2016, 1–18.

21. Choi, E.; Bahadori, M.T.; Schuetz, A.; Stewart, W.F.; Sun, J. Doctor AI: Predicting Clinical Events via Recurrent Neural Networks. Mach. Learn. Healthc. Conf. 2015, 301–318.

22. Imison, C.; Poteliakhoff, E.; Thompson, J. Older people and emergency bed use. Exploring variation. Ideas that Chang. Heal. care 2012, 1–24.

23. Rockwood, K.; Song, X.; MacKnight, C.; Bergman, H.; Hogan, D.B.; McDowell, I.; Mitnitski, A. A global clinical measure of fitness and frailty in elderly people. CMAJ 2005, 173, 489–495, doi:10.1503/cmaj.050051.

24. Sterne, J.A.C.; White, I.R.; Carlin, J.B.; Spratt, M.; Royston, P.; Kenward, M.G.; Wood, A.M.; Carpenter, J.R. Multiple imputation for missing data in epidemiological and clinical research: potential and pitfalls. BMJ 2009, 338, b2393–b2393, doi:10.1136/bmj.b2393.

25. Zhang, J.; Chen, D. Interpolation calculation made EZ. In Proceedings of the 4th Annual Conference Proceedings, NorthEast SAS Users Group NESUG; Baltimore, MD, 2001.

26. Wittenberg, R.; Sharpin, L.; McCormick, B.; Hurst, J. Understanding emergency hospital admissions of older people; Centre for Health Service Economics and Organisation, Oxford, UK, 2017;

27. Jin, Y.-Z.; Zheng, D.-H.; Duan, Z.-Y.; Lin, Y.-Z.; Zhang, X.-Y.; Wang, J.-R.; Han, S.; Wang, G.-F.; Zhang, Y.-J. Relationship Between Hematocrit Level and Cardiovascular Risk Factors in a Community-Based Population. J. Clin. Lab. Anal. 2015, 29, 289–93, doi:10.1002/jcla.21767.

28. Danesh, J. Haematocrit, viscosity, erythrocyte sedimentation rate: meta-analyses of prospective studies of coronary heart disease. Eur. Heart J. 2000, 21, 515–520, doi:10.1053/euhj.1999.1699.

29. Bone, R.C.; Balk, R.A.; Cerra, F.B.; Dellinger, R.P.; Fein, A.M.; Knaus, W.A.; Schein, R.M.H.; Sibbald, W.J. Definitions for Sepsis and Organ Failure and Guidelines for the Use of Innovative Therapies in Sepsis. Chest 1992, 101, 1644–1655, doi:10.1378/chest.101.6.1644.

30. Che, Z.; Purushotham, S.; Cho, K.; Sontag, D.; Liu, Y. Recurrent Neural Networks for Multivariate Time Series with Missing Values. Sci. Rep. 2018, 8, 6085, doi:10.1038/s41598-018-24271-9.

31. White, I.R.; Royston, P.; Wood, A.M. Multiple imputation using chained equations: Issues and guidance for practice. Stat. Med. 2011, 30, 377–399, doi:10.1002/sim.4067.

32. Yoon, J.; Zame, W.R.; Van Der Schaar, M. Estimating Missing Data in Temporal Data Streams Using Multi-Directional Recurrent Neural Networks. IEEE Trans. Biomed. Eng. 2019, 66, 1477–1490, doi:10.1109/TBME.2018.2874712.

33. Jensen, P.B.; Jensen, L.J.; Brunak, S. Mining electronic health records: Towards better research applications and clinical care. Nat. Rev. Genet. 2012, 13, 395–405, doi:10.1038/nrg3208.

34. Beaulieu-Jones, B.K.; Orzechowski, P.; Moore, J.H. Mapping Patient Trajectories using Longitudinal Extraction and Deep Learning in the MIMIC-III Critical Care Database. In Proceedings of the Biocomputing 2018; WORLD SCIENTIFIC, 2018; Vol. 0, pp. 123–132.

35. Shapiro, N.; Howell, M.D.; Bates, D.W.; Angus, D.C.; Ngo, L.; Talmor, D. The Association of Sepsis Syndrome and Organ Dysfunction With Mortality in Emergency Department Patients With Suspected Infection. Ann. Emerg. Med. 2006, 48, 583-590.e1, doi:10.1016/j.annemergmed.2006.07.007.

36. Farrah, K.; McIntyre, L.; Doig, C.J.; Talarico, R.; Taljaard, M.; Krahn, M.; Fergusson, D.; Forster, A.J.; Coyle, D.; Thavorn, K. Sepsis-Associated Mortality, Resource Use, and Healthcare Costs: A Propensity-Matched Cohort Study*. Crit. Care Med. 2021, 49, 215–227, doi:10.1097/CCM.0000000000004777.

37. Royal College of Physicians National Early Warning Score (NEWS) 2: Standardising the assessment of acute-illness severity in the NHS. Updated report of a working party.; 2017;

38. Gerry, S.; Bonnici, T.; Birks, J.; Kirtley, S.; Virdee, P.S.; Watkinson, P.J.; Collins, G.S. Early warning scores for detecting deterioration in adult hospital patients: systematic review and critical appraisal of methodology. BMJ 2020, m1501, doi:10.1136/bmj.m1501.

39. Fang, A.H. Sen Lim, W.T.; Balakrishnan, T. Early warning score validation methodologies and performance metrics: a systematic review. BMC Med. Inform. Decis. Mak. 2020, 20, 111, doi:10.1186/s12911-020-01144-8.

40. Ghosh, E.; Eshelman, L.; Yang, L.; Carlson, E.; Lord, B. Early Deterioration Indicator: Data-driven approach to detecting deterioration in general ward. Resuscitation 2018, 122, 99–105, doi:10.1016/j.resuscitation.2017.10.026.

41. National Institute for Health and Care Excellence Falls in older people: assessing risk and prevention;2013;

42. Hoffman, G.J.; Liu, H.; Alexander, N.B.; Tinetti, M.; Braun, T.M.; Min, L.C. Posthospital Fall Injuries and 30-Day Readmissions in Adults 65 Years and Older. JAMA Netw. Open 2019, 2, e194276, doi:10.1001/jamanetworkopen.2019.4276.

43. Arandjelovic, O. Discovering hospital admission patterns using models learnt from electronic hospital records. Bioinformatics 2015, 31, btv508, doi:10.1093/bioinformatics/btv508.

44. Jackson, C.H.; Sharples, L.D.; Thompson, S.G.; Duffy, S.W.; Couto, E. Multistate Markov models for disease progression with classification error. J. R. Stat. Soc. Ser. D (The Stat. 2003, 52, 193–209, doi:10.1111/1467-9884.00351.

