## Supplementary material for "Big Data Analysis of Electronic Health Records: Clinically interpretable representations of older adult inpatient trajectories using time-series numerical data and Hidden Markov Models"

**Supplementary Table 1**: Variables retrieved from Epic system

The NHS number and local patient identifiers are replaced by an irreversible pseudonym that is consistent for each patient. The free text fields are scanned for the NHS number, local patient identifier and patient demographics and these are removed. Information extracted is outlined in the table below, including how the information is converted during the anonymisation process to ensure the collective data for each patient is not identifiable.

| **Information for extraction** | **Conversion of potentially sensitive information** |
|---|---|
| Hospital number (patient identifier) | Anonymised unique patient study code |
| Admission ID (identifier for each admission) | Anonymised unique admission study code |
| Sex | M/F |
| Date of birth | 5-year age bands from 65 to >100 at admission |
| Physiological data (heart rate, blood pressure, respiratory rate, oxygen saturation, temp, MEWS-Score) | n/a<br>*Dates provided as consecutive days using date of admission as Day 1. Times are maintained as recorded in EPIC* |
| Relevant blood biochemistry | Liver function, renal function, clotting factor and blood glucose, full blood count |
| Frailty scores | n/a |
| Medication history | n/a |
| Date of admission | Month of admission |
| Admission specialty | Broad category (Department of Medicine for the Elderly, Medicine or Surgery) |
| Number of ward moves | 0,1,2,3,4,5,6,7,8,>8 |
| Length of stay | 1,2,3,4,5,6,7,8,9,10, ≥11 |
| Discharge to usual place of residence/ new institutionalization | n/a<br>Yes/No |
| Diagnostic codes at discharge | High level ICD-10 codes (I-XXII) |
| Discharge specialty | Broad category (Department of Medicine for the Elderly, Medicine or Surgery) |
| Inpatient mortality (death during this inpatient episode) | Yes/No |
| Death within 30 days of discharge | Yes/No |
| Readmission within 30 days after discharge | Yes/No |
| Delayed transfer of care (define as more than 24 hours after last recorded clinically fit date) | Yes/No |

**Supplementary Figure 1**: Missingness information in the final dataset. A: Overall fraction of missingness by laboratory test results and vital signs (i.e., proportion of total days with missing value for each variable); B: Correlation matrix between missingness patters for all variables; C: Correlation matrix between normalised values for all variables; D: Final predictors matrix used for multiple imputation.

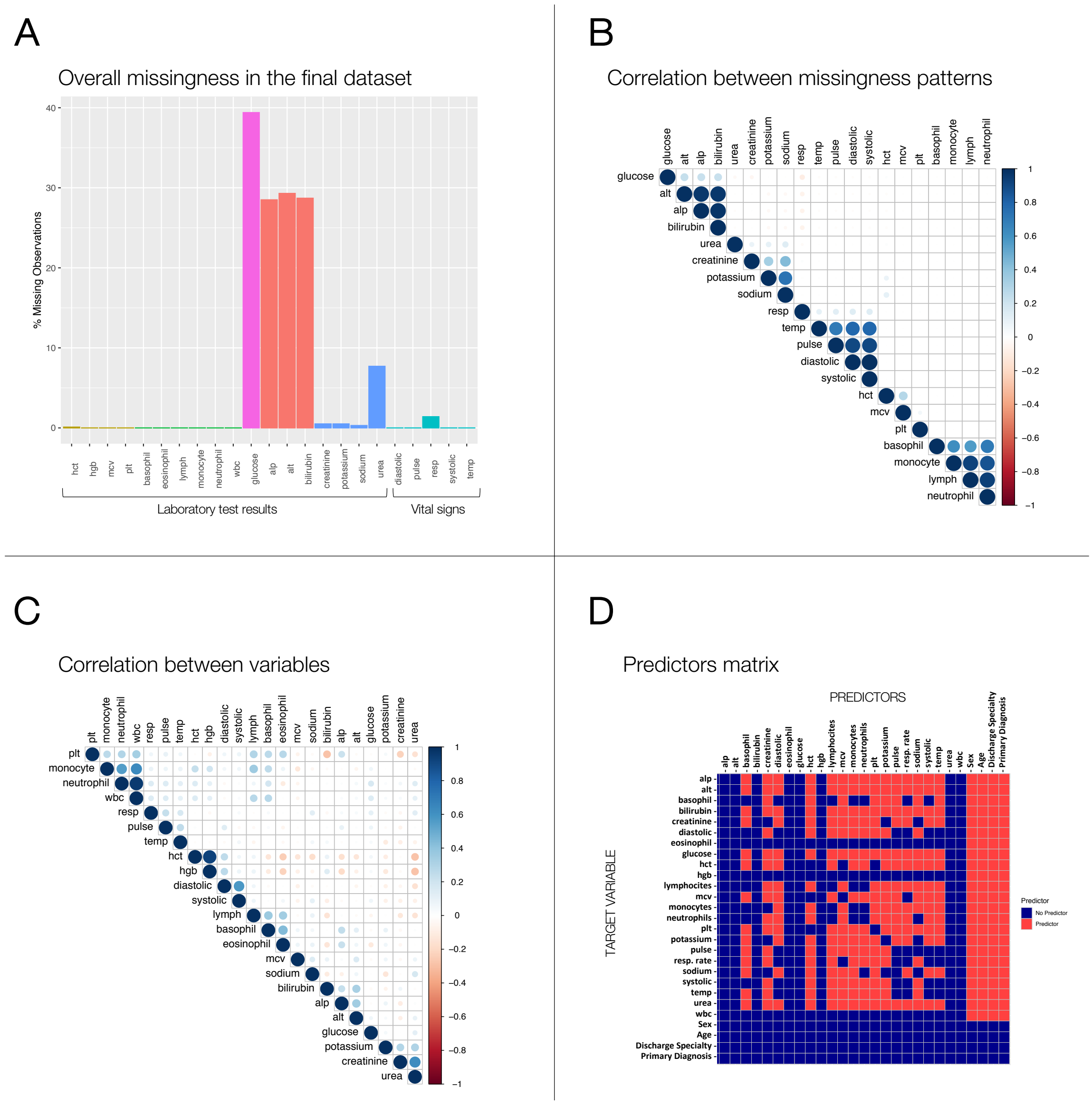

**Supplementary Figure 2**: Results of 2-fold cross-validation for selection of best number of states for HMM models

A

Average log likelihood for 2-fold cross-validation with different number of states

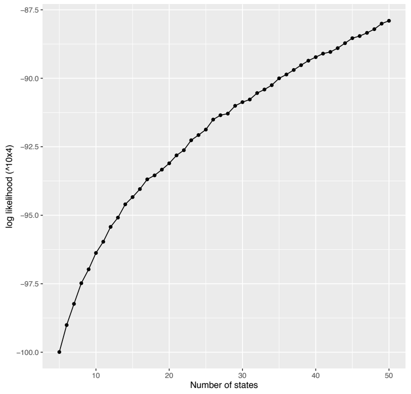


B

Total chi-square score for 2-fold cross-validation with different numbers of states (all p-values << 0.0001)

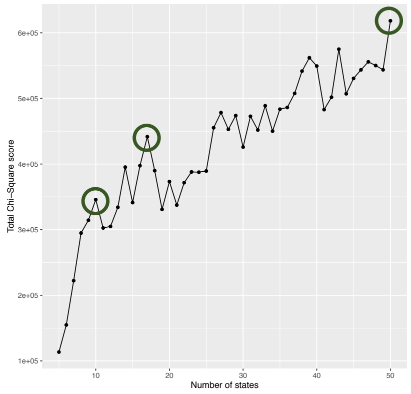


C

Results of 2-fold cross-validation chi-squared t-test as correlation matrices and Pearson’s residuals plots (blue: positive association; red: negative association)

Correlation matrix

Pearson’s residuals plot

10 states

17 states

50 states

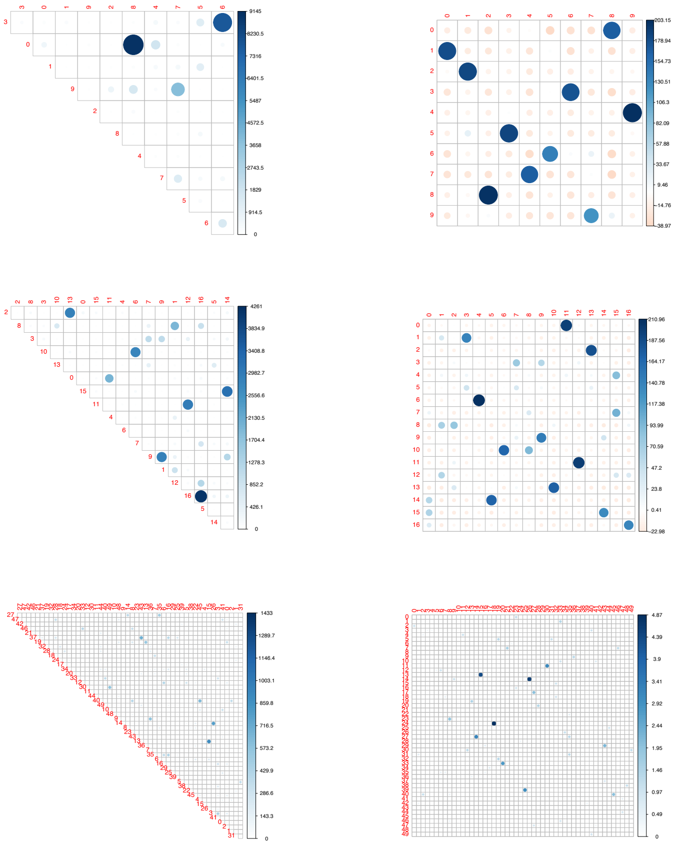

**Supplementary Table 2**: Summary of patients' characteristics in the final cohort divided as 'training and testing' (80 % of the patients) and 'hold-out validation' (20% of the patients) sets. The analyses and results in this publication are conducted only with the training and test set while the validation is kept apart for future confirmatory analyses.

| Characteristics | | Training and test | Validation |
|---|---|---|---|
| Number of patients | | 8926 | 2232 |
| Sex (%) | Women | 47.2 | 48.2 |
| Age-band (%) | 65-69 | 20.9 | 19.1 |
| | 70-74 | 19.8 | 21.9 |
| | 75-89 | 19.2 | 19.2 |
| | 80-84 | 17.9 | 18.5 |
| | 85-89 | 13.3 | 12.7 |
| | 90-94 | 7 | 6.5 |
| | 95-99 | 1.8 | 1.9 |
| | >=100 | 0.1 | 0.2 |
| Discharge Specialty (%) | Geriatric Medicine | 14.9 | 14.3 |
| | Other Medicine | 51.8 | 53.5 |
| | Surgery | 33.2 | 32.2 |
| Frailty category at admission (%) | 1 | 21.6 | 21.3 |
| | 2 | 7.9 | 8.1 |
| | 3 | 7.8 | 7.3 |
| | 4 | 4 | 4.6 |
| | Not recorded | 58.7 | 58.7 |
| Length of Stay (%) | <=3 | 28.1 | 29.8 |
| | 4-5 | 20.7 | 19.4 |
| | 6-10 | 26 | 25.8 |
| | >=11 | 25.1 | 25 |
| Number of diagnoses recorded at discharge (%) | 1-3 | 34.3 | 35 |
| | 4-5 | 20.8 | 21.1 |
| | 6-8 | 20.4 | 18.4 |
| | >=9 | 15.1 | 15.3 |
| | Not recorded | 9.4 | 10.2 |
| Inpatient Mortality (%) | Inpatient Death | 8.4 | 7.7 |
| | Discharged Alive | 91.6 | 92.3 |
| 30-day post-discharge readmission (%) | Readmitted | 14 | 15.1 |
| 30-day post-discharge mortality (%) | Recorded Death | 3.2 | 3.1 |
| Primary diagnosis at admission as top-level ICD-10 codes (%) | A | 6.9 | 7.7 |
| | B | 0.3 | 0.7 |
| | C | 5.4 | 5.2 |
| | D | 1.9 | 1.9 |
| | E | 2.4 | 2.6 |
| | F | 0.7 | 0.9 |
| | G | 1.6 | 1.8 |
| | H | 0.2 | 0.1 |
| | I | 16.2 | 15.9 |
| | J | 14.5 | 13.7 |
| | K | 16.5 | 17.4 |
| | L | 1.8 | 1.7 |
| | M | 2.3 | 1.9 |
| | N | 8.9 | 8 |
| | Q | 0.1 | 0.1 |
| | R | 8.3 | 8.1 |
| | S | 9.8 | 9.6 |
| | T | 2.3 | 2.5 |
| | Z | 0.1 | 0.04 |

### **Supplementary Figure 3**: Evaluation of missingness imputation using Multiple Imputation and Linear Interpolation imputation methods

A

Multiple Imputation:

Distribution of original (blue) and imputed (red) values for 10 imputed datasets

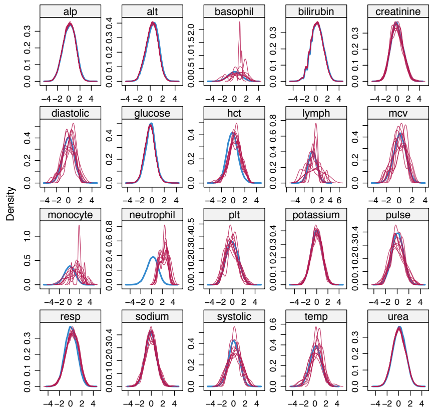


| Number of missing values per variable | | | | |
|---|---|---|---|---|
| 23117 | 23715 | **4** | 23270 | 429 |
| **22** | 31918 | 53 | **7** | **11** |
| **6** | **8** | **35** | 394 | 27 |
| 1130 | 226 | **22** | **36** | 6241 |

B

Multiple Imputation:

Converge plots for multiple iterations

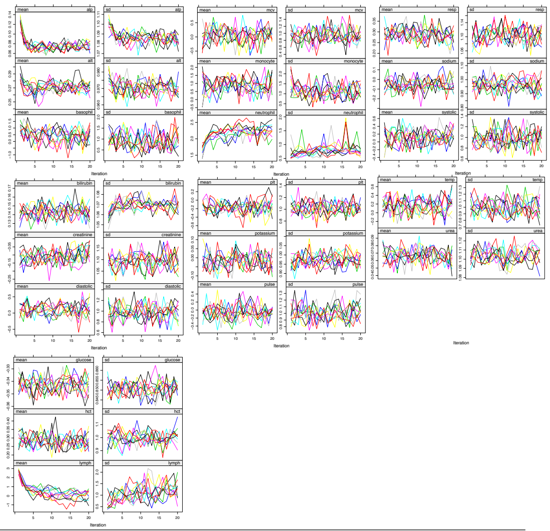

C

Multiple Imputation:

Distribution of randomly inserted (green) vs originally missing (blue) and non-missing (red) values

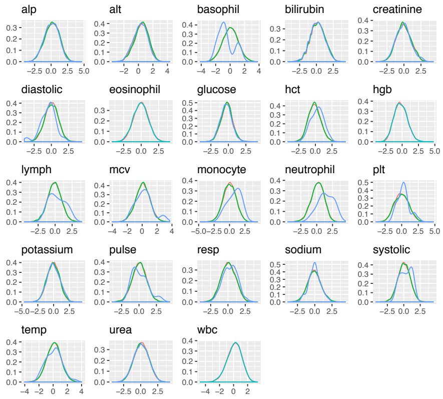


D

Linear Interpolation Imputation:

Distribution of randomly inserted (green) vs originally missing (blue), non-missing (red) and MI imputed (purple )values

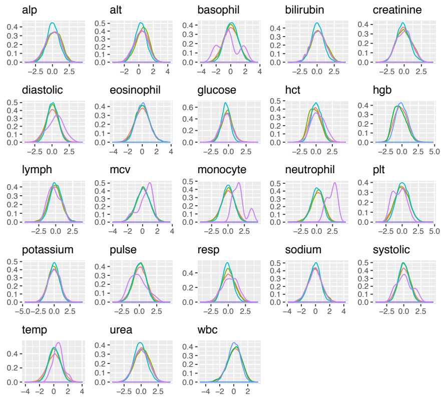


### **Supplementary Figures 4**: See separate document Supplementary_Figures_4.pdf

**Supplementary Table 3**: Clinical interpretation of HMM states; The most predominant features are highlighted in green

| States | | Features | | | Hospital Outcome |
|---|---|---|---|---|---|
| | | **Pattern of laboratory and physiological abnormalities** | **Association with Primary Diagnosis** | **Temporal relationship with Admission Episode** | **Association with LOS/ Inpatient death** |
| DISEASE-LIKE | Hepatic | High LFTs | **Digestive system (K) and neoplasms (C)** | Throughout admission episode | Long LOS |
| | Stable renal | High creatinine and urea and low haemaglobin and haematocrit | **Genitourinary system (N)** | Throughout admission episode, | Long LOS |
| | Unstable renal | High creatinine and urea and low haemaglobin and haematocrit | **Genitourinary system (N) (and other diagnostic codes to a lesser extent)** | Throughout admission episode | Inpatient death |
| | Stable but static renal | Low haemoglobin and haematocrit and mildly high urea and creatinine. Normal vital signs and WBC | Present in most diagnostic codes | Mid-late stages of the admission episode | Long LOS<br>Discharge alive |
| | Blood dyscrasia | All FBC parameters deranged in both directions with wide variance for WBC counts | **Neoplasms (C) and diseases of the blood (D)** | Throughout admission episode | - |
| | Bone marrow suppression | All FBC parameters low except MCV which is high | **Diseases of the blood (D) and neoplasms (C)** | Throughout admission episode | - |
| ADMISSION STAGE-LIKE | Acute presentation | High haemoglobin, haematocrit, total WBC, neutrophils and vital signs | Present in all diagnostic codes | **Strongly overrepresented on Day 1 of the admission episode, regardless of inpatient death or LOS** | - |
| | Treatment response (1) | Similar pattern to acute presentation state but all parameters nearer group mean | Present in all diagnostic codes | **Day 2 to Day 8 (or discharge) of the admission episode** | Short LOS<br>Discharge alive |
| | Treatment response (2) | Low lymphocytes. All other parameters near group mean. | Present in most diagnostic codes except genitourinary (N) and symptoms/ signs/ clinical findings (R) | **Day 2 onwards of the admission episode but usually resolved by discharge if discharged alive** | Inpatient death |
| | Early discharge | All parameters near group mean. Where slight deviations occur, these tend to be in directions that would be closer to a 'healthy population' mean e.g., slightly low WBC and neutrophils | Present in all diagnostic codes | **The last days of short admission episodes** | Short LOS<br>Discharge alive |

| | | | | | |
|---|---|---|---|---|---|
| | Pre-discharge | Low creatinine and urea and high platelets. Other parameters near group mean | Present in all diagnostic codes | **The last days of long admission episodes, more often in those discharged alive** | Long LOS Discharge alive |
| PHYSIOLOGICAL-LIKE | Early inflammatory response | **Markedly high WBC and neutrophils, High urea, respiratory rate, and heart rate** | Present in all diagnostic codes | Throughout admission episode | Inpatient death |
| | Resolving inflammatory response | **High WBC, neutrophils and platelets and all other parameters either higher or lower compared to group mean, except for respiratory rate** | Present in A, C, I, J, and K | **Throughout admission episode although more common after Day 1** | Long LOS Discharge alive |
| | Autoimmune/ atopic | **Markedly higher basophils, eosinophils and lymphocytes** | Uncommon in all diagnostic codes | Throughout admission episode | Long LOS |
| | Acute thrombotic | **High haemoglobin, haematocrit and lymphocytes** | Present in all diagnostic codes and especially in Diseases of the circulatory system (I) and symptoms/ signs/ clinical findings (R) | Throughout admission episode | Short LOS Discharge alive |
| | Prolonged illness | **Most parameters higher than group mean (especially respiratory rate) except haemoglobin, haematocrit and urea which are lower** | Present in most diagnostic codes except Genitourinary system (N) and symptoms/ signs/ clinical findings (R) | **More common after Day 4-5** | Long LOS |
| | Other illness presentation | All parameters at or near group mean | Present in most diagnostic codes with slight preference for Endocrine, nutritional and metabolic diseases (E) | More common in first 3-4 days of admission | Short LOS |

LOS: length of stay; LFTs: liver function tests; FBC: full blood count; WBC: white blood cells; MCV: mean cell volume; Not all states could be classified as confidently after initial visual clinical interpretation (either no or >1 blue predominant feature).

**Supplementary Figures 5**: See separate document Supplementary_Figures_5.pdf

**Supplementary Figures 6**: See separate document Supplementary_Figures_6.pdf

**Supplementary Figure 7**: Pearson residuals plots showing associations between patients' assigned main states and relevant characteristics. Blue: positive associations; red: negative associations.

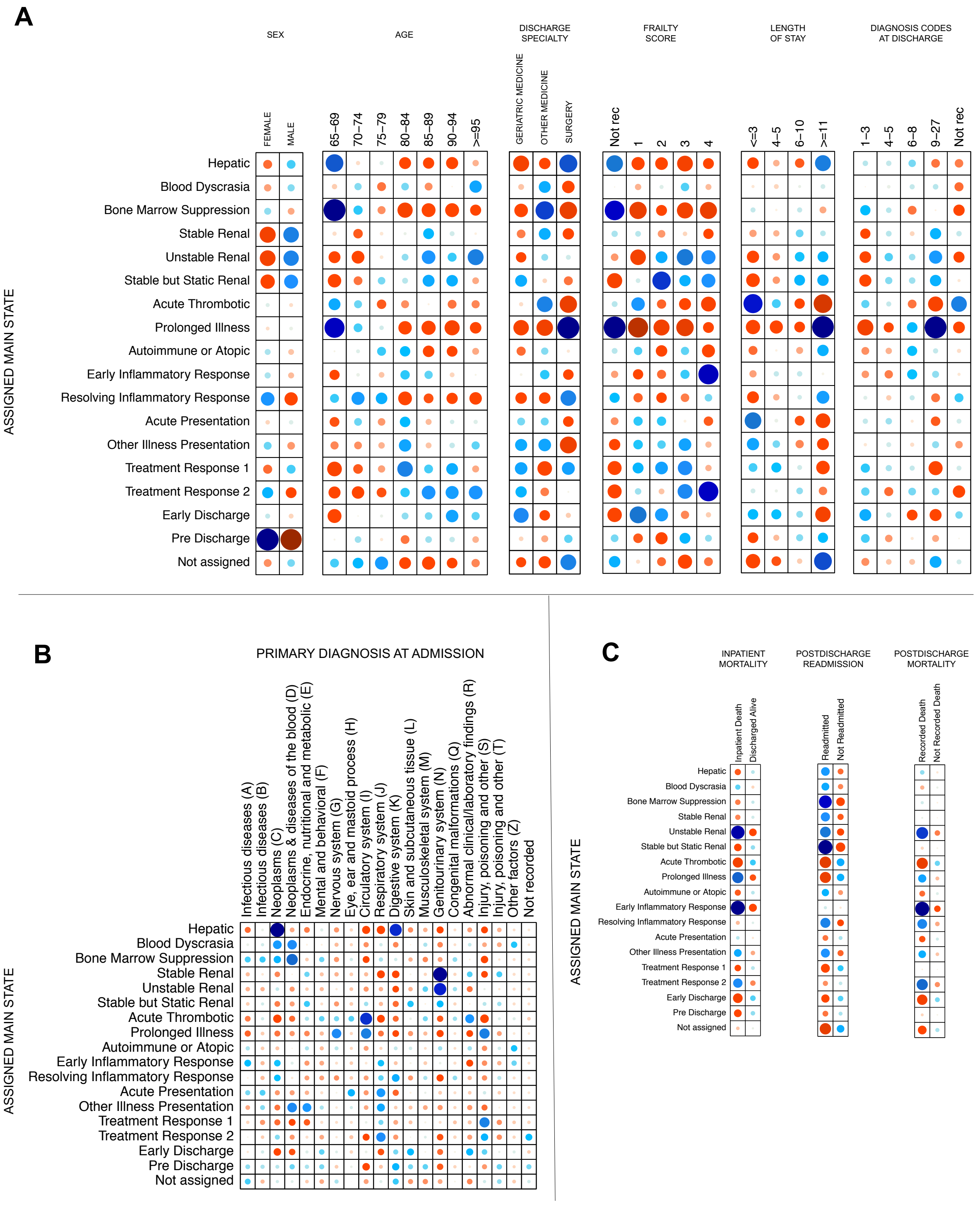

**Supplementary Table 4.** Hypotheses tested for prediction of clinical outcomes with discriminative models

| Model | Representation | Covariates | Input variables | Outcome | | | |
|---|---|---|---|---|---|---|---|
| | | | | Inpatient Mortality | Clinical outcome at 30-days | Diagnosis at Admission | Diagnosis at Discharge |
| LR | Multivariate time series | DM | D1 | x | | | |
| | | | D2 | x | | | |
| | | | D3 | x | | | |
| | | | D1D2 | x | | | |
| | | | D2D3 | x | | | |
| | | | D1D2D3 | x | | | |
| | | DM + PDA | D1 | x | | | |
| | | | D2 | x | | | |
| | | | D3 | x | | | |
| | | | D1D2 | x | | | |
| | | | D2D3 | x | | | |
| | | | D1D2D3 | x | | | |
| LR | HMM states | DM | D1 | x | | | |
| | | | D2 | x | | | |
| | | | D3 | x | | | |
| | | | D1D2 | x | | | |
| | | | D2D3 | x | | | |
| | | | D1D2D3 | x | | | |
| | | DM + PDA | D1 | x | | | |
| | | | D2 | x | | | |
| | | | D3 | x | | | |
| | | | D1D2 | x | | | |
| | | | D2D3 | x | | | |
| | | | D1D2D3 | x | | | |
| RF | Multivariate time series | DM | D1 | x | x | x | x |
| | | | D2 | x | x | x | x |
| | | | D3 | x | x | x | x |
| | | | D1D2 | x | x | x | x |
| | | | D2D3 | x | x | x | x |
| | | | D1D2D3 | x | x | x | x |
| | | DM + PDA | D1 | x | x | | |
| | | | D2 | x | x | | |
| | | | D3 | x | x | | |
| | | | D1D2 | x | x | | |
| | | | D2D3 | x | x | | |
| | | | D1D2D3 | x | x | | |
| RF | HMM states | DM | D1 | x | x | x | x |
| | | | D2 | x | x | x | x |
| | | | D3 | x | x | x | x |
| | | | D1D2 | x | x | x | x |
| | | | D2D3 | x | x | x | x |
| | | | D1D2D3 | x | x | x | x |
| | | DM + PDA | D1 | x | x | | |
| | | | D2 | x | x | | |
| | | | D3 | x | x | | |
| | | | D1D2 | x | x | | |
| | | | D2D3 | x | x | | |
| | | | D1D2D3 | x | x | | |

**Supplementary Table 5.** Selected best models (based on highest performance on ROC-AUC on test dataset) and their parameters identified using hyperparameter tunning.

| Predicted Outcome | Model | Input Variables | Data Representation | # trees | min sample size | min sample leaf | max features | max depth | bootstrap | Best Score (ROC AUC) |
|---|---|---|---|---|---|---|---|---|---|---|
| Inpatient Mortality | RF | D3 + PDA | MVTS | 1000 | 5 | 2 | log2 | 20 | TRUE | 0.849 |
| 30-day clinical outcome | RF | D1D2D3 + PDA | MVTS | 1800 | 2 | 4 | auto | 10 | FALSE | 0.588 |
| Primary Diagnosis at Admission | RF | D1D2D3 + NO PDA | MVTS | 1800 | 2 | 4 | auto | 10 | FALSE | 0.626 |
| Diagnosis at Discharge | RF | D1D2D3 + NO PDA | MVTS | 1800 | 2 | 4 | auto | 10 | FALSE | 0.584 |

**Supplementary Table 6A.** ROC-AUC results for prediction of inpatient mortality as mean (SD) in the *training and test* dataset

| INPUT VARIABLES | | RF | | LR | |
|---|---|---|---|---|---|
| | | MVTS | STATES | MVTS | STATES |
| NO PDA | D1 | 0.757 (0.001) | 0.686 (0.001) | 0.732 (0) | 0.698 (0) |
| | D2 | 0.799 (0.001) | 0.737 (0.001) | 0.785 (0) | 0.744 (0) |
| | D3 | 0.844 (0.001) | **0.778 (0)** | 0.825 (0) | **0.782 (0)** |
| | D1D2 | 0.814 (0.001) | 0.742 (0) | 0.803 (0) | 0.744 (0) |
| | D2D3 | 0.841 (0.001) | 0.764 (0.001) | 0.826 (0) | 0.776 (0) |
| | D1D2D3 | **0.848 (0.001)** | 0.765 (0.001) | **0.832 (0)** | 0.774 (0) |
| PDA | D1 | 0.77 (0.002) | 0.728 (0.001) | 0.746 (0) | 0.729 (0) |
| | D2 | 0.81 (0.003) | 0.764 (0) | 0.793 (0) | 0.76 (0) |
| | D3 | **0.851 (0.002)** | **0.792 (0.001)** | 0.829 (0) | **0.791 (0)** |
| | D1D2 | 0.816 (0.001) | 0.762 (0.001) | 0.805 (0) | 0.759 (0) |
| | D2D3 | 0.844 (0.001) | 0.78 (0) | 0.828 (0) | 0.786 (0) |
| | D1D2D3 | 0.847 (0.002) | 0.778 (0.001) | **0.833 (0)** | 0.786 (0) |

**Supplementary Table 6B.** ROC-AUC results for prediction of inpatient mortality as mean (SD) in the *hold-out validation* dataset

| INPUT VARIABLES | | RF | | LR | |
|---|---|---|---|---|---|
| | | MVTS | STATES | MVTS | STATES |
| NO PDA | D1 | 0.781 (0.002) | 0.632 (0.001) | 0.755 (0) | **0.639 (0)** |
| | D2 | 0.791 (0.002) | 0.623 (0.001) | 0.785 (0) | 0.624 (0) |
| | D3 | **0.853 (0.001)** | 0.575 (0.002) | 0.834 (0) | 0.559 (0) |
| | D1D2 | 0.815 (0.002) | **0.633 (0.002)** | 0.814 (0) | 0.556 (0) |
| | D2D3 | 0.849 (0.001) | 0.583 (0.001) | 0.839 (0) | 0.575 (0) |
| | D1D2D3 | **0.86 (0.001)** | 0.586 (0.005) | **0.859 (0)** | 0.529 (0) |
| PDA | D1 | 0.801 (0.003) | **0.683 (0.002)** | 0.789 (0) | **0.691 (0)** |
| | D2 | 0.81 (0.001) | 0.671 (0.002) | 0.822 (0) | 0.679 (0) |
| | D3 | **0.863 (0.001)** | 0.632 (0.005) | 0.853 (0) | 0.627 (0) |
| | D1D2 | 0.822 (0.002) | 0.669 (0.004) | 0.838 (0) | 0.622 (0) |
| | D2D3 | 0.856 (0.002) | 0.63 (0.002) | 0.856 (0) | 0.634 (0) |
| | D1D2D3 | **0.862 (0.002)** | 0.63 (0.004) | **0.869 (0)** | 0.625 (0) |

**Supplementary Table 7A.** Precision-Recall Curve AUC results for prediction of inpatient mortality as mean (SD) in the *training and test* dataset

| INPUT VARIABLES | | RF | | LR | |
|---|---|---|---|---|---|
| | | MVTS | STATES | MVTS | STATES |
| NO PDA | D1 | 0.242 (0.002) | 0.166 (0.001) | 0.217 (0) | 0.178 (0) |
| | D2 | 0.328 (0.004) | 0.212 (0.003) | 0.279 (0) | 0.212 (0) |
| | D3 | **0.378 (0.003)** | **0.248 (0.003)** | 0.371 (0) | **0.252 (0)** |
| | D1D2 | 0.32 (0.004) | 0.204 (0.001) | 0.315 (0) | 0.208 (0) |
| | D2D3 | 0.353 (0.003) | 0.231 (0.002) | 0.367 (0) | 0.244 (0) |
| | D1D2D3 | 0.368 (0.002) | 0.22 (0.001) | **0.39 (0)** | 0.23 (0) |
| PDA | D1 | 0.257 (0.003) | 0.221 (0.002) | 0.232 (0) | 0.193 (0) |
| | D2 | 0.34 (0.005) | 0.259 (0.001) | 0.295 (0) | 0.236 (0) |
| | D3 | **0.392 (0.005)** | 0.29 (0.003) | 0.369 (0) | **0.27 (0)** |
| | D1D2 | 0.314 (0.004) | 0.24 (0.005) | 0.323 (0) | 0.225 (0) |
| | D2D3 | 0.364 (0.003) | **0.268 (0.001)** | 0.364 (0) | 0.255 (0) |
| | D1D2D3 | 0.36 (0.005) | 0.255 (0.002) | **0.395 (0)** | 0.25 (0) |

**Supplementary Table 7B.** Precision-Recall Curve AUC results for prediction of inpatient mortality as mean (SD) in the *hold-out validation* dataset

| INPUT VARIABLES | | RF | | LR | |
|---|---|---|---|---|---|
| | | MVTS | STATES | MVTS | STATES |
| NO PDA | D1 | 0.209 (0.002) | **0.125 (0.001)** | 0.209 (0) | **0.167 (0)** |
| | D2 | 0.265 (0.005) | 0.113 (0.001) | 0.303 (0) | 0.134 (0) |
| | D3 | **0.364 (0.002)** | 0.089 (0.001) | 0.381 (0) | 0.099 (0) |
| | D1D2 | 0.269 (0.004) | 0.115 (0.001) | 0.34 (0) | 0.104 (0) |
| | D2D3 | 0.356 (0.003) | 0.088 (0) | 0.397 (0) | 0.101 (0) |
| | D1D2D3 | 0.355 (0.005) | 0.09 (0.001) | **0.422 (0)** | 0.085 (0) |
| PDA | D1 | 0.225 (0.005) | **0.169 (0.004)** | 0.236 (0) | **0.179 (0)** |
| | D2 | 0.276 (0.007) | 0.133 (0.004) | 0.329 (0) | 0.145 (0) |
| | D3 | **0.381 (0.004)** | 0.107 (0.003) | 0.411 (0) | 0.117 (0) |
| | D1D2 | 0.265 (0.003) | 0.13 (0.003) | 0.354 (0) | 0.116 (0) |
| | D2D3 | 0.361 (0.005) | 0.104 (0.002) | 0.422 (0) | 0.124 (0) |
| | D1D2D3 | 0.341 (0.004) | 0.104 (0.002) | **0.438 (0)** | 0.118 (0) |

**Supplementary Table 8A**. Precision, Recall and F1-scores results for prediction of inpatient mortality as mean (SD) in the *training and test* dataset

| PRECISION | | | | | |
|---|---|---|---|---|---|
| INPUT VARIABLES | | RF | | LR | |
| | | MVTS | STATES | MV TS | STATES |
| NO PDA | D1 | 0.018 (0) | 0.018 (0) | 0.018 (0) | 0.018 (0) |
| | D2 | 0.032 (0) | 0.032 (0) | 0.032 (0) | 0.032 (0) |
| | D3 | 0.596 (0.014) | **0.188 (0.001)** | 0.212 (0) | 0.172 (0) |
| | D1D2 | 0.5 (0.04) | 0.17 (0.001) | 0.2 (0) | 0.16 (0) |
| | D2D3 | 0.603 (0.031) | 0.184 (0.002) | 0.218 (0) | 0.17 (0) |
| | D1D2D3 | **0.617 (0.028)** | 0.179 (0.002) | **0.223 (0)** | **0.176 (0)** |
| PDA | D1 | 0.675 (0.026) | 0.173 (0.001) | 0.176 (0) | 0.167 (0) |
| | D2 | **1 (0)** | 0.18 (0.001) | 0.196 (0) | 0.166 (0) |
| | D3 | 0.718 (0.049) | **0.199 (0.002)** | **0.215 (0)** | **0.191 (0)** |
| | D1D2 | 0 (0) | 0.188 (0.003) | 0.208 (0) | 0.174 (0) |
| | D2D3 | 0.523 (0.139) | 0.19 (0.001) | 0.214 (0) | 0.183 (0) |
| | D1D2D3 | 0.571 (0.053) | 0.191 (0.003) | 0.223 (0) | 0.181 (0) |
| RECALL | | | | | |
| INPUT VARIABLES | | RF | | LR | |
| | | MVTS | STATES | MVTS | STATES |
| NO PDA | D1 | 0.018 (0) | 0.556 (0) | 0.673 (0) | 0.606 (0) |
| | D2 | 0.032 (0) | 0.647 (0) | 0.704 (0) | 0.686 (0) |
| | D3 | **0.167 (0)** | **0.701 (0)** | **0.743 (0)** | **0.726 (0)** |
| | D1D2 | 0.026 (0) | 0.623 (0) | 0.73 (0) | 0.721 (0) |
| | D2D3 | 0.054 (0) | 0.678 (0) | 0.735 (0) | **0.726 (0)** |
| | D1D2D3 | 0.092 (0) | 0.677 (0) | **0.743 (0)** | 0.712 (0) |
| PDA | D1 | 0.009 (0) | 0.581 (0) | 0.668 (0) | 0.664 (0) |
| | D2 | 0.004 (0) | 0.623 (0) | 0.717 (0) | 0.681 (0) |
| | D3 | 0.047 (0) | **0.693 (0)** | 0.712 (0) | **0.752 (0)** |
| | D1D2 | 0 (0) | 0.621 (0) | 0.735 (0) | 0.708 (0) |
| | D2D3 | 0.011 (0) | 0.678 (0) | 0.704 (0) | 0.73 (0) |
| | D1D2D3 | 0.046 (0) | 0.679 (0) | **0.739 (0)** | 0.708 (0) |
| F1-SCORE | | | | | |
| INPUT VARIABLES | | RF | | LR | |
| | | MVTS | STATES | MVTS | STATES |
| NO PDA | D1 | 0.035 (0.005) | 0.246 (0.002) | 0.271 (0) | 0.222 (0) |
| | D2 | 0.06 (0.014) | 0.26 (0.002) | 0.288 (0) | 0.25 (0) |
| | D3 | **0.261 (0.01)** | **0.296 (0.002)** | 0.329 (0) | 0.278 (0) |
| | D1D2 | 0.05 (0.005) | 0.267 (0.002) | 0.314 (0) | 0.262 (0) |
| | D2D3 | 0.099 (0.009) | 0.29 (0.002) | 0.336 (0) | 0.275 (0) |
| | D1D2D3 | 0.161 (0.008) | 0.283 (0.002) | **0.343 (0)** | **0.282 (0)** |
| PDA | D1 | 0.018 (0.003) | 0.267 (0.002) | 0.279 (0) | 0.267 (0) |
| | D2 | 0.009 (0) | 0.279 (0.002) | 0.308 (0) | 0.266 (0) |
| | D3 | **0.089 (0.008)** | **0.309 (0.002)** | 0.33 (0) | **0.305 (0)** |
| | D1D2 | 0 (0) | 0.289 (0.004) | 0.325 (0) | 0.279 (0) |
| | D2D3 | 0.022 (0.01) | 0.297 (0.002) | 0.329 (0) | 0.292 (0) |
| | D1D2D3 | 0.084 (0.011) | 0.298 (0.004) | **0.343 (0)** | 0.289 (0) |

**Supplementary Table 8B**. Precision, Recall and F1-scores results for prediction of inpatient mortality as mean (SD) in the *hold-out validation* dataset

| PRECISION | | | | | |
|---|---|---|---|---|---|
| INPUT VARIABLES | | RF | | LR | |
| | | MVTS | STATES | MV TS | STATES |
| NO PDA | D1 | 0.252 (0.049) | 0.133 (0.001) | 0.158 (0) | 0.117 (0) |
| | D2 | 0.623 (0.07) | 0.12 (0.003) | 0.162 (0) | **0.119 (0)** |
| | D3 | 0.547 (0.019) | 0.092 (0.003) | 0.214 (0) | 0.092 (0) |
| | D1D2 | 0.489 (0.092) | **0.135 (0.004)** | 0.191 (0) | 0.085 (0) |
| | D2D3 | **0.802 (0.078)** | 0.082 (0.002) | 0.218 (0) | 0.097 (0) |
| | D1D2D3 | 0.69 (0.042) | 0.089 (0.004) | **0.227 (0)** | 0.081 (0) |
| PDA | D1 | 0.1 (0.316) | 0.137 (0.002) | 0.177 (0) | 0.131 (0) |
| | D2 | 0 (0) | 0.136 (0.002) | 0.191 (0) | **0.135 (0)** |
| | D3 | 0.751 (0.048) | 0.121 (0.004) | 0.212 (0) | 0.114 (0) |
| | D1D2 | 0 (0) | **0.144 (0.004)** | 0.202 (0) | 0.099 (0) |
| | D2D3 | **0.9 (0.316)** | 0.12 (0.003) | 0.22 (0) | 0.11 (0) |
| | D1D2D3 | 0.708 (0.114) | 0.125 (0.005) | **0.238 (0)** | 0.11 (0) |
| RECALL | | | | | |
| INPUT VARIABLES | | RF | | LR | |
| | | MVTS | STATES | MVTS | STATES |
| NO PDA | D1 | 0.006 (0) | **0.485 (0.004)** | 0.667 (0) | 0.526 (0) |
| | D2 | 0.034 (0.006) | 0.377 (0.007) | 0.684 (0) | 0.485 (0) |
| | D3 | **0.151 (0.005)** | 0.364 (0.019) | 0.749 (0) | 0.503 (0) |
| | D1D2 | 0.016 (0.002) | 0.452 (0.02) | 0.713 (0) | **0.637 (0)** |
| | D2D3 | 0.051 (0.008) | 0.298 (0.006) | 0.743 (0) | 0.503 (0) |
| | D1D2D3 | 0.079 (0.003) | 0.329 (0.021) | **0.76 (0)** | 0.561 (0) |
| PDA | D1 | 0.001 (0.002) | **0.526 (0.005)** | 0.725 (0) | 0.532 (0) |
| | D2 | 0 (0) | 0.414 (0.013) | 0.76 (0) | 0.526 (0) |
| | D3 | **0.051 (0.007)** | 0.456 (0.022) | 0.719 (0) | 0.532 (0) |
| | D1D2 | 0 (0) | 0.462 (0.018) | 0.743 (0) | **0.632 (0)** |
| | D2D3 | 0.007 (0.004) | 0.395 (0.01) | 0.725 (0) | 0.515 (0) |
| | D1D2D3 | 0.032 (0.006) | 0.401 (0.03) | **0.778 (0)** | 0.596 (0) |
| F1-SCORE | | | | | |
| INPUT VARIABLES | | RF | | LR | |
| | | MVTS | STATES | MVTS | STATES |
| NO PDA | D1 | 0.011 (0) | **0.208 (0.001)** | 0.256 (0) | **0.192 (0)** |
| | D2 | 0.064 (0.011) | 0.182 (0.004) | 0.262 (0) | 0.191 (0) |
| | D3 | **0.237 (0.008)** | 0.146 (0.005) | 0.332 (0) | 0.156 (0) |
| | D1D2 | 0.032 (0.005) | **0.208 (0.004)** | 0.301 (0) | 0.15 (0) |
| | D2D3 | 0.097 (0.014) | 0.128 (0.003) | **0.337 (0)** | 0.163 (0) |
| | D1D2D3 | 0.142 (0.006) | 0.14 (0.006) | 0.35 (0) | 0.141 (0) |
| PDA | D1 | 0.001 (0.004) | 0.217 (0.003) | 0.285 (0) | 0.21 (0) |
| | D2 | 0 (0) | 0.204 (0.004) | 0.305 (0) | **0.214 (0)** |
| | D3 | **0.095 (0.012)** | 0.191 (0.006) | 0.327 (0) | 0.188 (0) |
| | D1D2 | 0 (0) | **0.219 (0.004)** | 0.318 (0) | 0.171 (0) |
| | D2D3 | 0.014 (0.007) | 0.184 (0.005) | 0.338 (0) | 0.181 (0) |
| | D1D2D3 | 0.061 (0.012) | 0.191 (0.007) | **0.365 (0)** | 0.186 (0) |

**Supplementary Table 9A.** Weighted ROC-AUC for prediction of 30-day clinical outcome in the *training and test* dataset

| INPUT VARIABLES | | RF | |
|---|---|---|---|
| | | MVTS | STATES |
| NO PDA | D1 | 0.634 (0.001) | 0.588 (0) |
| | D2 | 0.638 (0) | 0.625 (0.001) |
| | D3 | 0.66 (0) | 0.645 (0.001) |
| | D1D2 | 0.653 (0.001) | 0.628 (0.001) |
| | D2D3 | 0.664 (0) | **0.647 (0)** |
| | D1D2D3 | **0.674 (0.001)** | **0.647 (0)** |
| PDA | D1 | 0.649 (0) | 0.636 (0.001) |
| | D2 | 0.654 (0.001) | 0.654 (0) |
| | D3 | 0.679 (0.001) | **0.671 (0)** |
| | D1D2 | 0.661 (0.001) | 0.651 (0) |
| | D2D3 | 0.673 (0.001) | 0.667 (0) |
| | D1D2D3 | **0.681 (0.001)** | 0.664 (0) |

**Supplementary Table 9B.** Weighted ROC-AUC for prediction of 30-day clinical outcome in the *hold-out validation* dataset

| INPUT VARIABLES | | RF | |
|---|---|---|---|
| | | MV TS | STATES |
| NO PDA | D1 | 0.629 (0.001) | 0.553 (0.001) |
| | D2 | 0.64 (0.001) | 0.552 (0.001) |
| | D3 | 0.664 (0) | 0.54 (0.001) |
| | D1D2 | 0.657 (0.001) | **0.558 (0.001)** |
| | D2D3 | 0.668 (0.001) | 0.546 (0.001) |
| | D1D2D3 | **0.676 (0)** | 0.555 (0.002) |
| PDA | D1 | 0.641 (0.001) | **0.592 (0.002)** |
| | D2 | 0.656 (0) | 0.585 (0.001) |
| | D3 | 0.676 (0.001) | 0.575 (0.001) |
| | D1D2 | 0.663 (0.001) | 0.585 (0.001) |
| | D2D3 | 0.676 (0.001) | 0.577 (0.001) |
| | D1D2D3 | **0.681 (0.001)** | 0.582 (0.001) |

**Supplementary Table 10A.** Weighted precision, recall and F1-score for prediction of 30-day clinical outcome in the *training and test* dataset

| INPUT VARIABLES | | RF | | | | | |
|---|---|---|---|---|---|---|---|
| | | Precision | | Recall | | F1-score | |
| | | MVTS | MV TS | MVTS | STATES | MVTS | STATES |
| NO PDA | D1 | 0.64 (0.002) | 0.625 (0) | 0.662 (0.002) | 0.578 (0.002) | 0.648 (0.002) | 0.576 (0) |
| | D2 | 0.647 (0.002) | 0.641 (0.001) | 0.651 (0.002) | **0.592 (0.005)** | 0.646 (0.001) | 0.591 (0.003) |
| | D3 | 0.659 (0.001) | 0.651 (0.002) | 0.68 (0.002) | 0.591 (0.006) | **0.669 (0.002)** | **0.593 (0.005)** |
| | D1D2 | 0.651 (0.002) | 0.64 (0.002) | 0.672 (0.002) | 0.582 (0.007) | 0.656 (0.002) | 0.588 (0.004) |
| | D2D3 | **0.66 (0.001)** | **0.655 (0.001)** | 0.685 (0.002) | 0.574 (0.002) | 0.668 (0.001) | 0.585 (0.002) |
| | D1D2D3 | **0.66 (0.004)** | 0.652 (0.001) | **0.692 (0.002)** | 0.58 (0.003) | **0.669 (0.002)** | 0.588 (0.001) |
| PDA | D1 | 0.653 (0.003) | 0.645 (0.001) | 0.65 (0.002) | 0.618 (0.005) | 0.646 (0.002) | 0.611 (0.003) |
| | D2 | 0.652 (0.003) | 0.651 (0.001) | 0.641 (0.002) | **0.632 (0.005)** | 0.645 (0.002) | **0.62 (0.003)** |
| | D3 | 0.661 (0.001) | 0.655 (0.001) | 0.67 (0.002) | 0.624 (0.003) | 0.665 (0.001) | 0.617 (0.002) |
| | D1D2 | 0.655 (0.004) | 0.649 (0.001) | 0.667 (0.003) | 0.621 (0.001) | 0.655 (0.002) | 0.608 (0.001) |
| | D2D3 | 0.66 (0.002) | **0.658 (0.001)** | 0.678 (0.003) | 0.604 (0.002) | 0.665 (0.002) | 0.607 (0.001) |
| | D1D2D3 | **0.662 (0.002)** | 0.657 (0.002) | **0.688 (0.002)** | 0.606 (0.002) | **0.668 (0.001)** | 0.597 (0.001) |

**Supplementary Table 10B.** Weighted precision, recall and F1-score for prediction of 30-day clinical outcome in the *hold-out validation* dataset

| INPUT VARIABLES | | RF | | | | | |
|---|---|---|---|---|---|---|---|
| | | Precision | | Recall | | F1-score | |
| | | MVTS | MV TS | MVTS | STATES | MVTS | STATES |
| NO PDA | D1 | 0.634 (0.002) | 0.612 (0.001) | 0.659 (0.002) | **0.615 (0.006)** | 0.644 (0.001) | **0.61 (0.003)** |
| | D2 | 0.633 (0.002) | 0.61 (0.001) | 0.65 (0.002) | 0.542 (0.004) | 0.639 (0.002) | 0.556 (0.003) |
| | D3 | **0.651 (0.001)** | 0.601 (0.002) | 0.684 (0.002) | 0.493 (0.011) | **0.665 (0.001)** | 0.517 (0.007) |
| | D1D2 | 0.636 (0.003) | **0.613 (0.002)** | 0.672 (0.002) | 0.586 (0.012) | 0.648 (0.002) | 0.594 (0.007) |
| | D2D3 | 0.654 (0.002) | 0.612 (0.002) | 0.686 (0.001) | 0.479 (0.01) | 0.664 (0.001) | 0.511 (0.007) |
| | D1D2D3 | 0.649 (0.002) | 0.611 (0.002) | **0.695 (0.002)** | 0.55 (0.01) | **0.665 (0.001)** | 0.569 (0.006) |
| PDA | D1 | 0.637 (0.001) | 0.622 (0.002) | 0.649 (0.002) | **0.639 (0.01)** | 0.642 (0.002) | **0.623 (0.005)** |
| | D2 | 0.64 (0.001) | 0.619 (0.001) | 0.647 (0.002) | 0.617 (0.006) | 0.64 (0.001) | 0.602 (0.004) |
| | D3 | **0.657 (0.002)** | 0.615 (0.001) | 0.681 (0.002) | 0.557 (0.004) | **0.666 (0.002)** | 0.565 (0.002) |
| | D1D2 | 0.636 (0.001) | 0.617 (0.001) | 0.666 (0.002) | 0.603 (0.005) | 0.646 (0.001) | 0.597 (0.003) |
| | D2D3 | 0.656 (0.002) | 0.621 (0.001) | 0.683 (0.002) | 0.543 (0.004) | 0.665 (0.002) | 0.56 (0.003) |
| | D1D2D3 | 0.649 (0.002) | **0.624 (0.001)** | **0.691 (0.002)** | 0.553 (0.008) | 0.663 (0.002) | 0.568 (0.005) |

**Supplementary Table 11A**. Precision, Recall and F1-score by class for prediction of 30-day clinical outcome in the *training and test* dataset

| | | | | | | | | | | | |
|---|---|---|---|---|---|---|---|---|---|---|---|
| PRECISION | | | | | | | | | | | |
| INPUT VARIABLES | | RF | | | | | | | | | |
| | | MVTS | | | | | STATES | | | | |
| | | ID | DA | PDR | PDRM | PDM | ID | DA | PDR | PRDM | PDM |
| NO PDA | D1 | 0.318 (0.016) | 0.786 (0.001) | 0.161 (0.009) | 0 (0) | **0.076 (0.019)** | 0.134 (0.001) | 0.793 (0) | 0.133 (0.002) | 0.008 (0) | 0.04 (0.002) |
| | D2 | 0.326 (0.007) | 0.795 (0.001) | 0.159 (0.008) | 0 (0) | 0.067 (0.014) | 0.174 (0.001) | 0.806 (0.001) | 0.155 (0.003) | 0.011 (0) | 0.047 (0.002) |
| | D3 | 0.371 (0.009) | 0.803 (0.001) | 0.184 (0.006) | 0 (0) | 0.049 (0.018) | **0.191 (0.001)** | 0.815 (0.003) | **0.168 (0.005)** | 0.014 (0.002) | **0.048 (0.002)** |
| | D1D2 | 0.384 (0.012) | 0.795 (0.002) | 0.163 (0.008) | 0 (0) | 0 (0) | 0.182 (0.004) | 0.803 (0.002) | 0.154 (0.002) | 0.014 (0.001) | 0.037 (0.004) |
| | D2D3 | 0.381 (0.006) | **0.804 (0.001)** | 0.18 (0.013) | 0 (0) | 0 (0) | 0.19 (0.002) | **0.821 (0.001)** | **0.168 (0.002)** | 0.011 (0.001) | 0.039 (0.002) |
| | D1D2D3 | **0.4 (0.008)** | 0.799 (0.001) | **0.188 (0.015)** | 0 (0) | 0.039 (0.087) | 0.19 (0.002) | 0.816 (0.002) | 0.166 (0.003) | 0.008 (0.004) | 0.044 (0.004) |
| PDA | D1 | 0.324 (0.007) | 0.794 (0.001) | 0.161 (0.009) | 0 (0) | **0.368 (0.095)** | 0.174 (0.003) | 0.809 (0.001) | 0.157 (0.002) | 0.021 (0.001) | 0.062 (0.003) |
| | D2 | 0.329 (0.004) | 0.802 (0.001) | 0.158 (0.008) | 0 (0) | 0.062 (0.043) | 0.196 (0.003) | 0.813 (0.001) | **0.169 (0.004)** | 0.011 (0.011) | **0.064 (0.004)** |
| | D3 | 0.363 (0.009) | **0.807 (0.001)** | 0.181 (0.005) | 0 (0) | 0 (0) | **0.209 (0.001)** | 0.817 (0.001) | 0.171 (0.003) | 0.005 (0.006) | 0.055 (0.003) |
| | D1D2 | 0.379 (0.012) | 0.798 (0.001) | 0.16 (0.009) | 0 (0) | 0.12 (0.14) | 0.193 (0.001) | 0.815 (0.001) | 0.156 (0.002) | 0.009 (0.002) | 0.043 (0.001) |
| | D2D3 | 0.371 (0.011) | 0.805 (0.001) | 0.184 (0.008) | 0 (0) | 0 (0) | 0.203 (0.001) | 0.821 (0.001) | 0.176 (0.003) | 0.012 (0.001) | 0.058 (0.004) |
| | D1D2D3 | **0.408 (0.004)** | 0.801 (0.001) | **0.196 (0.013)** | 0 (0) | 0 (0) | 0.204 (0.001) | **0.823 (0.002)** | 0.158 (0.003) | 0.019 (0.002) | 0.051 (0.002) |
| RECALL | | | | | | | | | | | |
| INPUT VARIABLES | | RF | | | | | | | | | |
| | | MVTS | | | | | STATES | | | | |
| | | ID | DA | PDR | PDRM | PDM | ID | DA | PDR | PRDM | PDM |
| NO PDA | D1 | 0.167 (0.009) | 0.841 (0.002) | 0.121 (0.009) | 0 (0) | 0.017 (0.005) | 0.484 (0.003) | 0.636 (0.001) | 0.399 (0.016) | 0.095 (0) | 0.255 (0.013) |
| | D2 | 0.266 (0.007) | **0.822 (0.001)** | 0.083 (0.006) | 0 (0) | 0.017 (0.005) | 0.635 (0.011) | 0.644 (0.006) | 0.349 (0.015) | 0.095 (0) | 0.302 (0.013) |
| | D3 | **0.329 (0.012)** | 0.843 (0.002) | 0.146 (0.005) | 0 (0) | 0.02 (0.007) | 0.655 (0.008) | 0.634 (0.009) | 0.383 (0.013) | 0.152 (0.02) | 0.343 (0.013) |
| | D1D2 | 0.21 (0.006) | 0.86 (0.002) | 0.065 (0.003) | 0 (0) | 0 (0) | 0.617 (0.014) | 0.64 (0.009) | 0.329 (0.015) | 0.095 (0) | 0.212 (0.034) |
| | D2D3 | 0.293 (0.005) | 0.866 (0.002) | 0.073 (0.004) | 0 (0) | 0 (0) | 0.663 (0.005) | 0.615 (0.004) | 0.362 (0.007) | 0.095 (0) | 0.303 (0.016) |
| | D1D2D3 | 0.262 (0.01) | **0.882 (0.003)** | 0.064 (0.005) | 0 (0) | 0.003 (0.006) | **0.669 (0.01)** | 0.625 (0.003) | 0.349 (0.008) | 0.048 (0.022) | 0.315 (0.035) |
| PDA | D1 | 0.188 (0.004) | 0.823 (0.002) | 0.12 (0.008) | 0 (0) | 0.025 (0.008) | 0.577 (0.023) | 0.686 (0.006) | 0.346 (0.01) | 0.095 (0) | 0.306 (0.018) |
| | D2 | 0.299 (0.004) | 0.806 (0.002) | 0.082 (0.005) | 0 (0) | 0.011 (0.007) | 0.669 (0.007) | 0.692 (0.006) | 0.36 (0.01) | 0.043 (0.042) | 0.306 (0.026) |
| | D3 | **0.361 (0.007)** | 0.826 (0.003) | 0.147 (0.005) | 0 (0) | 0 (0) | 0.68 (0.005) | 0.679 (0.003) | 0.371 (0.009) | 0.019 (0.025) | 0.291 (0.011) |
| | D1D2 | 0.223 (0.006) | 0.852 (0.003) | 0.065 (0.004) | 0 (0) | 0.009 (0.011) | 0.675 (0.004) | 0.667 (0.001) | 0.424 (0.006) | 0.11 (0.023) | 0.26 (0.009) |
| | D2D3 | 0.297 (0.006) | 0.856 (0.003) | 0.079 (0.003) | 0 (0) | 0 (0) | 0.673 (0.004) | 0.654 (0.002) | 0.369 (0.007) | 0.048 (0) | 0.289 (0.02) |
| | D1D2D3 | 0.272 (0.007) | **0.874 (0.001)** | 0.067 (0.004) | 0 (0) | 0 (0) | **0.705 (0.006)** | 0.632 (0.002) | 0.461 (0.01) | 0.219 (0.025) | 0.365 (0.016) |
| F1-SCORE | | | | | | | | | | | |
| INPUT VARIABLES | | RF | | | | | | | | | |
| | | MVTS | | | | | STATES | | | | |
| | | ID | DA | PDR | PDRM | PDM | ID | DA | PDR | PRDM | PDM |
| NO PDA | D1 | 0.219 (0.011) | 0.813 (0.001) | 0.138 (0.009) | 0 (0) | **0.028 (0.008)** | 0.21 (0.001) | 0.706 (0) | 0.199 (0.004) | 0.014 (0.001) | 0.07 (0.003) |
| | D2 | 0.293 (0.007) | 0.808 (0.001) | 0.109 (0.007) | 0 (0) | 0.027 (0.007) | 0.274 (0.001) | 0.716 (0.003) | 0.215 (0.004) | 0.019 (0.001) | 0.082 (0.003) |
| | D3 | **0.349 (0.01)** | 0.822 (0.001) | **0.162 (0.005)** | 0 (0) | **0.028 (0.01)** | 0.296 (0.002) | **0.713 (0.006)** | 0.233 (0.005) | **0.026 (0.003)** | **0.085 (0.003)** |
| | D1D2 | 0.271 (0.007) | 0.826 (0.002) | 0.093 (0.004) | 0 (0) | 0 (0) | 0.281 (0.004) | 0.712 (0.006) | 0.21 (0.004) | 0.024 (0.001) | 0.063 (0.008) |
| | D2D3 | 0.331 (0.004) | 0.834 (0.001) | 0.104 (0.006) | 0 (0) | 0 (0) | **0.296 (0.002)** | 0.703 (0.002) | **0.23 (0.003)** | 0.019 (0.001) | 0.069 (0.004) |

| | | | | | | | | | | | |
|---|---|---|---|---|---|---|---|---|---|---|---|
| | D1D2D3 | 0.317 (0.009) | **0.838 (0.002)** | 0.095 (0.007) | 0 (0) | 0.006 (0.012) | **0.296 (0.003)** | 0.707 (0.001) | 0.225 (0.003) | 0.014 (0.007) | 0.078 (0.007) |
| PDA | D1 | 0.238 (0.004) | 0.808 (0.001) | 0.137 (0.009) | 0 (0) | **0.046 (0.015)** | 0.267 (0.004) | 0.742 (0.004) | 0.216 (0.003) | **0.034 (0.002)** | 0.104 (0.006) |
| | D2 | 0.313 (0.003) | 0.804 (0.001) | 0.108 (0.006) | 0 (0) | 0.018 (0.013) | 0.304 (0.004) | **0.748 (0.004)** | 0.23 (0.005) | 0.017 (0.017) | **0.106 (0.007)** |
| | D3 | **0.362 (0.007)** | 0.817 (0.002) | **0.162 (0.004)** | 0 (0) | 0 (0) | **0.32 (0.002)** | 0.742 (0.002) | 0.234 (0.005) | 0.007 (0.01) | 0.093 (0.004) |
| | D1D2 | 0.28 (0.008) | 0.824 (0.001) | 0.093 (0.005) | 0 (0) | 0.017 (0.02) | 0.3 (0.002) | 0.734 (0.001) | 0.228 (0.003) | 0.017 (0.004) | 0.073 (0.002) |
| | D2D3 | 0.33 (0.008) | 0.829 (0.002) | 0.111 (0.004) | 0 (0) | 0 (0) | 0.312 (0.002) | 0.728 (0.002) | **0.239 (0.004)** | 0.019 (0.001) | 0.096 (0.007) |
| | D1D2D3 | 0.327 (0.006) | **0.836 (0.001)** | 0.1 (0.006) | 0 (0) | 0 (0) | 0.316 (0.002) | 0.715 (0.001) | 0.235 (0.004) | **0.034 (0.004)** | 0.09 (0.003) |

**Supplementary Table 11B**. Precision, Recall and F1-score by class for prediction of 30-day clinical outcome in the *hold-out validation* dataset

| PRECISION | | | | | | | | | | | |
|---|---|---|---|---|---|---|---|---|---|---|---|
| INPUT VARIABLES | | RF | | | | | | | | | |
| | | MVTS | | | | | STATES | | | | |
| | | ID | DA | PDR | PDRM | PDM | ID | DA | PDR | PRDM | PDM |
| NO PDA | D1 | 0.224 (0.009) | 0.785 (0.001) | 0.18 (0.006) | 0 (0) | **0.123 (0.053)** | **0.143 (0.003)** | 0.771 (0.001) | 0.157 (0.001) | 0 (0) | 0.034 (0.001) |
| | D2 | 0.257 (0.006) | 0.789 (0.001) | 0.151 (0.009) | 0 (0) | 0 (0) | 0.117 (0.003) | 0.773 (0.002) | 0.146 (0.002) | 0.014 (0.001) | 0.035 (0.001) |
| | D3 | 0.326 (0.005) | 0.796 (0.001) | **0.2 (0.009)** | 0 (0) | 0.064 (0.009) | 0.092 (0.003) | 0.764 (0.003) | 0.142 (0.004) | 0.012 (0.001) | **0.036 (0.003)** |
| | D1D2 | 0.302 (0.011) | 0.786 (0.001) | 0.148 (0.015) | 0 (0) | 0.101 (0.111) | 0.135 (0.004) | 0.774 (0.001) | 0.153 (0.008) | 0.013 (0.008) | 0.031 (0.002) |
| | D2D3 | 0.359 (0.004) | **0.797 (0.001)** | 0.197 (0.009) | 0 (0) | 0.033 (0.07) | 0.088 (0.003) | **0.779 (0.002)** | 0.147 (0.001) | 0.014 (0.001) | 0.04 (0.003) |
| | D1D2D3 | **0.373 (0.01)** | 0.794 (0.001) | 0.177 (0.01) | 0 (0) | 0 (0) | 0.089 (0.004) | 0.775 (0.002) | **0.161 (0.005)** | **0.019 (0.006)** | **0.036 (0.004)** |
| PDA | D1 | 0.234 (0.009) | 0.791 (0.001) | 0.18 (0.01) | 0 (0) | 0 (0) | **0.156 (0.005)** | 0.779 (0.002) | **0.174 (0.008)** | 0.015 (0.013) | 0.064 (0.004) |
| | D2 | 0.268 (0.005) | 0.795 (0.001) | 0.165 (0.01) | 0 (0) | 0 (0) | 0.135 (0.003) | 0.778 (0.001) | 0.167 (0.006) | **0.027 (0.017)** | **0.067 (0.003)** |
| | D3 | 0.335 (0.006) | **0.8 (0.001)** | **0.198 (0.005)** | 0 (0) | **0.145 (0.062)** | 0.123 (0.003) | 0.776 (0.002) | 0.16 (0.002) | 0.017 (0.006) | 0.063 (0.002) |
| | D1D2 | 0.307 (0.01) | 0.787 (0.001) | 0.155 (0.012) | 0 (0) | 0 (0) | 0.147 (0.003) | 0.775 (0.002) | 0.171 (0.003) | 0.013 (0.002) | 0.042 (0.002) |
| | D2D3 | 0.372 (0.007) | 0.799 (0.001) | **0.199 (0.011)** | 0 (0) | 0.02 (0.063) | 0.109 (0.003) | 0.786 (0.002) | 0.159 (0.002) | 0.009 (0.009) | 0.059 (0.003) |
| | D1D2D3 | **0.375 (0.011)** | 0.795 (0.001) | 0.169 (0.009) | 0 (0) | 0 (0) | 0.105 (0.006) | **0.789 (0.001)** | 0.165 (0.001) | 0.015 (0) | 0.041 (0.002) |
| RECALL | | | | | | | | | | | |
| INPUT VARIABLES | | RF | | | | | | | | | |
| | | MVTS | | | | | STATES | | | | |
| | | ID | DA | PDR | PDRM | PDM | ID | DA | PDR | PRDM | PDM |
| NO PDA | D1 | 0.137 (0.008) | 0.838 (0.002) | **0.134 (0.005)** | 0 (0) | **0.033 (0.016)** | 0.282 (0.016) | **0.753 (0.007)** | 0.175 (0.002) | 0 (0) | 0.169 (0.019) |
| | D2 | 0.251 (0.006) | 0.828 (0.003) | 0.069 (0.004) | 0 (0) | 0 (0) | 0.353 (0.02) | 0.611 (0.006) | 0.356 (0.011) | **0.105 (0)** | 0.245 (0.014) |
| | D3 | **0.365 (0.006)** | 0.85 (0.003) | 0.128 (0.005) | 0 (0) | 0.02 (0) | **0.388 (0.019)** | 0.542 (0.014) | 0.352 (0.01) | **0.105 (0)** | 0.241 (0.023) |
| | D1D2 | 0.212 (0.008) | 0.864 (0.002) | 0.053 (0.006) | 0 (0) | 0.018 (0.019) | 0.311 (0.03) | 0.707 (0.015) | 0.191 (0.018) | 0.047 (0.03) | 0.214 (0.019) |
| | D2D3 | 0.323 (0.006) | 0.868 (0.002) | 0.073 (0.003) | 0 (0) | 0.004 (0.008) | 0.325 (0.009) | 0.522 (0.014) | **0.388 (0.007)** | **0.105 (0)** | **0.275 (0.013)** |
| | D1D2D3 | 0.285 (0.011) | **0.888 (0.003)** | 0.054 (0.002) | 0 (0) | 0 (0) | 0.295 (0.014) | 0.648 (0.012) | 0.247 (0.02) | 0.074 (0.027) | 0.245 (0.019) |
| PDA | D1 | 0.158 (0.007) | 0.825 (0.003) | 0.132 (0.009) | 0 (0) | 0 (0) | **0.418 (0.015)** | **0.76 (0.012)** | 0.2 (0.013) | 0.032 (0.027) | **0.384 (0.025)** |
| | D2 | 0.295 (0.009) | 0.82 (0.003) | 0.069 (0.004) | 0 (0) | 0 (0) | 0.404 (0.015) | 0.695 (0.009) | **0.405 (0.013)** | 0.068 (0.043) | 0.314 (0.013) |
| | D3 | **0.392 (0.01)** | 0.842 (0.003) | **0.136 (0.005)** | 0 (0) | **0.018 (0.006)** | 0.451 (0.008) | 0.618 (0.005) | 0.364 (0.004) | 0.047 (0.017) | 0.312 (0.011) |
| | D1D2 | 0.222 (0.01) | 0.855 (0.003) | 0.058 (0.005) | 0 (0) | 0 (0) | 0.411 (0.015) | 0.688 (0.008) | 0.337 (0.007) | **0.158 (0.025)** | 0.294 (0.013) |

| | | | | | | | | | | | |
|---|---|---|---|---|---|---|---|---|---|---|---|
| | D2D3 | 0.353 (0.006) | 0.861 (0.003) | 0.076 (0.004) | 0 (0) | 0.002 (0.006) | 0.344 (0.011) | 0.606 (0.006) | 0.392 (0.007) | 0.026 (0.028) | 0.263 (0.017) |
| | D1D2D3 | 0.289 (0.011) | **0.882 (0.002)** | 0.053 (0.002) | 0 (0) | 0 (0) | 0.326 (0.023) | 0.623 (0.01) | 0.376 (0.01) | **0.158 (0)** | 0.29 (0.012) |

| F1-SCORE | | | | | | | | | | | |
|---|---|---|---|---|---|---|---|---|---|---|---|
| INPUT VARIABLES | | RF | | | | | | | | | |
| | | MVTS | | | | | STATES | | | | |
| | | ID | DA | PDR | PDRM | PDM | ID | DA | PDR | PRDM | PDM |
| NO PDA | D1 | 0.17 (0.009) | 0.811 (0.001) | 0.153 (0.005) | 0 (0) | **0.052 (0.025)** | 0.19 (0.003) | **0.762 (0.004)** | 0.166 (0.001) | 0 (0) | 0.056 (0.002) |
| | D2 | 0.254 (0.005) | 0.808 (0.002) | 0.094 (0.006) | 0 (0) | 0 (0) | 0.175 (0.005) | 0.683 (0.003) | 0.207 (0.003) | 0.024 (0.001) | 0.062 (0.002) |
| | D3 | **0.344 (0.004)** | 0.822 (0.001) | **0.156 (0.006)** | 0 (0) | 0.03 (0.001) | 0.149 (0.005) | 0.634 (0.009) | 0.203 (0.005) | 0.022 (0.001) | **0.063 (0.006)** |
| | D1D2 | 0.249 (0.009) | 0.823 (0.001) | 0.078 (0.008) | 0 (0) | 0.03 (0.033) | **0.188 (0.007)** | 0.739 (0.008) | 0.17 (0.01) | 0.021 (0.012) | 0.055 (0.004) |
| | D2D3 | **0.34 (0.005)** | 0.831 (0.001) | 0.107 (0.004) | 0 (0) | 0.007 (0.015) | 0.139 (0.004) | 0.625 (0.01) | **0.213 (0.002)** | **0.025 (0.001)** | 0.07 (0.005) |
| | D1D2D3 | 0.323 (0.011) | **0.838 (0.002)** | 0.083 (0.004) | 0 (0) | 0 (0) | 0.136 (0.005) | 0.706 (0.007) | 0.195 (0.01) | 0.03 (0.01) | **0.063 (0.007)** |
| PDA | D1 | 0.189 (0.008) | 0.808 (0.001) | 0.152 (0.009) | 0 (0) | 0 (0) | **0.227 (0.005)** | **0.769 (0.006)** | 0.186 (0.01) | 0.02 (0.017) | 0.11 (0.007) |
| | D2 | 0.281 (0.007) | 0.807 (0.002) | 0.098 (0.006) | 0 (0) | 0 (0) | 0.202 (0.005) | 0.734 (0.005) | 0.236 (0.008) | **0.038 (0.024)** | **0.111 (0.004)** |
| | D3 | **0.362 (0.008)** | 0.821 (0.002) | **0.161 (0.005)** | 0 (0) | **0.031 (0.011)** | 0.193 (0.004) | 0.688 (0.003) | 0.223 (0.002) | 0.025 (0.009) | 0.104 (0.004) |
| | D1D2 | 0.257 (0.01) | 0.82 (0.001) | 0.085 (0.007) | 0 (0) | 0 (0) | 0.216 (0.004) | 0.729 (0.004) | 0.227 (0.004) | 0.024 (0.003) | 0.074 (0.003) |
| | D2D3 | **0.362 (0.006)** | 0.829 (0.002) | 0.11 (0.005) | 0 (0) | 0.004 (0.011) | 0.166 (0.005) | 0.685 (0.004) | 0.226 (0.003) | 0.013 (0.014) | 0.096 (0.006) |
| | D1D2D3 | 0.326 (0.011) | **0.836 (0.001)** | 0.081 (0.003) | 0 (0) | 0 (0) | 0.158 (0.009) | 0.696 (0.006) | **0.229 (0.002)** | 0.027 (0.001) | 0.072 (0.003) |

**Supplementary Table 12A**. Weighted ROC-AUC for prediction of primary diagnosis at admission (PDA) in the *training and test* dataset

| INPUT VARIABLES | | MVTS | STATES |
|---|---|---|---|
| NO PDA | D1 | 0.74 (0) | 0.62 (0) |
| | D2 | 0.71 (0) | 0.63 (0.001) |
| | D3 | 0.69 (0) | 0.63 (0.001) |
| | D1D2 | 0.75 (0) | 0.64 (0) |
| | D2D3 | 0.73 (0) | 0.64 (0) |
| | D1D2D3 | **0.76 (0)** | **0.65 (0)** |

**Supplementary Table 12B**. Weighted ROC-AUC for prediction of primary diagnosis at admission (PDA) in the *hold-out validation* dataset

| INPUT VARIABLES | | RF | |
|---|---|---|---|
| | | MV TS | STATES |
| NO PDA | D1 | 0.732 (0) | 0.54 (0.001) |
| | D2 | 0.696 (0) | 0.532 (0.001) |
| | D3 | 0.686 (0) | 0.537 (0.001) |
| | D1D2 | 0.738 (0) | 0.537 (0.001) |
| | D2D3 | 0.715 (0) | 0.539 (0.001) |
| | D1D2D3 | **0.748 (0)** | **0.541 (0.001)** |

**Supplementary Table 13A**. Weighted precision, recall and F1-score for prediction of primary diagnosis at admission (PDA) in the *training and test* dataset

| INPUT VARIABLES | | RF | | RF | | RF | |
|---|---|---|---|---|---|---|---|
| | | Precision | | Recall | | F1-score | |
| | | MVTS | STATES | MVTS | STATES | MVTS | STATES |
| NO PDA | D1 | 0.29 (0.002) | 0.16 (0.001) | **0.39 (0.002)** | 0.49 (0.005) | 0.32 (0.002) | 0.24 (0.002) |
| | D2 | 0.28 (0.002) | 0.17 (0.001) | 0.36 (0.002) | 0.46 (0.002) | 0.31 (0.002) | 0.24 (0.001) |
| | D3 | 0.27 (0.002) | 0.17 (0.001) | 0.28 (0.002) | 0.46 (0.007) | 0.27 (0.002) | 0.24 (0.002) |
| | D1D2 | 0.32 (0.001) | 0.18 (0.002) | 0.37 (0.002) | 0.49 (0.004) | **0.34 (0.001)** | 0.25 (0.002) |
| | D2D3 | 0.33 (0.002) | 0.18 (0.001) | 0.31 (0.002) | 0.48 (0.003) | 0.31 (0.002) | 0.25 (0.001) |
| | D1D2D3 | **0.35 (0.003)** | **0.19 (0.001)** | 0.35 (0.002) | **0.5 (0.002)** | **0.34 (0.002)** | **0.26 (0.001)** |

**Supplementary Table 13B**. Weighted precision, recall and F1-score for prediction of primary diagnosis at admission (PDA) in the *hold-out validation* dataset

| INPUT VARIABLES | | RF | | RF | | RF | |
|---|---|---|---|---|---|---|---|
| | | Precision | | Recall | | F1-score | |
| | | MVTS | STATES | MVTS | STATES | MVTS | STATES |
| NO PDA | D1 | 0.29 (0.001) | **0.15 (0.003)** | **0.39 (0.001)** | 0.27 (0.007) | 0.32 (0.001) | 0.17 (0.004) |
| | D2 | 0.27 (0.002) | 0.12 (0.002) | 0.34 (0.002) | 0.3 (0.005) | 0.3 (0.002) | 0.16 (0.003) |
| | D3 | 0.27 (0.003) | 0.13 (0.002) | 0.28 (0.003) | 0.32 (0.007) | 0.27 (0.002) | 0.18 (0.003) |
| | D1D2 | 0.32 (0.001) | 0.14 (0.002) | 0.37 (0.001) | 0.26 (0.005) | **0.34 (0.001)** | 0.16 (0.002) |
| | D2D3 | 0.32 (0.002) | 0.13 (0.002) | 0.3 (0.001) | **0.32 (0.004)** | 0.3 (0.001) | **0.18 (0.002)** |
| | D1D2D3 | **0.35 (0.001)** | 0.14 (0.001) | 0.34 (0.002) | 0.27 (0.004) | 0.33 (0.001) | 0.17 (0.002) |

**Supplementary Figure 8A**. Precision, recall and F1-score for prediction of primary diagnosis at admission (PDA) by ICD-10 code in the *training and test dataset*. Dashed line is the half value of the highest metric

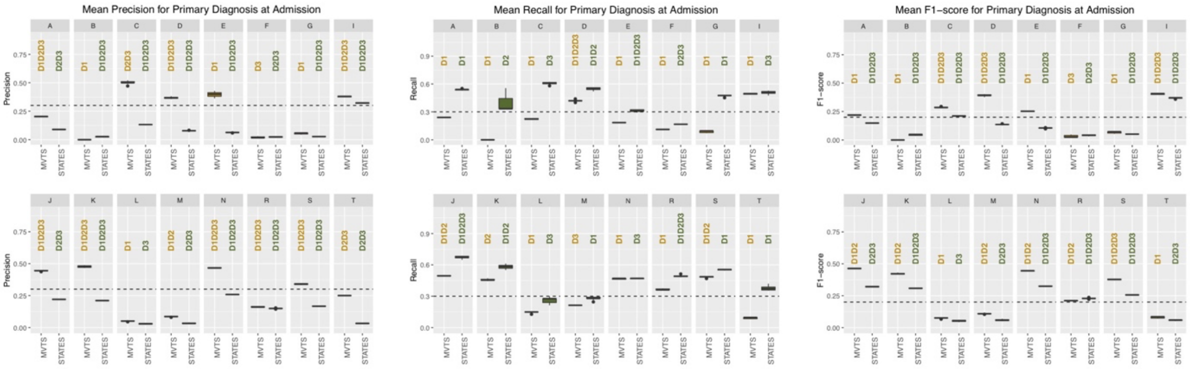


**Supplementary Figure 8B**. Precision, recall and F1-score for prediction of primary diagnosis at admission (PDA) by ICD-10 code in the *hold-out validation dataset*. Dashed line is the half value of the highest metric

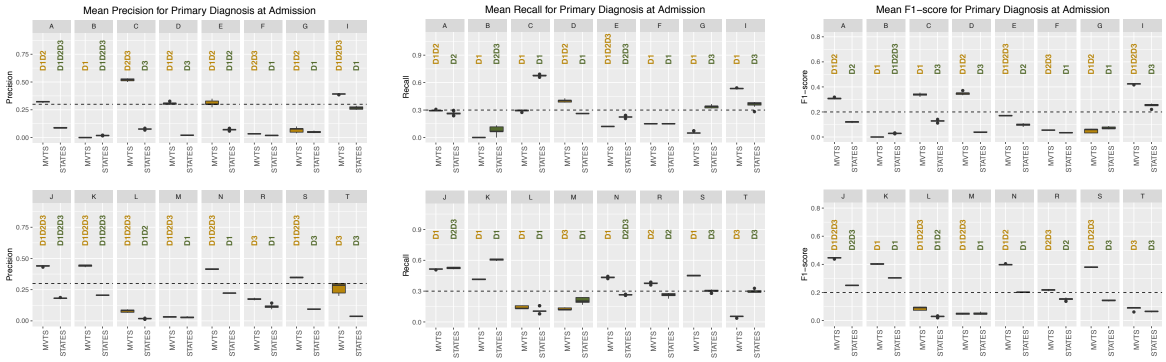

**Supplementary Table 14A**. Weighted ROC-AUC for prediction of diagnosis at discharge (DD) in the *training and test* dataset

| INPUT VARIABLES | | RF | |
|---|---|---|---|
| | | MVTS | STATES |
| NO PDA | D1 | 0.63 (0) | 0.58 (0) |
| | D2 | 0.63 (0) | 0.59 (0) |
| | D3 | 0.62 (0) | 0.59 (0.001) |
| | D1D2 | 0.64 (0) | 0.59 (0) |
| | D2D3 | 0.64 (0) | **0.6 (0)** |
| | D1D2D3 | **0.65 (0)** | **0.6 (0)** |

**Supplementary Table 14B**. Weighted ROC-AUC for prediction of diagnosis at discharge (DD) in the *hold-out validation* dataset

| INPUT VARIABLES | | RF | |
|---|---|---|---|
| | | MVTS | STATES |
| NO PDA | D1 | 0.732 (0) | 0.54 (0.001) |
| | D2 | 0.696 (0) | 0.532 (0.001) |
| | D3 | 0.686 (0) | 0.537 (0.001) |
| | D1D2 | 0.738 (0) | 0.537 (0.001) |
| | D2D3 | 0.715 (0) | 0.539 (0.001) |
| | D1D2D3 | **0.748 (0)** | **0.541 (0.001)** |

**Supplementary Table 15A**. Weighted precision, recall and F1-score for prediction of diagnosis at discharge (DD) in the *training and test* dataset

| INPUT VARIABLES | | RF | | RF | | RF | |
|---|---|---|---|---|---|---|---|
| | | Precision | | Recall | | F1-score | |
| | | MVTS | MV TS | MVTS | STATES | MVTS | STATES |
| NO PDA | D1 | 0.4 (0.001) | 0.32 (0.001) | **0.38 (0.001)** | 0.47 (0.003) | 0.38 (0.001) | 0.36 (0.001) |
| | D2 | 0.38 (0.001) | 0.33 (0.001) | **0.38 (0.001)** | **0.48 (0.002)** | 0.38 (0.001) | 0.37 (0.001) |
| | D3 | 0.38 (0.001) | 0.33 (0.001) | 0.37 (0.001) | 0.47 (0.003) | 0.37 (0.001) | 0.37 (0.001) |
| | D1D2 | **0.42 (0.001)** | **0.34 (0.001)** | 0.37 (0.001) | 0.47 (0.002) | **0.39 (0.001)** | 0.37 (0.001) |
| | D2D3 | 0.4 (0.001) | **0.34 (0)** | 0.37 (0.001) | 0.47 (0.002) | 0.38 (0.001) | 0.37 (0.001) |
| | D1D2D3 | **0.42 (0.001)** | **0.34 (0.001)** | 0.36 (0.001) | 0.47 (0.002) | 0.38 (0.001) | **0.38 (0.001)** |

**Supplementary Table 15B**. Weighted precision, recall and F1-score for prediction of diagnosis at discharge (DD) in the *hold-out validation* dataset

| INPUT VARIABLES | | RF | | RF | | RF | |
|---|---|---|---|---|---|---|---|
| | | Precision | | Recall | | F1-score | |
| | | MVTS | MV TS | MVTS | STATES | MVTS | STATES |
| NO PDA | D1 | 0.291 (0.001) | 0.151 (0.003) | **0.386 (0.001)** | 0.271 (0.007) | 0.323 (0.001) | 0.165 (0.004) |
| | D2 | 0.265 (0.002) | 0.115 (0.002) | 0.338 (0.002) | 0.298 (0.005) | 0.29 (0.002) | 0.159 (0.003) |
| | D3 | 0.274 (0.003) | 0.125 (0.002) | 0.281 (0.003) | **0.315 (0.007)** | 0.271 (0.002) | 0.175 (0.003) |
| | D1D2 | 0.324 (0.001) | 0.141 (0.002) | 0.369 (0.001) | 0.26 (0.005) | **0.336 (0.001)** | 0.161 (0.002) |
| | D2D3 | 0.319 (0.002) | 0.127 (0.002) | 0.299 (0.001) | **0.317 (0.004)** | 0.298 (0.001) | **0.176 (0.002)** |
| | D1D2D3 | **0.346 (0.001)** | **0.139 (0.001)** | 0.335 (0.002) | 0.273 (0.004) | 0.332 (0.001) | 0.17 (0.002) |

**Supplementary Figure 9A**. Precision, recall and F1-score for prediction of diagnosis at discharge (DD) by ICD-10 code in the *training and test* dataset. Dashed line is the half value of the highest metric

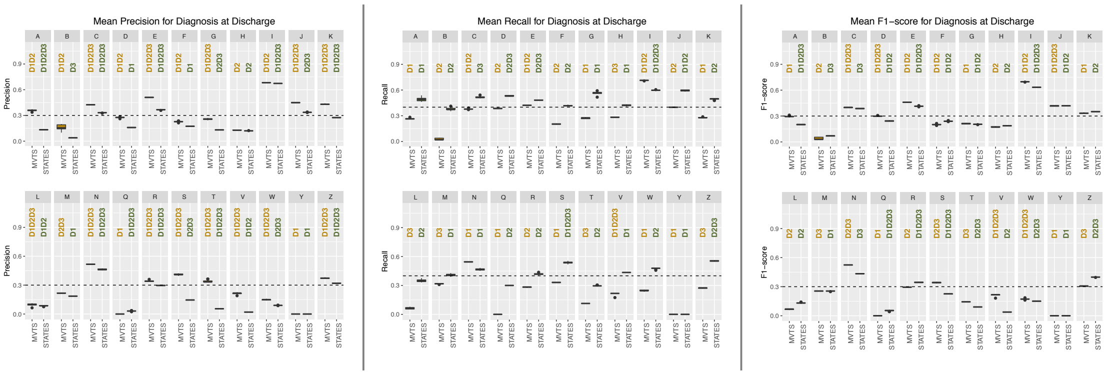


**Supplementary Figure 9B**. Precision, recall and F1-score for prediction of diagnosis at discharge (DD) by ICD-10 code in the *hold-out validation* dataset. Dashed line is the half value of the highest metric

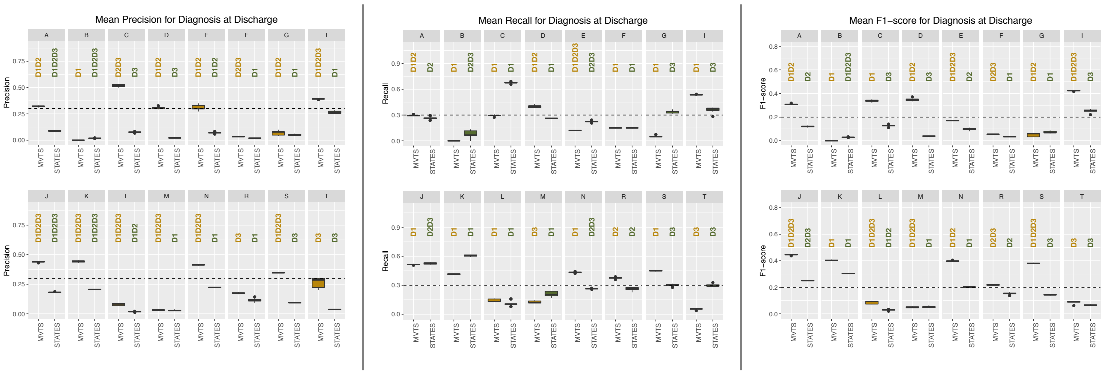
