## Supplementary Figures 4 for "Big Data Analysis of Electronic Health Records: Clinically interpretable representations of older adult inpatient trajectories using time-series numerical data and Hidden Markov Models"

### INFLUENCE OF IMPUTED VALUES BY STATES AND VARIABLES

### INTRODUCTION

- Most common variables with missing (not recorded) observations are glucose, liver function tests (alp, alt, bilirubin), urea and respiratory rate (compared to other vital signs).
- The fraction of imputed values by linear interpolation (LI) is larger than by multiple imputation (MI) because the days imputed by MI can only have a maximum of five missing variables (4 lab tests and 1 vital sign)

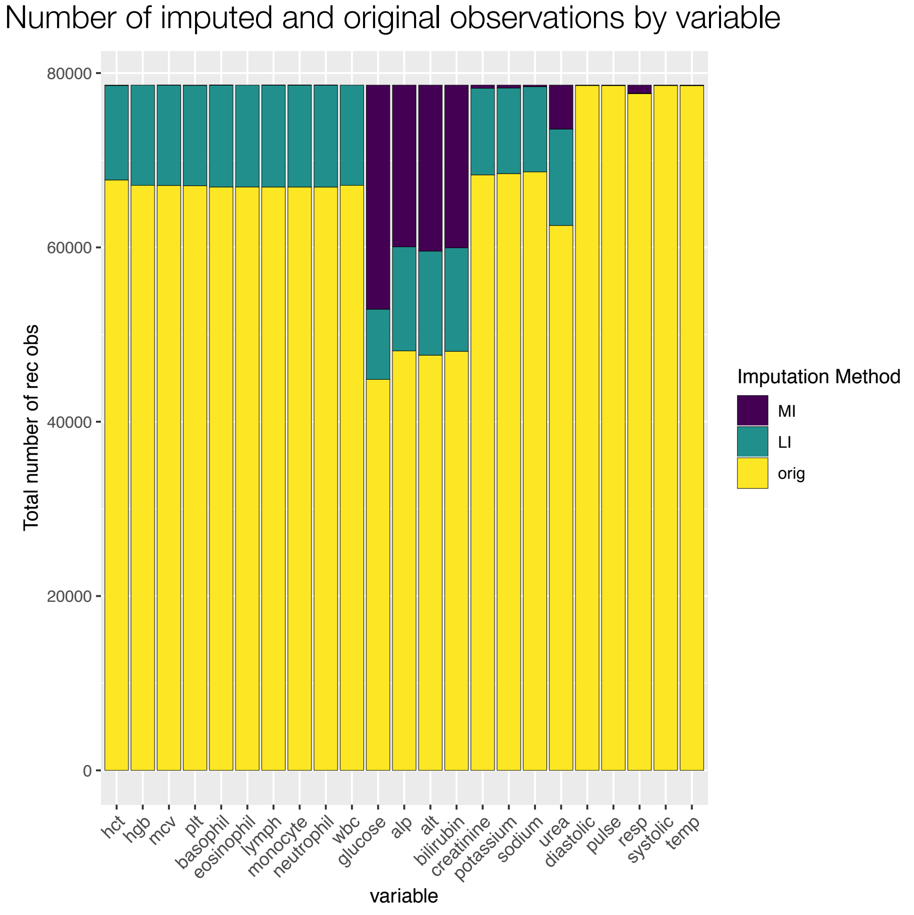


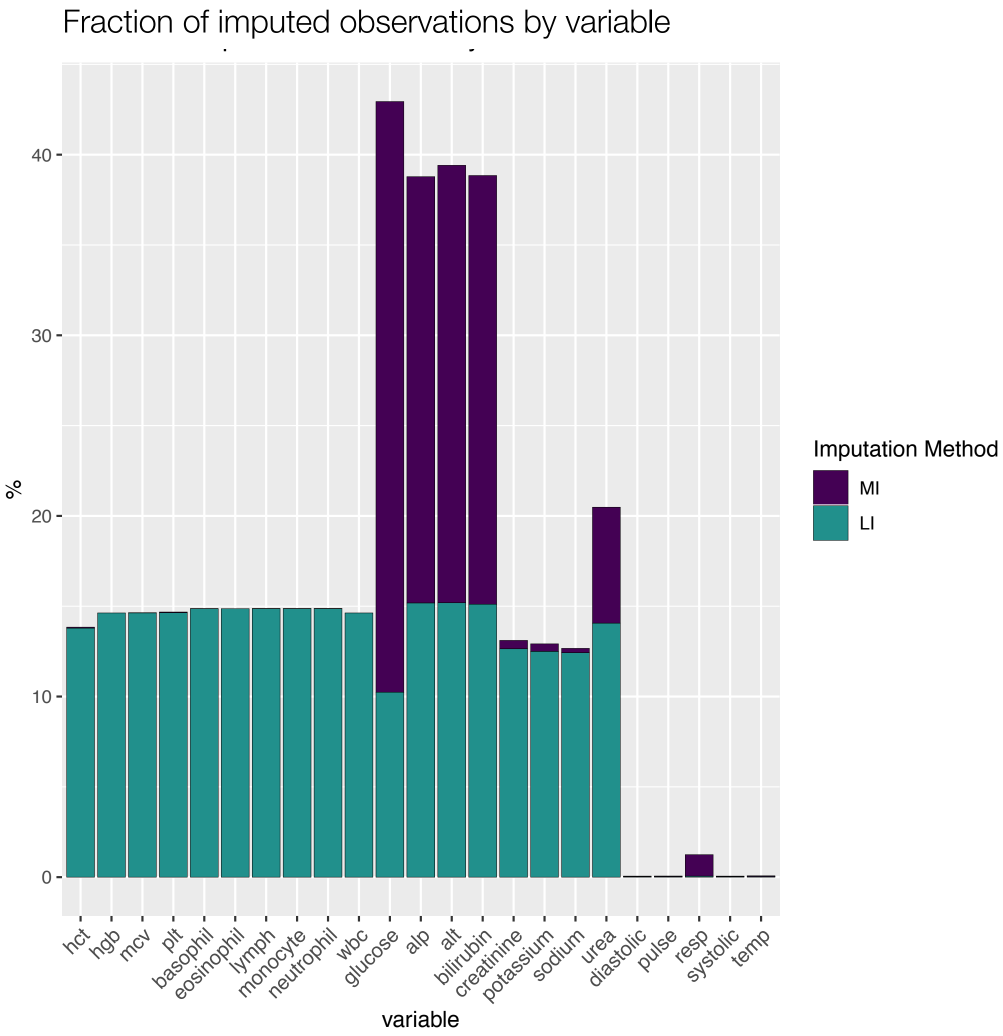

### INTRODUCTION

- Clinical visual interpretation of distribution of lab tests and vital signs by states is conducted independently of information on missingness (top-left plot).

- Distribution of orginal vs imputed values are compared (bottom-left plot)

- Distributions by imputation method (MI, LI or original value) are presented (bottom-centre plot). To illustrate the potential impact of the imputed values on the results for that state, the number of imputed values for each variable by imputation method are shown (bottom-right plot).

- A comparison of the *total number* (i.e., for all variables) of recorded observations (both original and imputed) by state are shown (top-right plot). Note that each variable has the same number of observations by state. The number of recorded observations for one specific variable is therefore the *total number* for that state divided by 23 (i.e., number of variables)

### DISEASE-LIKE STATES

- Hepatic
- Stable renal
- Unstable renal
- Stable but static renal
- Blood dyscrasia
- Bone marrow suppression

### HEPATIC-LIKE STATE

Distribution of values for this state

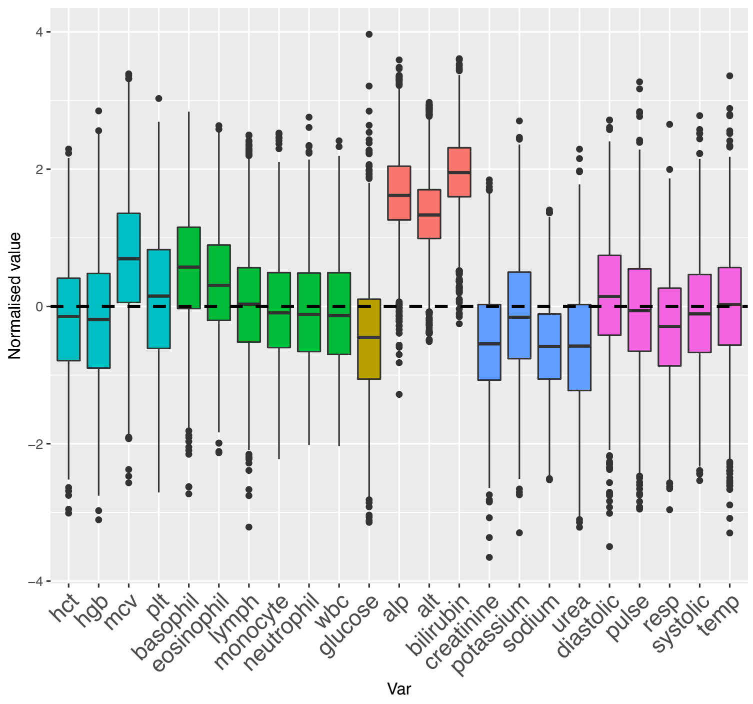


#### Clinical visual interpretation:

- **Very high**: alt, alp and bilirubin;
- **High**: mcv;
- **Lower**: urea, glucose, creatinine, sodium

Number of imputed and original observations by state
(current state highlighted with a green dot)

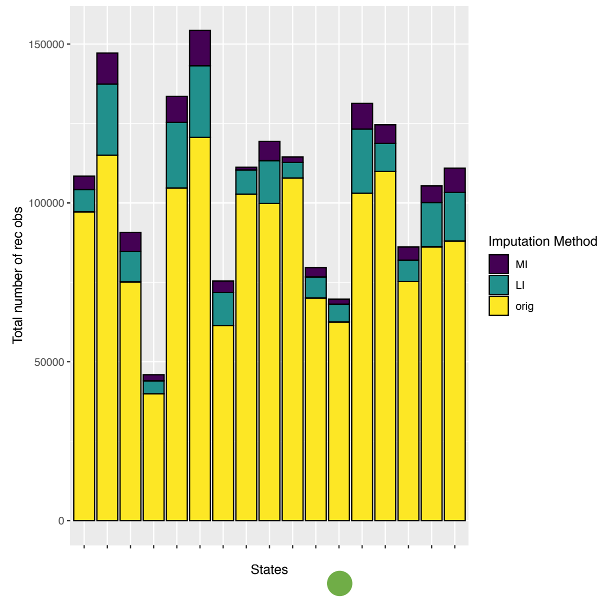


Distribution of imputed vs original values

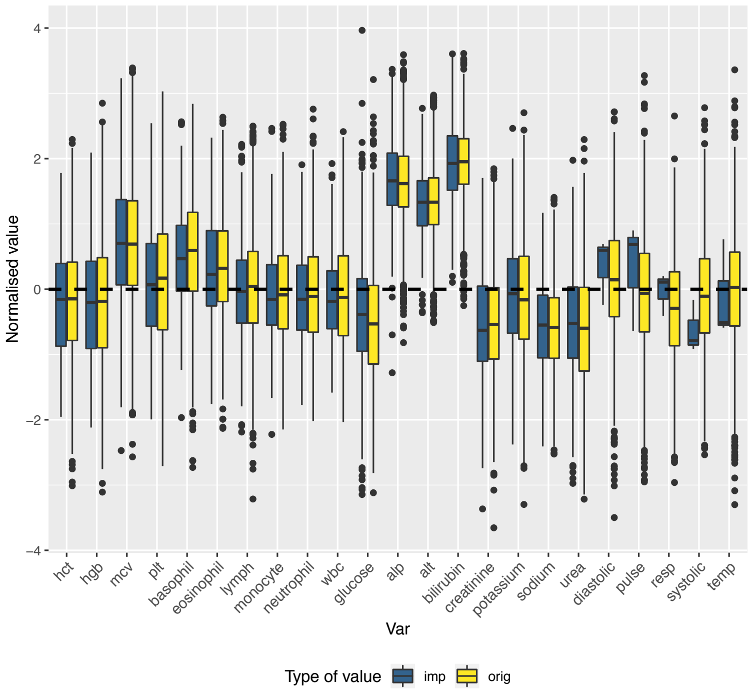


Distribution of original vs imputed values by imputation method

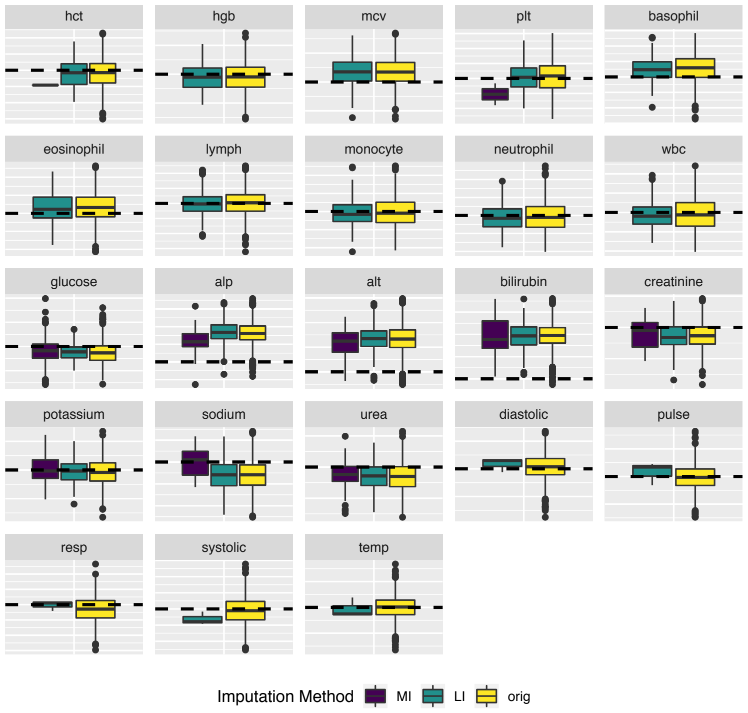


Number of imputed and original observations

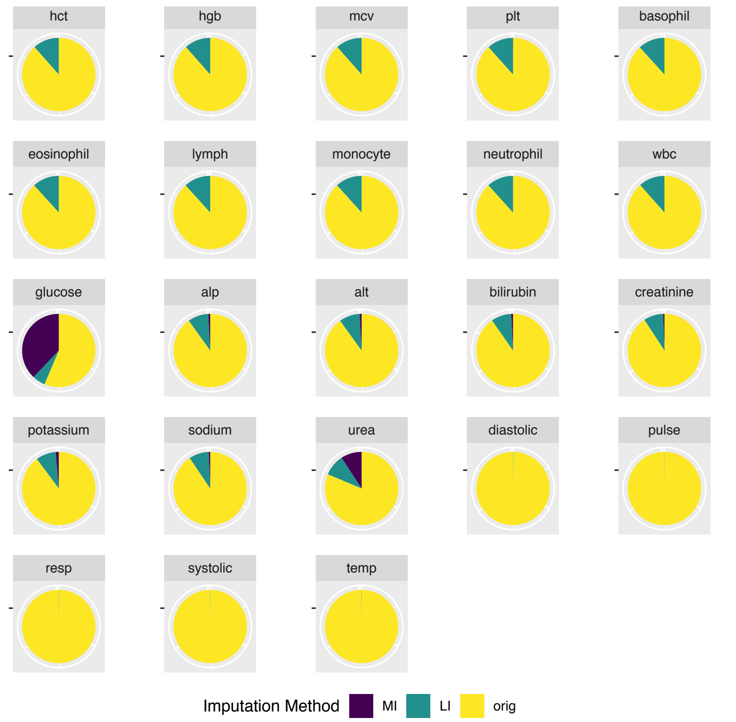

### STABLE RENAL-LIKE STATE

Distribution of values for this state

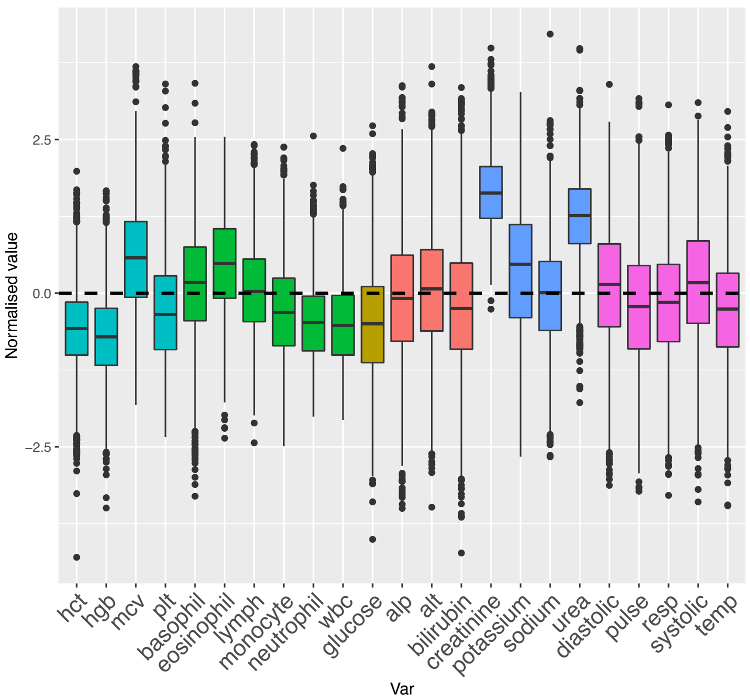


#### Clinical visual interpretation:

- **Very high**: creatinine, urea (with small variance);
- **High**: potassium, eosinophils, mcv;
- **Low**: hgb, hct, wbc, neutrophils, glucose;
- Liver Function Tests and Vital Signs all **near group mean or slightly lower** (i.e., maybe closer to healthy population mean)

Number of imputed and original observations by state
(current state highlighted with a green dot)

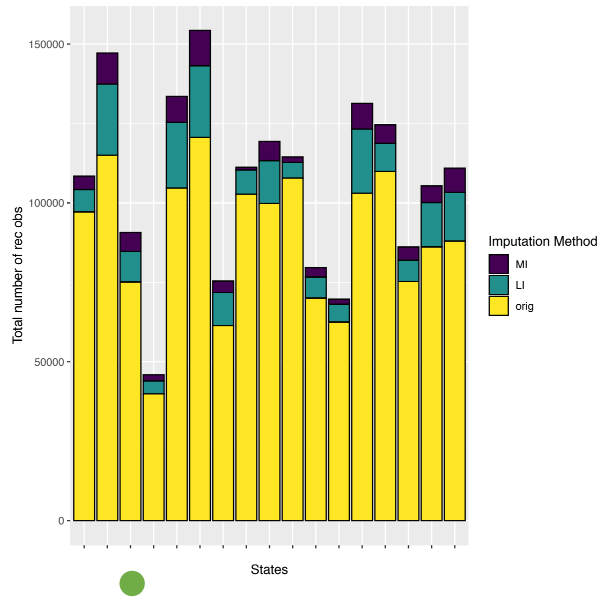


Distribution of imputed vs original values

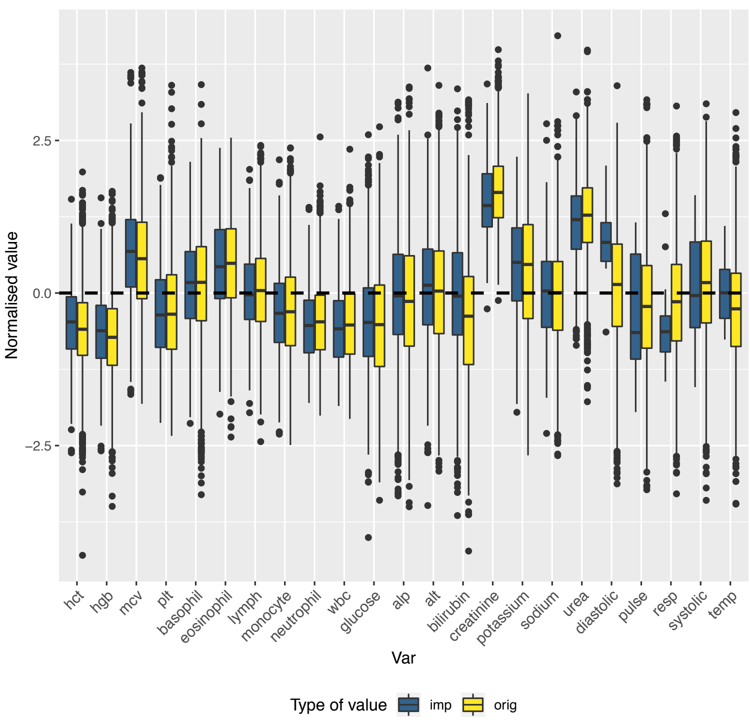


Distribution of original vs imputed values by imputation method

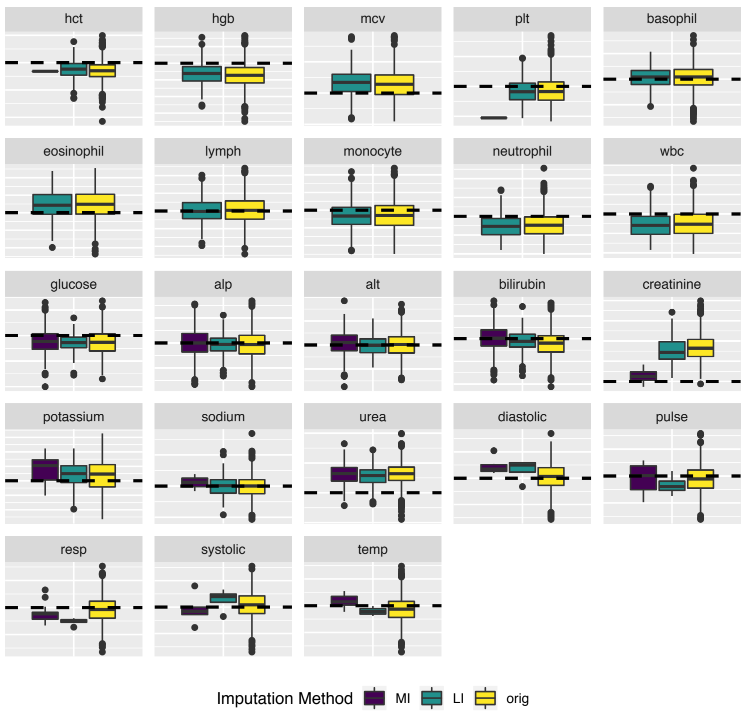


Number of imputed and original observations

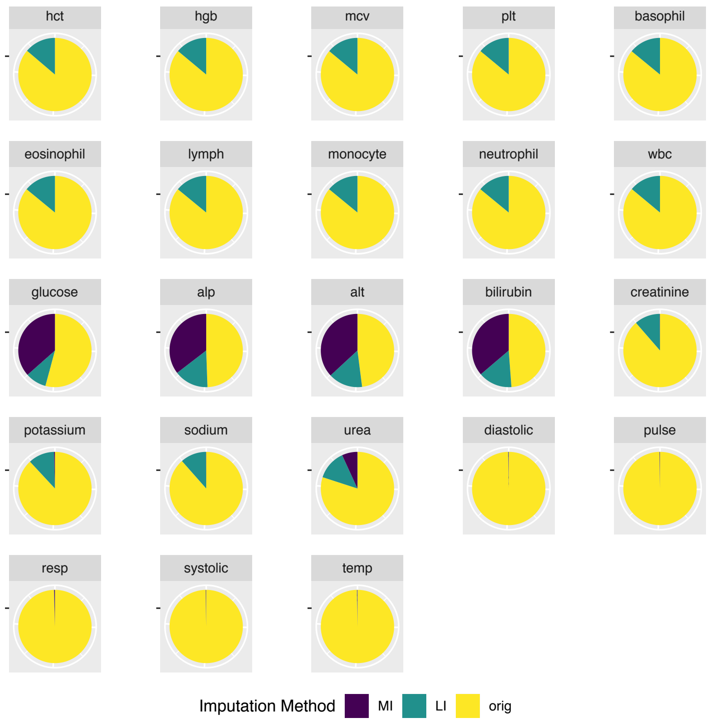

### UNSTABLE RENAL-LIKE STATE

Distribution of values for this state

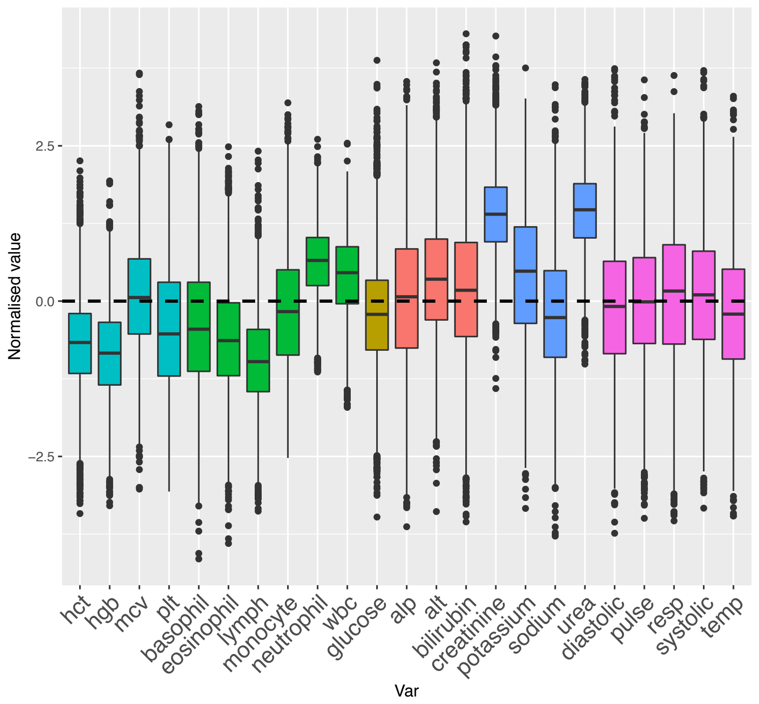


#### Clinical visual interpretation:

- **Very high**: urea, creatinine (with low variance), potassium;
- **Higher**: wbc, neutrophils;
- **Lower**: hgb, hct, lymphs;
- Liver Function Tests and Vital Signs all **around group mean or** on **higher** side of group mean

Number of imputed and original observations by state
(current state highlighted with a green dot)

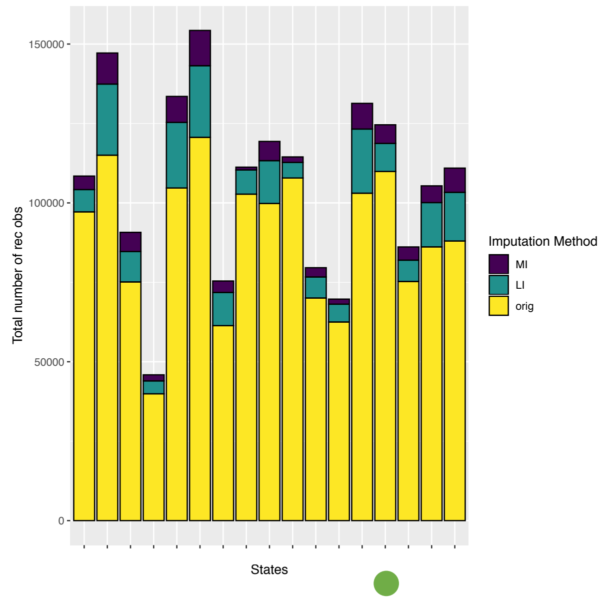


Distribution of imputed vs original values

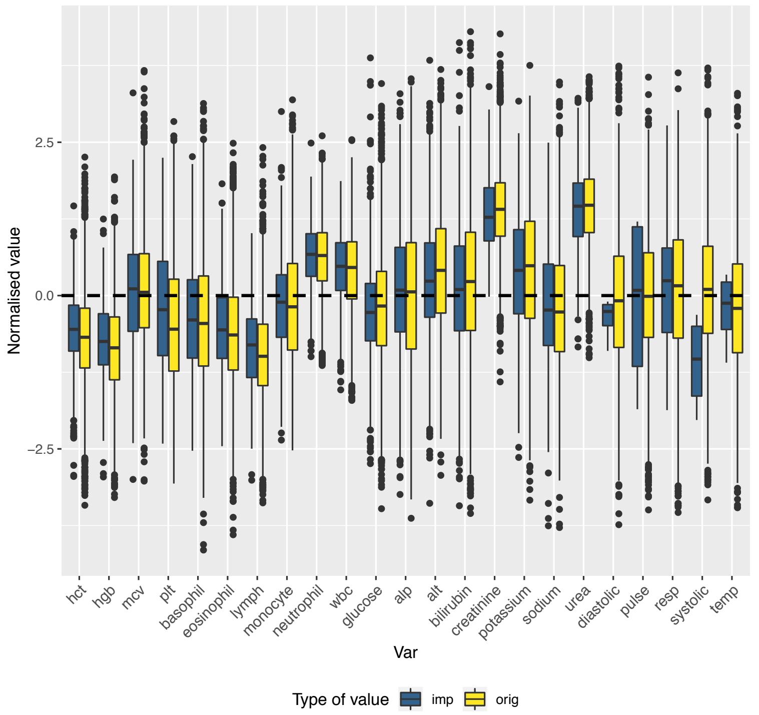


Distribution of original vs imputed values by imputation method

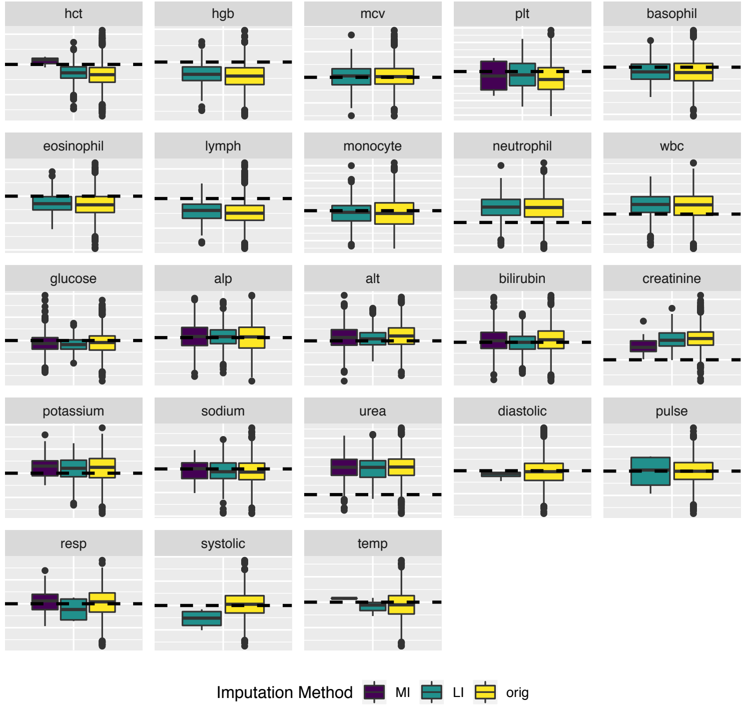


Number of imputed and original observations

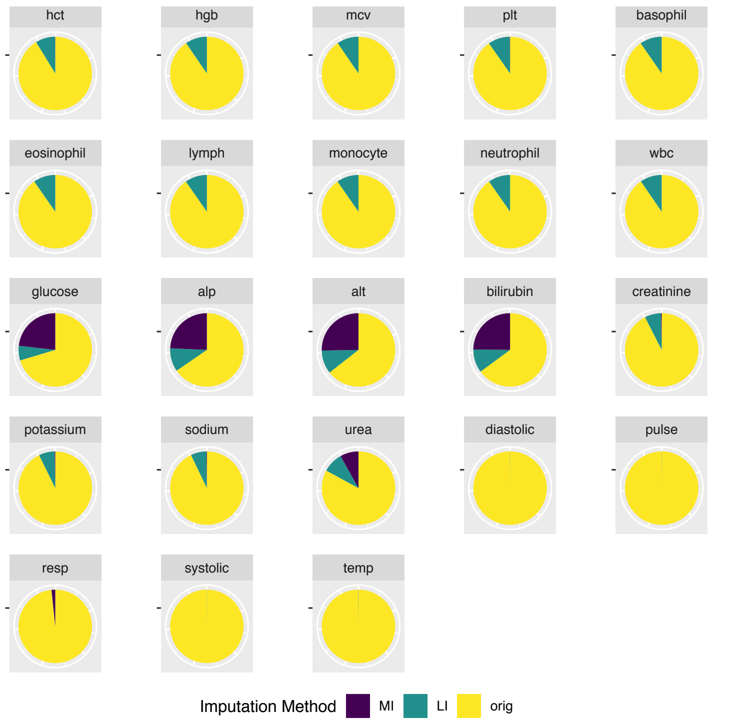

### STABLE BUT STATIC RENAL-LIKE STATE

Distribution of values for this state

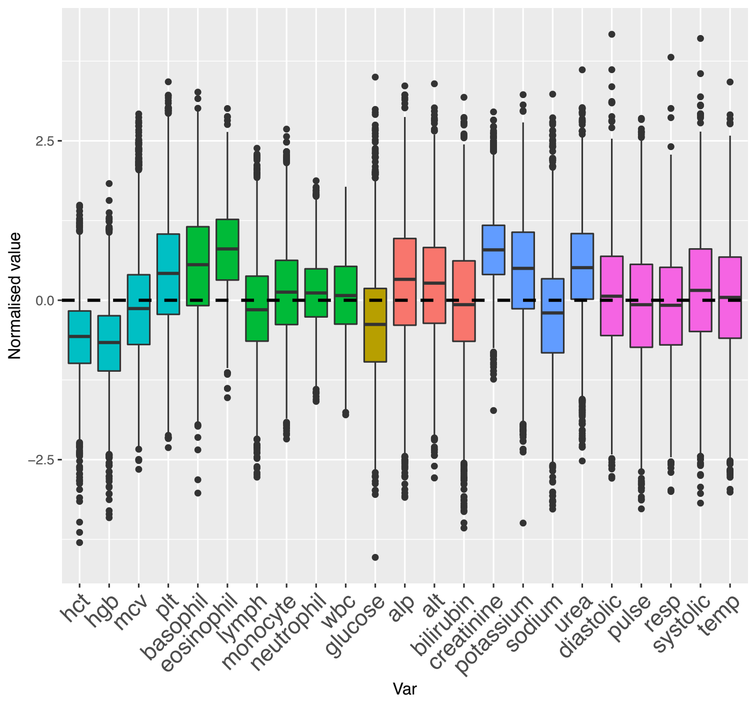


#### Clinical visual interpretation:

- **Lower**: hgb, hct, glucose;
- **Higher**: eosonophils, creatinine, urea, potassium;
- **‘Normal’**: Vital signs, white blood cells

Number of imputed and original observations by state
(current state highlighted with a green dot)

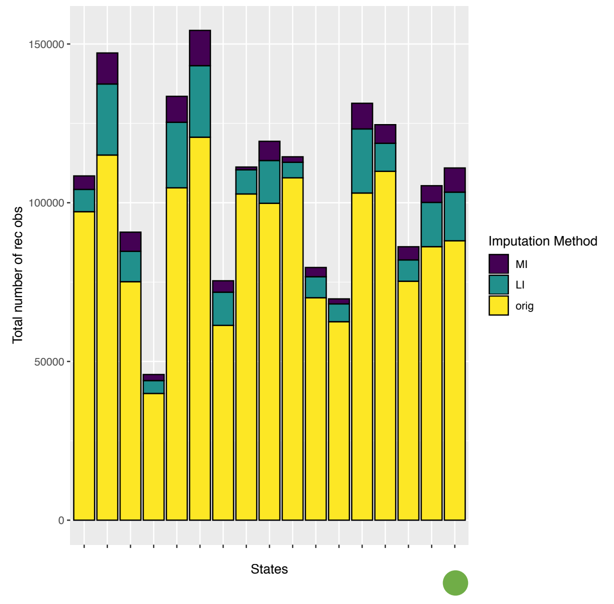


Distribution of imputed vs original values

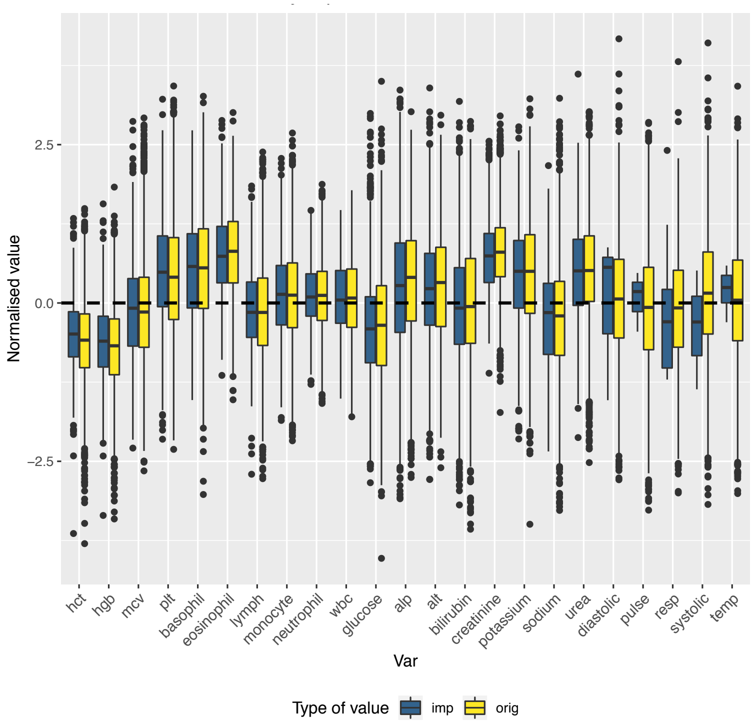


Distribution of original vs imputed values by imputation method

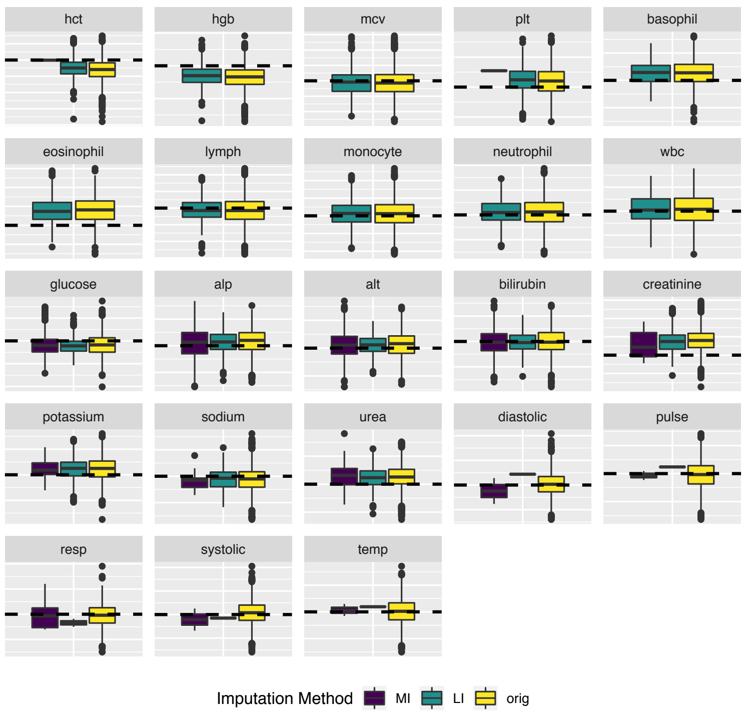


Number of imputed and original observations

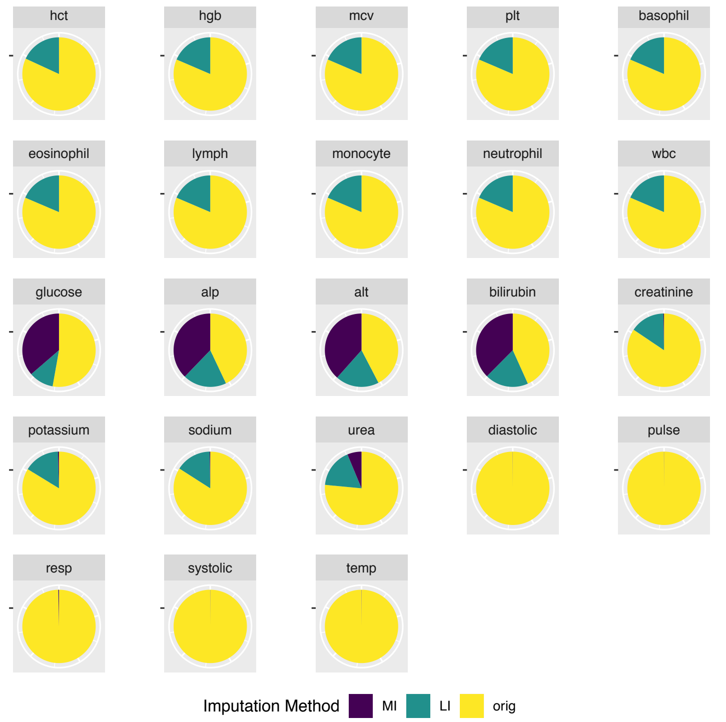

### BLOOD DYSCRASIA-LIKE STATE

Distribution of values for this state

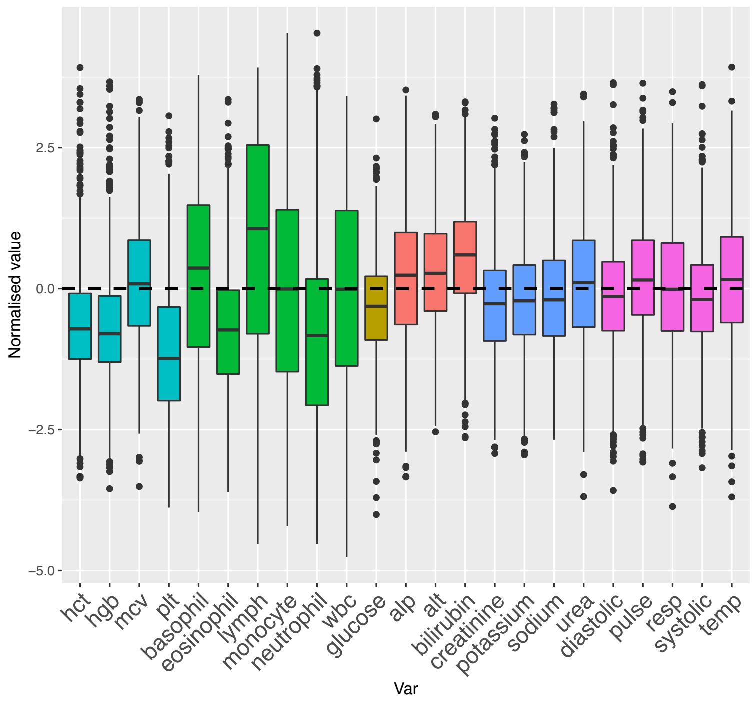


#### Clinical visual interpretation:

- **Lower**: hgb, hct, plts;
- **Deranged** wbc in both directions with wide variance;
- **Higher**: bilirubin;
- Other Liver and Renal Function Tests and Vital Signs all **very close to group mean**

Number of imputed and original observations by state
(current state highlighted with a green dot)

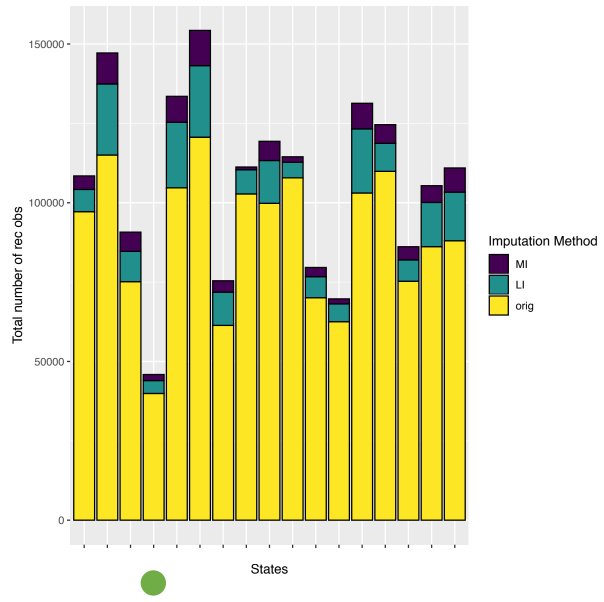


Distribution of imputed vs original values

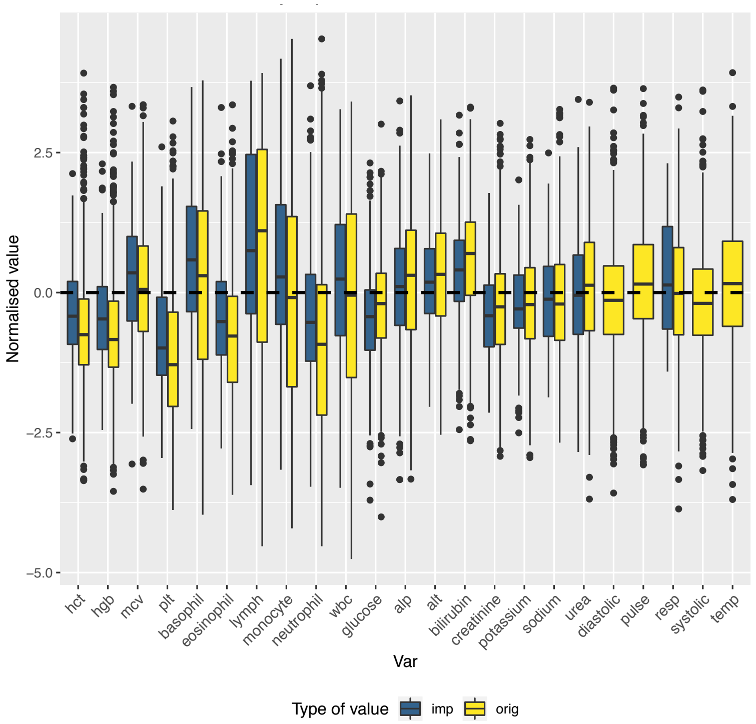


Distribution of original vs imputed values by imputation method

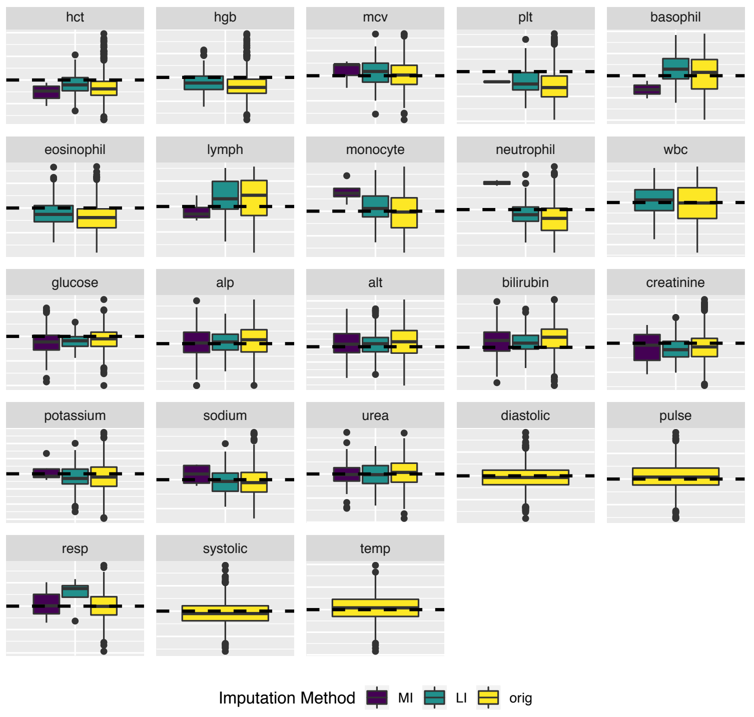


Number of imputed and original observations

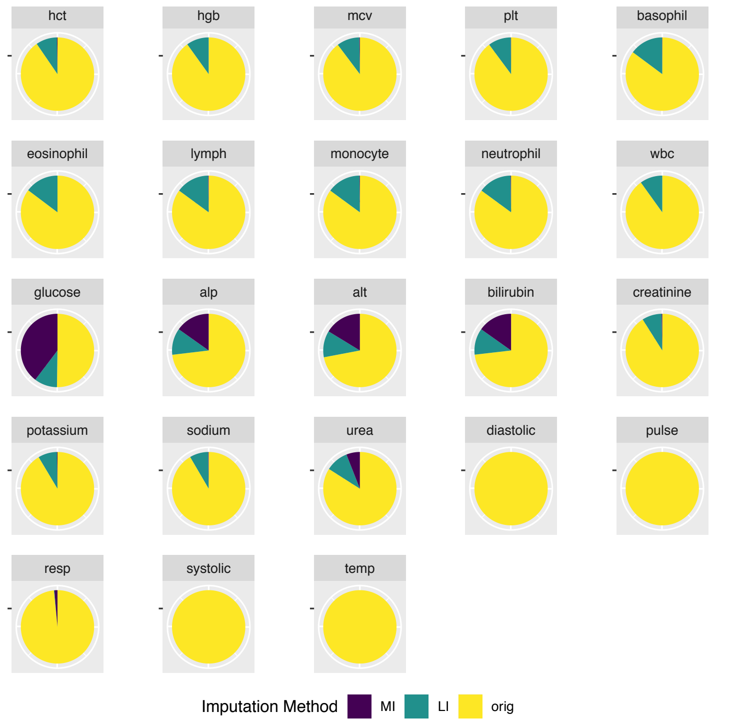

### BONE MARROW SUPPRESSION-LIKE STATE

Distribution of values for this state

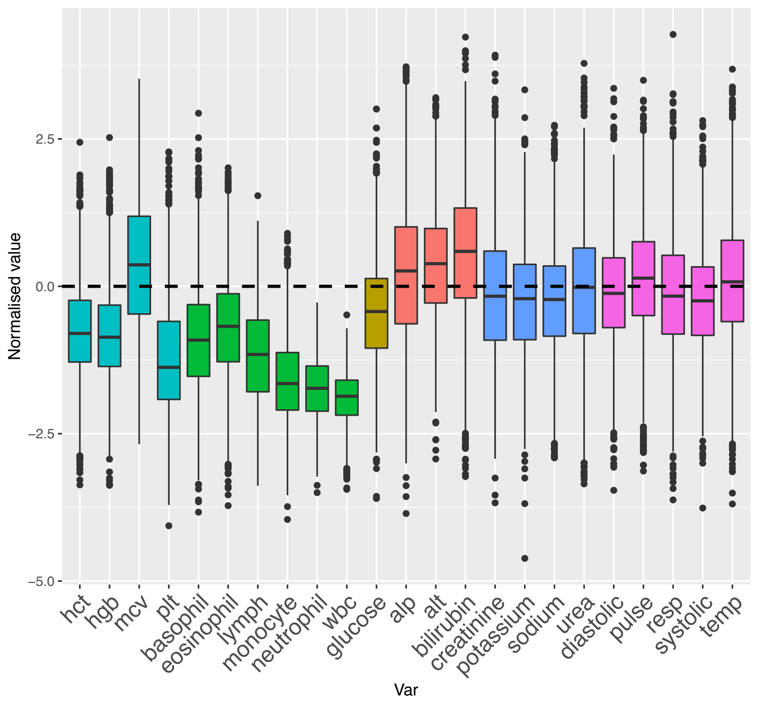


#### Clinical visual interpretation:

- ‘*Pancytopenia*’: all blood count* parameters **lower** except MCV which is **higher**;
- **Mildly high**: Liver Function Tests* (especially bilirubin);
- **Slightly lower**: glucose*;
- Renal Function Tests and Vital Signs just under group mean but generally unremarkable

Number of imputed and original observations by state
(current state highlighted with a green dot)

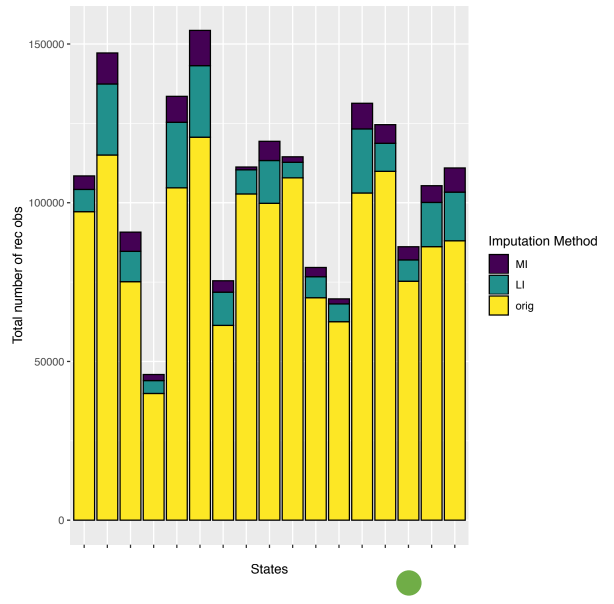


Distribution of imputed vs original values

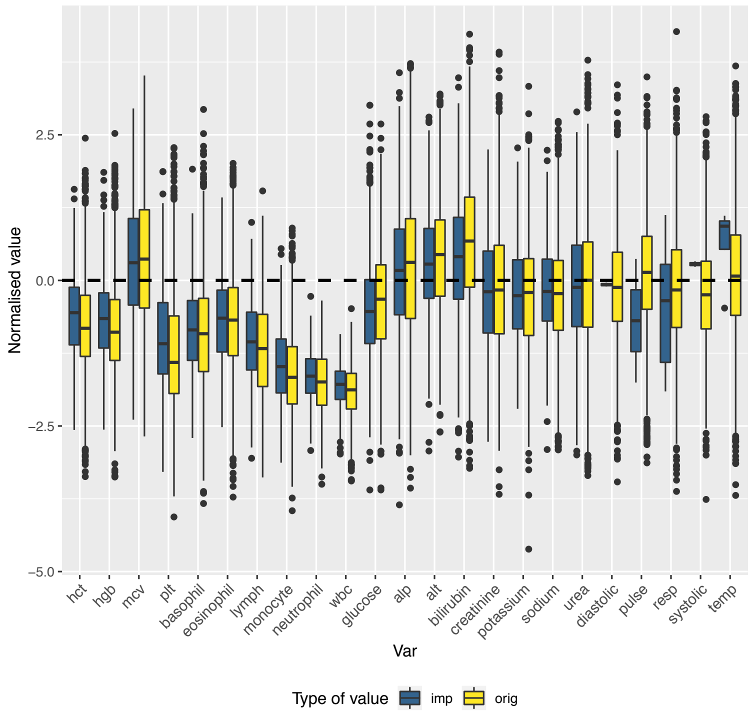


Distribution of original vs imputed values by imputation method

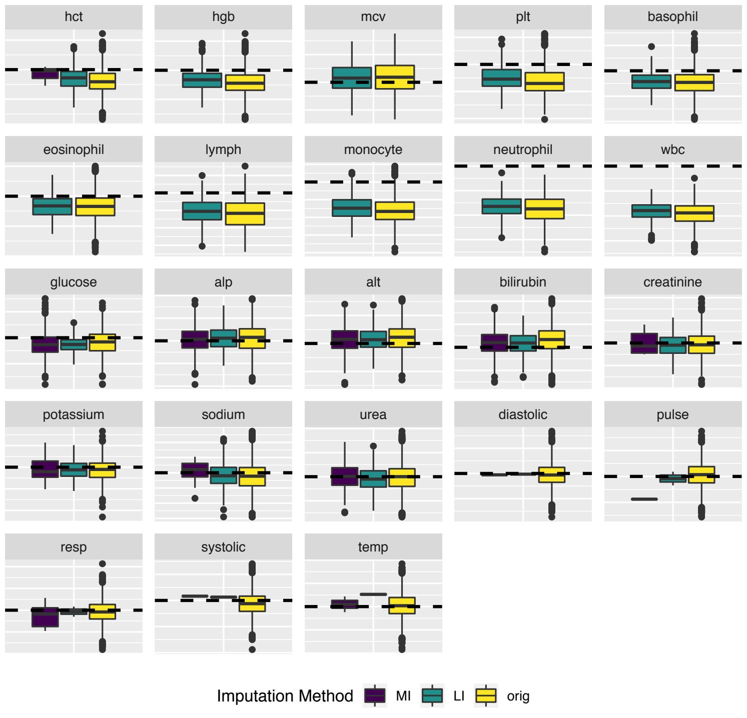


Number of imputed and original observations

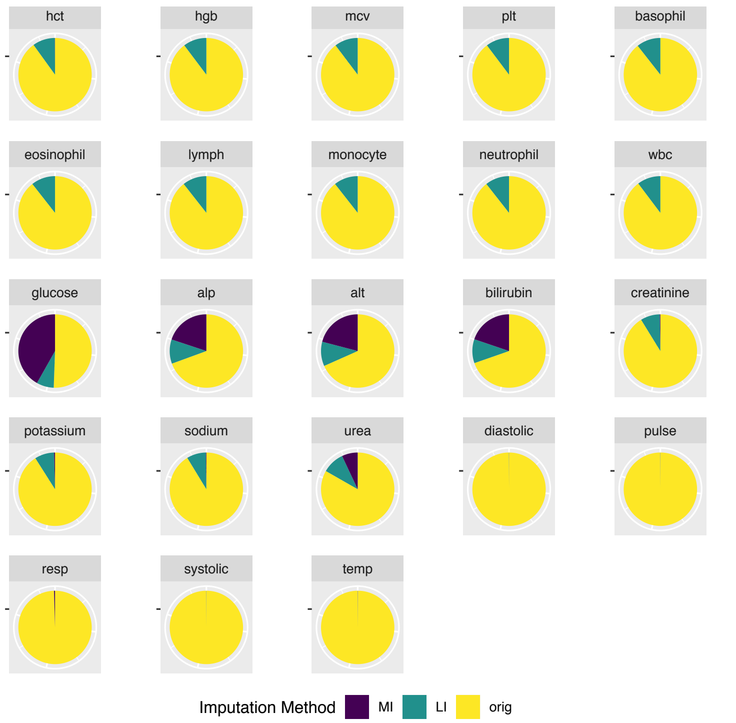

### ADMISSION STAGE-LIKE STATES

- Acute presentation
- Treatment response (1)
- Treatment response (2)
- Early discharge
- Pre-discharge

### ACUTE PRESENTATION-LIKE STATE

Distribution of values for this state

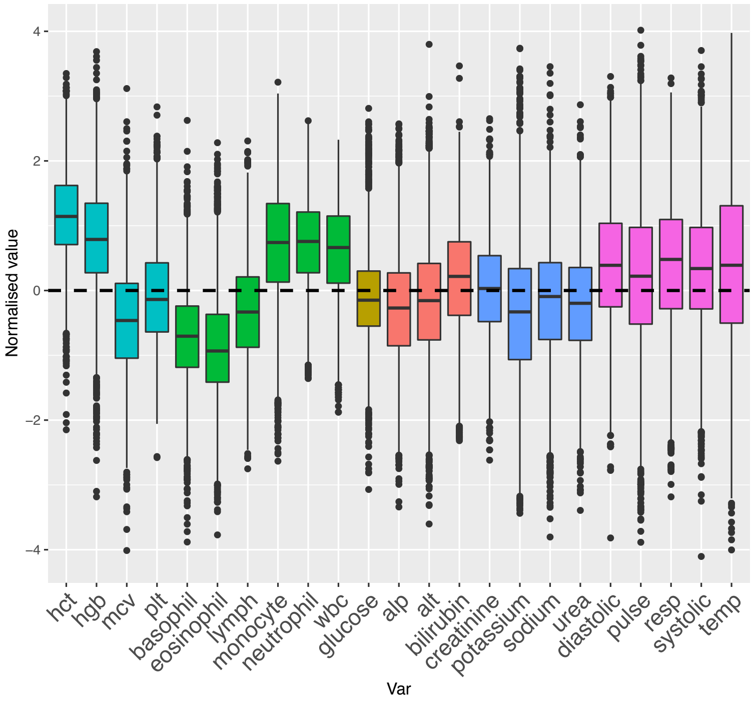


#### Clinical visual interpretation:

- **Higher**: hct, hgb, wbc, neutrophils, monocytes;
- **Slightly higher**: Vital signs;
- **Lower:** basophils, eosinophils, mcv

Number of imputed and original observations by state
(current state highlighted with a green dot)

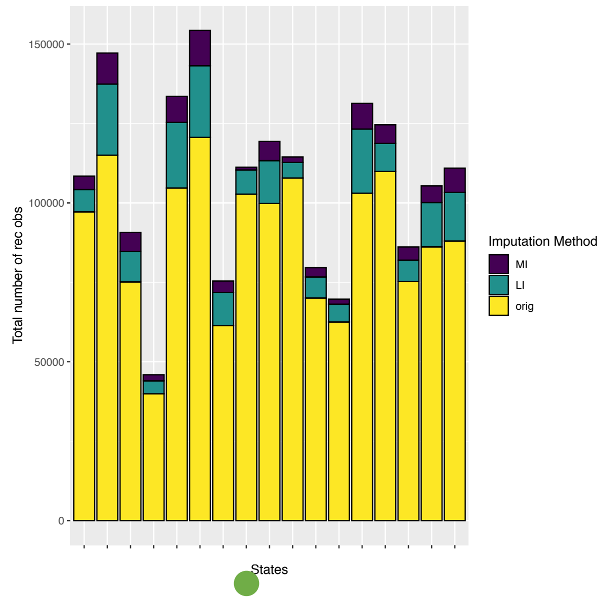


Distribution of imputed vs original values

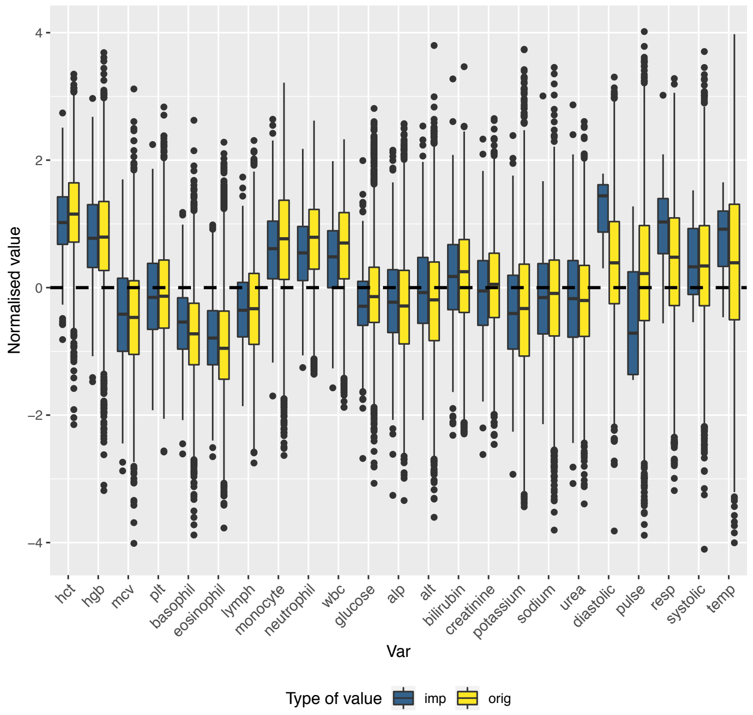


Distribution of original vs imputed values by imputation method

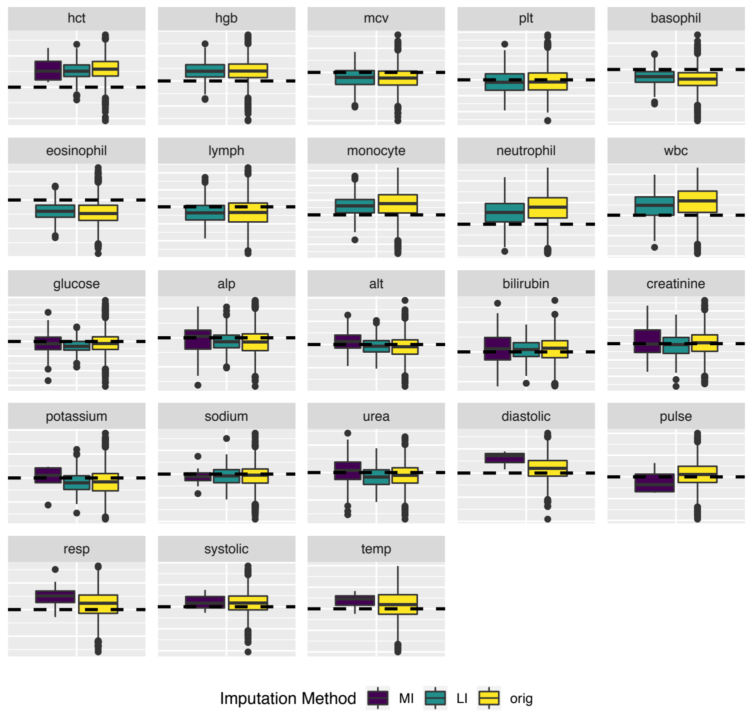


Number of imputed and original observations

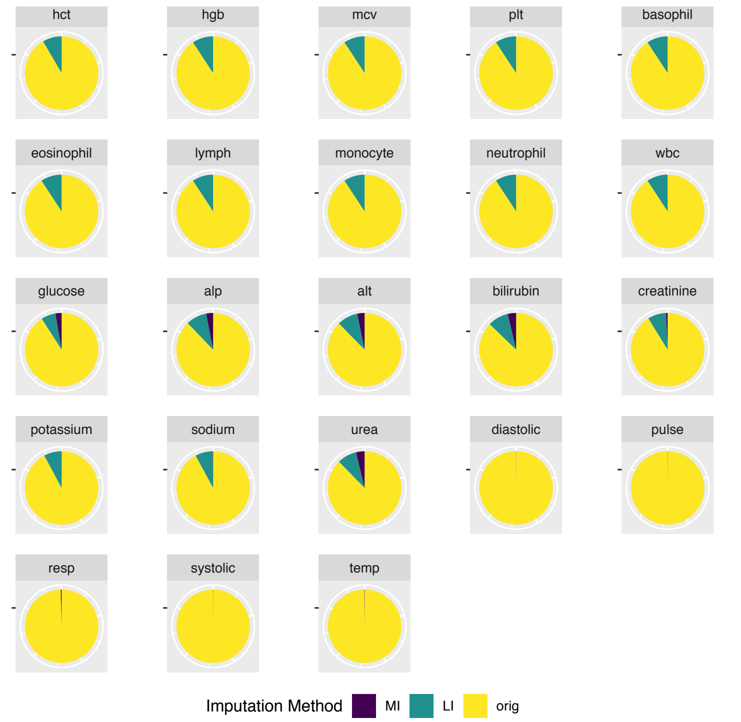

### TREATMENT RESPONSE-LIKE (1) STATE

Distribution of values for this state

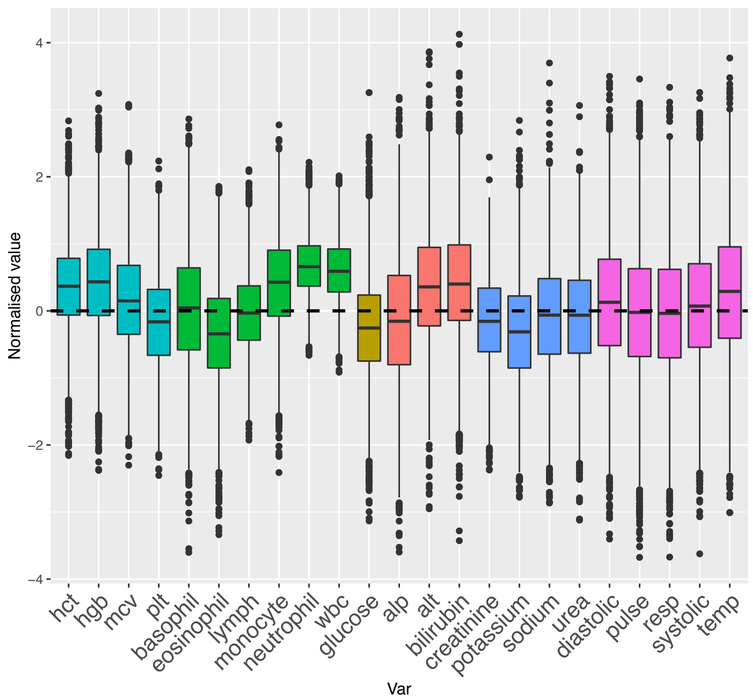


#### Clinical visual interpretation:

- **Higher**: hct, hgb, wbc, neutrophils, monocytes;
- **Slightly higher**: alt, bilirubin.

Number of imputed and original observations by state
(current state highlighted with a green dot)

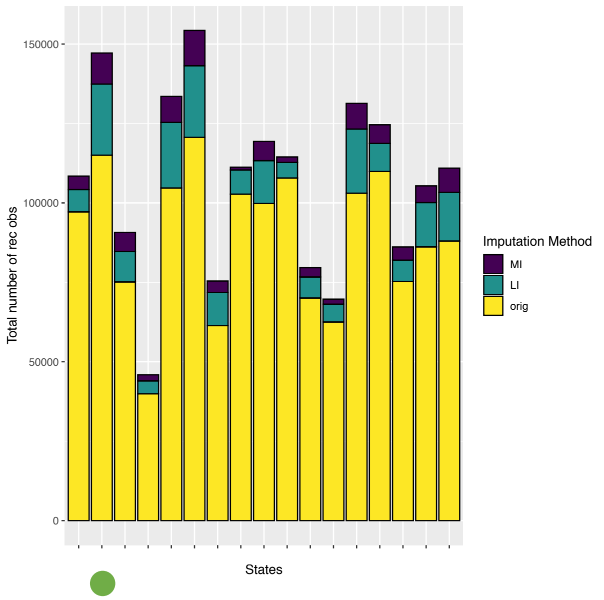


Distribution of imputed vs original values

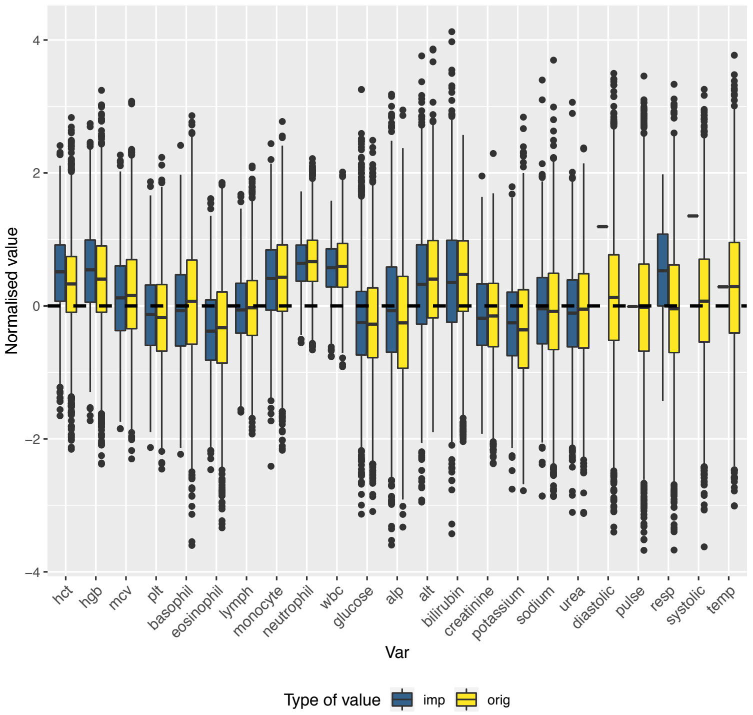


Distribution of original vs imputed values by imputation method

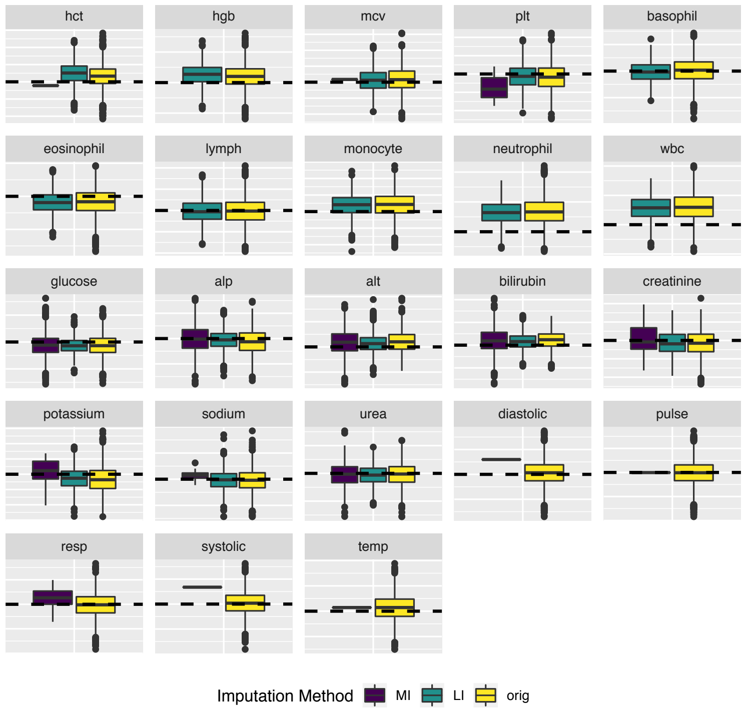


Number of imputed and original observations

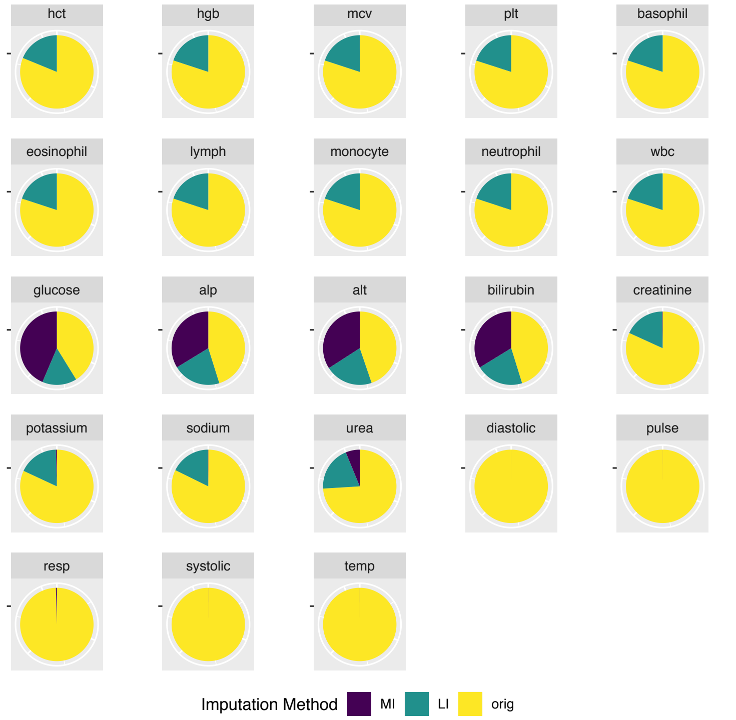

### TREATMENT RESPONSE-LIKE (2) STATE

Distribution of values for this state

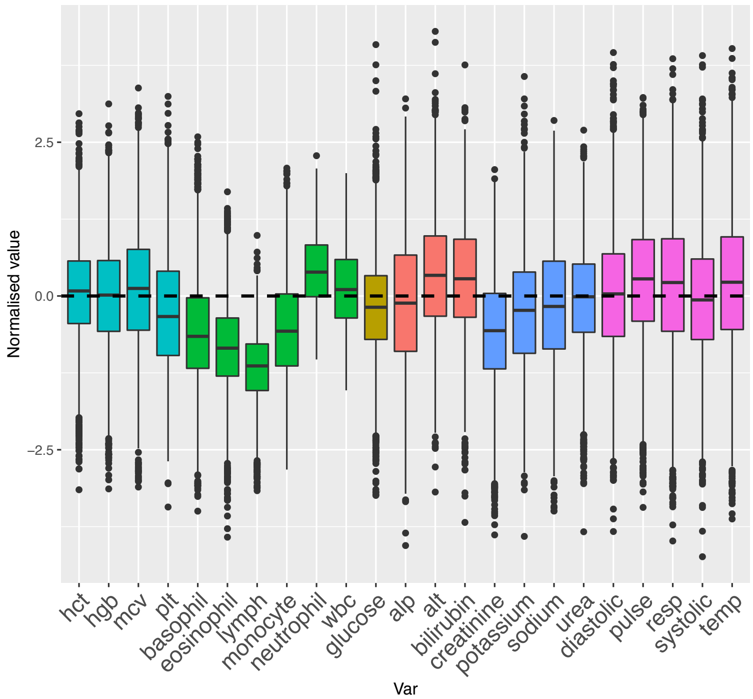


#### Clinical visual interpretation:

- **Lower**: basophils, monocytes, eosinophils, lypmhs, creatinine;
- **Higher**: neutrophils;
- **Slightly higher**: pulse, resp, temp, alt, bilirubin

Number of imputed and original observations by state
(current state highlighted with a green dot)

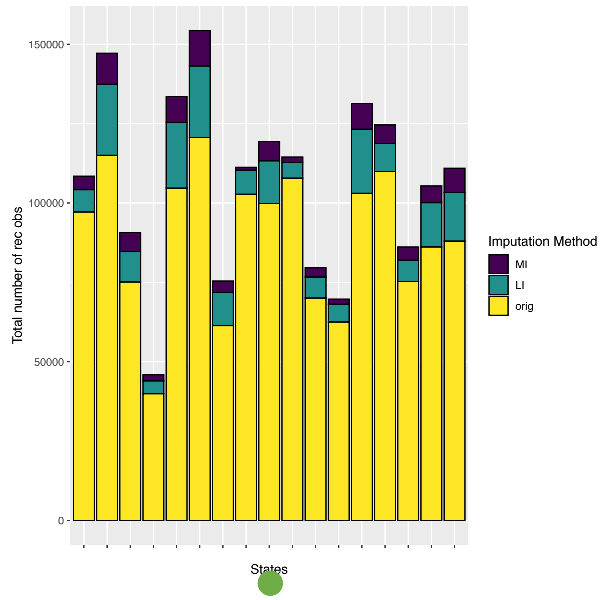


Distribution of imputed vs original values

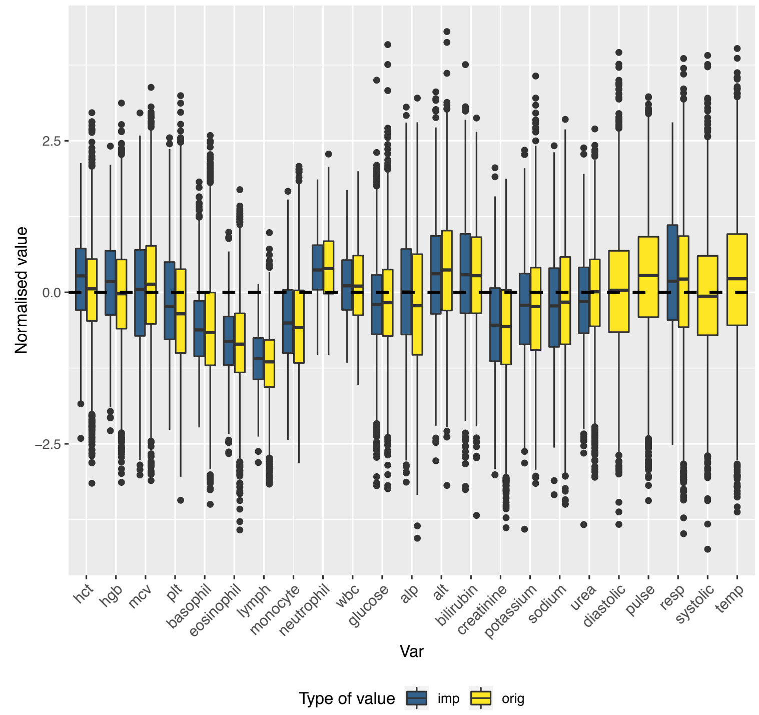


Distribution of original vs imputed values by imputation method

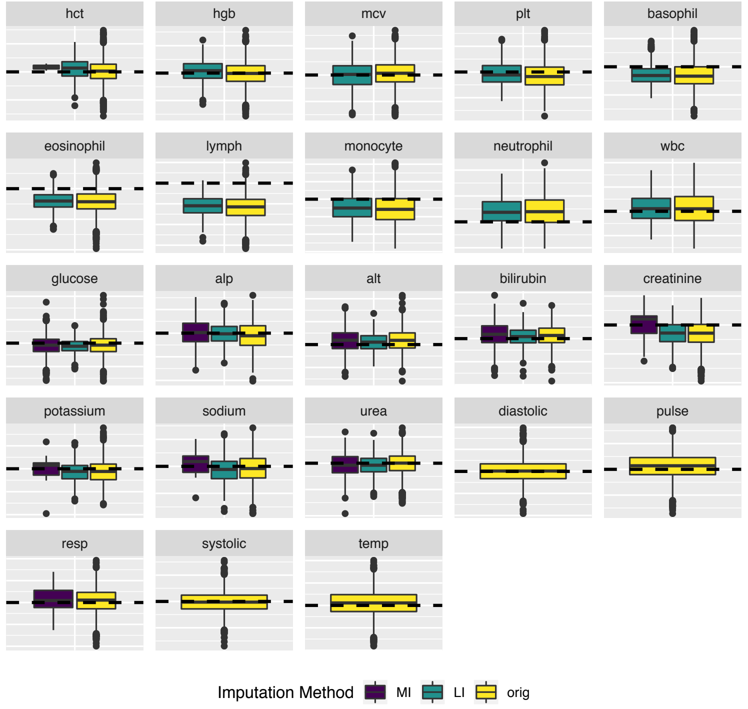


Number of imputed and original observations

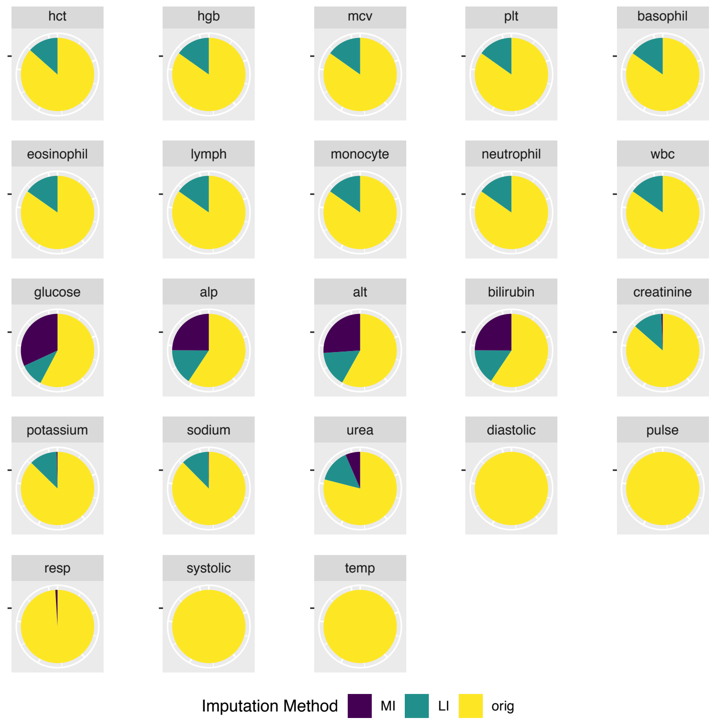

### EARLY DISCHARGE-LIKE STATE

Distribution of values for this state

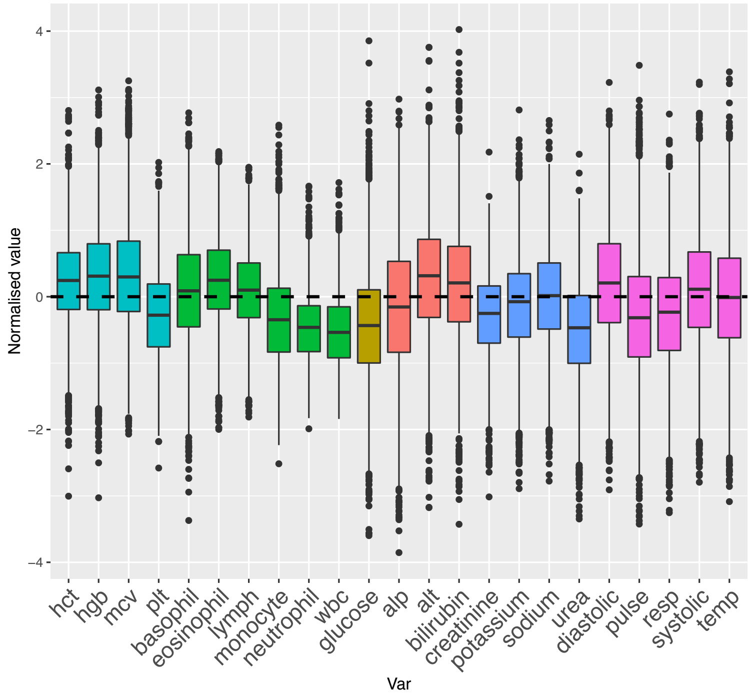


#### Clinical visual interpretation:

- All parameters near group mean with some **slightly higher**: hgb, hct, alt, bilirubin;
- **Slightly lower**: wbc, neutrophil, monocytes, glucose, urea, pulse, resp;
- In most cases the parameters that are slightly higher or lower probably indicate the values are nearer a 'healthy population' norm than our 'unwell population norm'.

Number of imputed and original observations by state
(current state highlighted with a green dot)

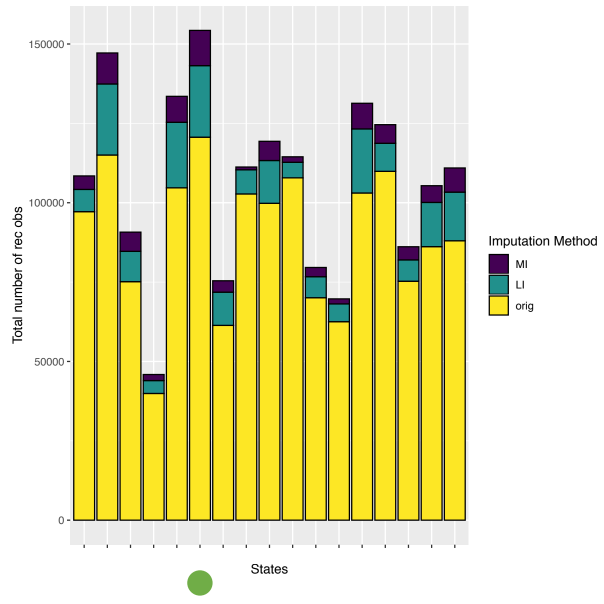


Distribution of imputed vs original values

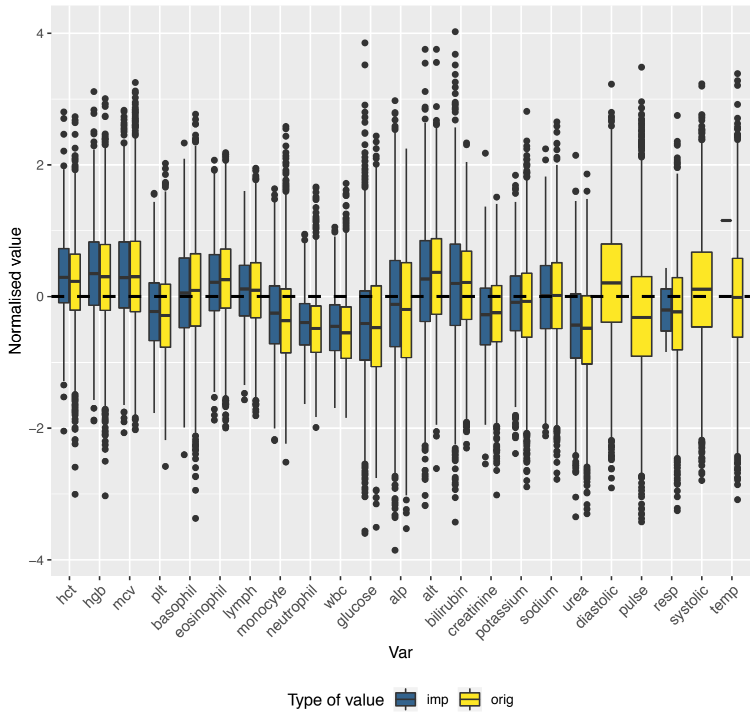


Distribution of original vs imputed values by imputation method

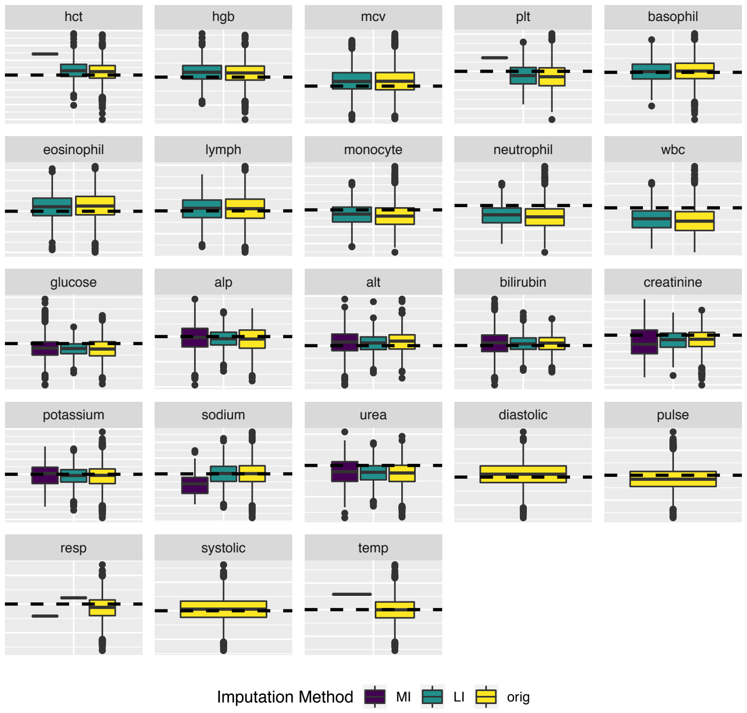


Number of imputed and original observations

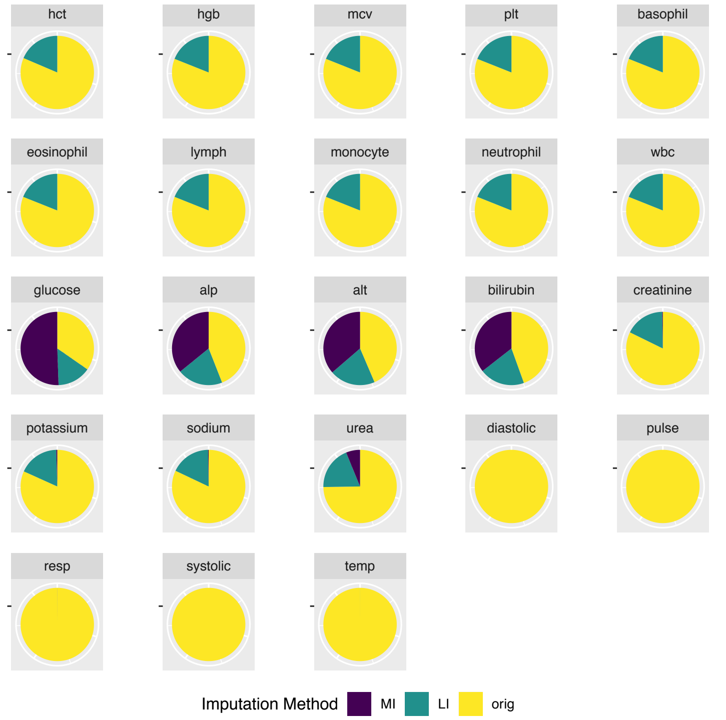

### PRE-DISCHARGE-LIKE STATE

Distribution of values for this state

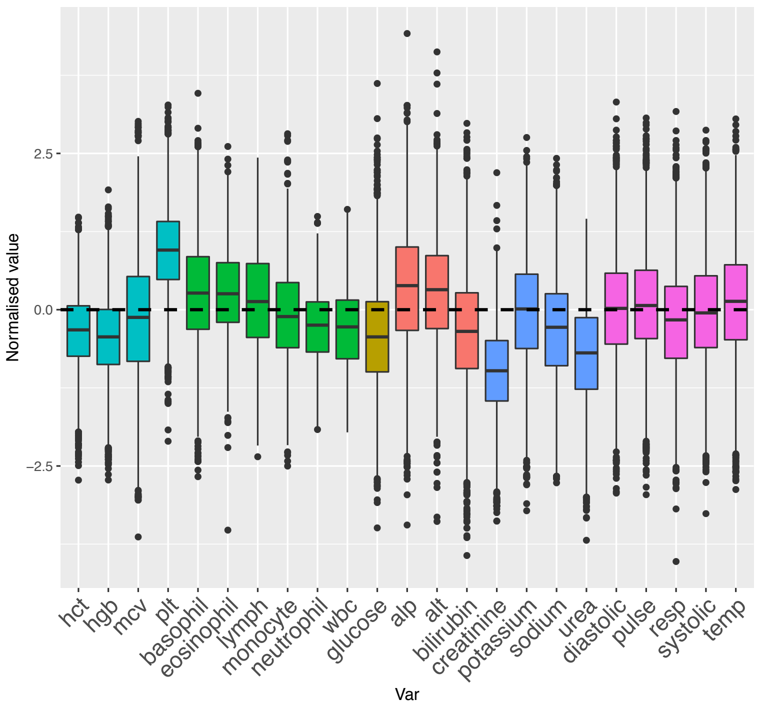


#### Clinical visual interpretation:

- **Low**: hgb, hct, creatinine, urea, glucose
- **High**: plt, alt, alp
- Everything else quite **near group me**an, especially Vital Signs.

Number of imputed and original observations by state
(current state highlighted with a green dot)

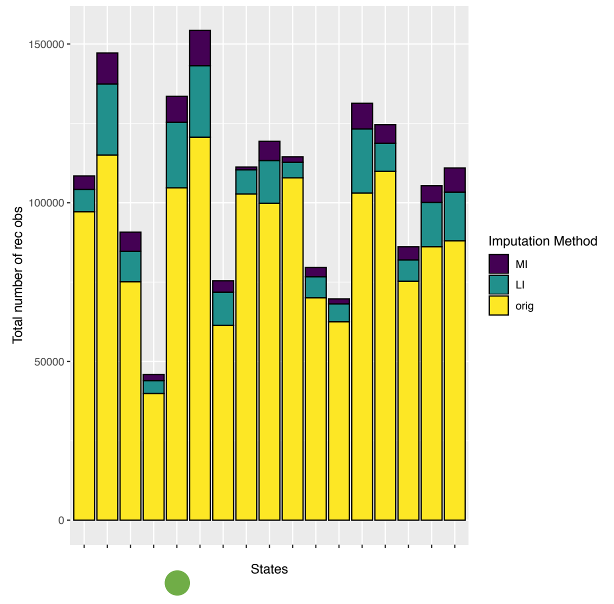


Distribution of imputed vs original values

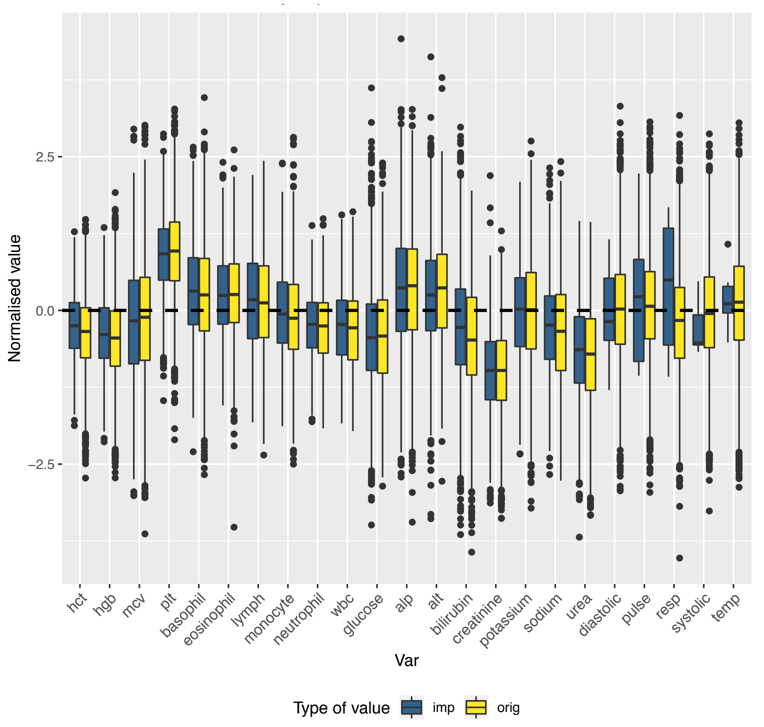


Distribution of original vs imputed values by imputation method

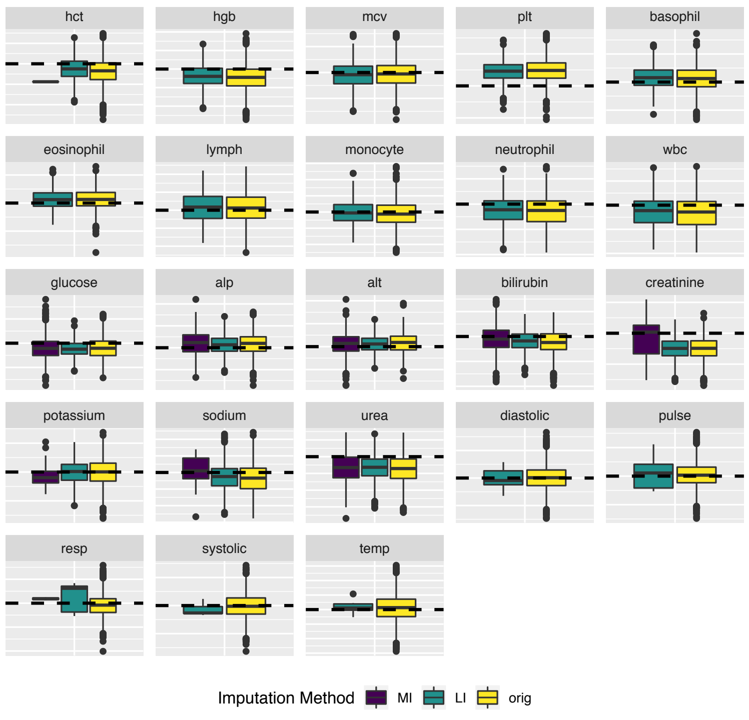


Number of imputed and original observations

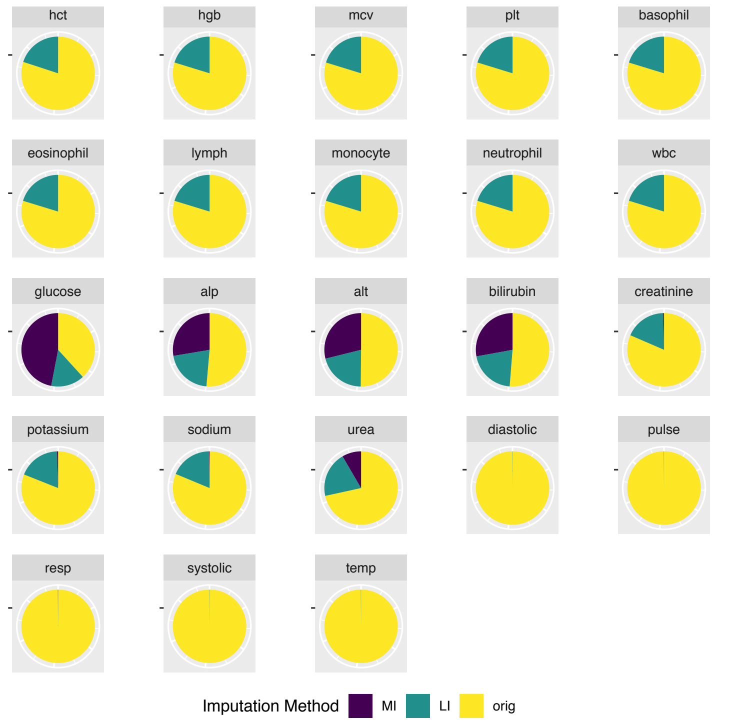

### PHYSIOLOGICAL-LIKE STATES

- Early inflammatory response
- Resolving inflammatory response
- Autoimmune/ Atopic
- Acute thrombotic
- Prolonged illness
- Other illness presentation

### EARLY INFLAMMATORY RESPONSE-LIKE STATE

Distribution of values for this state

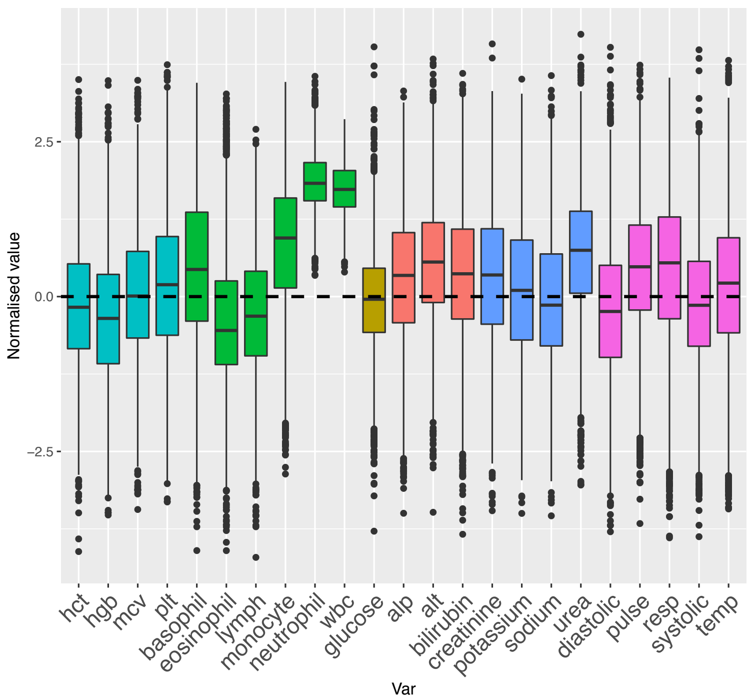


#### Clinical visual interpretation:

- **Markedly hig**h: wbc, neutrophils;
- **High**: monocytes, urea;
- **Relatively high**: pulse, resp, liver function tests, creatinine
- **Slightly high**: temp
- **Mildly low**: hgb, hct, eosinophils, lymph, diastolic and systolic BP

Number of imputed and original observations by state
(current state highlighted with a green dot)

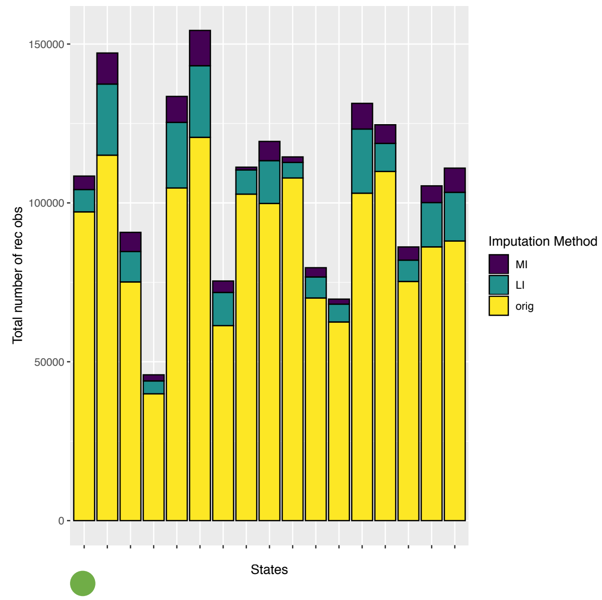


Distribution of imputed vs original values

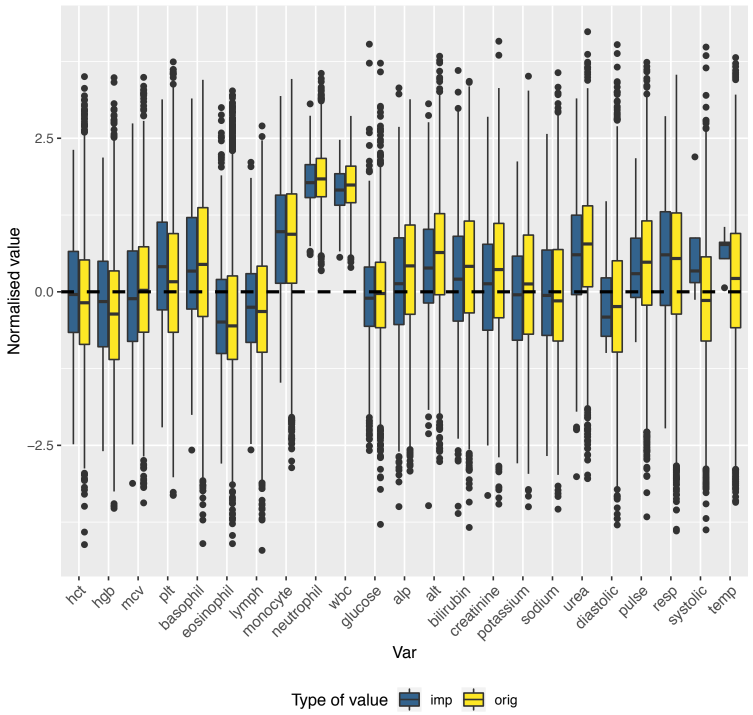


Distribution of original vs imputed values by imputation method

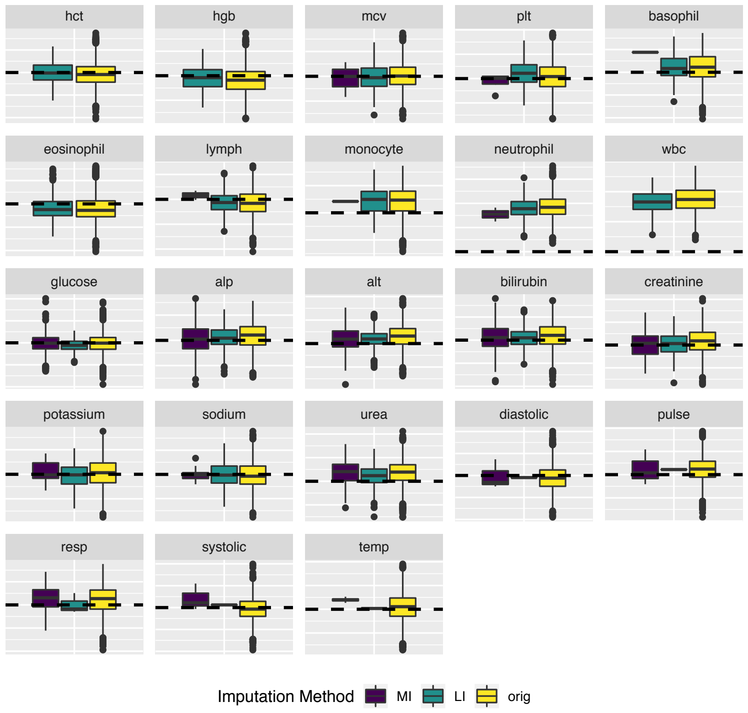


Number of imputed and original observations

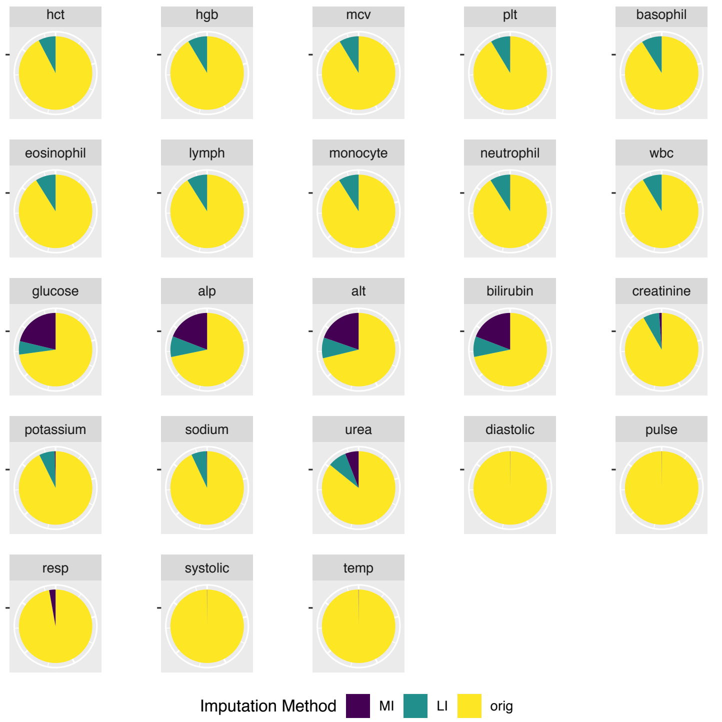

### RESOLVING INFLAMMATORY RESPONSE-LIKE STATE

Distribution of values for this state

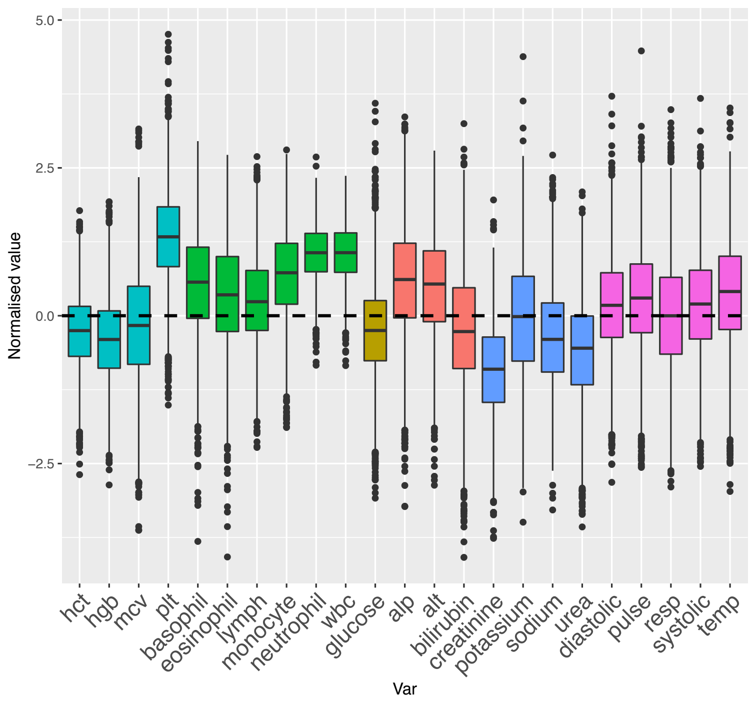


#### Clinical visual interpretation:

- **Higher**: wbc, neutrophils (with small variance);
- **Higher:** plt, monocytes, alp, alt (wider variance);
- **Slightly high**: Vitals Signs (except resp) and other white cells
- **Lower**: hgb, hct, creatinine, sodium

Number of imputed and original observations by state
(current state highlighted with a green dot)

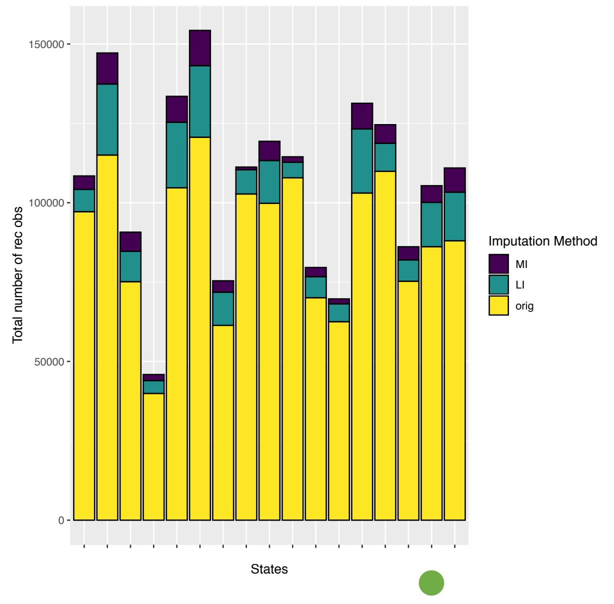


Distribution of imputed vs original values

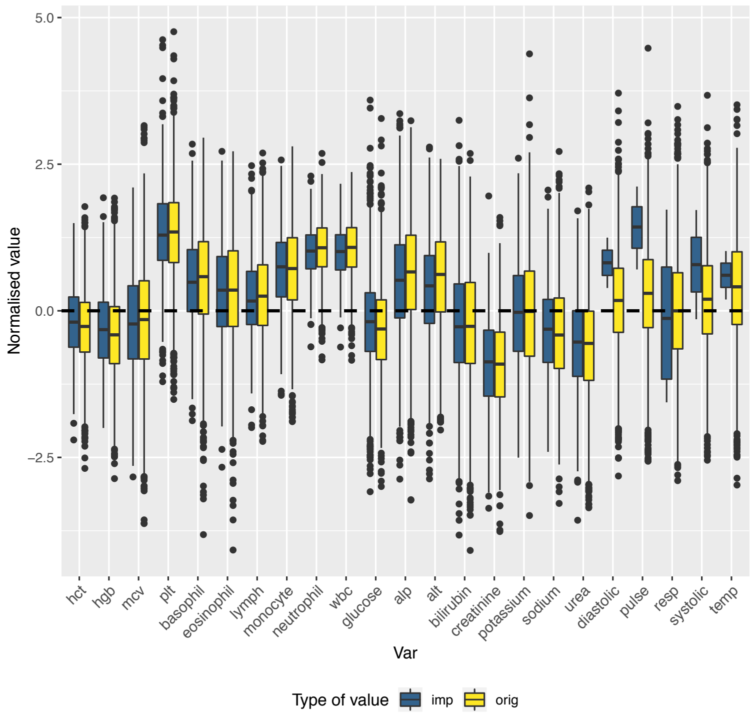


Distribution of original vs imputed values by imputation method

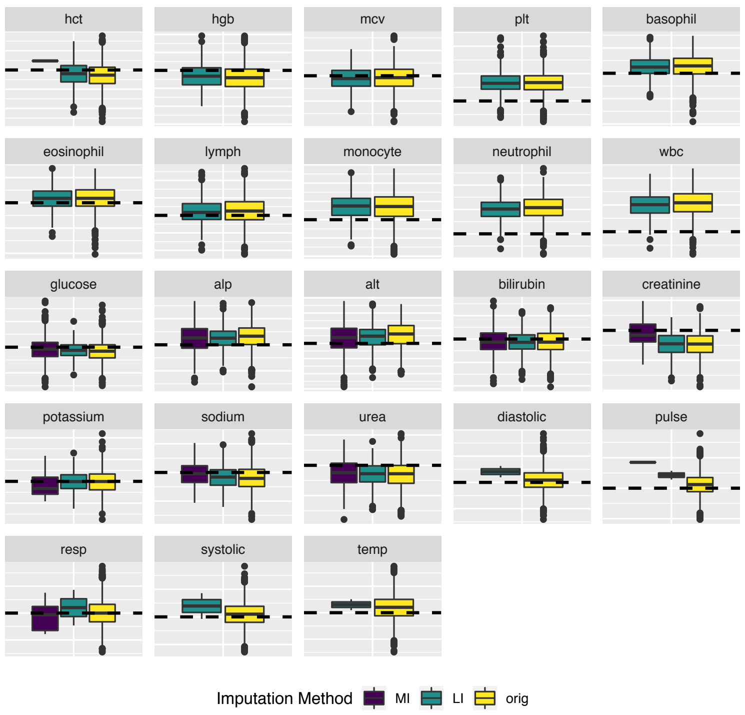


Number of imputed and original observations

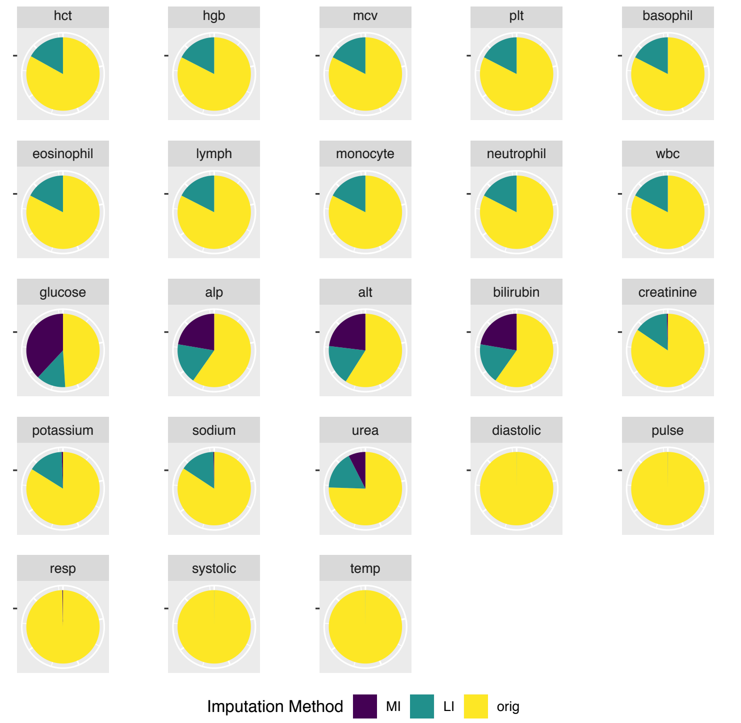

### AUTOIMMUNE/ ATOPIC-LIKE STATE

Distribution of values for this state

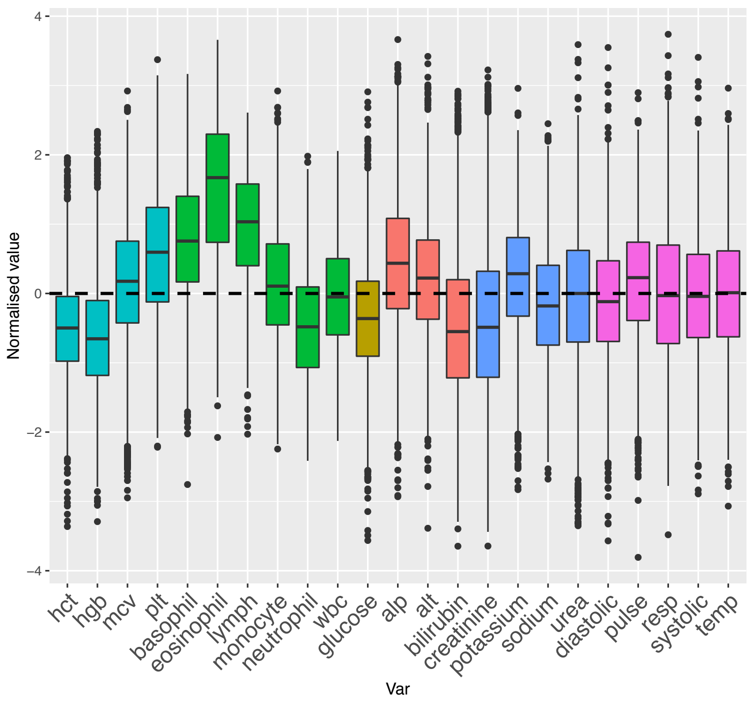


#### Clinical visual interpretation:

- **Markedly higher:** basophils, eosinophils and lymphs;
- **High**: plts;
- **Lower:** hgb, hct, neutrophils, glucose;
- **Slightly low**: bilirubin, creatinine.

Number of imputed and original observations by state
(current state highlighted with a green dot)

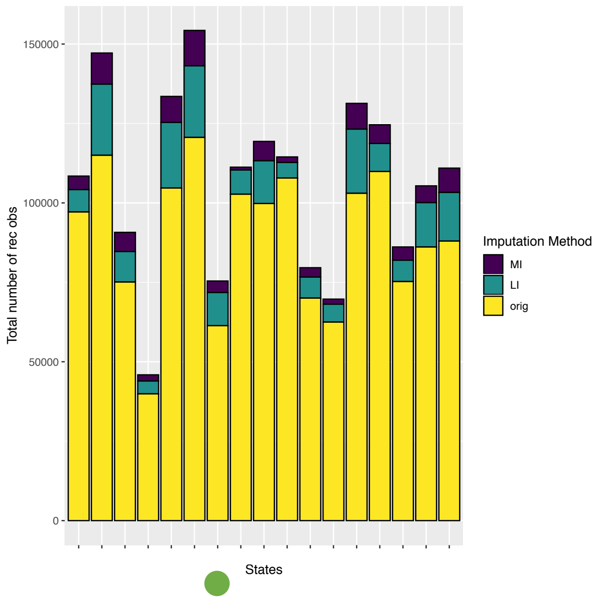


Distribution of imputed vs original values

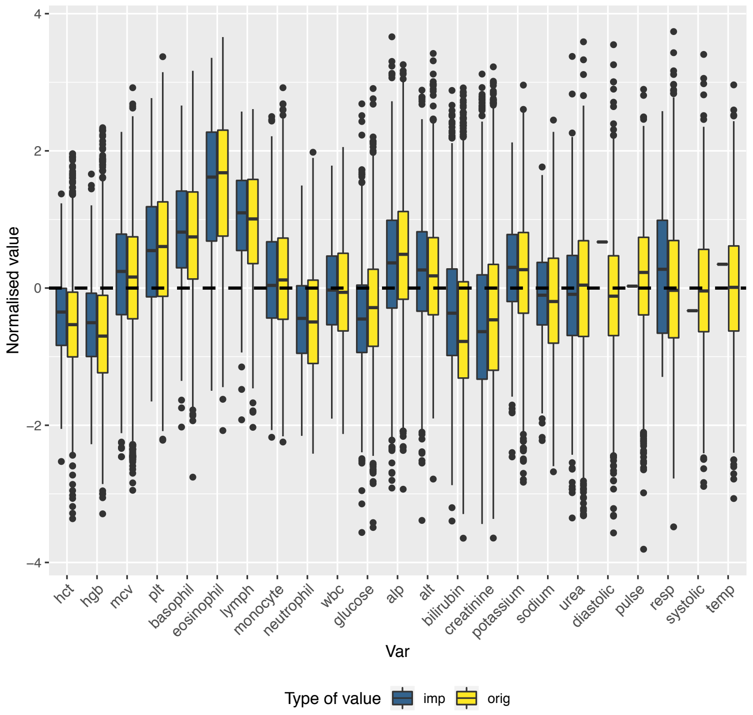


Distribution of original vs imputed values by imputation method

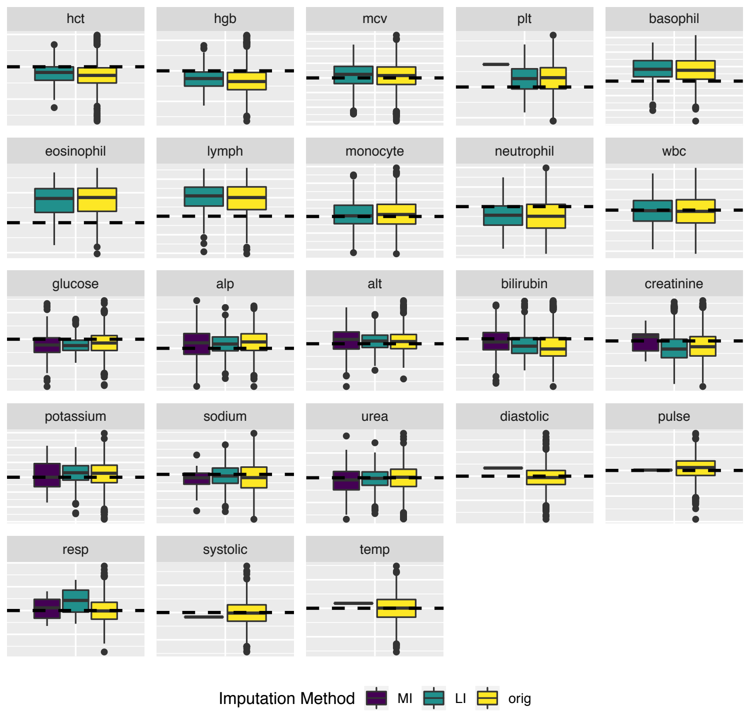


Number of imputed and original observations

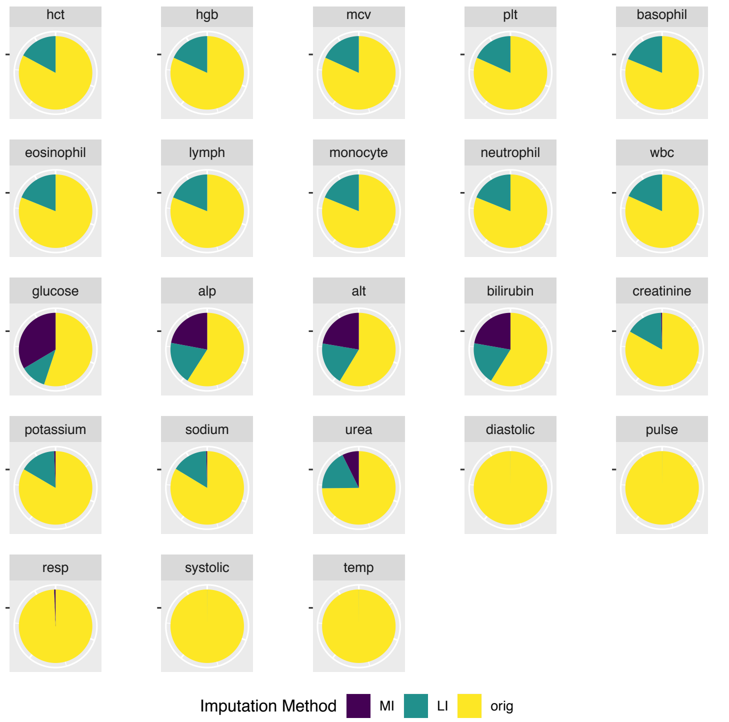

### ACUTE THROMBOTIC-LIKE STATE

Distribution of values for this state

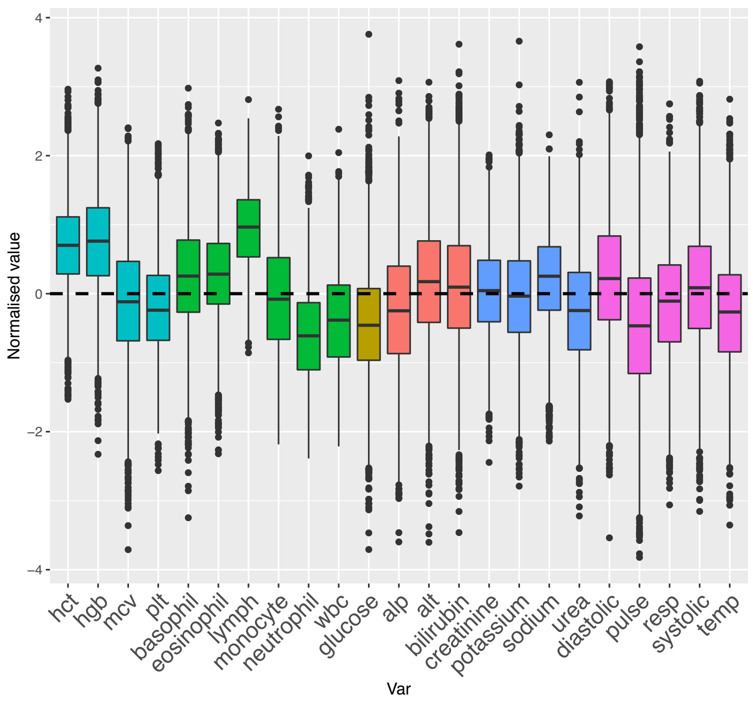


#### Clinical visual interpretation:

- **Higher**: hgb, hct, lypmph;
- **Lower**: neutrophils, wbc, glucose, pulse
- **Mildly low**: temp

Number of imputed and original observations by state
(current state highlighted with a green dot)

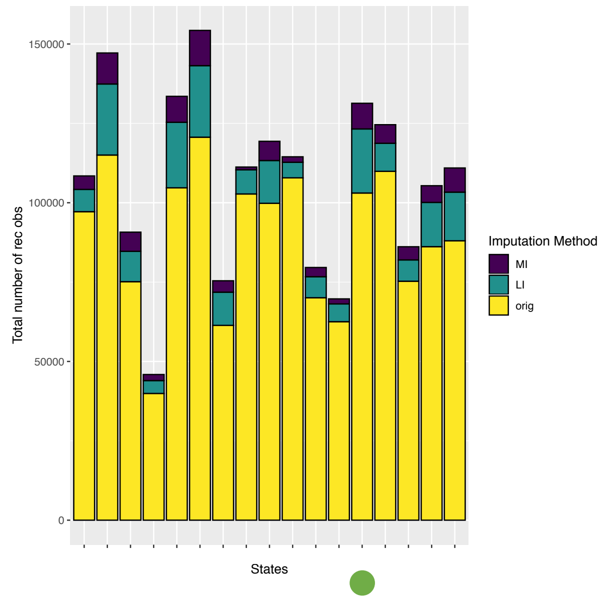


Distribution of imputed vs original values

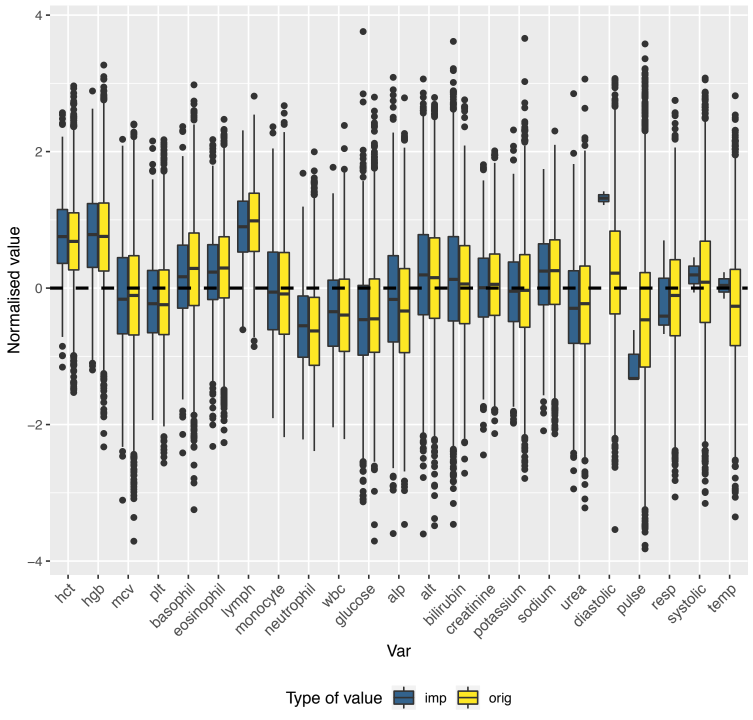


Distribution of original vs imputed values by imputation method

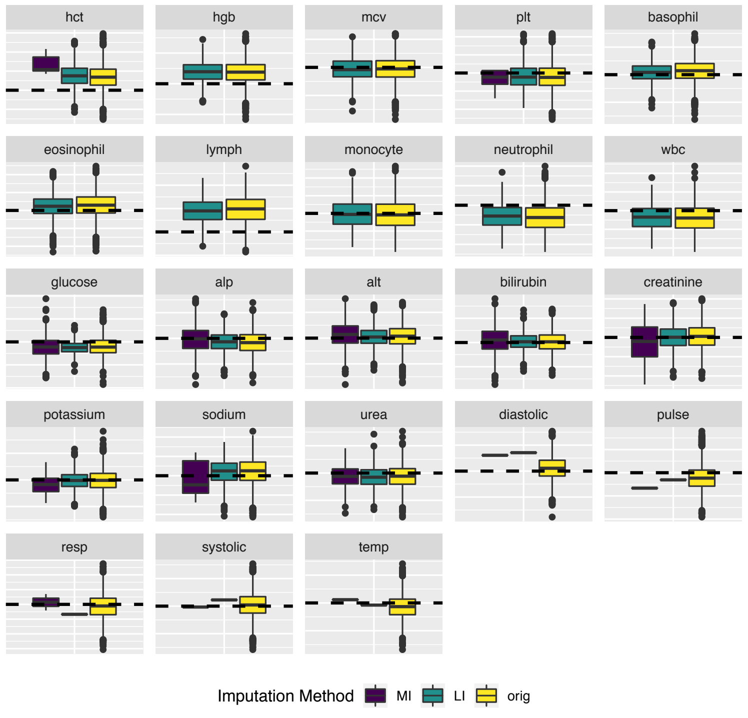


Number of imputed and original observations

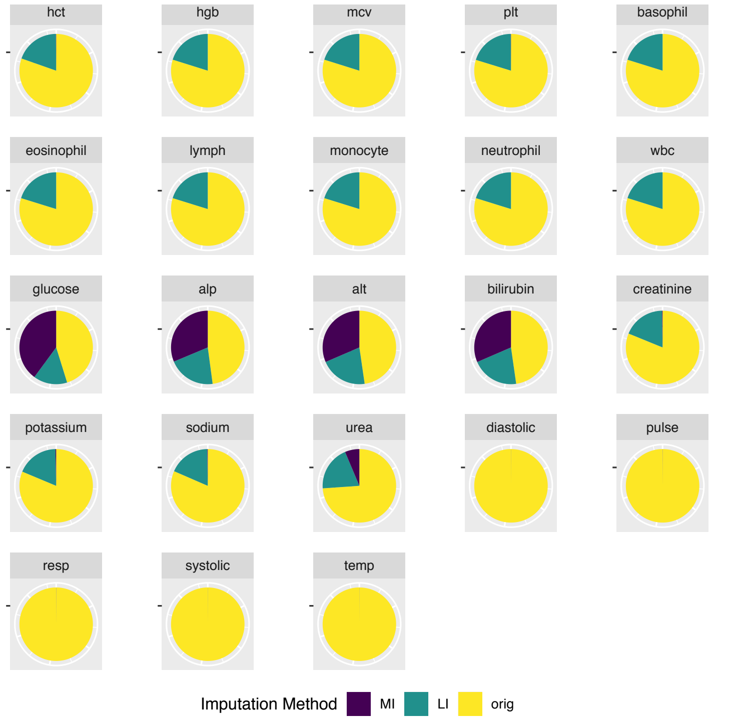

### PROLONGED ILLNESS-LIKE STATE

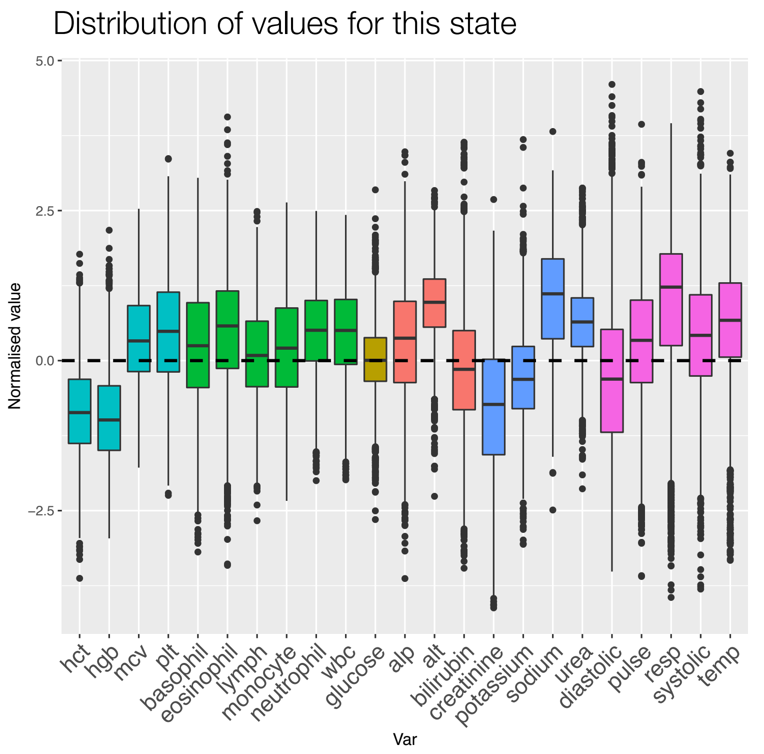


#### Clinical visual interpretation:

- **Lower**: hgb, hct, creatinine;
- **Higher**: mcv, plt, wbc, neutrophils, eosinophils, alt, sodium, urea, resp.
- **Mildly high**: other Vital Signs (except diastolic BP)

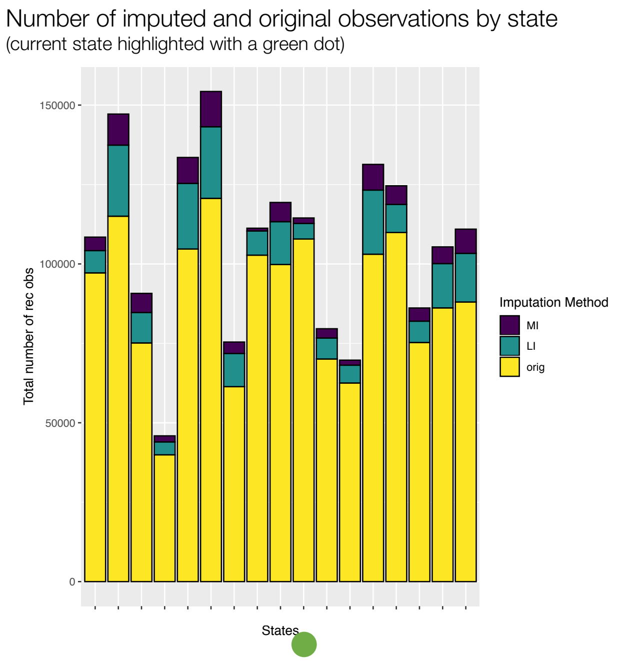


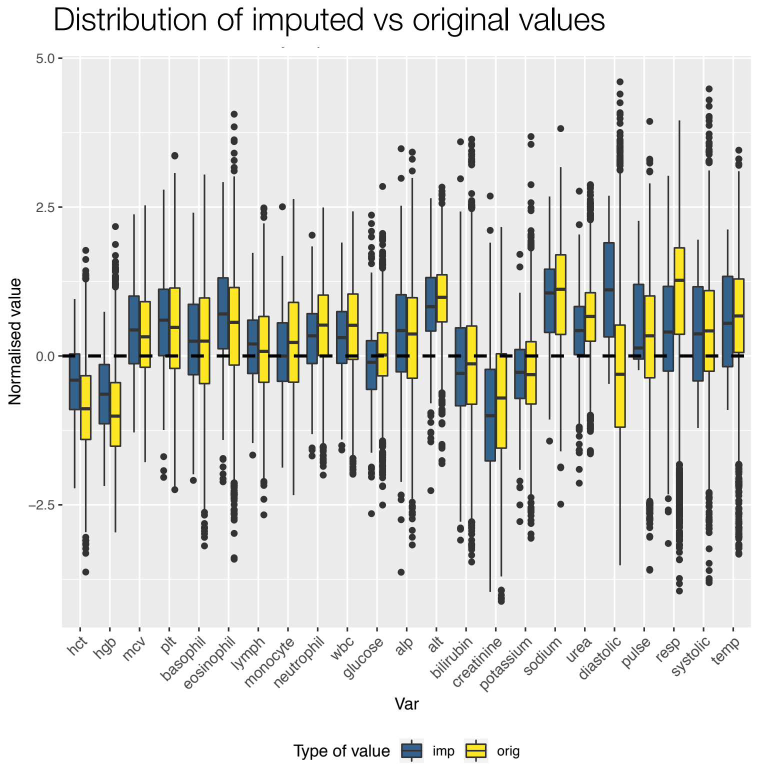


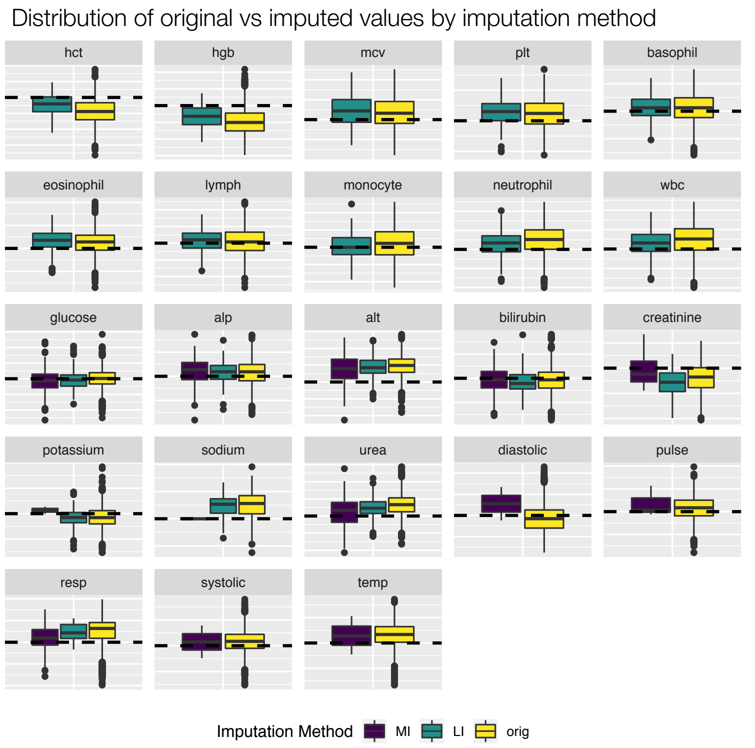


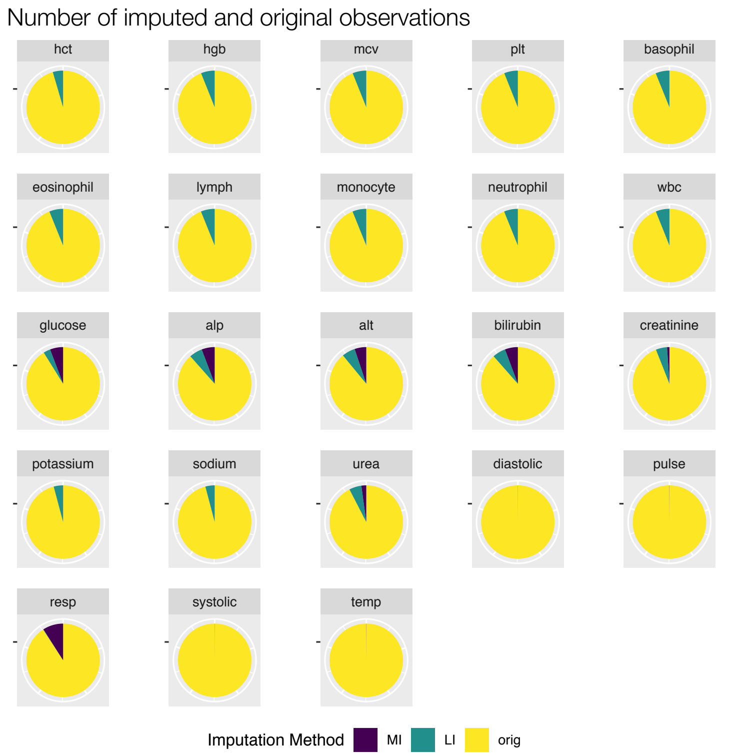

### OTHER ILLNESS PRESENTATION-LIKE STATE

Distribution of values for this state

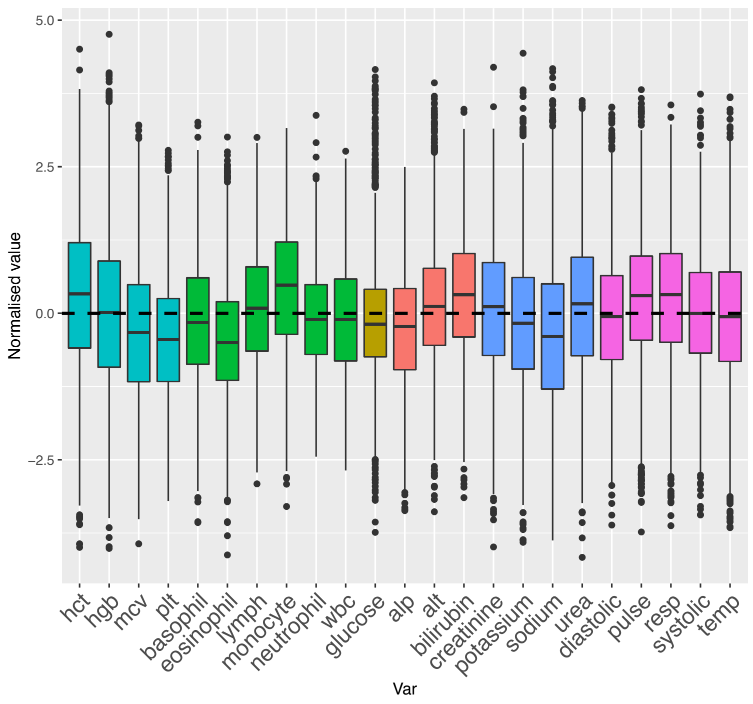


#### Clinical visual interpretation:

- Most obvious feature is ‘normality’: no stand out derangement of Vital signs or lab tests

Number of imputed and original observations by state
(current state highlighted with a green dot)

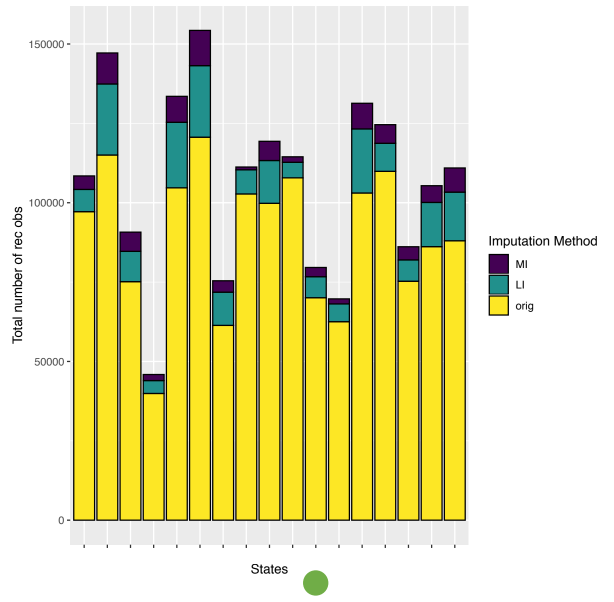


Distribution of imputed vs original values

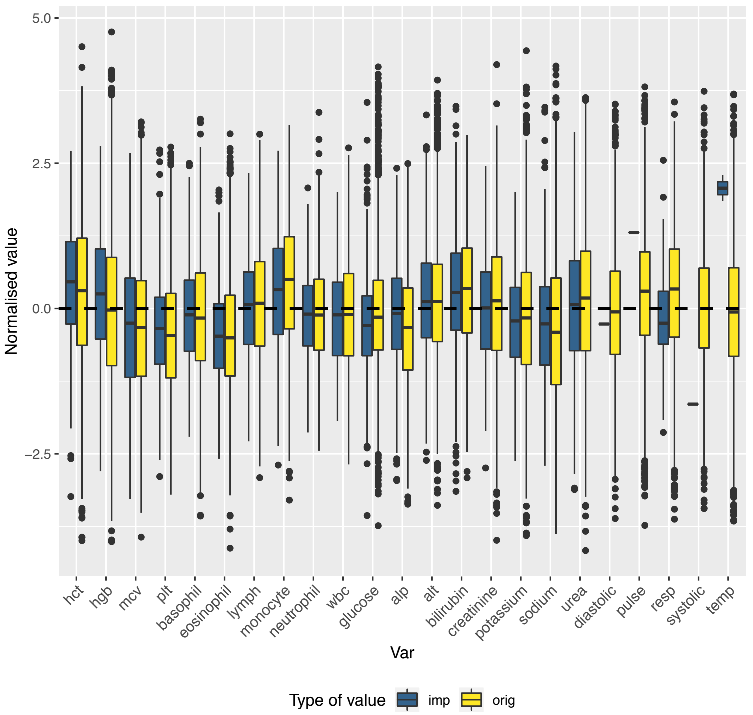


Distribution of original vs imputed values by imputation method

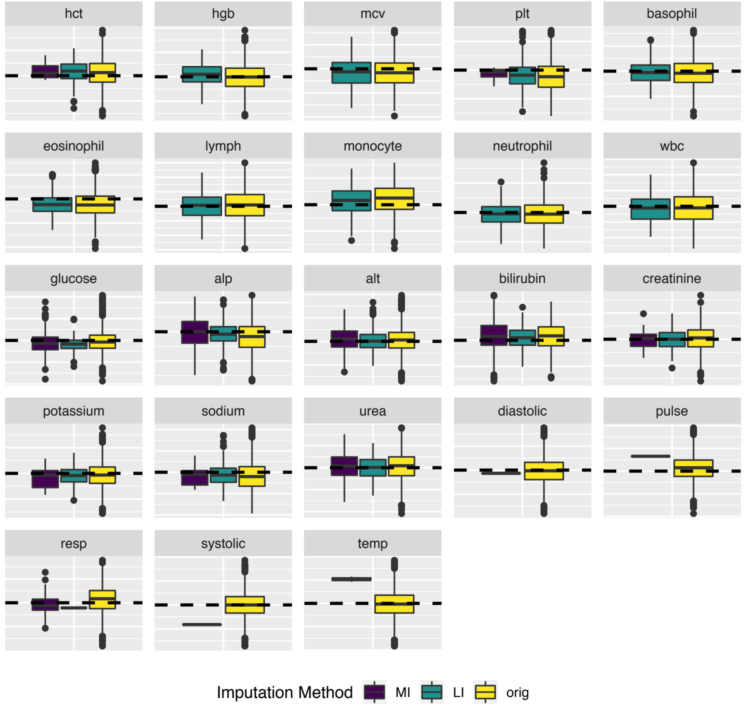


Number of imputed and original observations

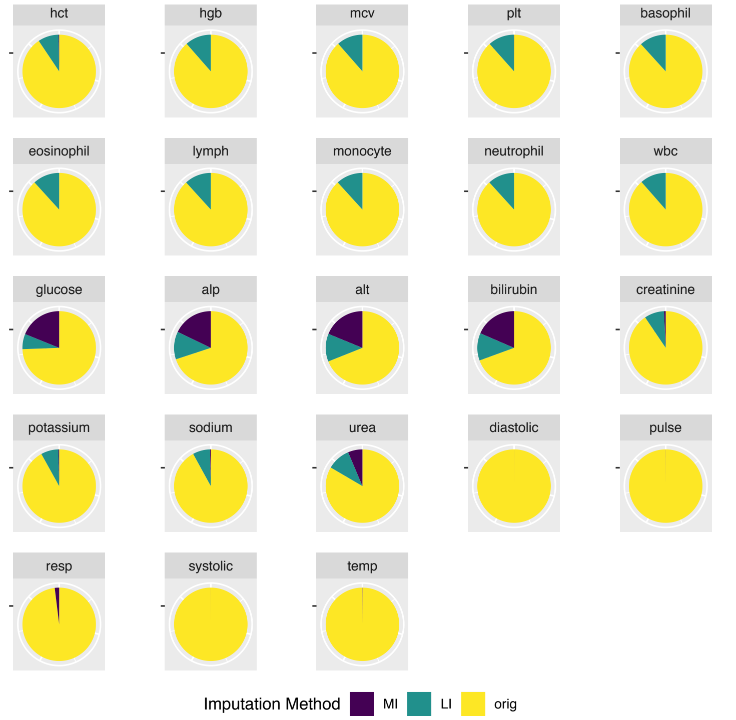
