## Supplementary Figures 5 for "Big Data Analysis of Electronic Health Records: Clinically interpretable representations of older adult inpatient trajectories using time-series numerical data and Hidden Markov Models"

### DISEASE-LIKE STATES

- Hepatic
- Stable renal
- Unstable renal
- Stable but static renal
- Blood dyscrasia
- Bone marrow suppression

### HEPATIC-LIKE STATE

#### AEs grouped by Primary Diagnosis

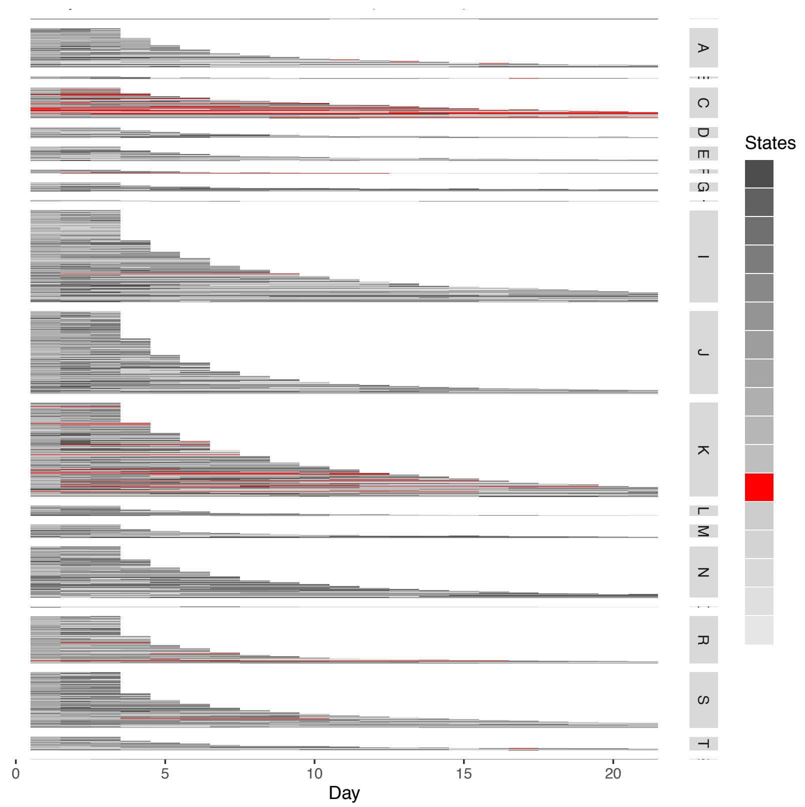


**Associated with:**

C: *Neoplasms*;

K: *Diseases of the digestive system;*

#### AEs grouped by Inpatient Mortality

Inpatient Death (ID) or Discharged Alive (DA)

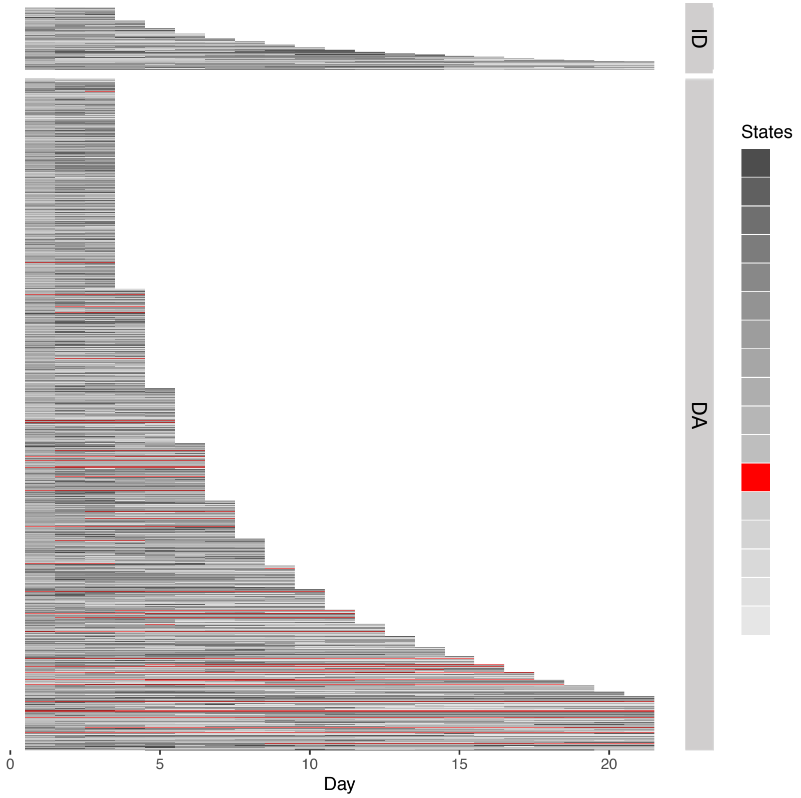


**Associated with:**

Medium/ long LOS

**Not associated with:**

Discharge status (although more in discharged alive patients with long LOS)

#### Distribution of Lab Tests and Vital Signs

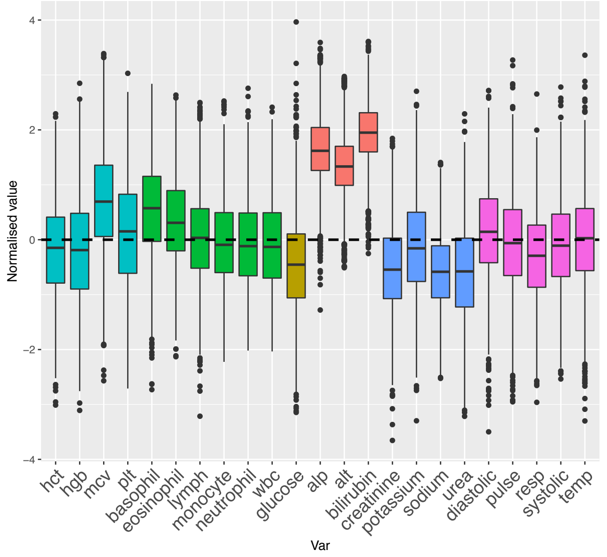


**Very high**: alt, alp and bilirubin;

**High**: mcv;

**Lower**: urea, glucose, creatinine, sodium

### STABLE RENAL-LIKE STATE

#### AEs grouped by Primary Diagnosis

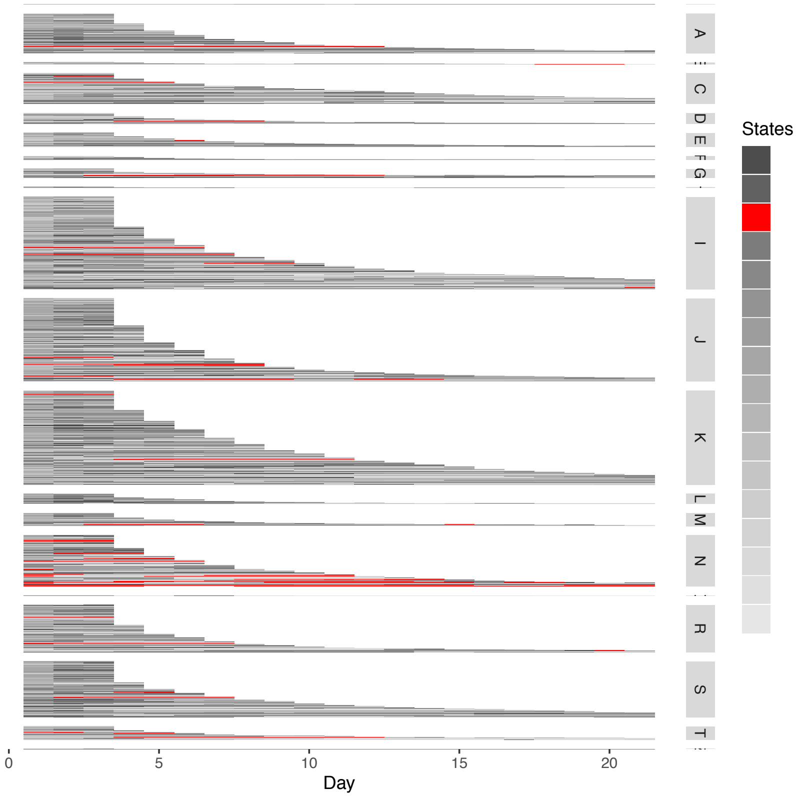


**Associated with:**

N: *Genitourinary system*;
I: *Circulatory system*;

#### AEs grouped by Inpatient Mortality

Inpatient Death (ID) or Discharged Alive (DA)

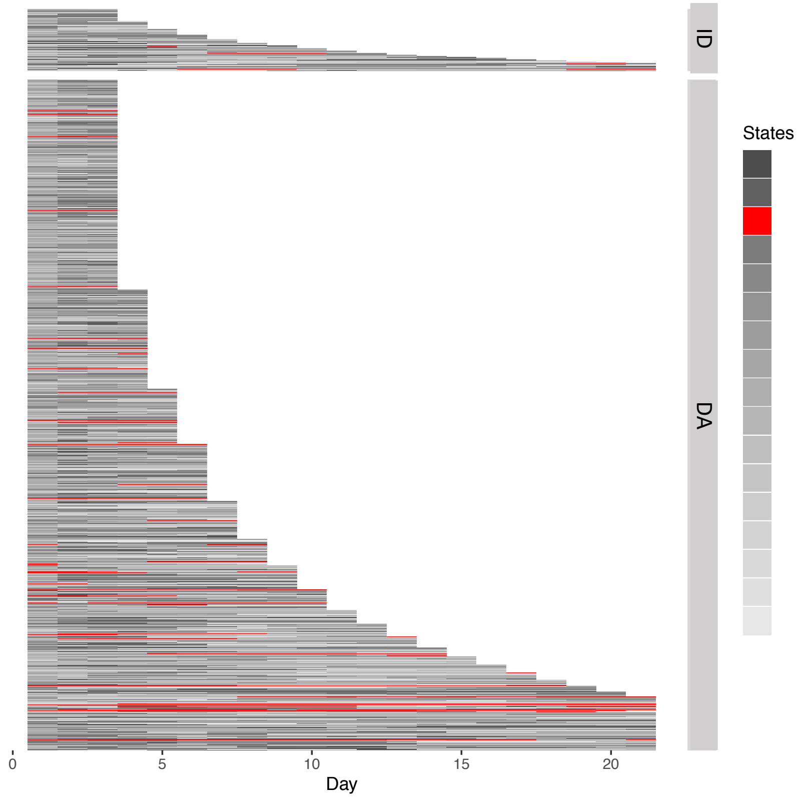


**Associated with:**

Medium and long LOS (present at various stages of AE)

**Not associated with:**

Discharge status

#### Distribution of Lab Tests and Vital Signs

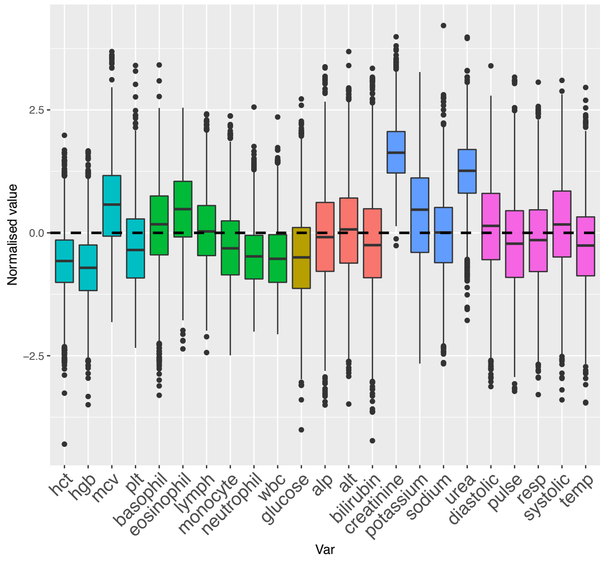


**Very high**: creatinine, urea (with small variance);

**High**: potassium, eosinophils, mcv;

**Low**: hgb, hct, wbc, neutrophils, glucose;

Liver Function Tests and Vital Signs all **near group mean or slightly lower** (i.e., maybe closer to healthy population mean)

### UNSTABLE RENAL-LIKE STATE

#### AEs grouped by Primary Diagnosis

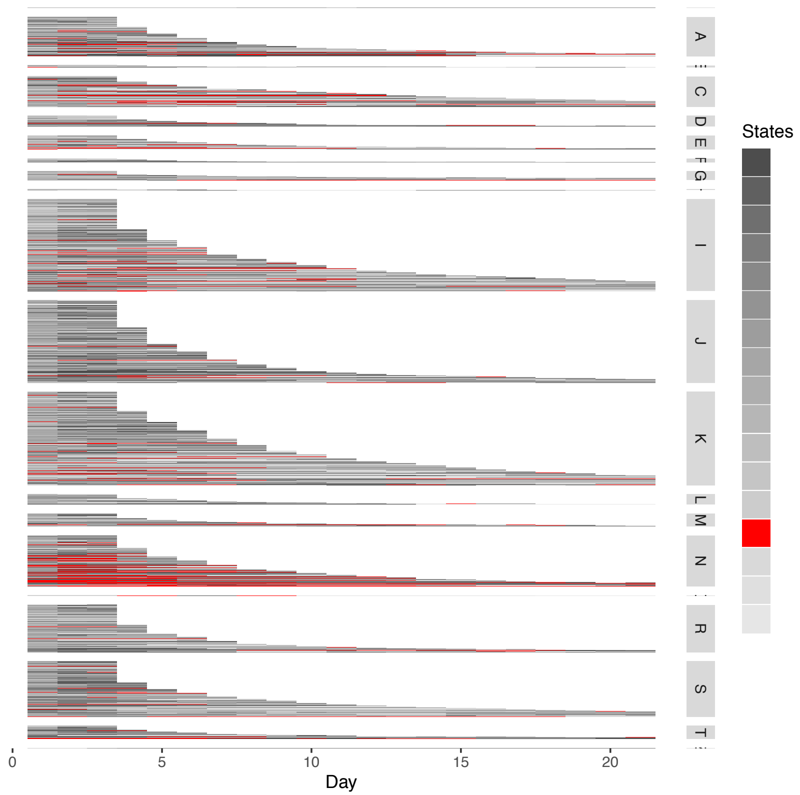


**Associated with:**

N: *Genitourinary system;*

Also present in other diagnostic codes to lesser extent;

#### AEs grouped by Inpatient Mortality

Inpatient Death (ID) or Discharged Alive (DA)

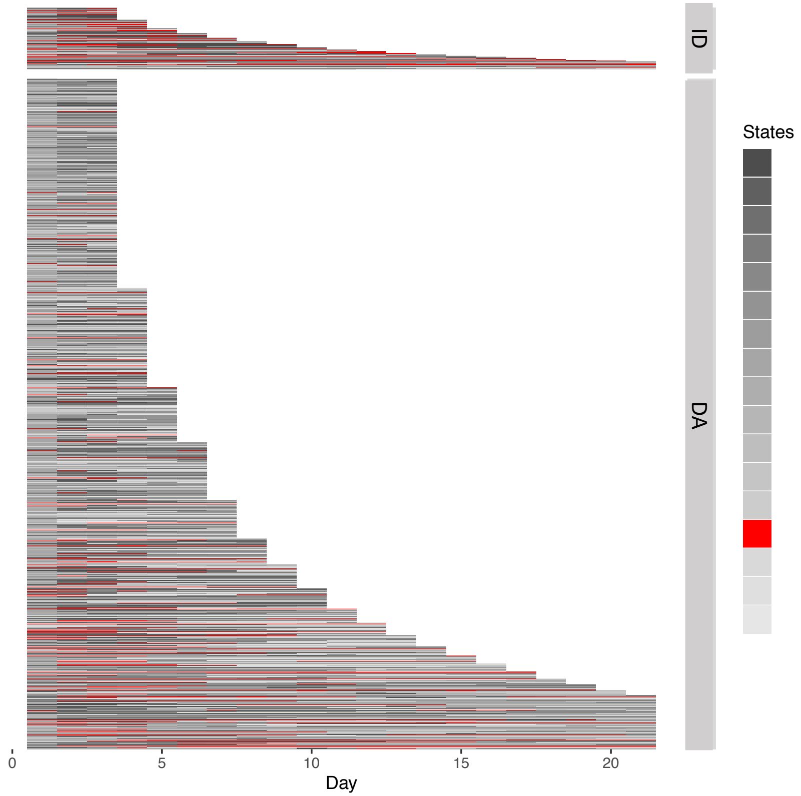


**Associated with:**

Slightly more prevalent in patients with long LOS;

Discharge as deceased.

#### Distribution of Lab Tests and Vital Signs

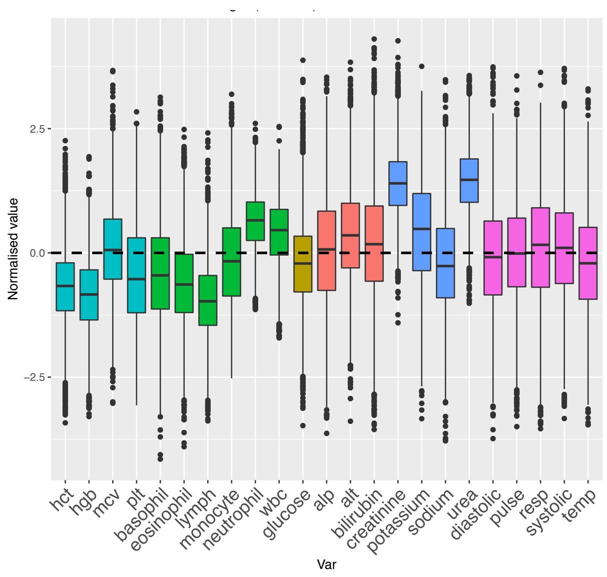


**Very high**: urea, creatinine (with low variance), potassium;

**Higher**: wbc, neutrophils;

**Lower**: hgb, hct, lymphs;

Liver Function Tests and Vital Signs all **around group mean or** on **higher** side of group mean

### STABLE BUT STATIC RENAL-LIKE STATE

#### AEs grouped by Primary Diagnosis

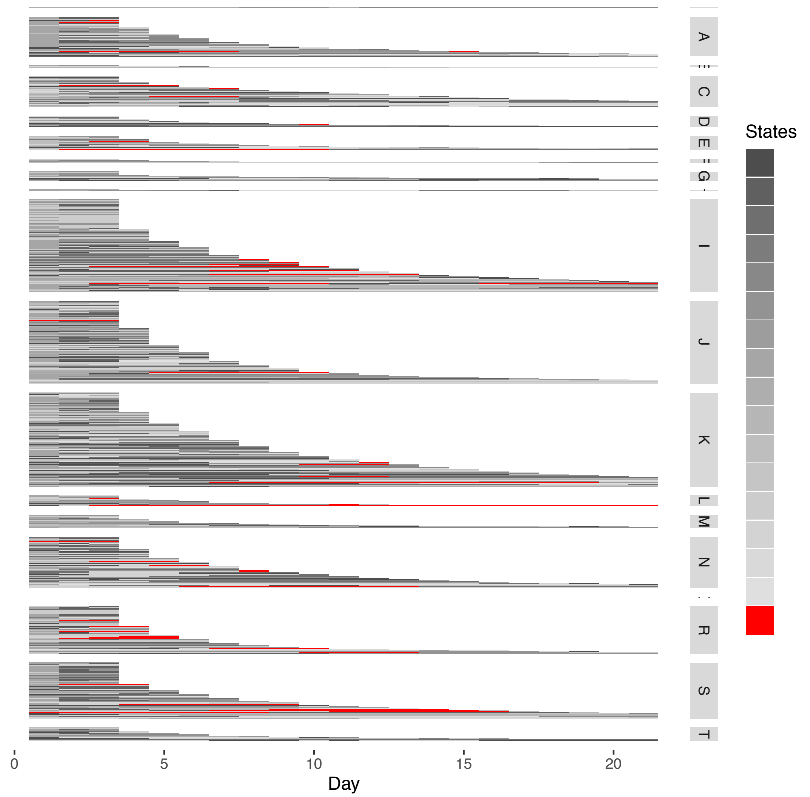


**Associated with:**

Slight increase in:

I: *Circulatory system*;

N: *Genitourinary system*;

S: *Injury, poisoning or other external causes;*

(could be as more medium/ long AE)

#### AEs grouped by Inpatient Mortality

Inpatient Death (ID) or Discharged Alive (DA)

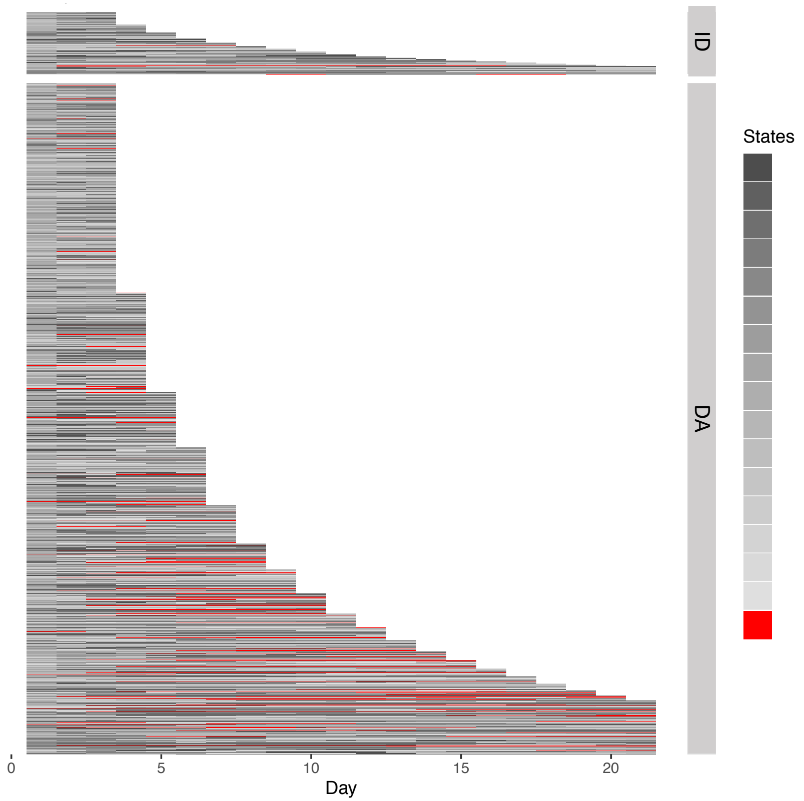


**Associated with:**

Discharge alive;

Mid-late stage of admission episode;

Long LOS.

#### Distribution of Lab Tests and Vital Signs

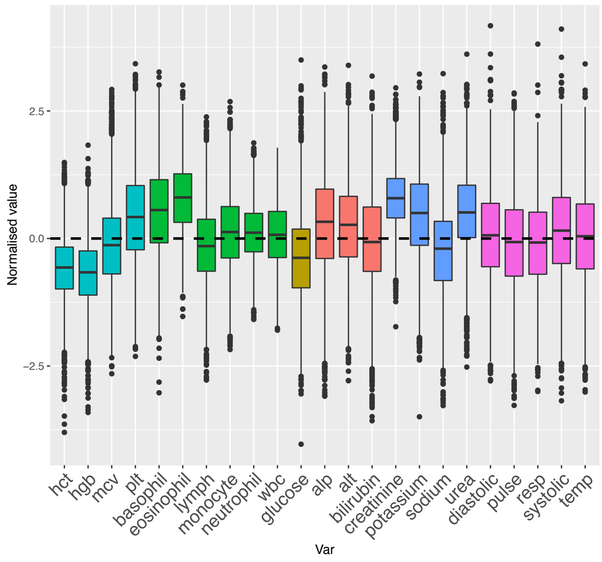


**Lower**: hgb, hct, glucose;

**Higher**: eosonophils, creatinine, urea, potassium;

**‘Normal’**: Vital signs, white blood cells

### BLOOD DYSCRASIA-LIKE STATE

#### AEs grouped by Primary Diagnosis

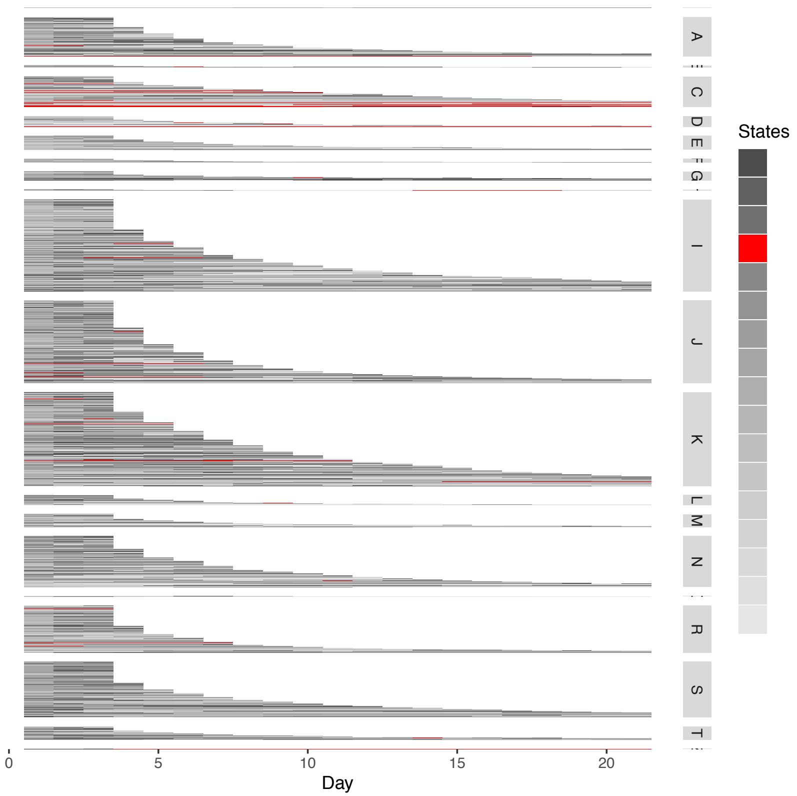


**Associated with:**

A: *Infectious and parasitic diseases*;

C: *Neoplasms*;

D: *Blood and blood-forming organs and certain disorders involving the immune mechanism*;

#### AEs grouped by Inpatient Mortality

Inpatient Death (ID) or Discharged Alive (DA)

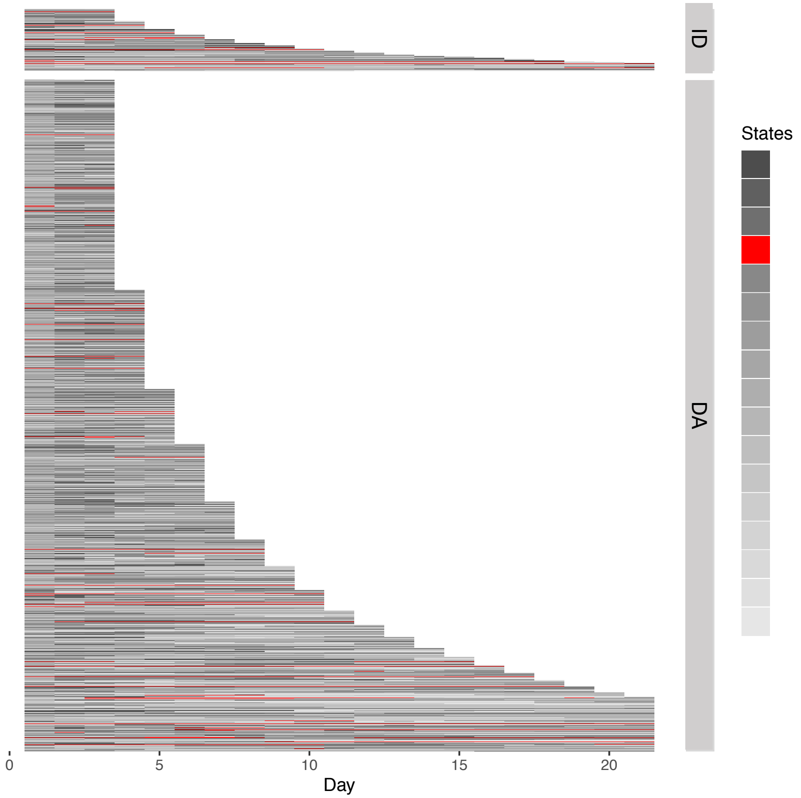


**Not associated with:**

No strong association with discharge status or LOS

#### Distribution of Lab Tests and Vital Signs

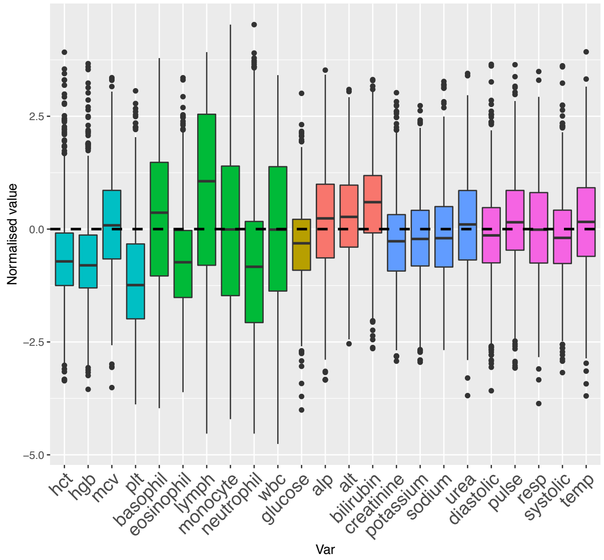


**Lower**: hgb, hct, plts;

**Deranged** wbc in both directions with wide variance;

**Higher**: bilirubin;

Other Liver and Renal Function Tests and Vital Signs all **very close to group mean**

### BONE MARROW SUPPRESSION-LIKE STATE

#### AEs grouped by Primary Diagnosis

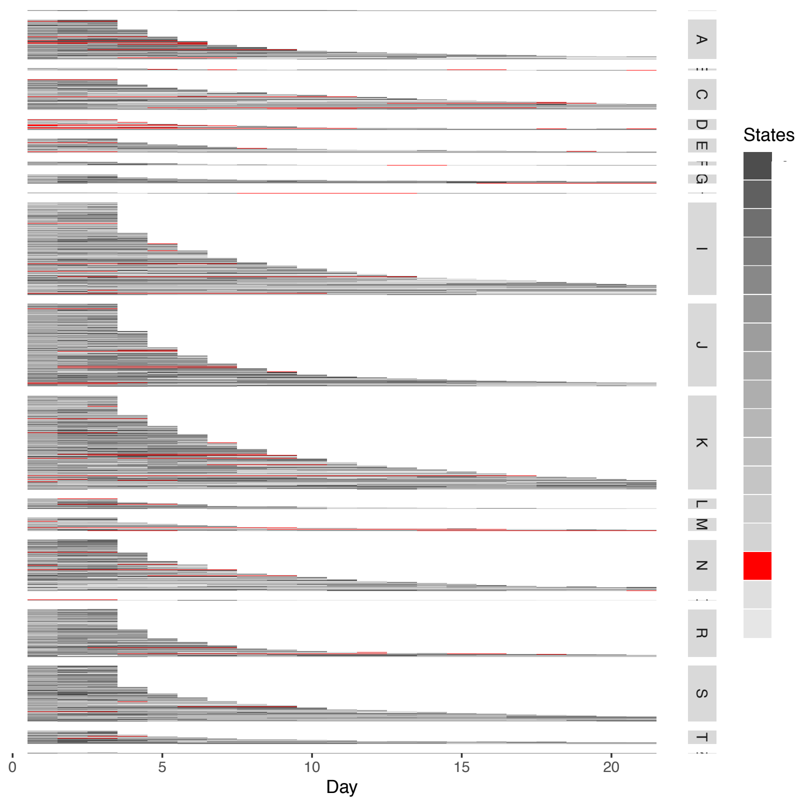


**Associated with:**

D: *Blood and blood-forming organs and certain disorders involving the immune mechanism*;
A: *Infectious and parasitic diseases;*
C: *Neoplasms*;

#### AEs grouped by Inpatient Mortality

Inpatient Death (ID) or Discharged Alive (DA)

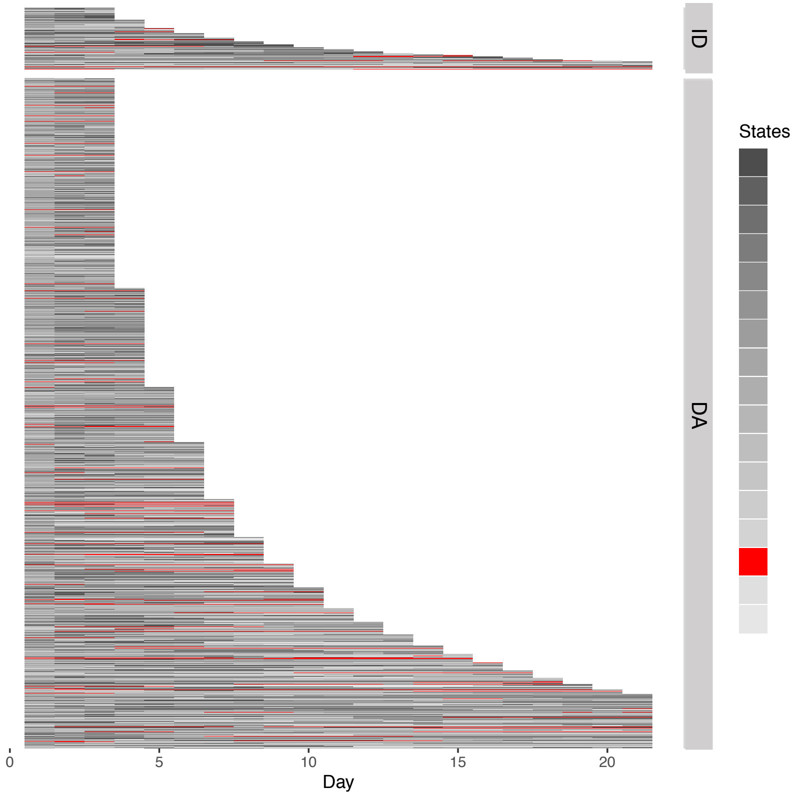


**Associated with:**

Weakly associated with Discharged alive and with long LOS

#### Distribution of Lab Tests and Vital Signs

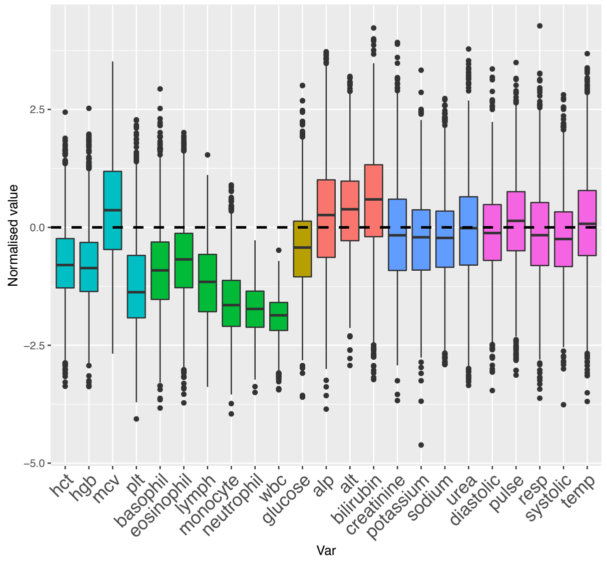


‘*Pancytopenia*’: all blood count parameters **lower** except MCV which is **higher**;

**Mildly high**: Liver Function Tests (especially bilirubin);

**Slightly lower**: glucose;

Renal Function Tests and Vital Signs just under group mean but generally unremarkable

### ADMISSION STAGE-LIKE STATES

- Acute presentation
- Treatment response (1)
- Treatment response (2)
- Early discharge
- Pre-discharge

### ACUTE PRESENTATION-LIKE STATE

#### AEs grouped by Primary Diagnosis

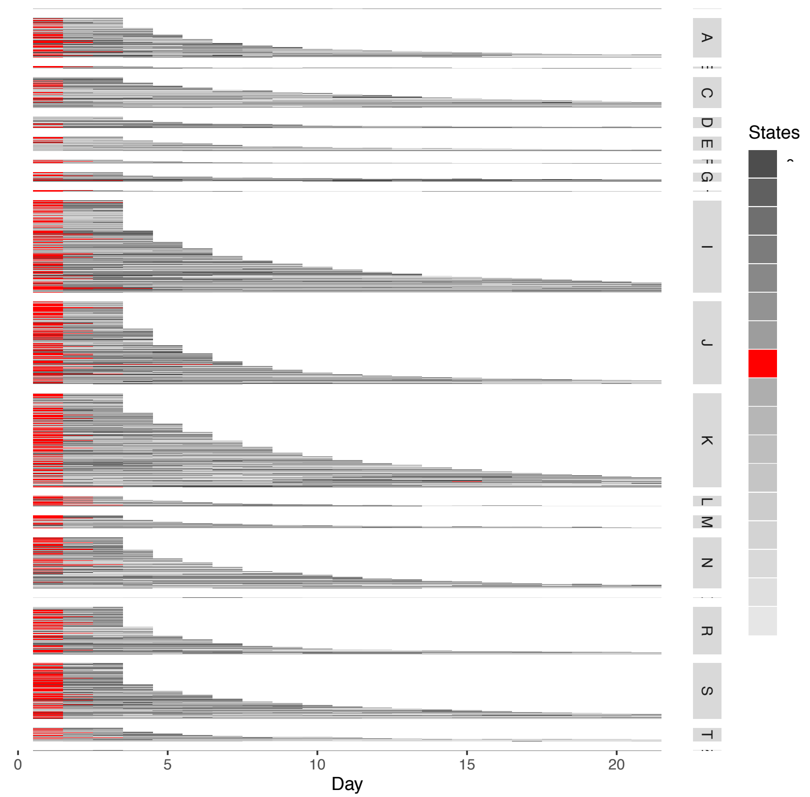


**Not associated with:**

Diagnostic codes;

#### AEs grouped by Inpatient Mortality

Inpatient Death (ID) or Discharged Alive (DA)

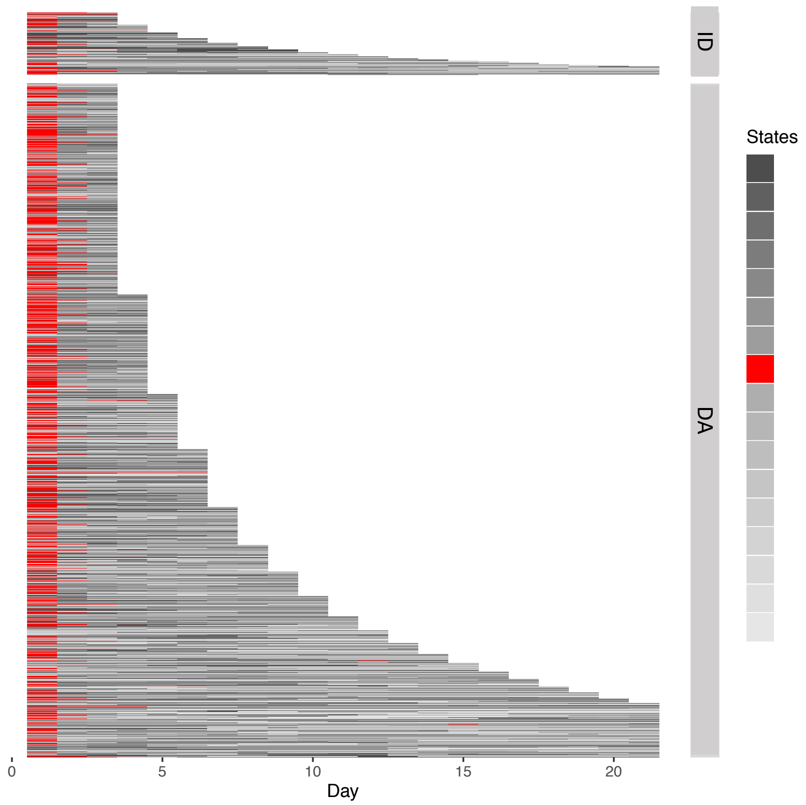


**Associated with:**

Day 1 of admission;

**Not associated with:**

Discharge status;
LOS.

#### Distribution of Lab Tests and Vital Signs

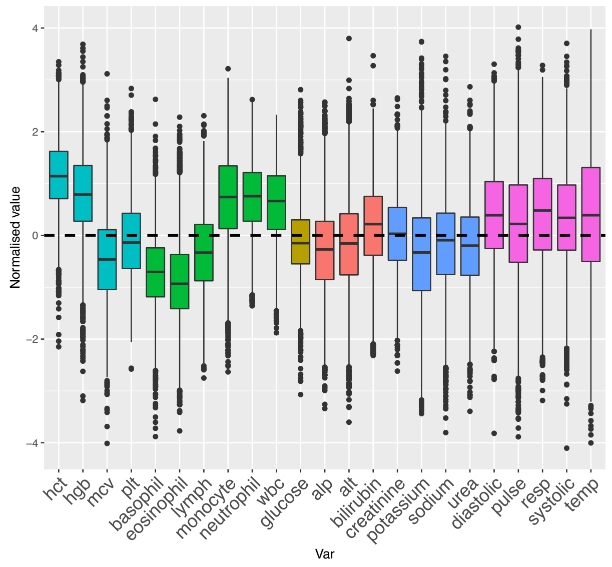


**Higher**: hct, hgb, wbc, neutrophils, monocytes;

**Slightly higher**: Vital signs;

**Lower:** basophils, eosinophils, mcv.

### TREATMENT RESPONSE-LIKE STATE (1)

#### AEs grouped by Primary Diagnosis

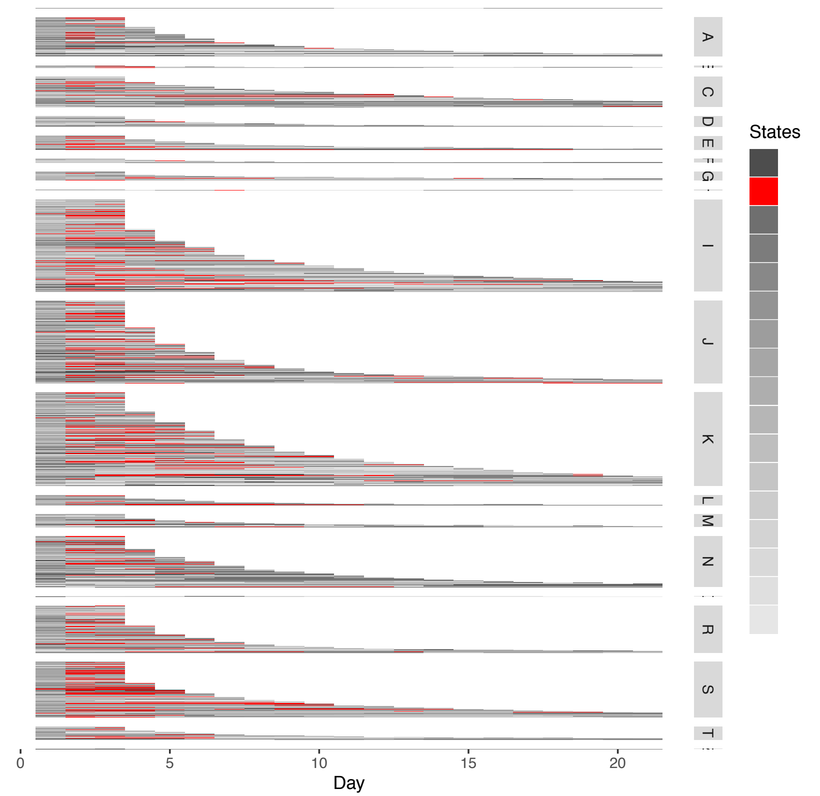


**Not associated with:**

Diagnostic codes;

#### AEs grouped by Inpatient Mortality

Inpatient Death (ID) or Discharged Alive (DA)

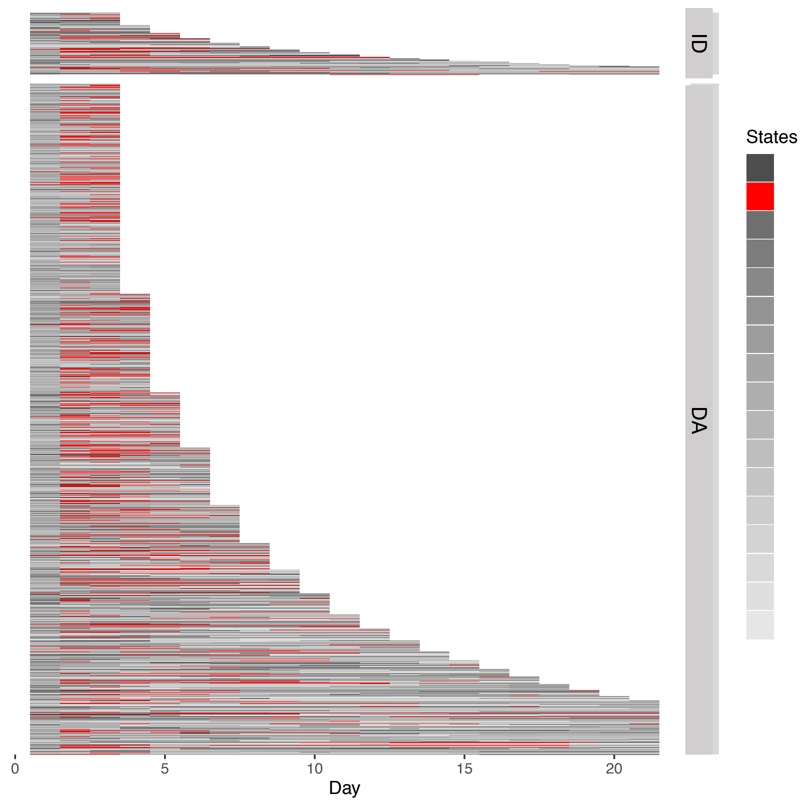


**Associated with:**

Day 2-8 of admission;
More common in short/ medium LOS (where it can be present on discharge);
Discharged alive.

#### Distribution of Lab Tests and Vital Signs

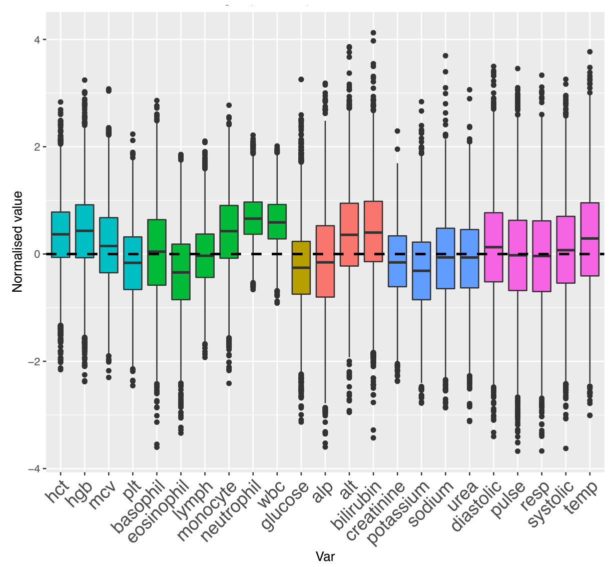


**Higher**: hct, hgb, wbc, neutrophils, monocytes;

**Slightly higher**: alt, bilirubin.

Lab Tests seem to be in a pattern that is similar to “Acute presentation state” – but less severe derangement.

### TREATMENT RESPONSE-LIKE STATE (2)

#### AEs grouped by Primary Diagnosis

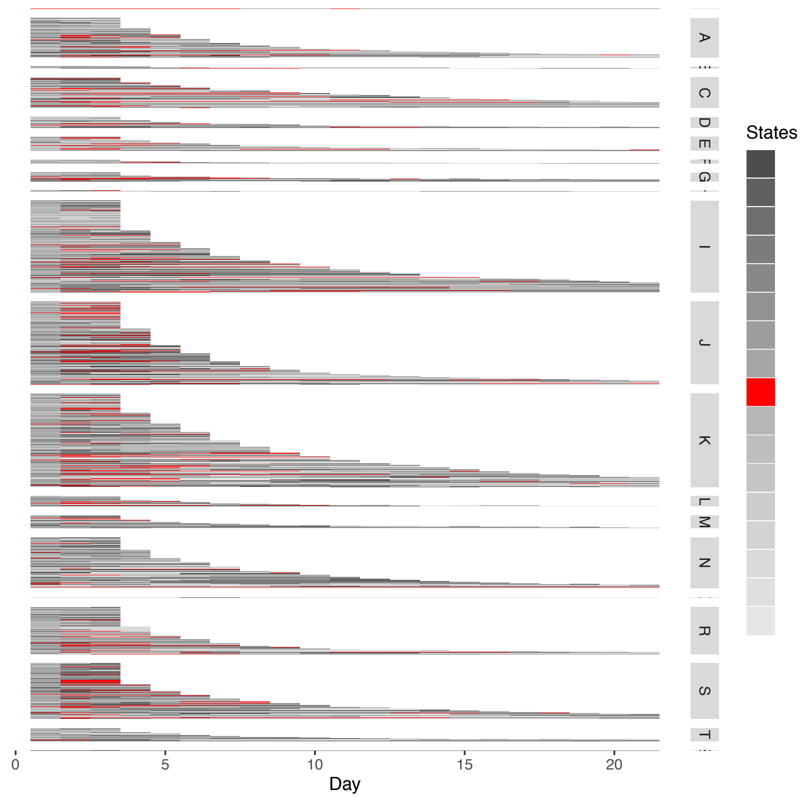


**Not associated with:**

Diagnostic codes

#### AEs grouped by Inpatient Mortality

Inpatient Death (ID) or Discharged Alive (DA)

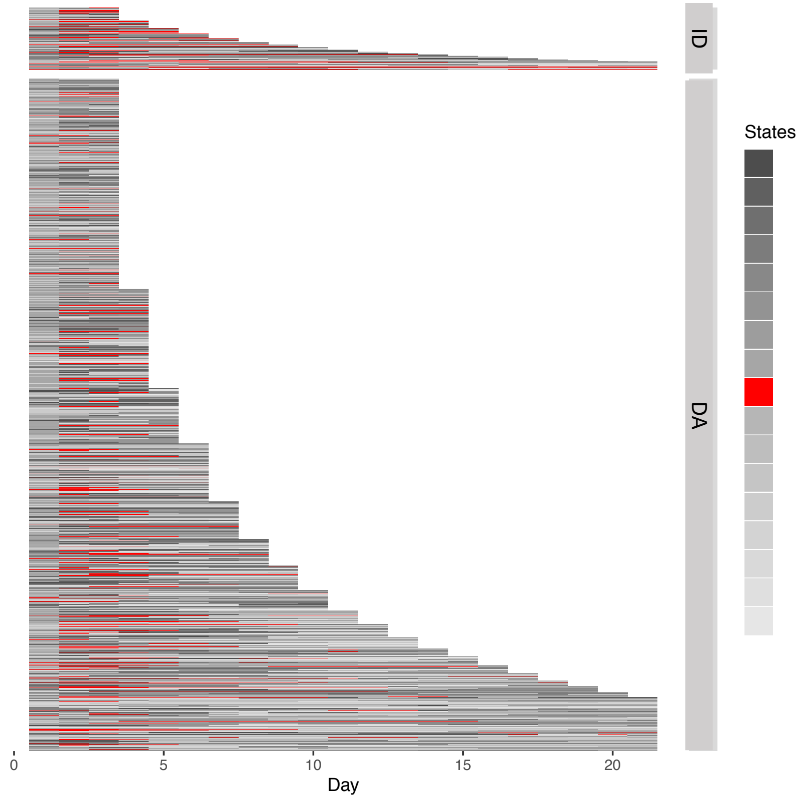


**Associated with:**

Day 2 of admission (can persist thereafter but usually resolved by discharge in long LOS - i.e., by around day 10)

Weakly with discharge as deceased

#### Distribution of Lab Tests and Vital Signs

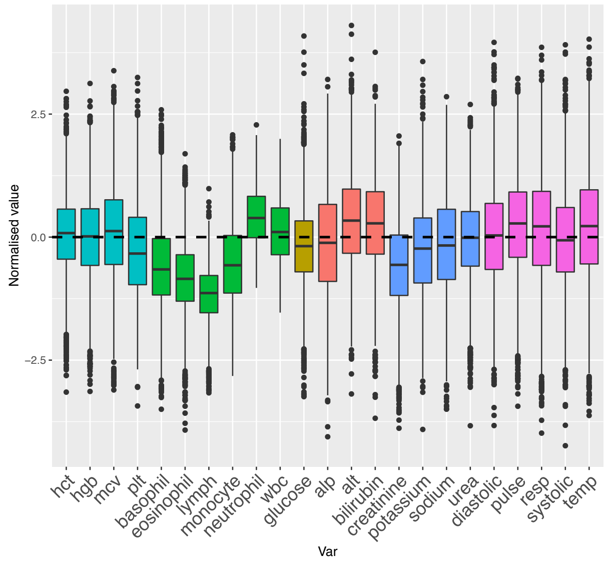


**Lower**: basophils, monocytes, eosinophils, lypmhs, creatinine;
**Higher**: neutrophils;
**Slightly higher**: pulse, resp, temp, alt, bilirubin

### EARLY DISCHARGE-LIKE STATE

#### AEs grouped by Primary Diagnosis

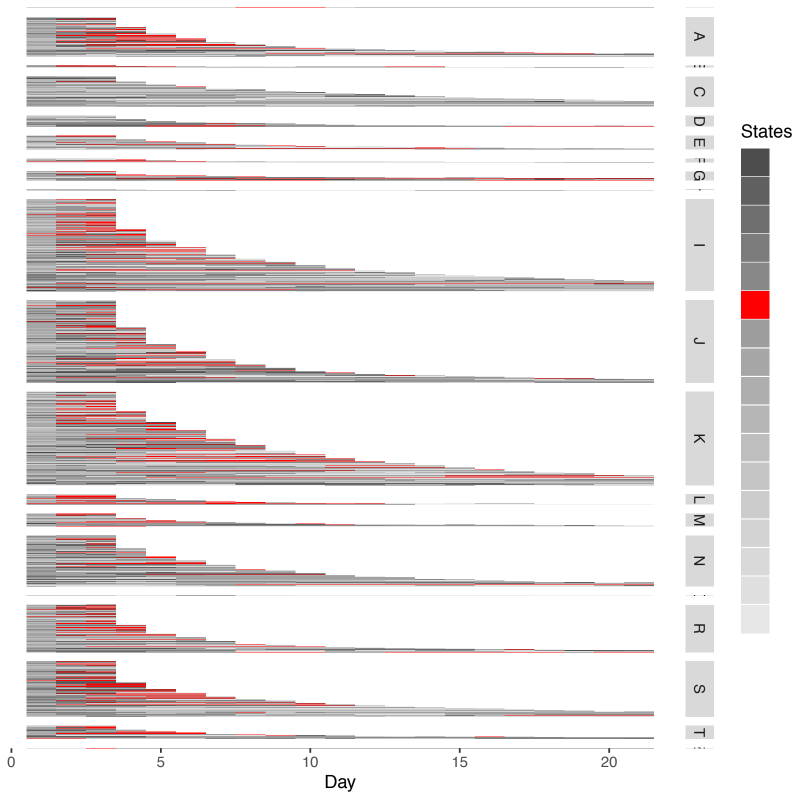


**Not associated with:**

Diagnostic codes;

#### AEs grouped by Inpatient Mortality

Inpatient Death (ID) or Discharged Alive (DA)

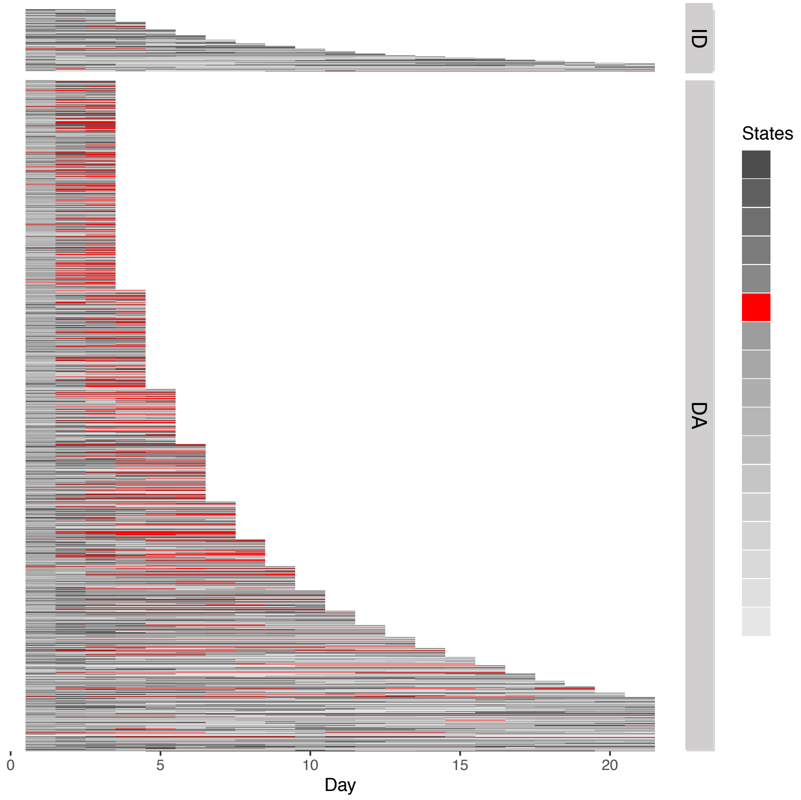


**Associated with:**

Shorter LOS (specially the last days of these admission episodes);
Discharge alive.

#### Distribution of Lab Tests and Vital Signs

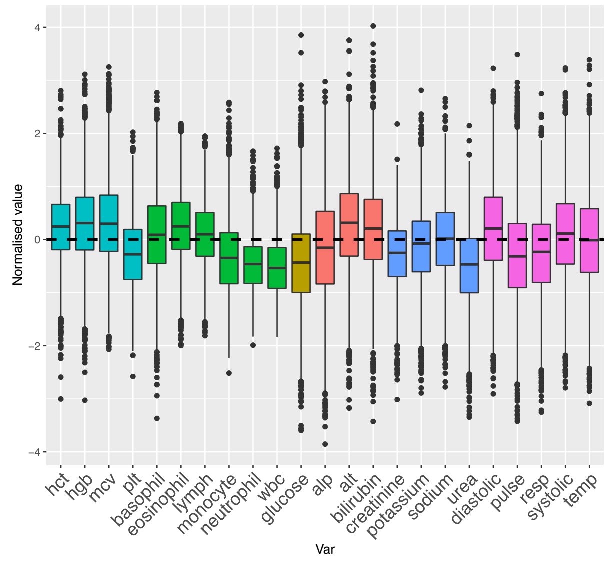


All parameters near group mean with some **slightly higher**: hgb, hct, alt, bilirubin;

**Slightly lower**: wbc, neutrophil, monocytes, glucose, urea, pulse, resp;

In most cases the parameters that are slightly higher or lower probably indicate the values are nearer a 'healthy population' norm than our 'unwell population norm'.

### PRE-DISCHARGE-LIKE STATE

#### AEs grouped by Primary Diagnosis

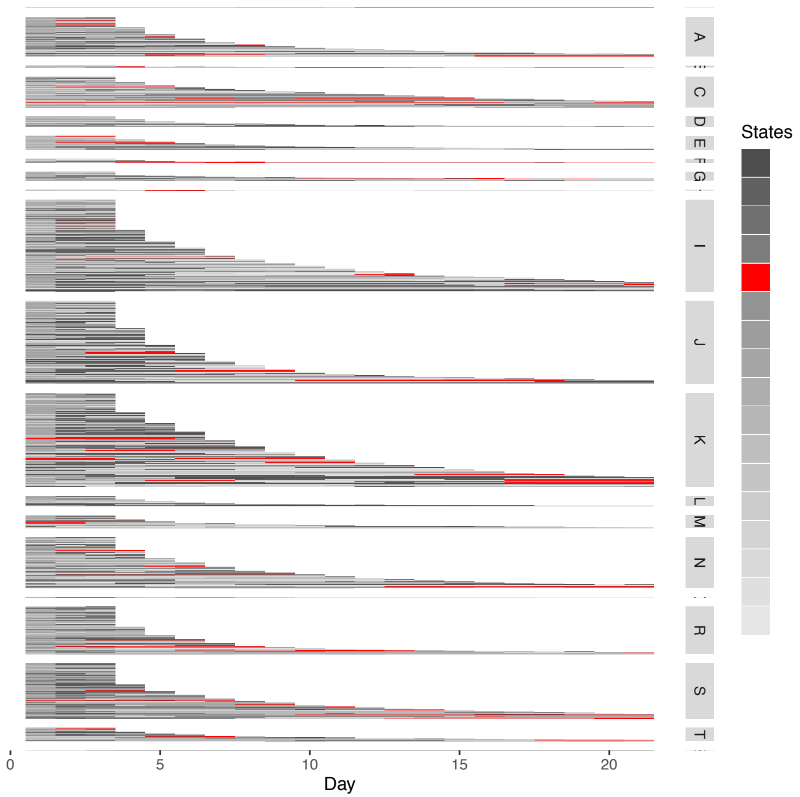


**Not associated with:**

Diagnostic codes;

#### AEs grouped by Inpatient Mortality

Inpatient Death (ID) or Discharged Alive (DA)

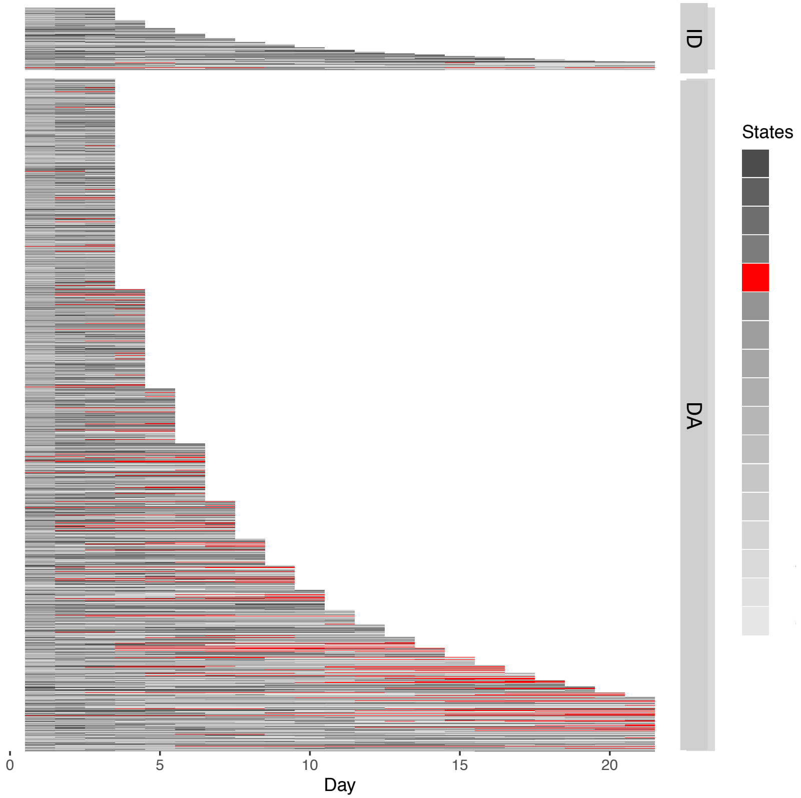


**Associated with:**

Discharged alive;
Predominantly in the last days of admission;
More common with long LOS.

#### Distribution of Lab Tests and Vital Signs

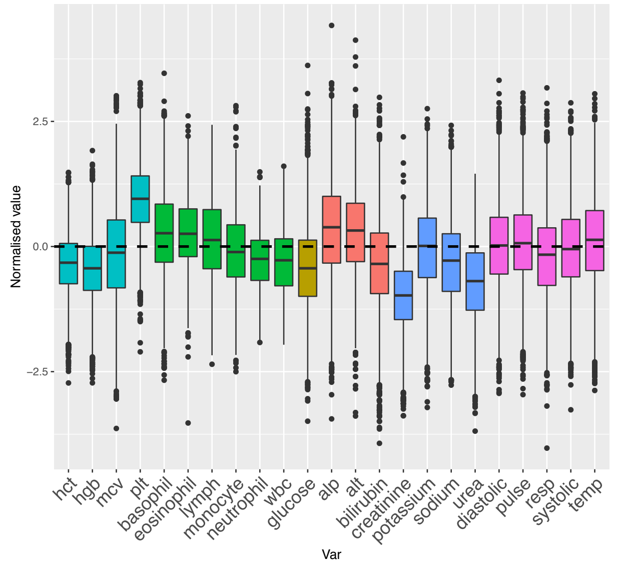


**Low**: hgb, hct, creatinine, urea, glucose

**High**: plt, alt, alp

Everything else quite **near group mea**n, especially Vital Signs.

### PHYSIOLOGIAL-LIKE STATES

- Early inflammatory response
- Resolving inflammatory response
- Autoimmune/ Atopic
- Acute thrombotic
- Prolonged illness
- Other illness presentation

### EARLY INFLAMMATORY RESPONSE-LIKE STATE

#### AEs grouped by Primary Diagnosis

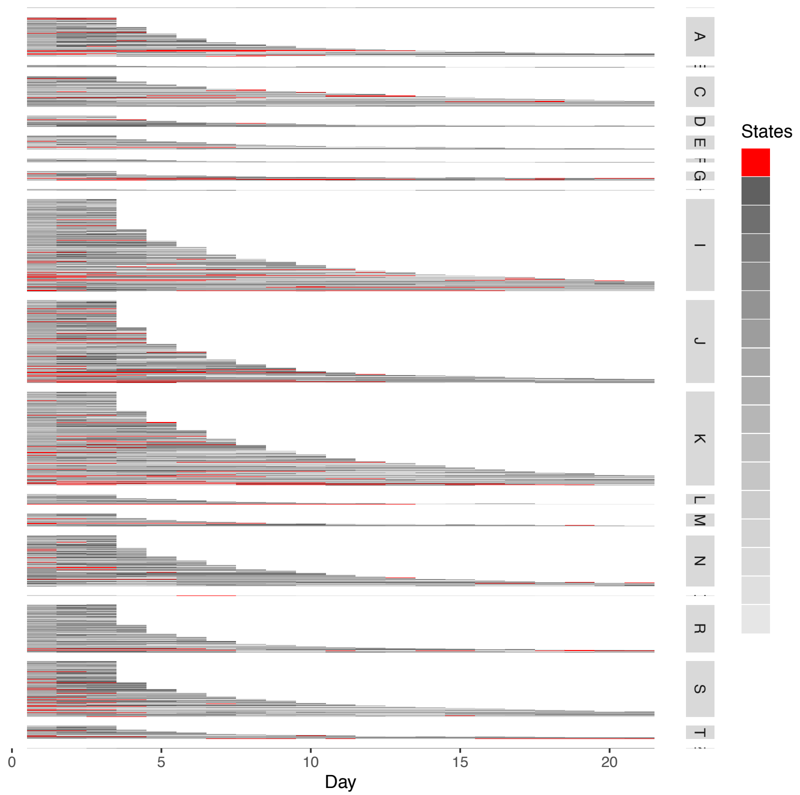


**Not associated with:**

Diagnostic codes

#### AEs grouped by Inpatient Mortality

Inpatient Death (ID) or Discharged Alive (DA)

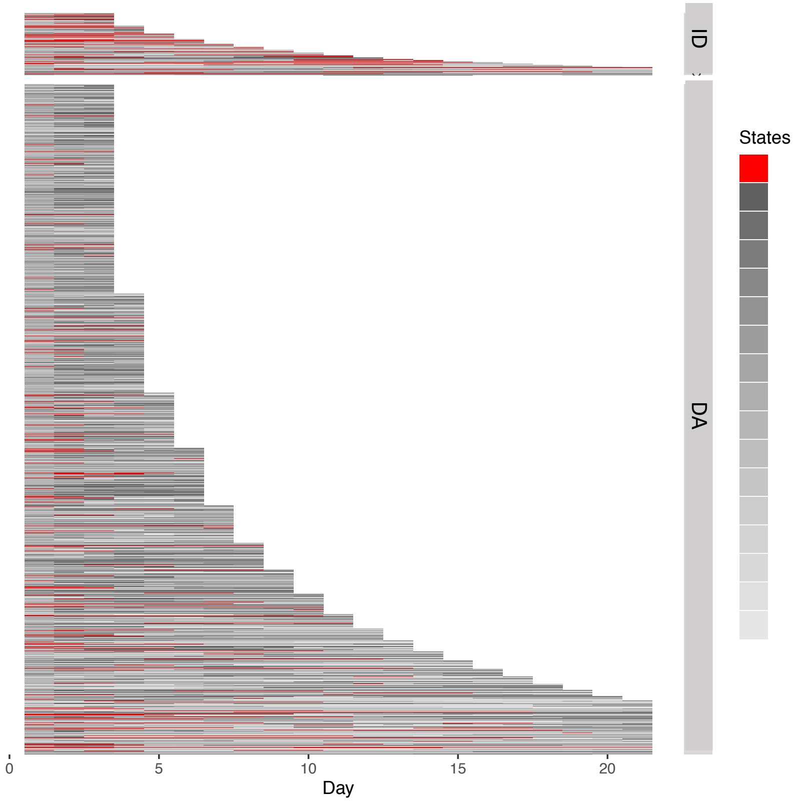


**Associated with:**

Discharged as deceased;

Tends to be present at beginning and resolve by discharge if outcome is discharged as alive.

#### Distribution of Lab Tests and Vital Signs

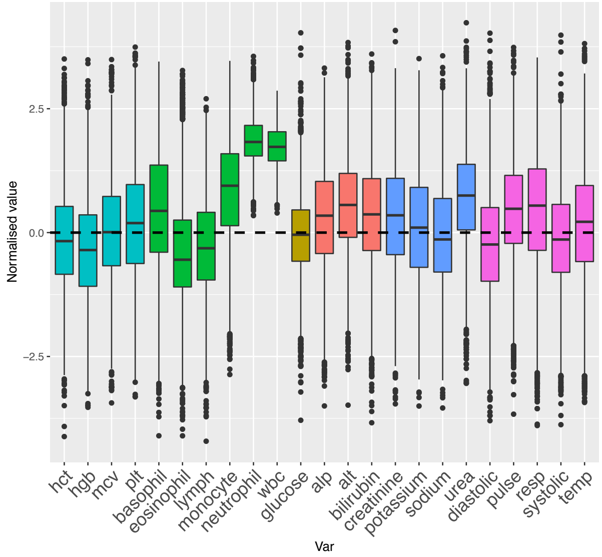


**Markedly high**: wbc, neutrophils;

**High**: monocytes, urea;

**Relatively high**: pulse, resp, liver function tests, creatinine

**Slightly high**: temp

**Mildly low**: hgb, hct, eosinophils, lymph, diastolic and systolic BP

### RESOLVING INFLAMMATORY RESPONSE-LIKE STATE

#### AEs grouped by Primary Diagnosis

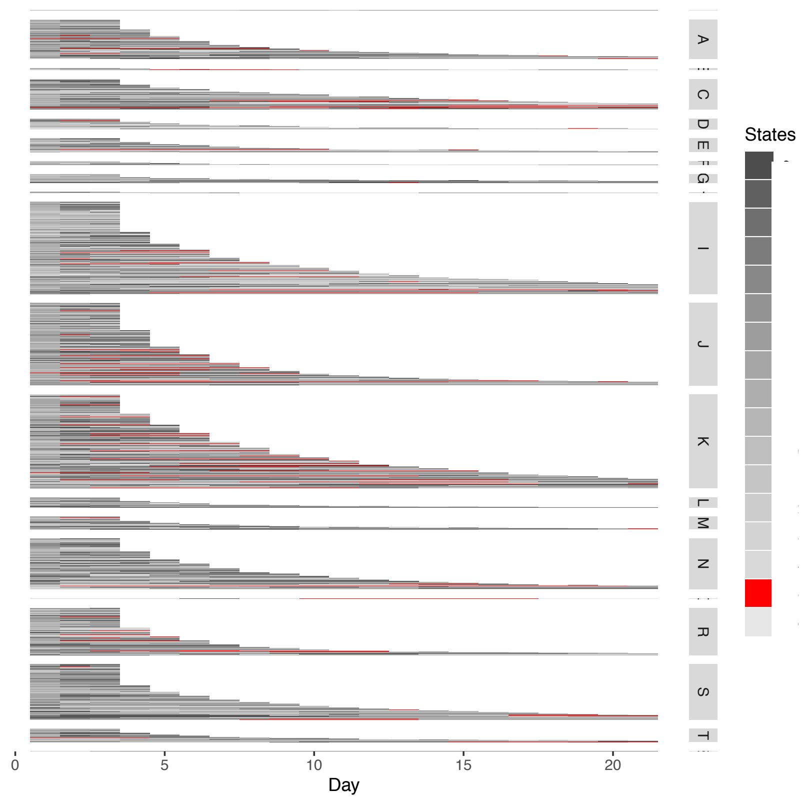


**Associated with:**
A: *Infectious and parasitic diseases*;
C: *Neoplasms*;
I: *Circulatory system*;
J: *Respiratory system*;
K: *Digestive system.*

#### AEs grouped by Inpatient Mortality

Inpatient Death (ID) or Discharged Alive (DA)

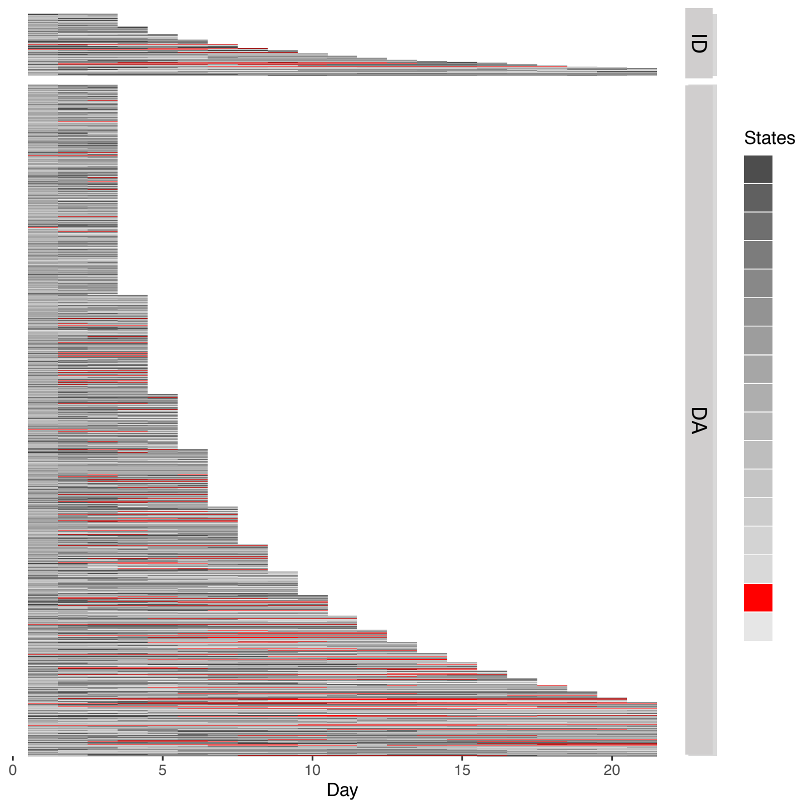


**Associated with:**
Discharge alive (but weak and mostly due to presence in long LOS);
Long LOS: tends to be present after day 1 and can persist until discharge.

#### Distribution of Lab Tests and Vital Signs

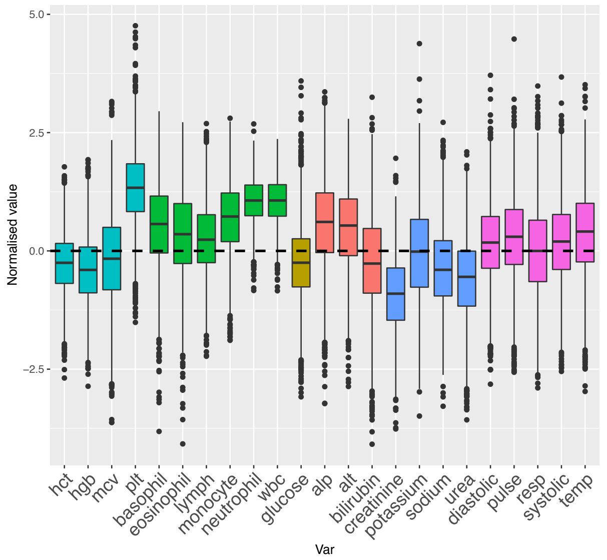


**Higher**: wbc, neutrophils (with small variance);

**Higher:** plt, monocytes, alp, alt (wider variance);

**Slightly high**: Vitals Signs (except resp) and other white cells;

**Lower**: hgb, hct, creatinine, sodium.

### AUTOIMMUNE/ ATOPIC-LIKE STATE

#### AEs grouped by Primary Diagnosis

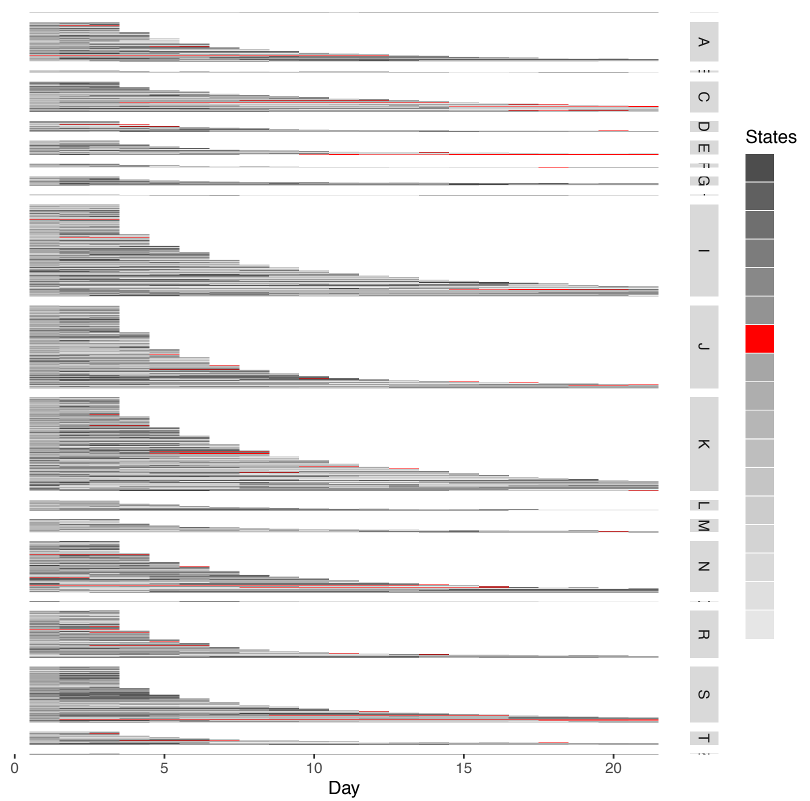


**Not associated with:**

Diagnostic codes

#### AEs grouped by Inpatient Mortality

Inpatient Death (ID) or Discharged Alive (DA)

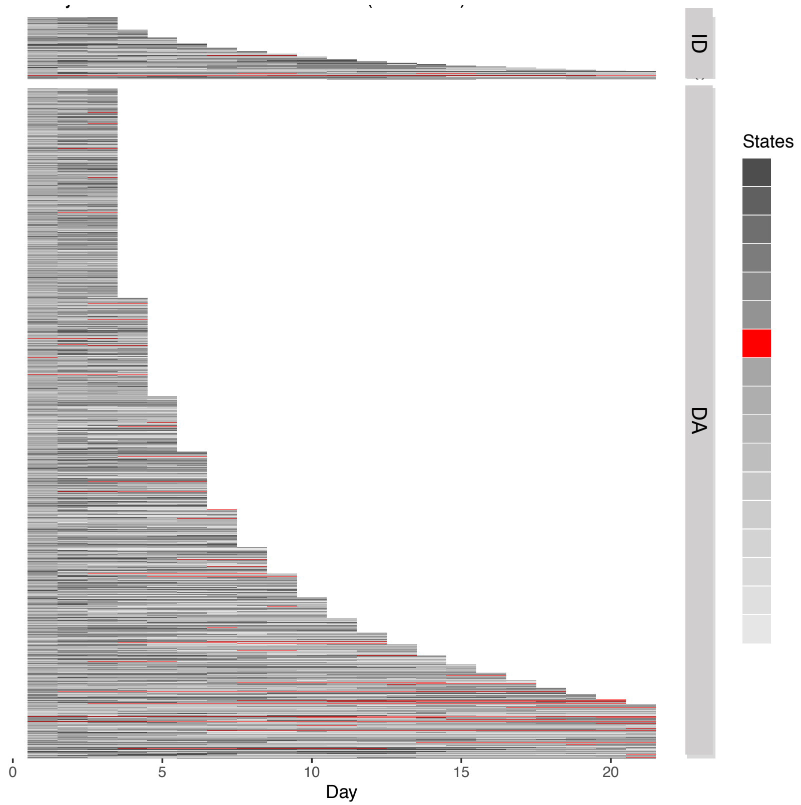


**Associated with:**

Discharged alive;

Overall not very common but seen more frequently in patients with long LOS.

#### Distribution of Lab Tests and Vital Signs

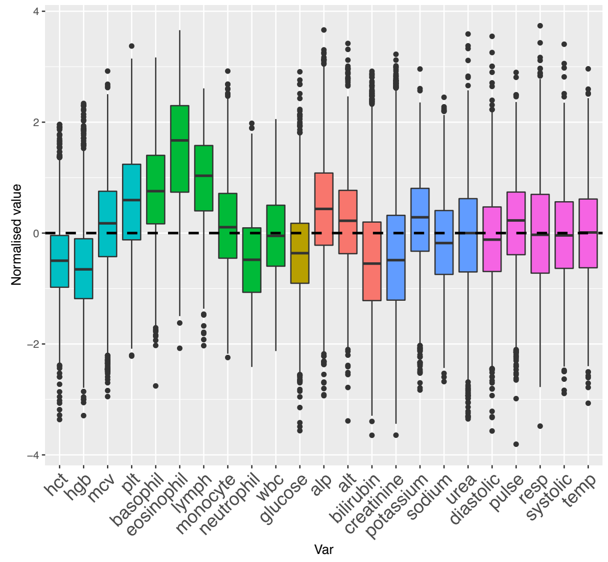


**Markedly higher:** basophils, eoisinophils and lymphs;

**High**: plts;

**Lower:** hgb, hct, neutrophils, glucose;

**Slightly low**: bilirubin, creatinine.

### ACUTE THROMBOTIC-LIKE STATE

#### AEs grouped by Primary Diagnosis

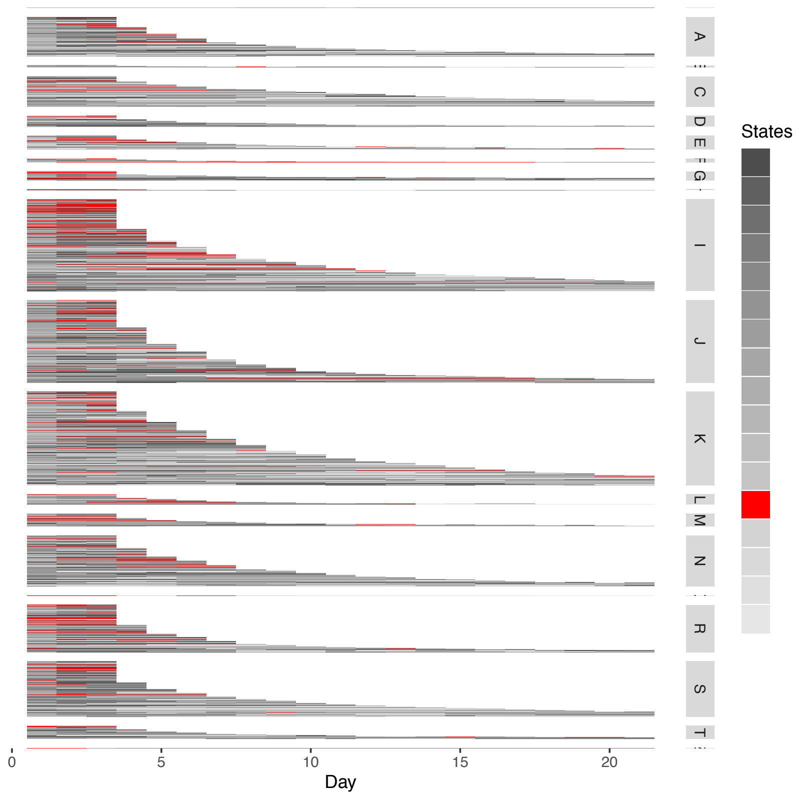


**Associated with:**

I: *Circulatory system*;

R: *Symptoms, signs and abnormal clinical and laboratory findings*;

Also seen to some extent in most other diagnostic categories;

#### AEs grouped by Inpatient Mortality

Inpatient Death (ID) or Discharged Alive (DA)

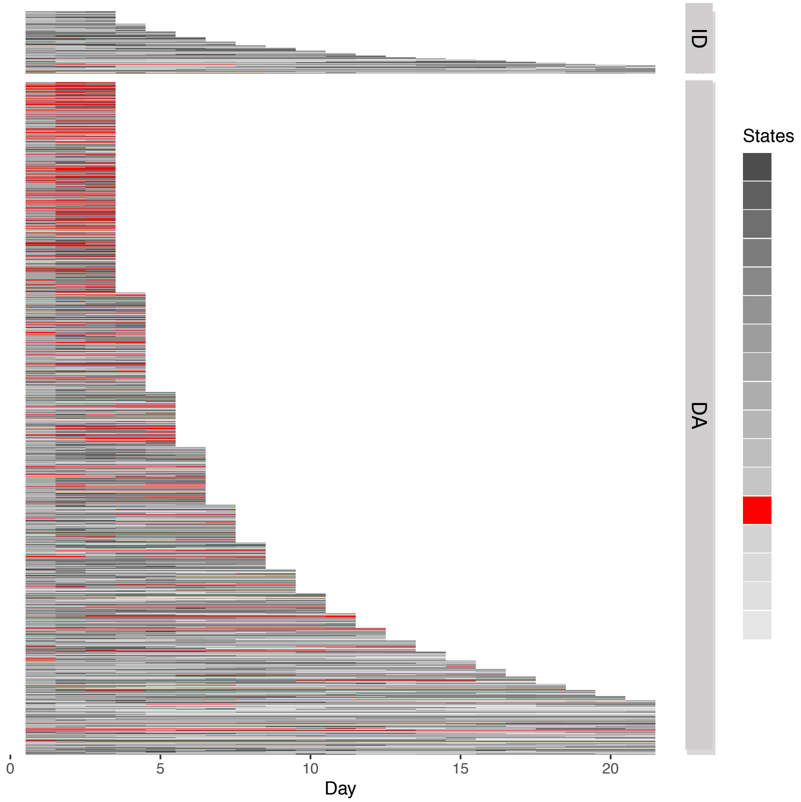


**Associated with:**

Short / medium LOS (common from first day onwards);

Discharge alive.

#### Distribution of Lab Tests and Vital Signs

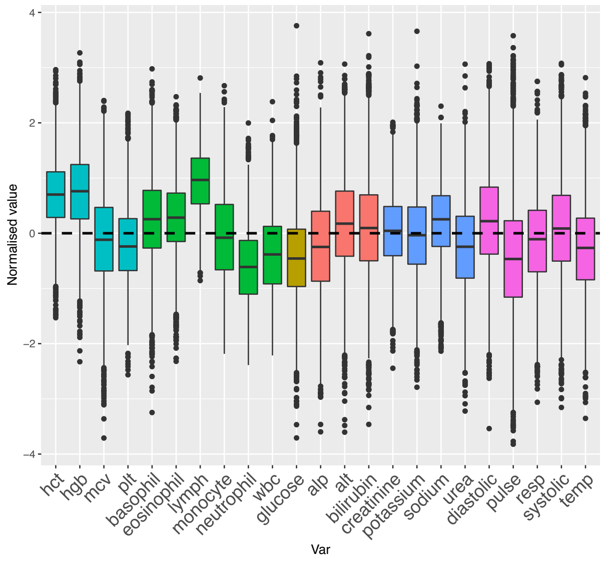


**Higher**: hgb, hct, lypmph;

**Lower**: neutrophils, wbc, glucose, pulse

**Mildly low**: temp

### PROLONGED ILLNESS-LIKE STATE

#### AEs grouped by Primary Diagnosis

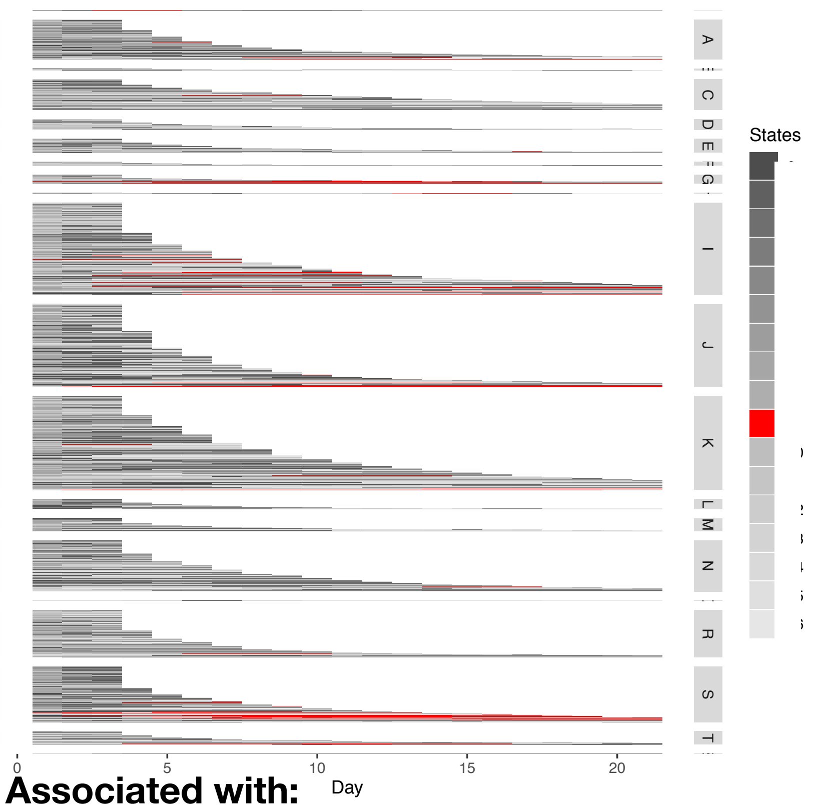


**Associated with:**

I: *Circulatory system*;
S: *Injury, poisoning and other external causes*;
Also G (*Nerv system*), J (*Res system*), K (*Digestive system*);
Notable that it is not in N (*Genitourinary system*) & R (*Symptoms, signs and abnormal clinical and lab findings*) even if long LOS

#### AEs grouped by Inpatient Mortality

Inpatient Death (ID) or Discharged Alive (DA)

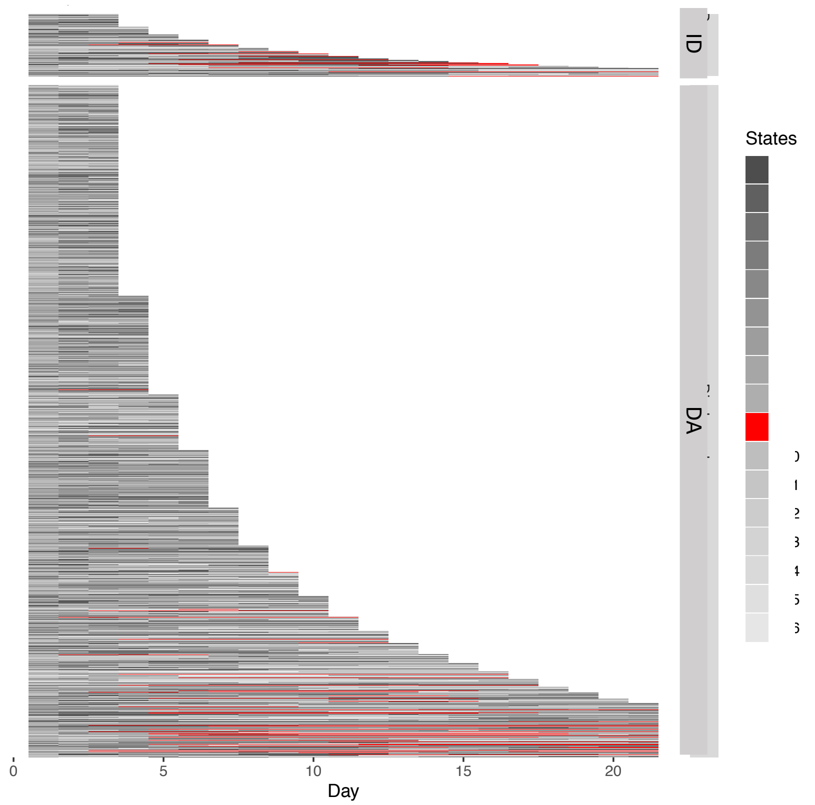


**Associated with:**

Long LOS and latter stages of admission episodes (never in first few days)

**Not associated with:**

Discharge status;

#### Distribution of Lab Tests and Vital Signs

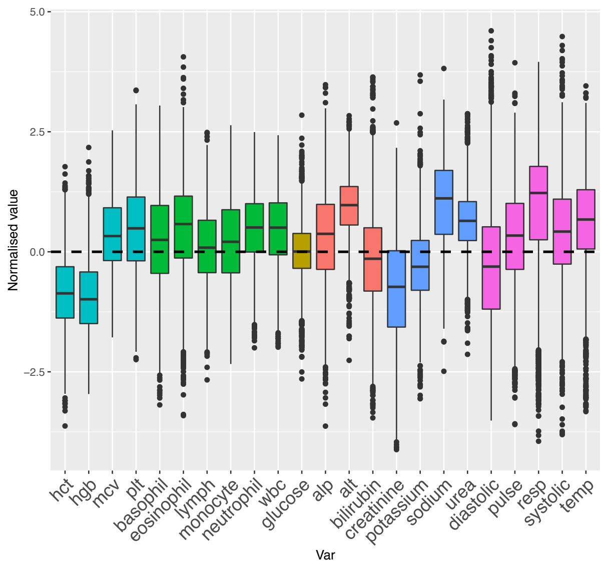


**Lower**: hgb, hct, creatinine;
**Higher**: mcv, plt, wbc, neutrophils, eosinophils, alt, sodium, urea, resp.
**Mildly high**: other Vital Signs (except diastolic BP)

### OTHER ILLNESS PRESENTATION-LIKE STATE

#### AEs grouped by Primary Diagnosis

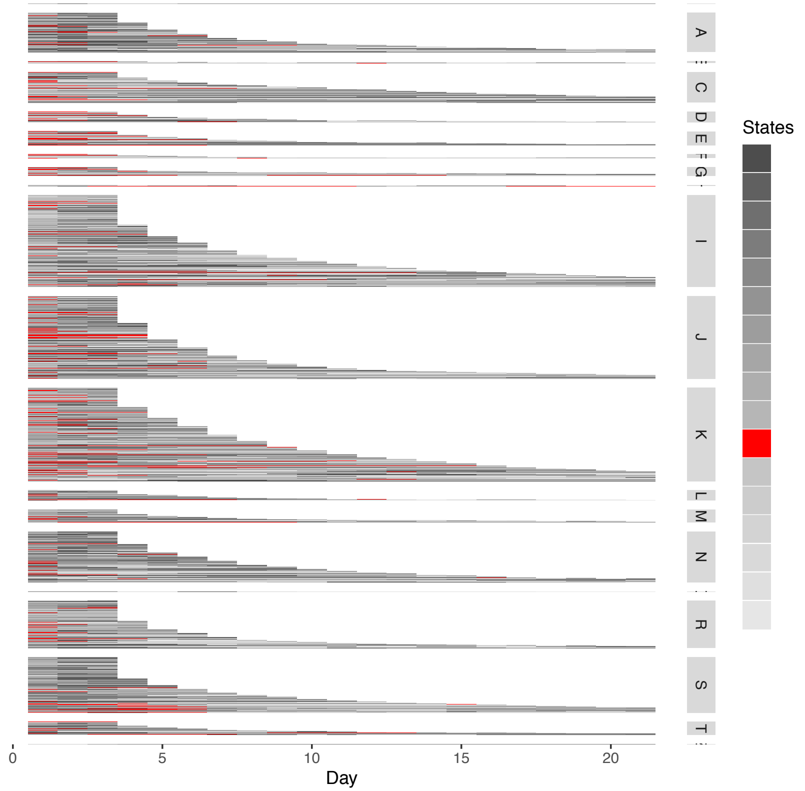


**Associated with:**

Not very clear, but maybe with:
E: *Endocrine, metabolic and nutritional diseases;*
D: *Blood and blood-forming organs and certain disorders involving the immune mechanism*;

#### AEs grouped by Inpatient Mortality

Inpatient Death (ID) or Discharged Alive (DA)

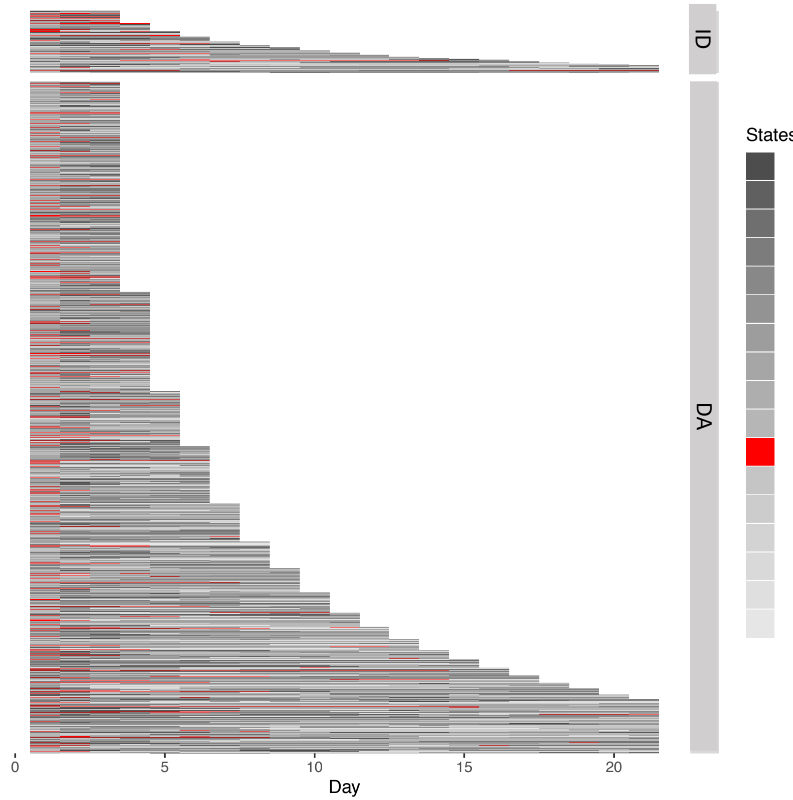


**Associated with:**

Early-mid stage of admission (rarely at the end of the admission unless very short);

**Not associated with:**

Discharge status

#### Distribution of Lab Tests and Vital Signs

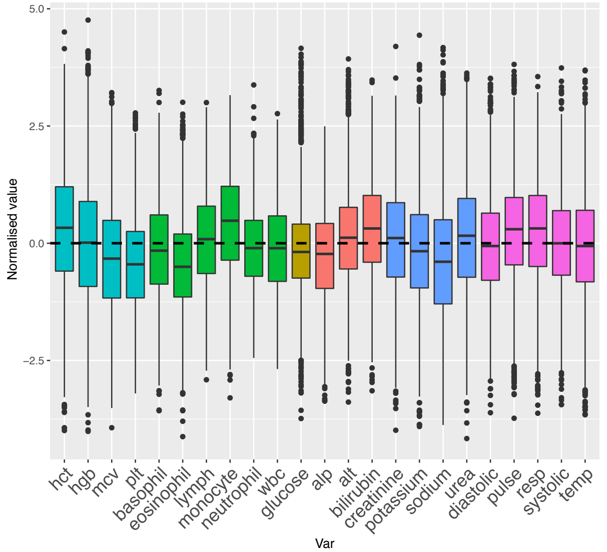


Most obvious feature is ‘normality’: no stand out derangement of Vital signs or Lab Tests

### List of abbreviations & ICD-10 codes

LOS: length of stay

LFTs: liver function tests

FBC: full blood count

WBC: white blood cells

MCV: mean cell volume

#### ICD-10 CODES

**A & B** – Infectious and parasitic diseases
**C & D** – Neoplasms
**D** – Diseases of the blood and blood-forming organs and certain disorders involving the immune mechanism
**E** – Endocrine, nutritional and metabolic diseases
**F** – Mental and behavioural disorders
**G** – Diseases of the Nervous System
**H** – Diseases of the eye and adnexa, the ear and mastoid process
**I** – Diseases of the circulatory system
**J** – Diseases of the respiratory system
**K** – Diseases of the digestive system
**L** – Skin and subcutaneous tissue
**M** – Musculoskeletal system and connective tissue
**N** – Genitourinary system
**P** – Conditions originating in the perinatal period
**Q** - Congenital malformations, deformations and chromosomal abnormalities
**R** – Symptoms, signs and abnormal clinical and laboratory findings, not elsewhere classified
**S & T** – Injury, poisoning and certain other consequences of external causes
**Z** – Factors influencing health status and contact with health services
