## Supplementary Figures 6 for "Big Data Analysis of Electronic Health Records: Clinically interpretable representations of older adult inpatient trajectories using time-series numerical data and Hidden Markov Models"

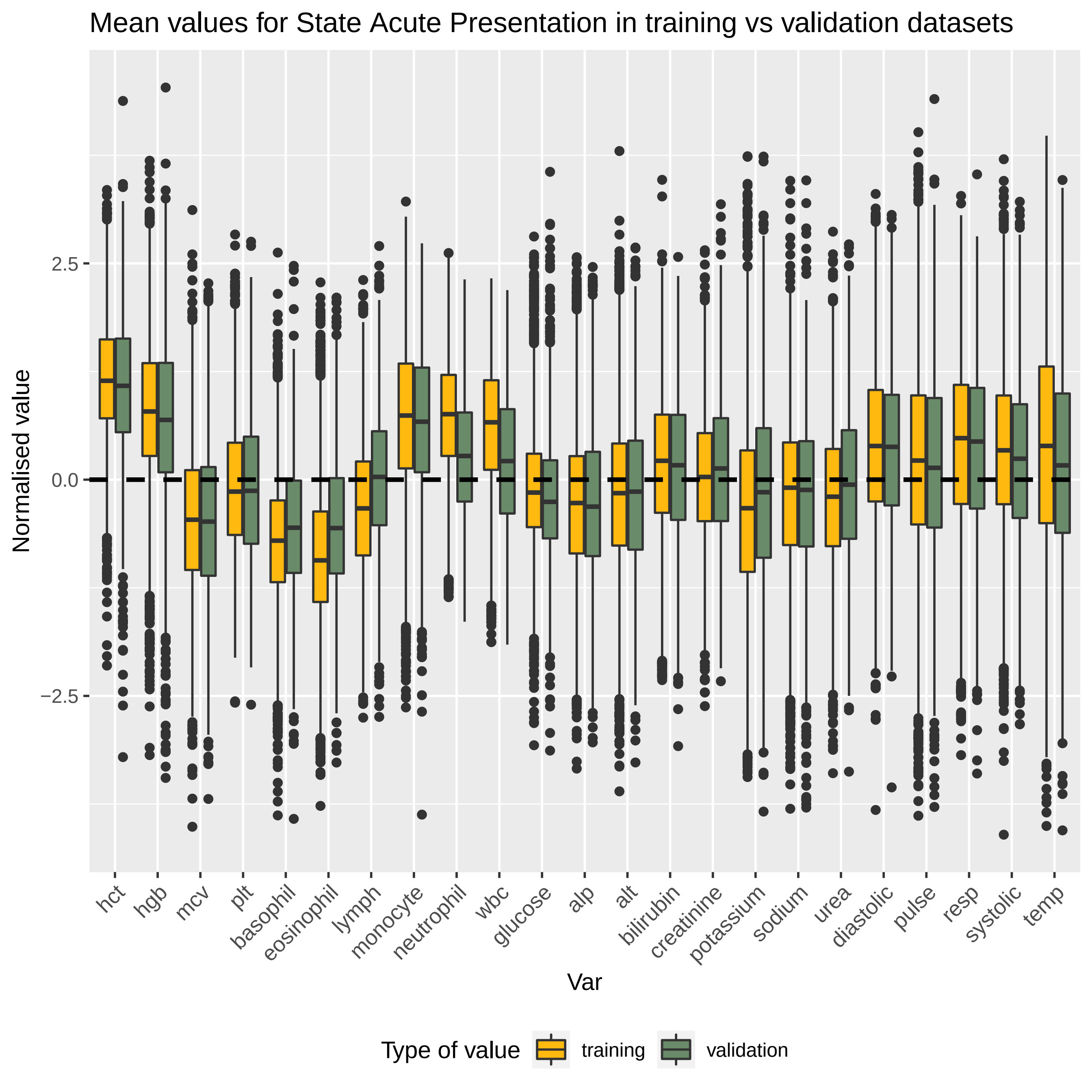

Mean values for State Acute Presentation in training vs validation datasets
Normalised value
2.5
0.0
−2.5
hct
hgb
mcv
plt
basophil
eosinophil
lymph
monocyte
neutrophil
wbc
glucose
alp
alt
bilirubin
creatinine
potassium
sodium
urea
diastolic
pulse
resp
systolic
temp
Var
Type of value
training
validation

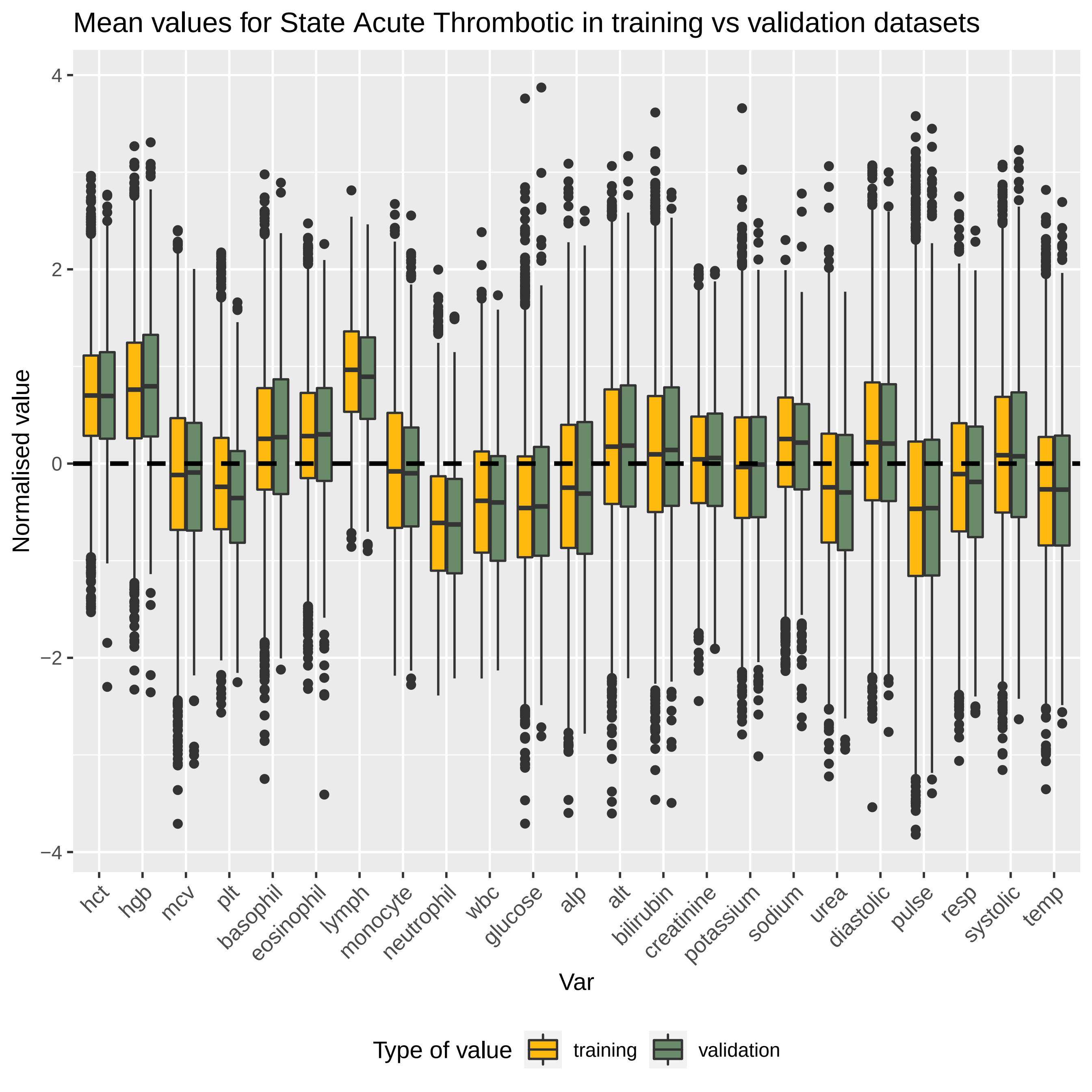
Mean values for State Acute Thrombotic in training vs validation datasets
Normalised value
4
2
0
−2
−4
hct
hgb
mcv
plt
basophil
eosinophil
lymph
monocyte
neutrophil
wbc
glucose
alp
alt
bilirubin
creatinine
potassium
sodium
urea
diastolic
pulse
resp
systolic
temp
Var
Type of value
training
validation

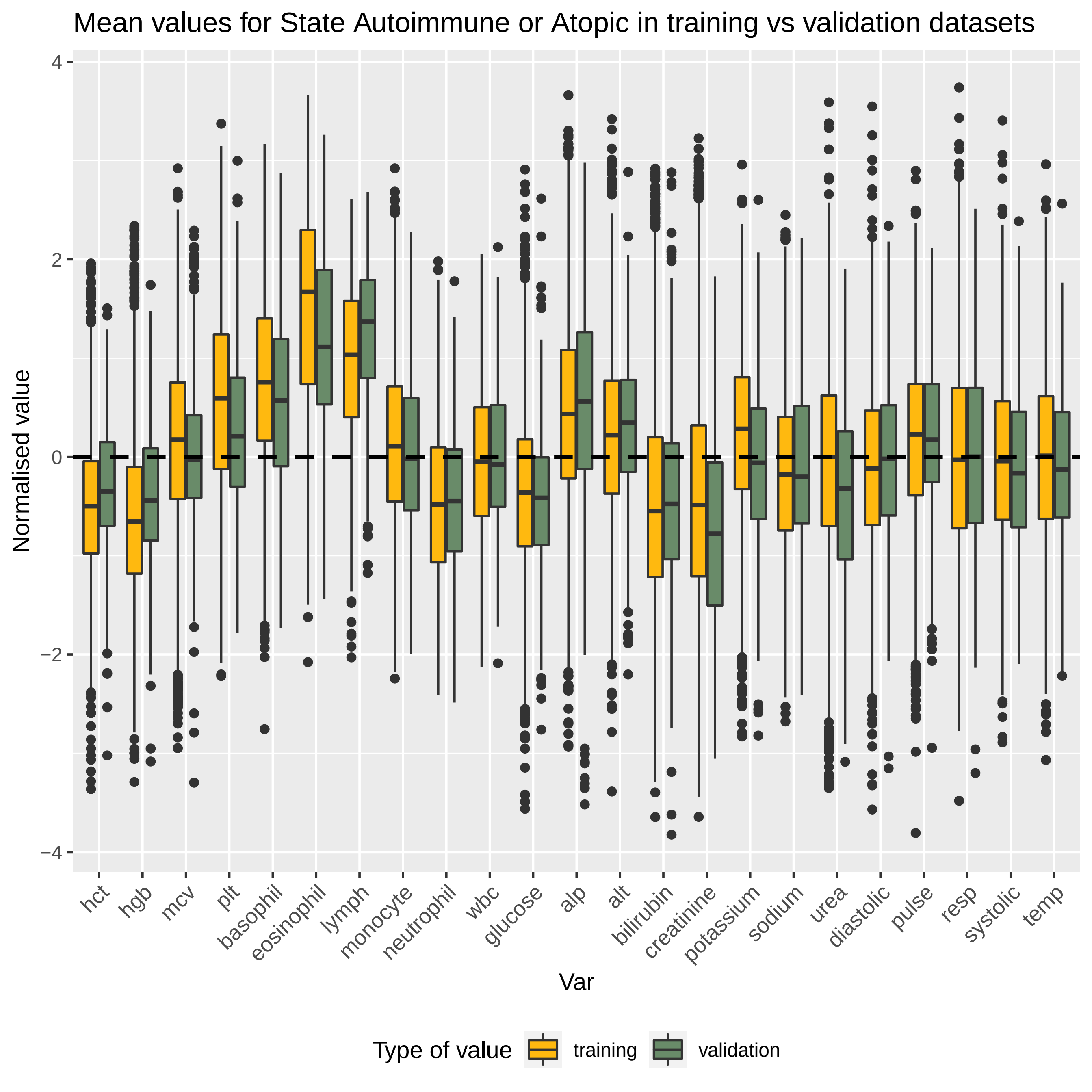

Mean values for State Autoimmune or Atopic in training vs validation datasets
Normalised value
4
2
0
−2
−4
hct
hgb
mcv
plt
basophil
eosinophil
lymph
monocyte
neutrophil
wbc
glucose
alp
alt
bilirubin
creatinine
potassium
sodium
urea
diastolic
pulse
resp
systolic
temp
Var
Type of value
training
validation

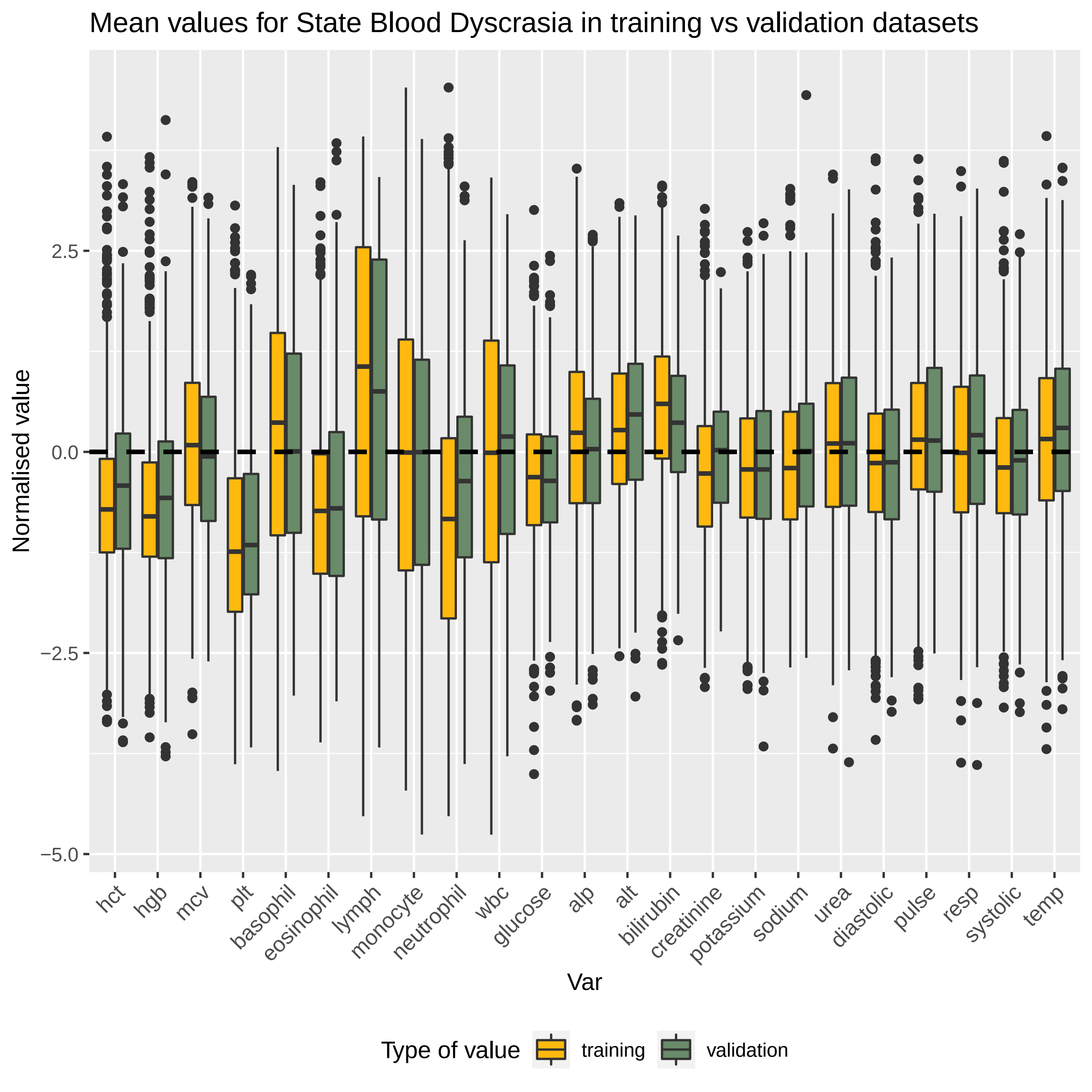
Mean values for State Blood Dyscrasia in training vs validation datasets
Normalised value
2.5
0.0
−2.5
−5.0
hct
hgb
mcv
plt
basophil
eosinophil
lymph
monocyte
neutrophil
wbc
glucose
alp
alt
bilirubin
creatinine
potassium
sodium
urea
diastolic
pulse
resp
systolic
temp
Var
Type of value
training
validation

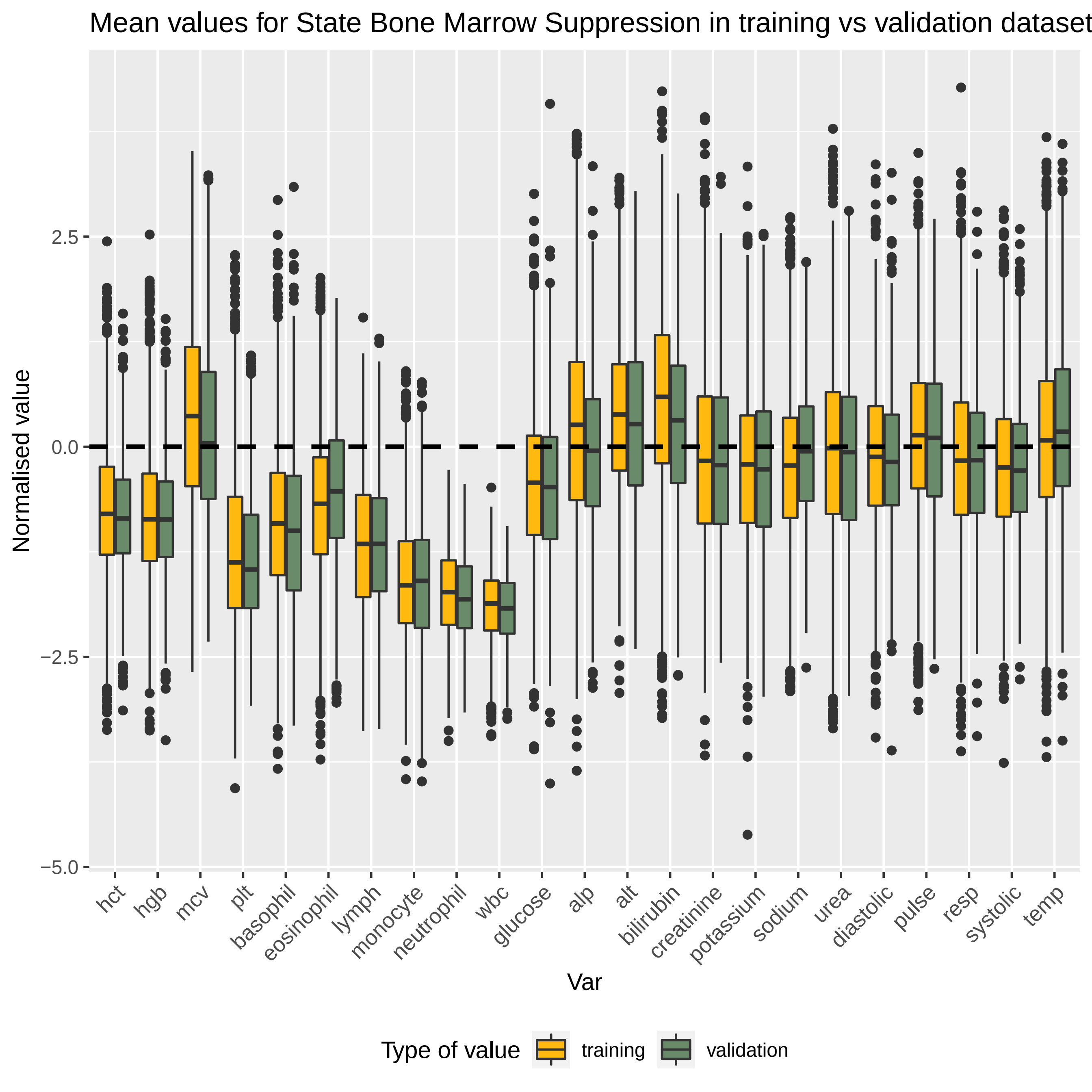

Mean values for State Bone Marrow Suppression in training vs validation dataset
Normalised value
2.5
0.0
−2.5
−5.0
hct
hgb
mcv
plt
basophil
eosinophil
lymph
monocyte
neutrophil
wbc
glucose
alp
alt
bilirubin
creatinine
potassium
sodium
urea
diastolic
pulse
resp
systolic
temp
Var
Type of value
training
validation

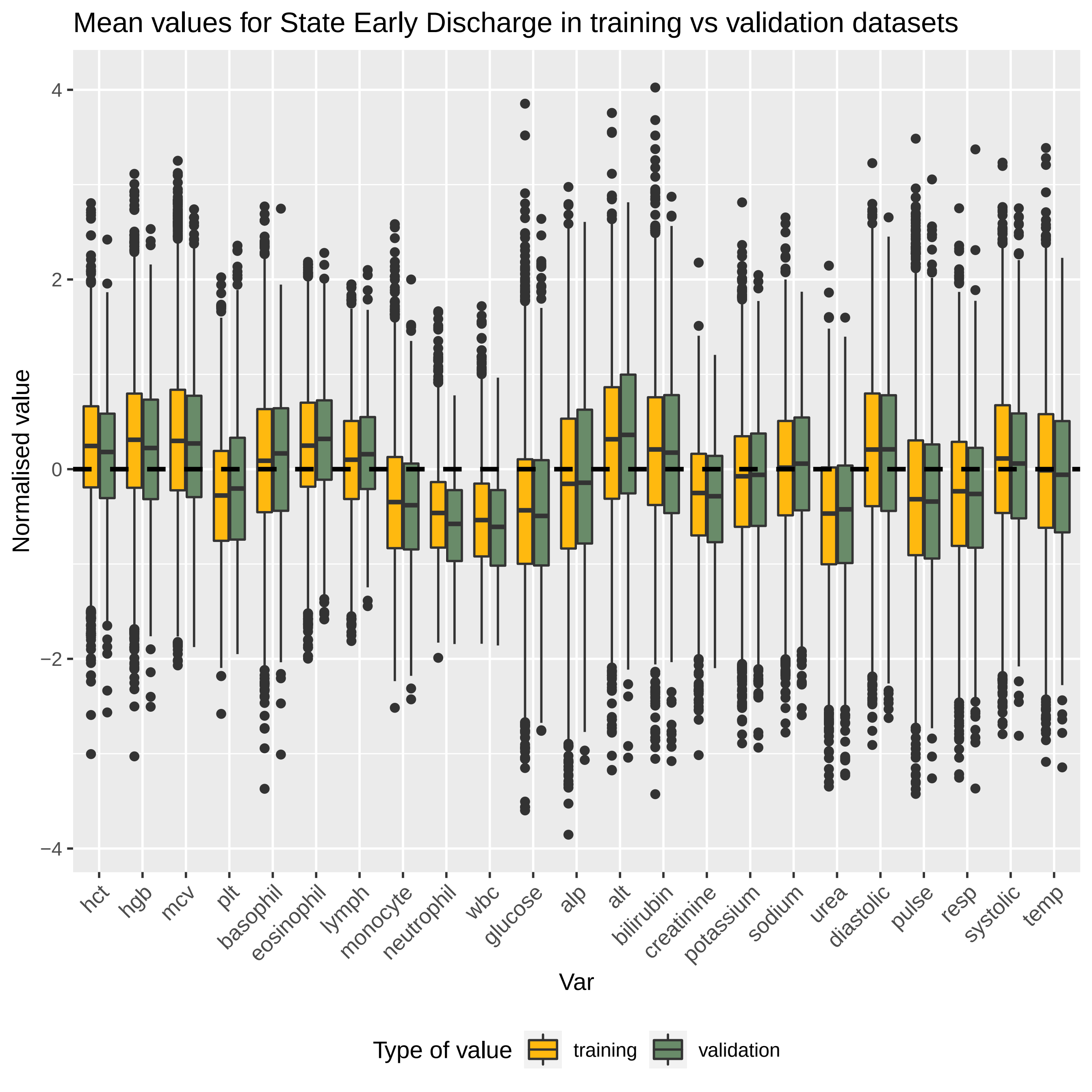
Mean values for State Early Discharge in training vs validation datasets
Normalised value
4
2
0
−2
−4
hct
hgb
mcv
plt
basophil
eosinophil
lymph
monocyte
neutrophil
wbc
glucose
alp
alt
bilirubin
creatinine
potassium
sodium
urea
diastolic
pulse
resp
systolic
temp
Var
Type of value
training
validation

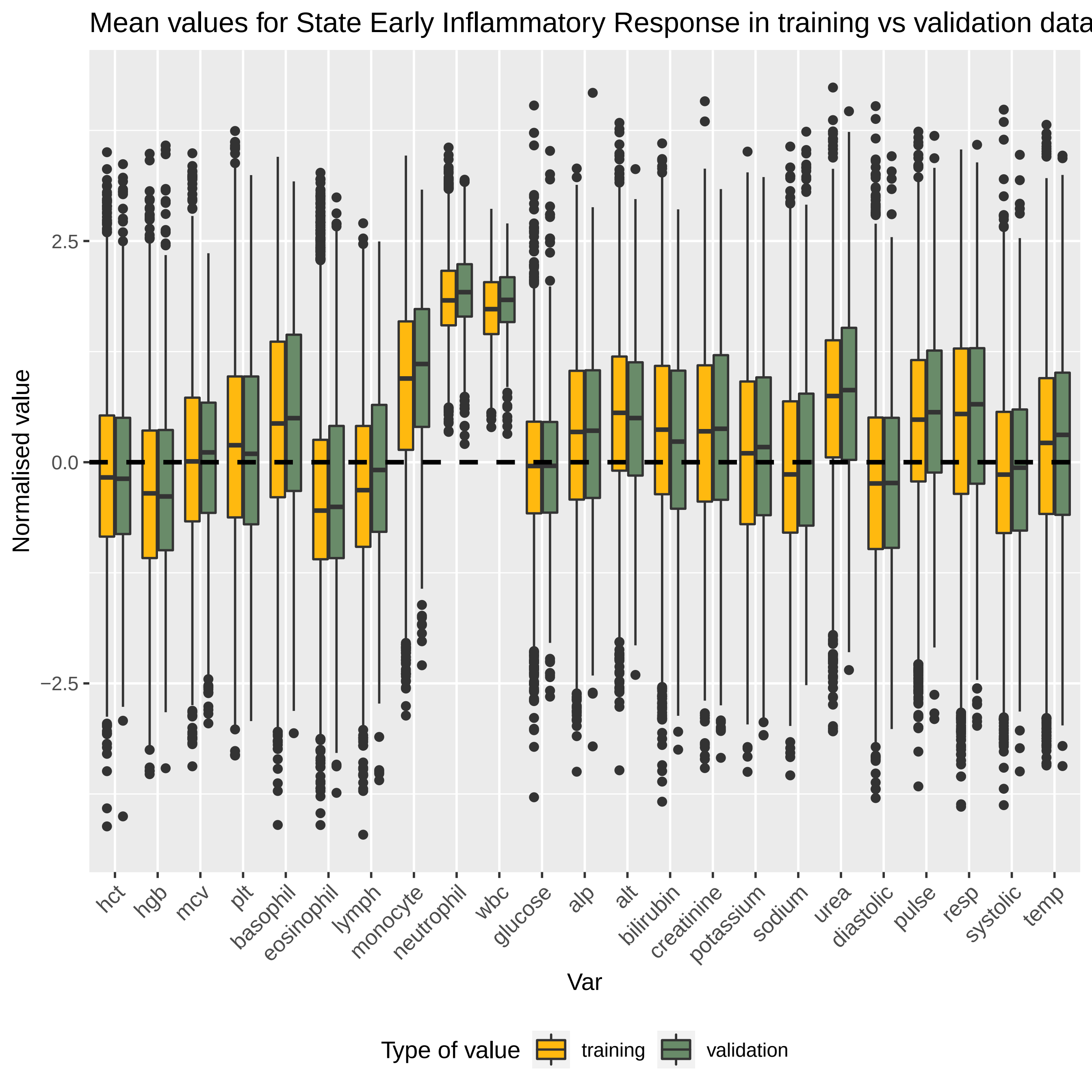

Mean values for State Early Inflammatory Response in training vs validation data
Normalised value
2.5
0.0
−2.5
hct
hgb
mcv
plt
basophil
eosinophil
lymph
monocyte
neutrophil
wbc
glucose
alp
alt
bilirubin
creatinine
potassium
sodium
urea
diastolic
pulse
resp
systolic
temp
Var
Type of value
training
validation

Mean values for State Hepatic in training vs validation datasets

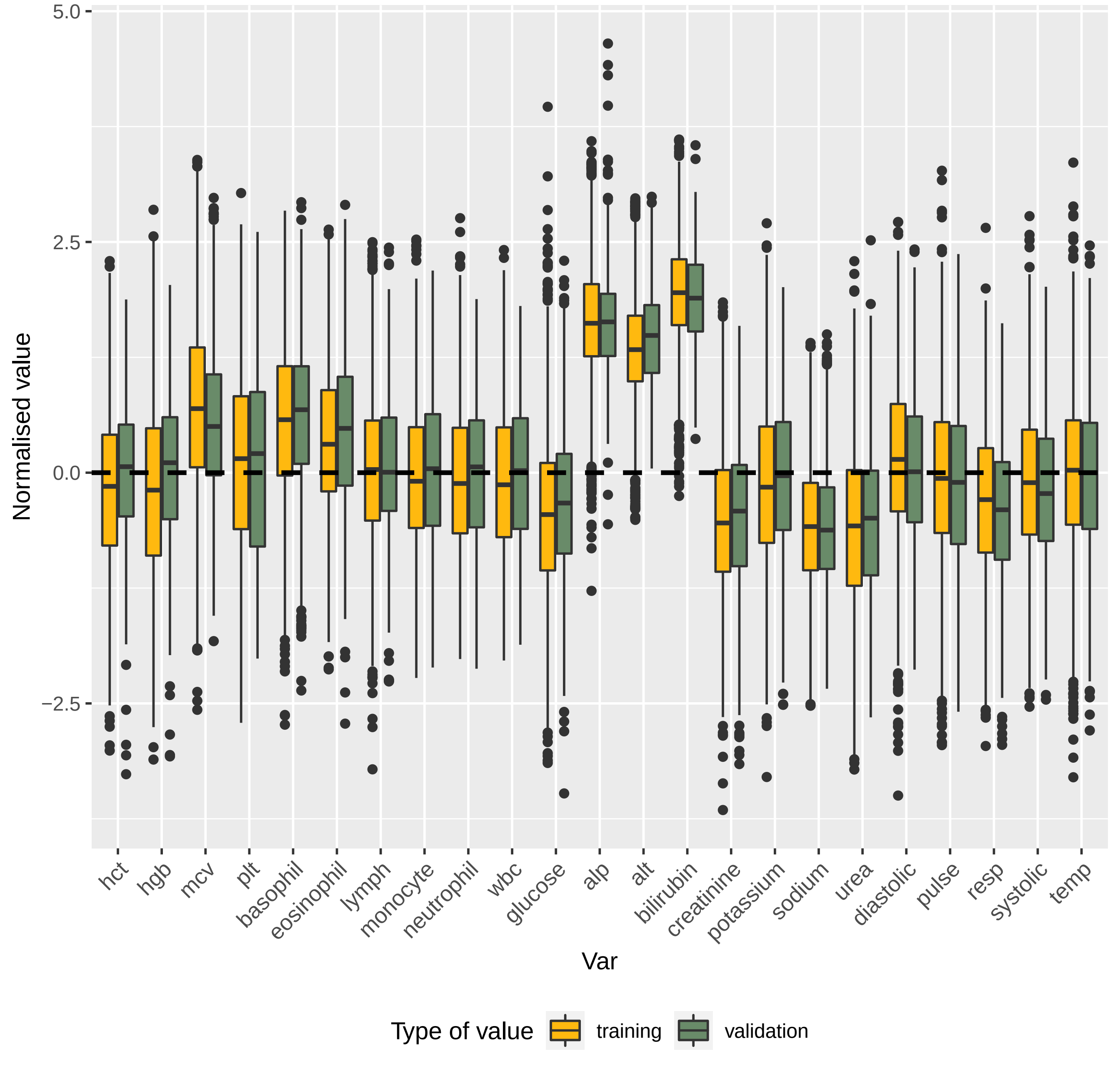

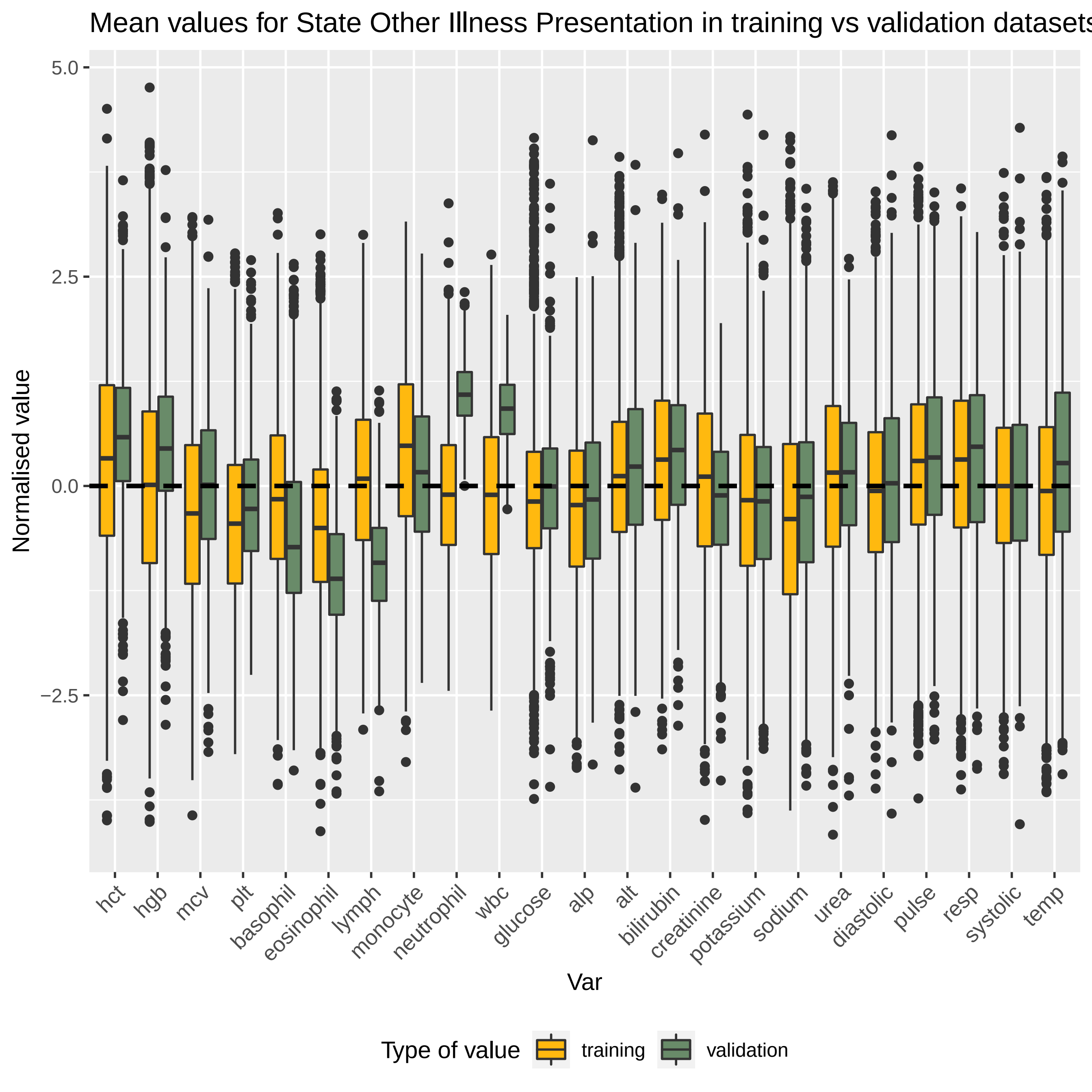
Mean values for State Other Illness Presentation in training vs validation datasets
Normalised value
5.0
2.5
0.0
-2.5
hct
hgb
mcv
plt
basophil
eosinophil
lymph
monocyte
neutrophil
wbc
glucose
alp
alt
bilirubin
creatinine
potassium
sodium
urea
diastolic
pulse
resp
systolic
temp
Var
Type of value
training
validation

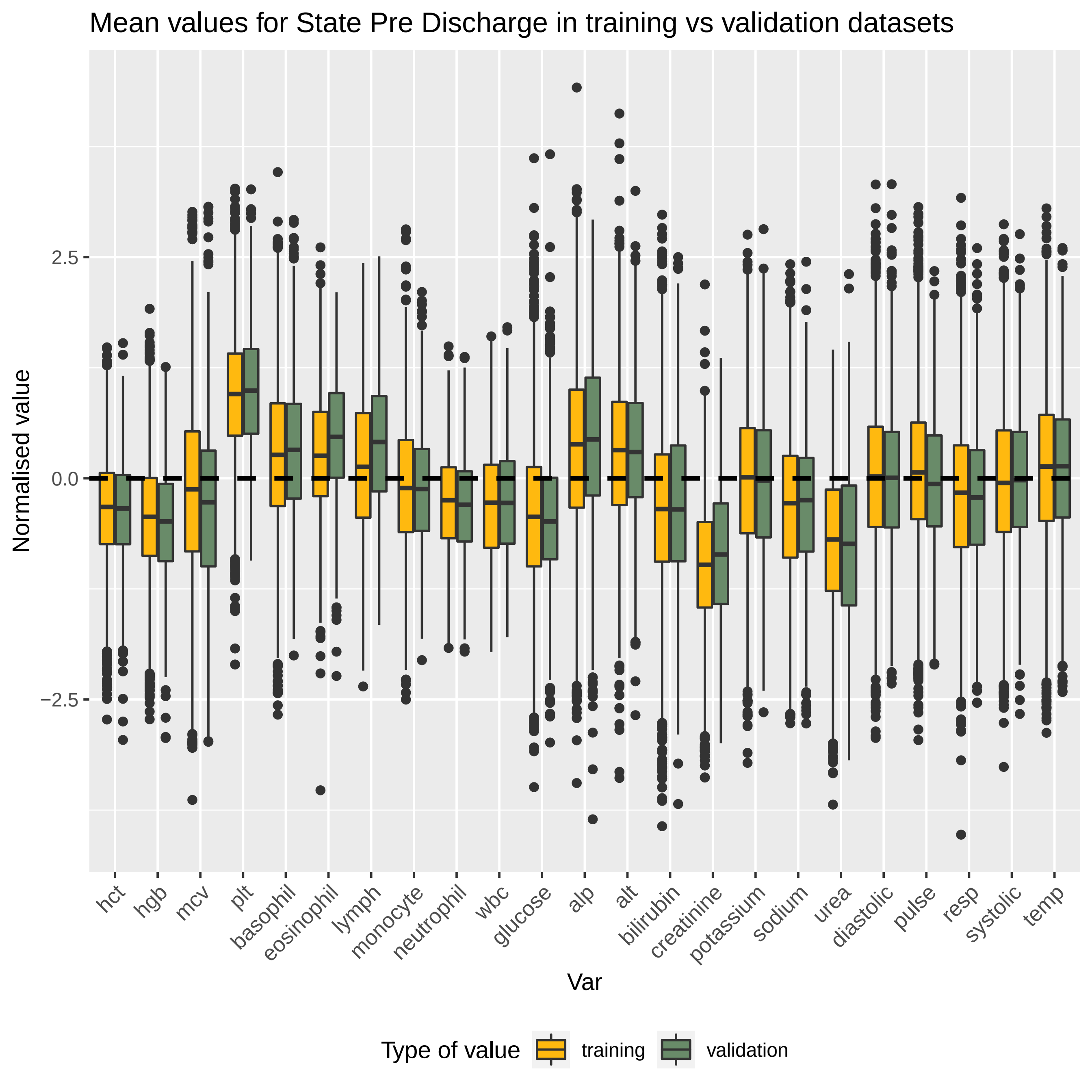

Mean values for State Pre Discharge in training vs validation datasets
Normalised value
2.5
0.0
−2.5
hct
hgb
mcv
plt
basophil
eosinophil
lymph
monocyte
neutrophil
wbc
glucose
alp
alt
bilirubin
creatinine
potassium
sodium
urea
diastolic
pulse
resp
systolic
temp
Var
Type of value
training
validation

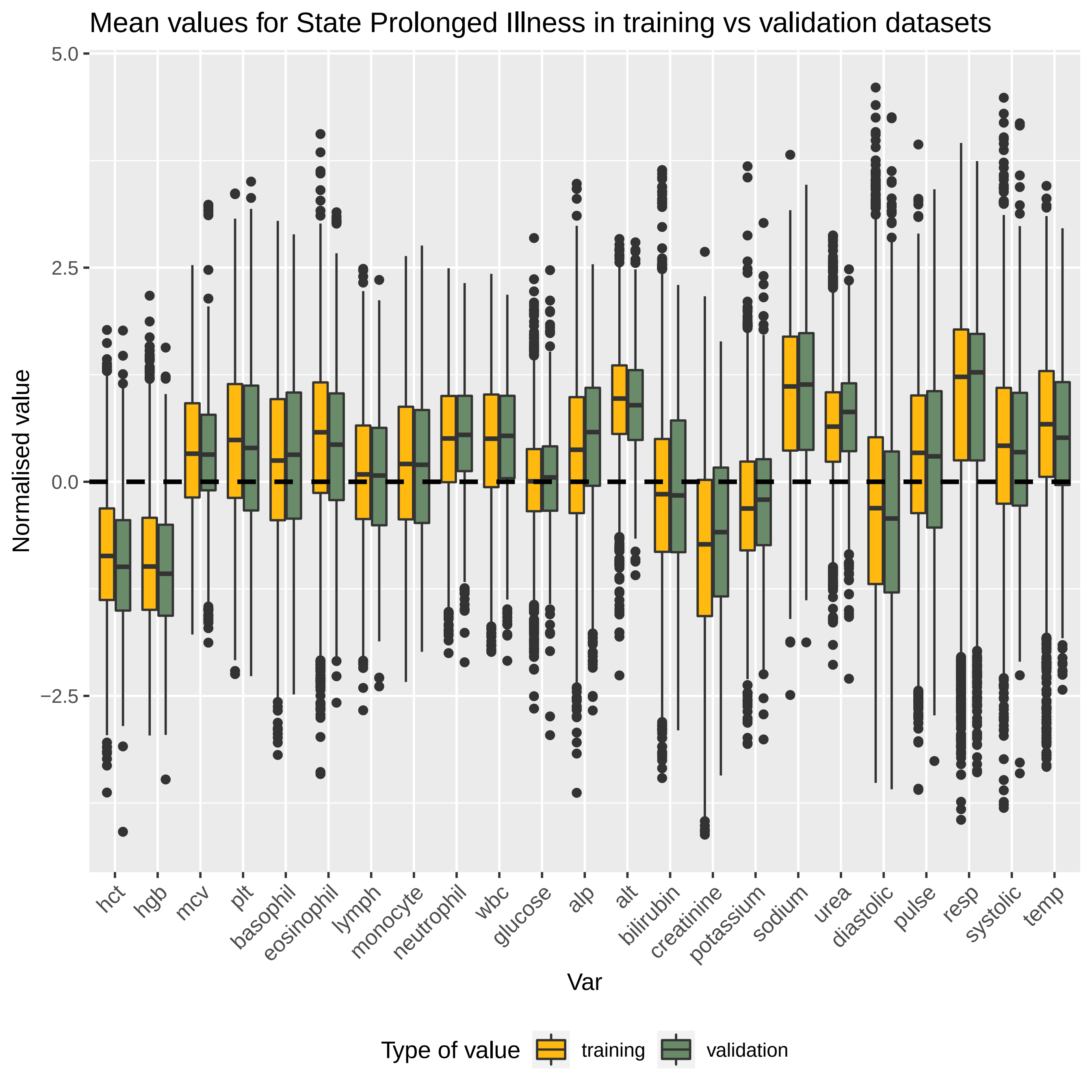

Mean values for State Prolonged Illness in training vs validation datasets
Normalised value
5.0
2.5
0.0
−2.5
hct
hgb
mcv
plt
basophil
eosinophil
lymph
monocyte
neutrophil
wbc
glucose
alp
alt
bilirubin
creatinine
potassium
sodium
urea
diastolic
pulse
resp
systolic
temp
Var
Type of value
training
validation

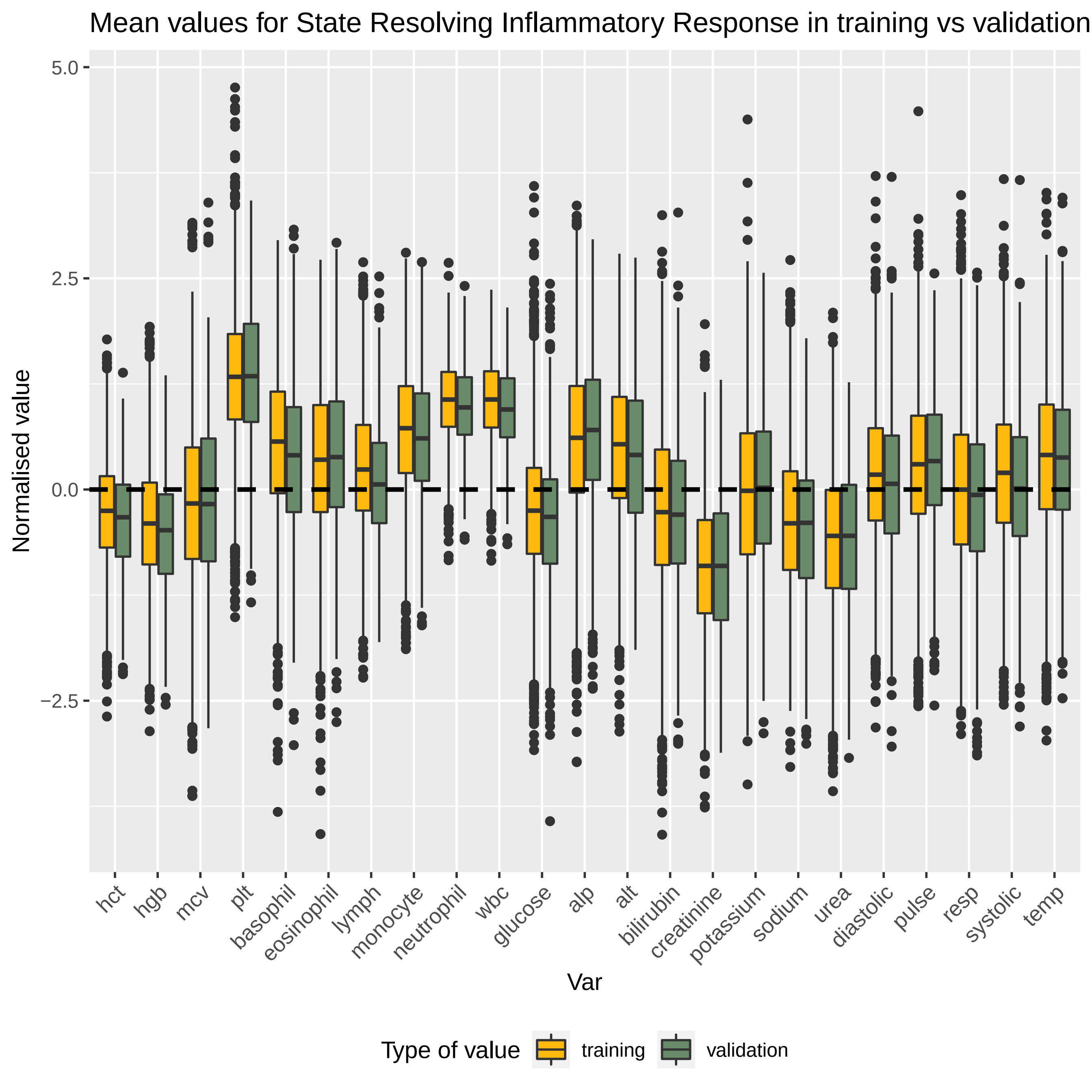

Mean values for State Resolving Inflammatory Response in training vs validation
Normalised value
5.0
2.5
0.0
−2.5
hct
hgb
mcv
plt
basophil
eosinophil
lymph
monocyte
neutrophil
wbc
glucose
alp
alt
bilirubin
creatinine
potassium
sodium
urea
diastolic
pulse
resp
systolic
temp
Var
Type of value
training
validation

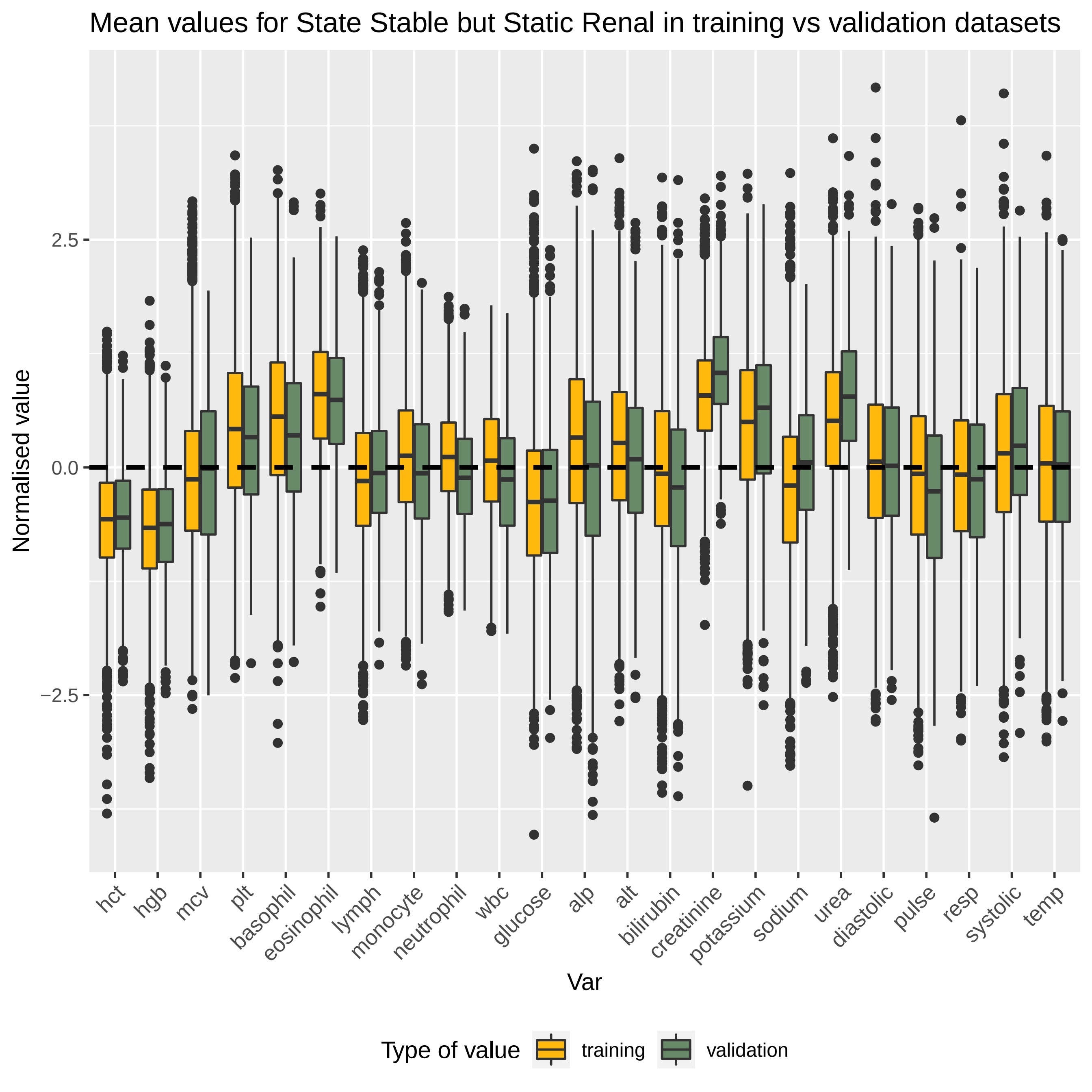

Mean values for State Stable but Static Renal in training vs validation datasets
Normalised value
2.5
0.0
−2.5
hct
hgb
mcv
plt
basophil
eosinophil
lymph
monocyte
neutrophil
wbc
glucose
alp
alt
bilirubin
creatinine
potassium
sodium
urea
diastolic
pulse
resp
systolic
temp
Var
Type of value
training
validation

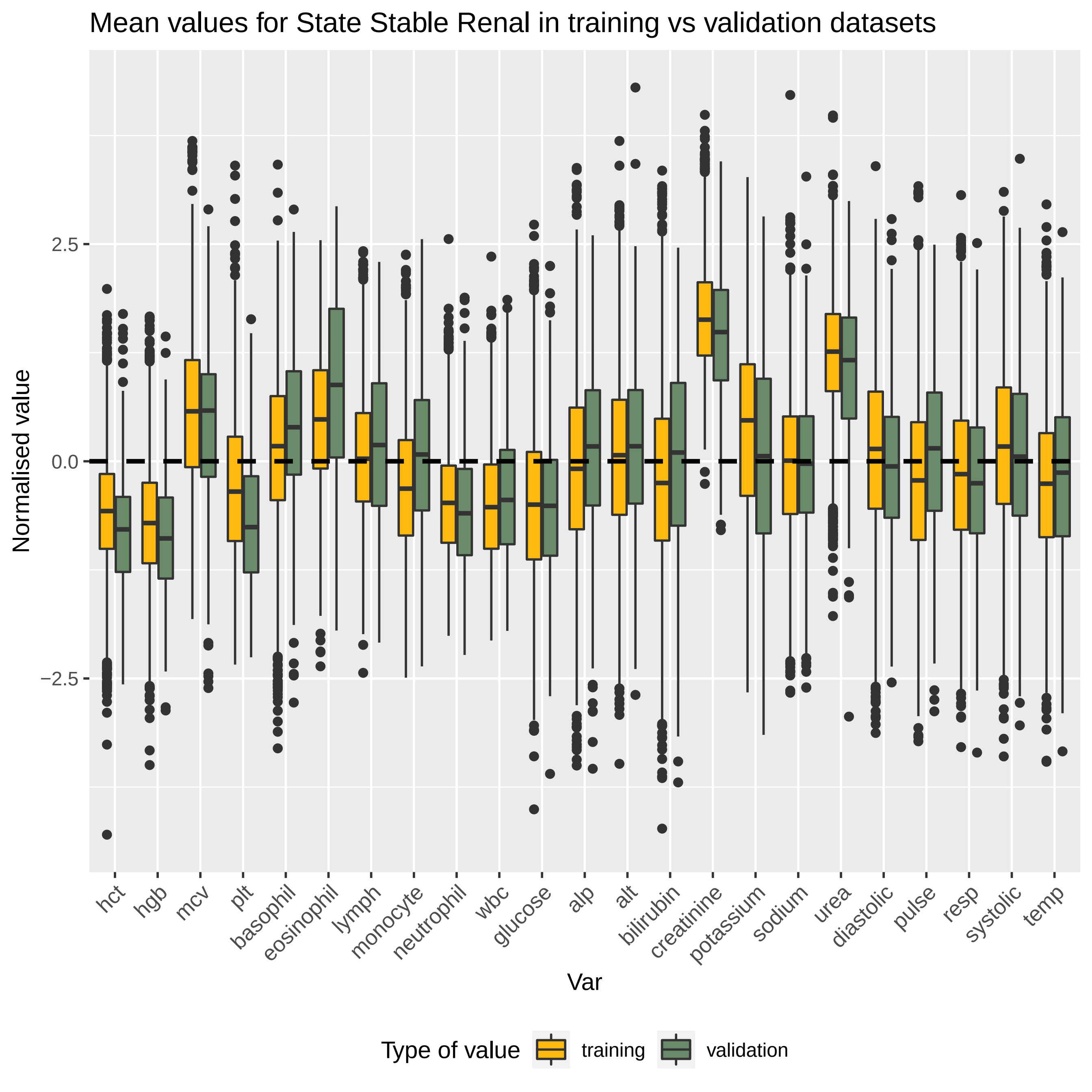

Mean values for State Stable Renal in training vs validation datasets
Normalised value
2.5
0.0
−2.5
hct
hgb
mcv
plt
basophil
eosinophil
lymph
monocyte
neutrophil
wbc
glucose
alp
alt
bilirubin
creatinine
potassium
sodium
urea
diastolic
pulse
resp
systolic
temp
Var
Type of value
training
validation

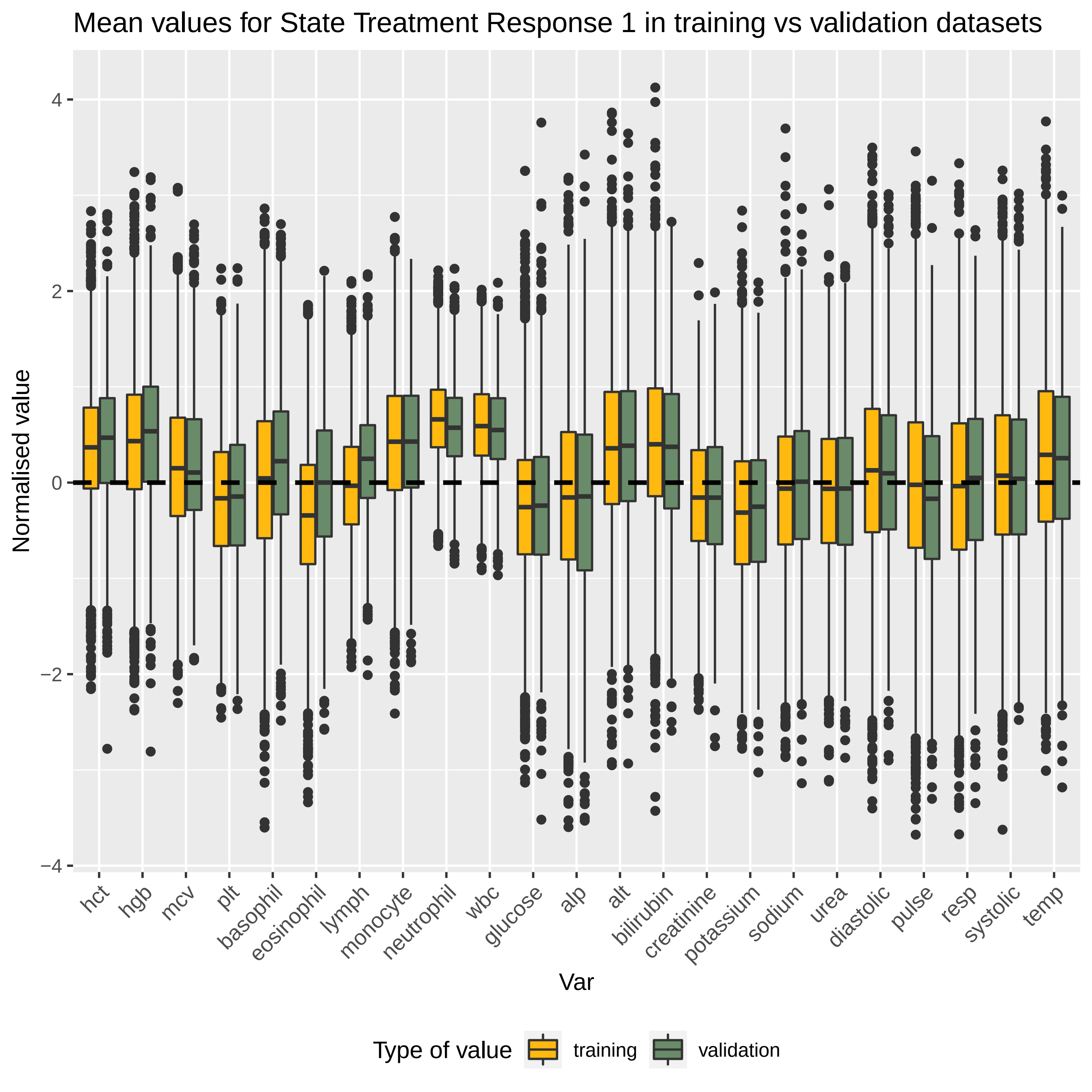
Mean values for State Treatment Response 1 in training vs validation datasets
Normalised value
4
2
0
−2
−4
hct
hgb
mcv
plt
basophil
eosinophil
lymph
monocyte
neutrophil
wbc
glucose
alp
alt
bilirubin
creatinine
potassium
sodium
urea
diastolic
pulse
resp
systolic
temp
Var
Type of value
training
validation

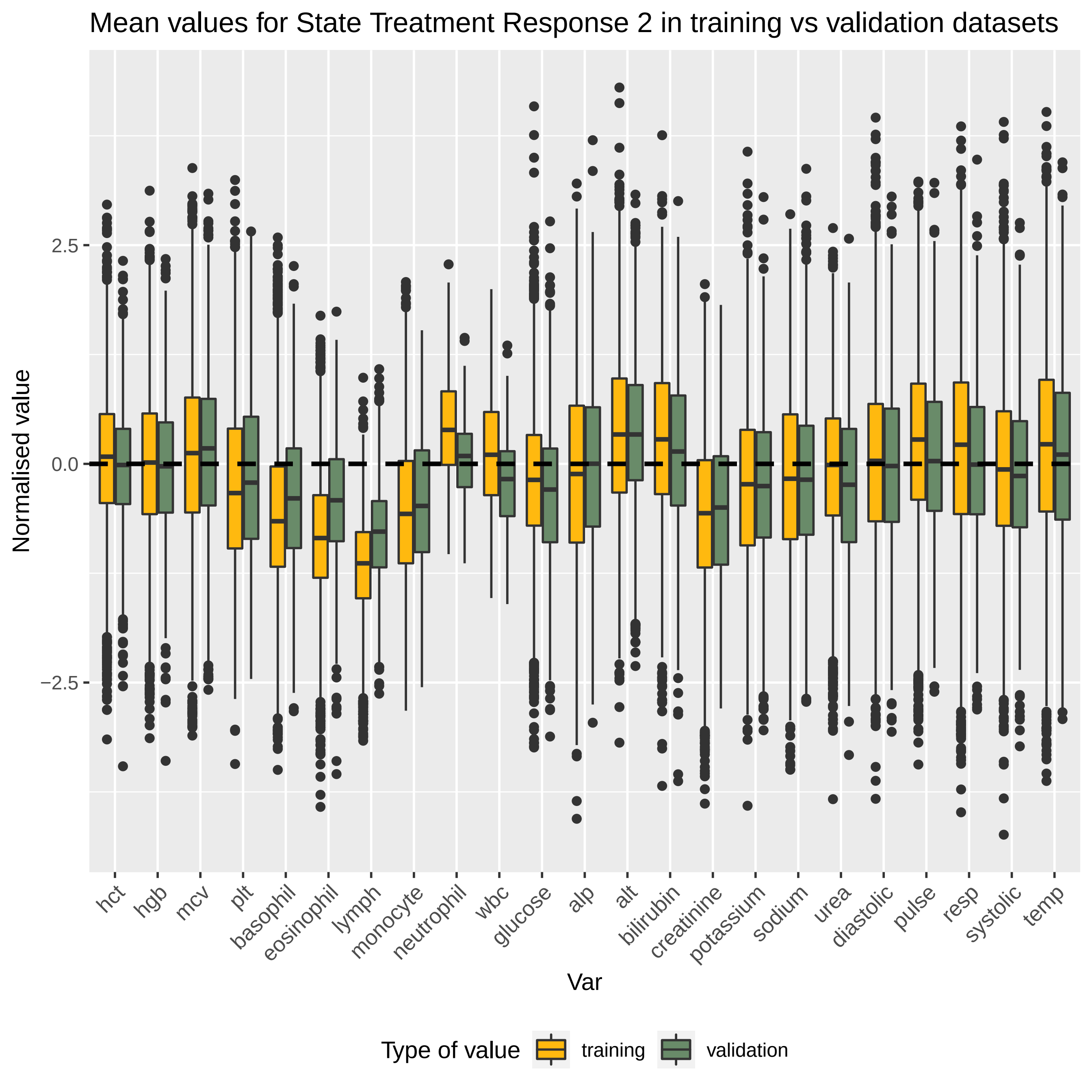

Mean values for State Treatment Response 2 in training vs validation datasets
Normalised value
2.5
0.0
−2.5
hct
hgb
mcv
plt
basophil
eosinophil
lymph
monocyte
neutrophil
wbc
glucose
alp
alt
bilirubin
creatinine
potassium
sodium
urea
diastolic
pulse
resp
systolic
temp
Var
Type of value
training
validation

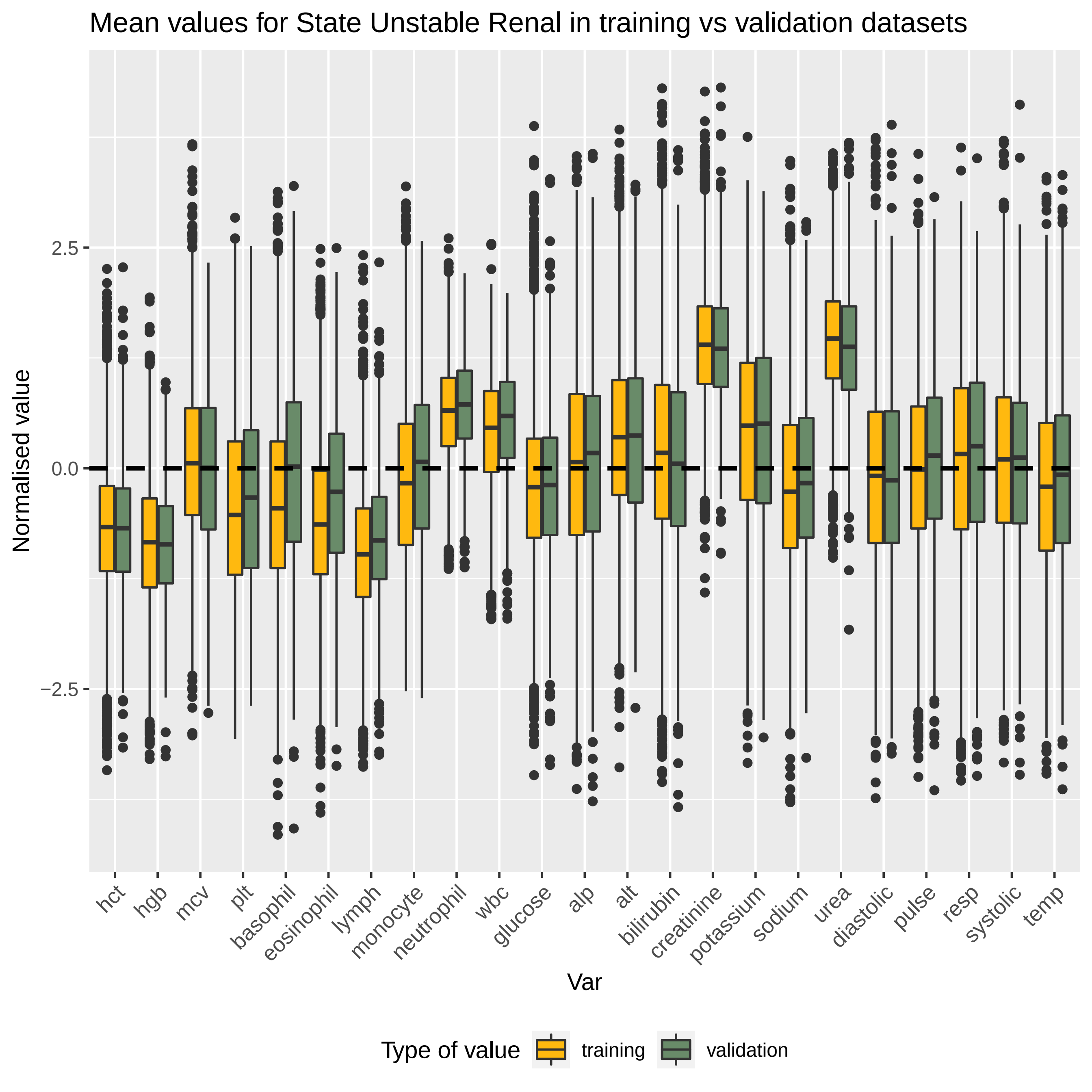

Mean values for State Unstable Renal in training vs validation datasets
Normalised value
2.5
0.0
−2.5
hct
hgb
mcv
plt
basophil
eosinophil
lymph
monocyte
neutrophil
wbc
glucose
alp
alt
bilirubin
creatinine
potassium
sodium
urea
diastolic
pulse
resp
systolic
temp
Var
Type of value
training
validation
